# Mendelian Randomization Study Reveals That Combined Spontaneous Free Cholesterol Diffusion and Reverse Cholesterol Transport Pathways Shift from Pro-Atherogenic to Anti-Atherogenic Effects

**DOI:** 10.1101/2025.05.21.25327960

**Authors:** Zhanqiang Yan, Jia Guo

## Abstract

**BACKGROUND:** The atherogenic properties of lipoproteins are well-characterized, in contrast to their putative atheroprotective effects which remain underinvestigated.

**METHODS:** We conducted this Mendelian randomization (MR) study to analyze the causal relationship between free cholesterol (FC) levels/total lipids/FC to total lipids ratio in lipoprotein subfractions and emergency coronary revascularization (for acute coronary syndromes [ACS]) (no controls excluded).

**RESULTS:** Positive causal association between the FC to total lipid ratio/FC levels in small HDL and emergency coronary revascularization (for ACS) suggest that spontaneous FC efflux from small HDL mediates pro-atherosclerotic effects independently.

Replacing FC levels with the FC-to-total-lipids ratio converts LDL from pro-atherogenic to anti-atherosclerotic particles. Or reduces pro-atherogenic effects in VLDL. In contrast, applying the same replacement in HDL particles reduces their anti-atherogenic properties.

The most remarkable finding is the negative causal relationship between the FC to total lipids ratio in (large LDL [P=1.88E-02, OR:0.78, 95%CI:0.64-0.96], medium LDL [P=1.76E-02, OR:0.78, 95%CI:0.64-0.96]) and emergency coronary revascularization (for ACS). This indicates that combined spontaneous FC diffusion and reverse cholesterol transport (RCT) can shift from pro-atherogenic to anti-atherogenic effects. The loss of bidirectional FC diffusion reduces the anti-atherosclerotic effect.

**CONCLUSIONS:** HDL can exhibit pro-atherosclerotic effects through spontaneously diffused FC. However, the bidirectional exchange of FC between HDL and other lipoproteins/cells—coupled with RCT and its alternative pathways—may attenuate or even reverse atherosclerosis, implying that the lack of this specific mechanism could drive proatherogenic processes. Notably, excessive cholesteryl ester (CE) production can counteract these protective mechanisms, potentially restoring a pro-atherosclerotic state.

## Introduction

Whereas the pro-atherosclerotic role of lipoproteins has been widely confirmed, comparatively little attention has been paid to their possible anti-atherosclerotic functions.

The consequences of this higher solubility in phospholipid vs. low solubility in water are important to free cholesterol trafficking among phospholipid surfaces. Similar to a water molecule evaporating from a rain drop, free cholesterol molecules desorb from phospholipid surfaces[19]. The tendency of free cholesterol to desorb from a phospholipid surface has been expressed as “ active cholesterol” and in the traditional physico-chemical term, as fugacity. Active cholesterol has been implicated in one model of reverse cholesterol transport [19][26], As with other lipids [19][27], the spontaneous transfer rate follows Kelvin’s law and increases with decreasing particle radius. For example, the halftimes for free cholesterol transfer from HDL (D□=□∼10□nm) and LDL (D□=□∼22□nm) to other lipid surfaces are 5 and 45□min respectively[19][28]. First, in humans HDL-FC has a plasma halftime of nearly 10□min [19][20], too fast for meaningful esterification by LCAT and far shorter than the lifetime of HDL-proteins (∼6□days)[19][21]. Thus, spontaneous transfer is responsible for much of the total cholesterol biodistribution and cytotoxicity, particularly in the state of high plasma free cholesterol [19][22][23][24]. Indeed, cellular free cholesterol accumulation underlies foam cell death [19][25].

High-density lipoprotein (HDL)-free cholesterol (FC) transfers to other lipoproteins and cells, the former by a spontaneous mechanism and the latter by both spontaneous and receptor-mediated mechanisms[3]. Our previous research data have verified the atherosclerotic effect of free cholesterol in small HDL. Free cholesterol can diffuse from HDL, causing cytotoxicity, especially in small HDL. Which means that the pro-atherosclerotic effect of small HDL is minimally affected by the changes in lipid level and lipid ratio. This is demonstrated by comparing data from lines 14(TG to total lipids ratio in small HDL [P=4.19E-05, OR:1.44,95% CI:1.21-1.71]) with those in lines 30(TG levels in small HDL [P=3.66E-04, OR:1.44,95% CI:1.18-1.76]) of Figure 3[4].

FC flux is bidirectional. In our earlier work[3][5]we studied the effects of HDL FC content on FC flux between HDL and human skin fibroblasts. At low HDL-FC content, HDL was an acceptor of FC from the cells but above 15 mol% FC, HDL was an FC donor to cells rather than an acceptor. Moreover, FC influx from the FC-rich HDL of Scarb1−/− mice to macrophages is higher (+300%) than that of wild-type mice with little difference in FC efflux[3][6]. Thus, the HDL-FC content determined the direction of the FC movement to both fibroblasts and macrophage cells. The plasma halftimes of LDL and HDL are on the order of 3 and 5 days [3][7][8]. In contrast, the spontaneous transfer halftimes of HDL- and LDL lipids is shorter, 5 and 45 min respectively [3][9]. Thus, during the long plasma halftimes of HDL and LDL, FC can exchange between the two lipoproteins many times thereby achieving dynamic equilibrium[3].

But the bidirectional spontaneous transfer of FC between HDL and other lipoproteins and cells can also exert anti-atherogenic effects. Moreover, which play a huge and irreplaceable role on a broader scale and to a greater extent. It doesn’t conflict with the mechanisms related to lipid and lipoprotein metabolism in the body. On the contrary, it is extremely and unimaginably beneficial to lipid and lipoprotein metabolism, thereby exerting an anti-atherosclerotic effect.

Whether the transfer of FC to lipoproteins or cells results in a harmful effect or a beneficial protective effect depends on the subsequent processing of FC. Lecithin: Cholesterol Acyltransferase (LCAT) synthesizes cholesteryl esters (CE) in HDL. Cholesteryl Ester Transfer Protein (CETP) transfers CE from HDL to various lipoproteins (e.g., CM, VLDL, IDL, LDL), which are then recognized and processed by diverse receptors. cholesterol can be recognized and processed by a wider range of receptors, such as LDL receptors, LDL Receptor Related Protein 1(LRP-1), VLDL receptors and other hepatic receptors, expanding the clearance pathway of cholesterol, expanding the liver’s ability to uptake, process and expel cholesterol, promoting the RCT and its alternative pathway. thereby exerting an anti-atherosclerotic effect.

Whereas excessive CE was transferred to other lipoproteins, lead to too many CE-enriched lipoproteins having elevated CE levels. Excessive CE loading alters lipoprotein surface topology, sterically hindering apolipoprotein B-100 recognition by LDL receptors, thereby establishing a cholesterol retention phenotype conducive to foam cell formation. thereby exerting pro-atherosclerotic effect.

Cholesterol in HDL is primarily transported to other lipoproteins or metabolized by the liver, minimizing harmful deposition. In contrast, cholesterol in VLDL, LDL, and peripheral cells has limited metabolic pathways, increasing its potential to promote atherosclerosis.

The spontaneous bidirectional transport of FC addresses this issue effectively:

1. FC diffuses from peripheral cells to HDL, where it is esterified to form CE.
2. HDL-CE can then be either metabolized by the liver or transferred to VLDL/LDL via CETP.
3. VLDL and LDL act as temporary reservoirs, supplying FC back to HDL when needed and preventing excessive CE accumulation. (Figure 1)

**Figure 1:**
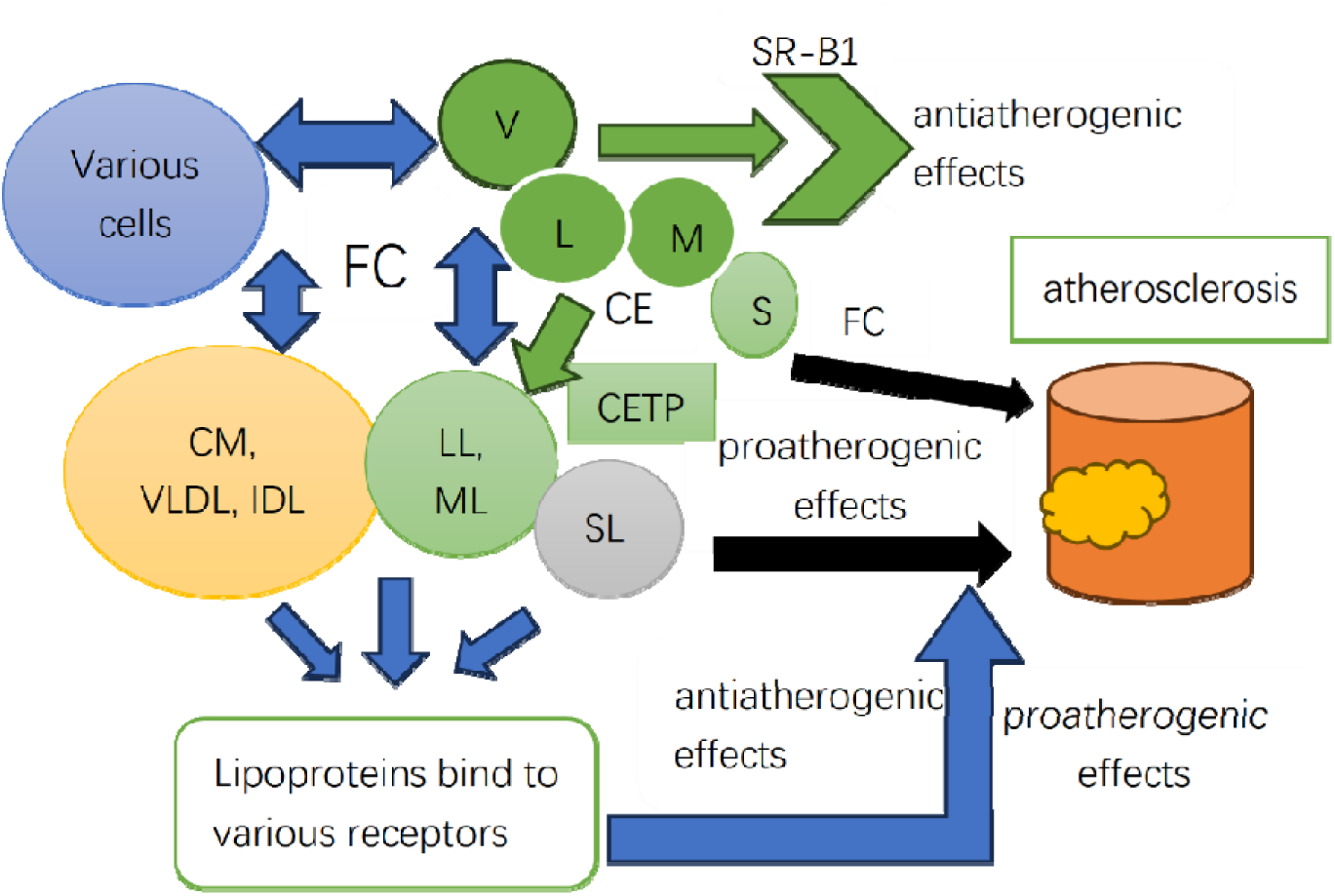
Lipids and lipoproteins FC free cholesterol, CE cholesterol ester, CETP: Cholesteryl Ester Transfer Protein, HDL: High-Density Lipoproteins, V: very large HDL, L: large HDL, M: medium HDL, S: small HDL, SR-B1: Class B Scavenger Receptor B1. CM: chylomicrons. VLDL: Very Low-Density Lipoproteins, IDL: Intermediate - Density Lipoproteins, LOL Low-Density Lipoproteins, LL: large LDL, ML medium LDL, SL small LDL.

Since the early discovery of the LDL receptor, many other related receptors that bind to native or modified lipoproteins have been identified[10]. LDLR, LRP1, LRP 2, LRP5/6, ApoER2, and VLDLR belong to the LDL receptor family. In addition to the initially assigned role as cholesterol transporters, these LDL receptor-related proteins have a wide range of additional functions that influence diverse physiological and pathological processes, including embryonic development, inflammation, haemodynamics, thrombosis, neointima hyperplasia, and atherosclerosis[10]-[15]. Cluster of differentiation 36 (CD36 also referred as SR-B2), scavenger receptor class B type 1 (SR-B1), and lectin-like oxidized low-density lipoprotein receptor-1 (LOX-1) belong to a large family of pattern recognition receptors. They are expressed in multiple cell types including hepatocytes, vascular SMCs, neurons, macrophages, fibroblasts, and endothelial cells[10][16][17][18]. These receptors interact with a broad range of ligands including LDL, IDL, VLDL, oxLDL, circulating native or modified lipoproteins, to promote or inhibit atherogenesis.

Mendelian randomization is an application of instrumental variable analysis, which aims to test a causal hypothesis in non-experimental data. In an MR analysis, genetic variants, commonly single nucleotide polymorphisms (SNPs), are used as instrumental variables (IVs) for the putative risk factor[2]. Since the transmission of genetic material from parents to offspring follows Mendel’s laws of inheritance, MR mimics the randomization process in randomized controlled trials (RCTs). This “natural randomization” helps mitigate confounding biases and avoids reverse causation, thereby strengthening causal inference.

The rapid growth of large-scale genome-wide association studies (GWAS) has identified numerous genetic variants associated with various traits, such as coronary heart disease and lipid profiles. These discoveries have expanded the pool of potential IVs, enabling broader applications of MR in diverse research areas.

## Methods

### Design

We employed a two-sample Mendelian randomization (MR) framework to infer causal relationships between [exposure] and [outcome], leveraging independent GWAS summary statistics for both instrument-exposure and instrument-outcome associations. The primary analysis was conducted using inverse-variance weighted (IVW) regression, with single-nucleotide polymorphisms (SNPs) serving as instrumental variables (IVs).

For valid causal inference, the selected genetic variants must satisfy the three key assumptions of MR:

The genetic variant (or multiple genetic variants) used as instrumental variable for the risk factor must (i) reliably associate with the risk factor under investigation (relevance assumption); (ii) not associate with any known or unknown confounding factors (independence assumption); and (iii) influence the outcome only through the risk factor and not through any direct causal pathway (exclusion restriction assumption) [2]. (Figure 2)

**Figure 2.**
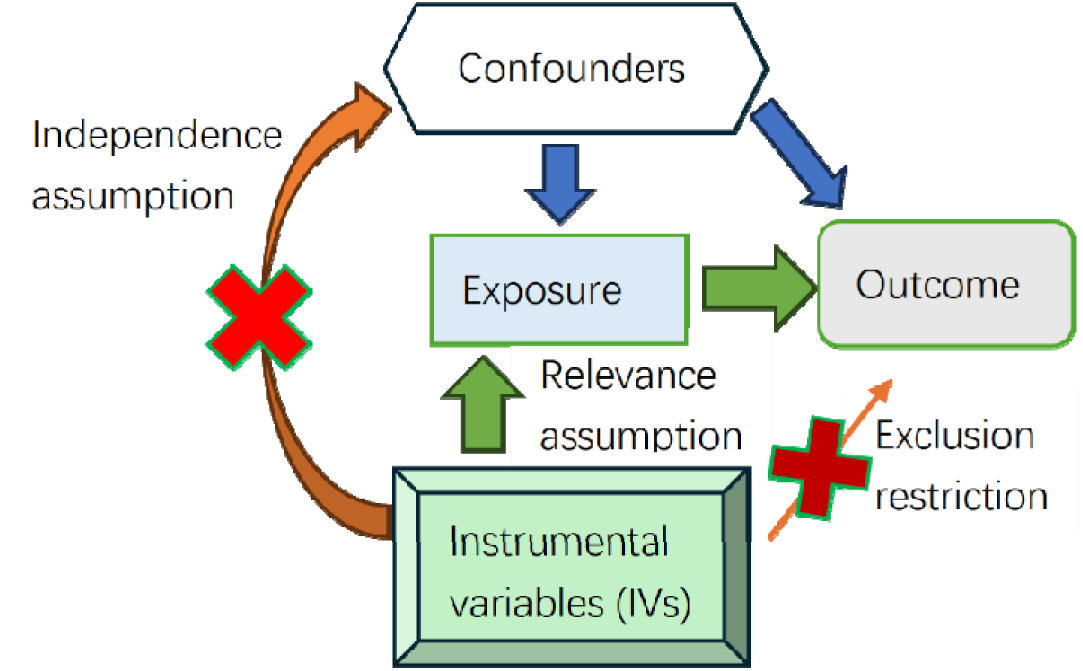
Graphical Abstract Exposure: Free cholesterol (FC) levels in lipoprotein subtractions Total lipid levels in lipoprotein subtractions FC to total lipids ratio in lipoprotein subtractions Outcome: Emergency coronary revascularization (for ACS) (no controls excluded)

### Data

The outcome data were derived from the FinnGen consortium, whereas the exposure data were obtained from the IEU OpenGWAS project (https://gwas.mrcieu.ac.uk/datasets/), To avoid potential population stratification bias, we confirmed no sample overlap between the two sources. All included participants were of European ancestry.

Table 1 presents detailed information on the datasets. All exposure datasets correspond to free cholesterol levels in HDL, differing only in their GWAS IDs (additional GWAS IDs are provided in Table 3). For brevity, only three representative exposure datasets are included in Table 1. Since all data were obtained from previously published, publicly available sources, no additional ethical approval was required

**Table 1.**
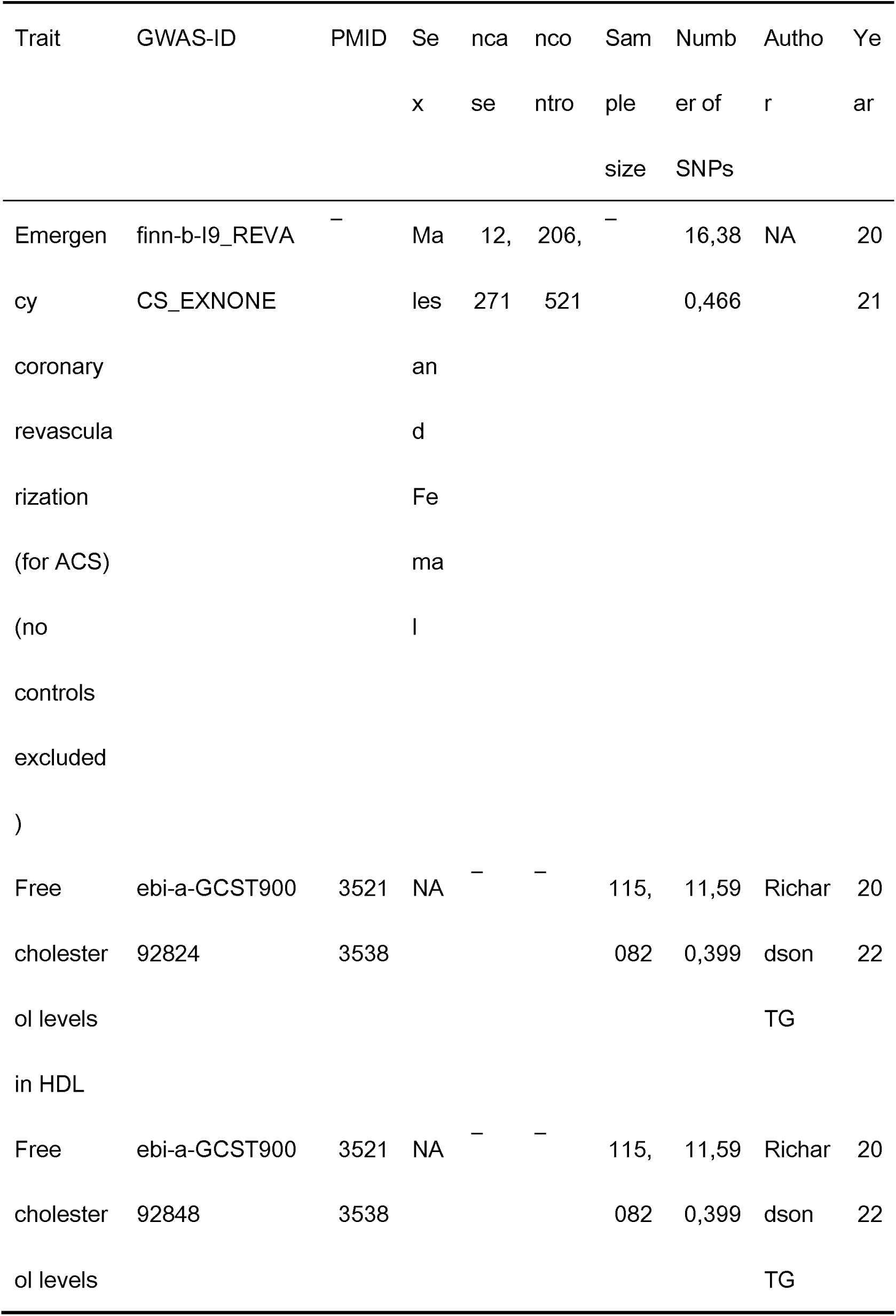
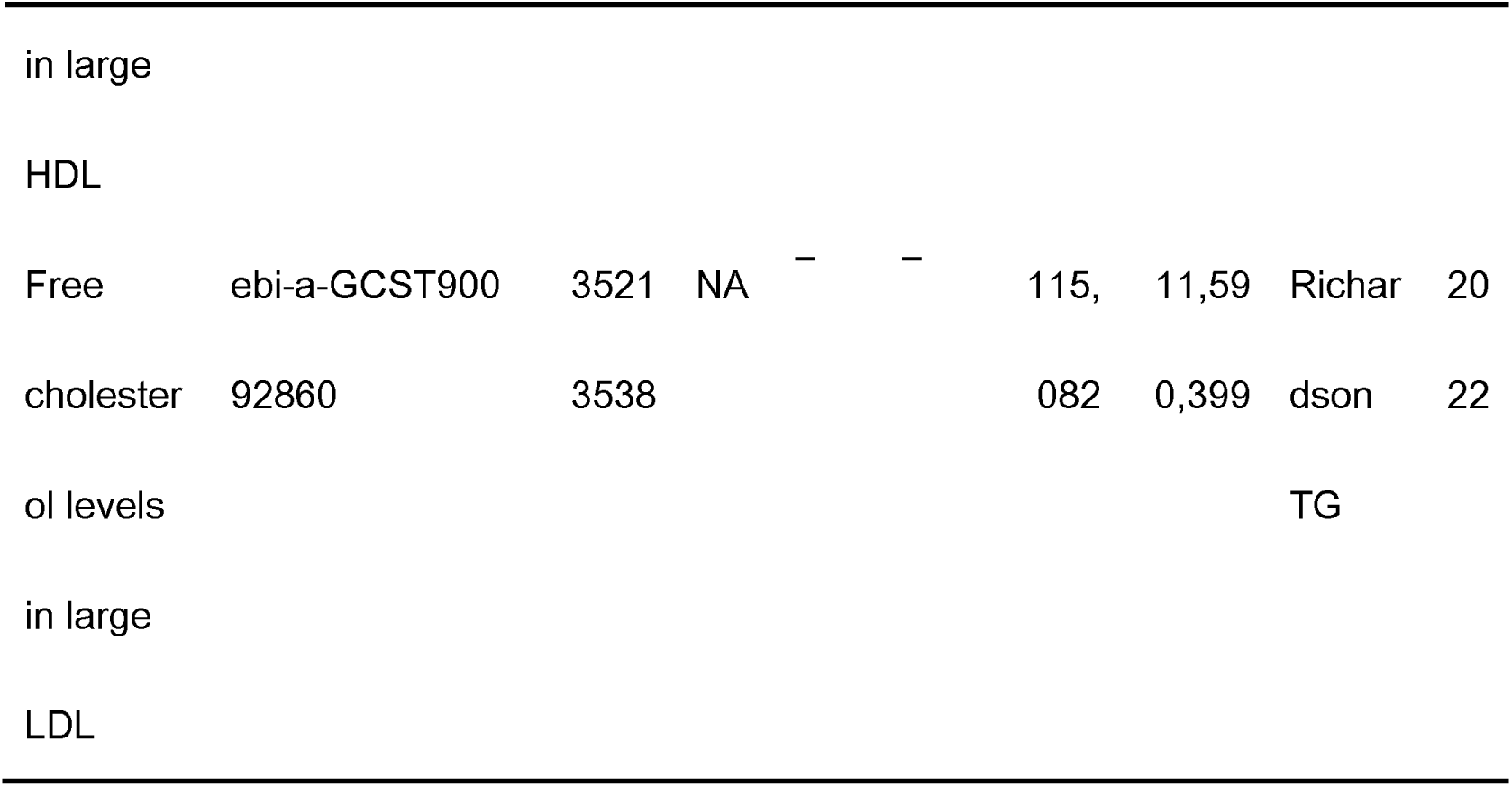
Characteristics of data of Exposures/Outcomes. Population: European.

We extracted instrumental variables (IVs) using the “extract_instruments” function from the R package “TwoSampleMR”. Only single nucleotide polymorphisms (SNPs) significantly associated with the exposure traits (p□<□5□×□10^-8^) were selected as IVs. To minimize linkage disequilibrium (LD) effects, we performed clumping with stringent parameters (r^2^ < 0.001, clump window = 10,000 kb). Additionally, we computed the F-statistic for each SNP, retaining only those with F > 10 to ensure strong instrument validity and reduce weak instrument bias.

### Statistical Analyses

In a two-sample MR study based on multiple genetic variants and summarized data, the causal estimate can be obtained by the inverse-variance weighted method, which is a meta-analysis of the single Wald ratios and is the most efficient method (greatest statistical power)[2]. Analyses were performed in R (version 4.4.2) using the “TwoSampleMR” and “MR-PRESSO” packages. Five MR methods were applied: IVW, MR-Egger, weighted median, simple mode, and weighted mode. Effect alleles and sizes were harmonized across datasets using the “harmonise_data”function in “TwoSampleMR”.

Heterogeneity was assessed via Cochran’s Q statistic. A significant Q test (p< 0.05) under the IVW method indicated substantial heterogeneity, warranting random-effects IVW regression. To evaluate horizontal pleiotropy, we used the “mr_pleiotropy_test” function; if pleiotropy was detected (p< 0.05), outlier SNPs were identified and removed via “MR-PRESSO” to correct the estimates.

## Results

### The MR Results

Figure 3 shows the results of IVW method. The random-effect IVW method was the primary analysis. The Mendelian randomization (MR) study results of the other four methods can be seen in supplemental material.

**Figure 3:**
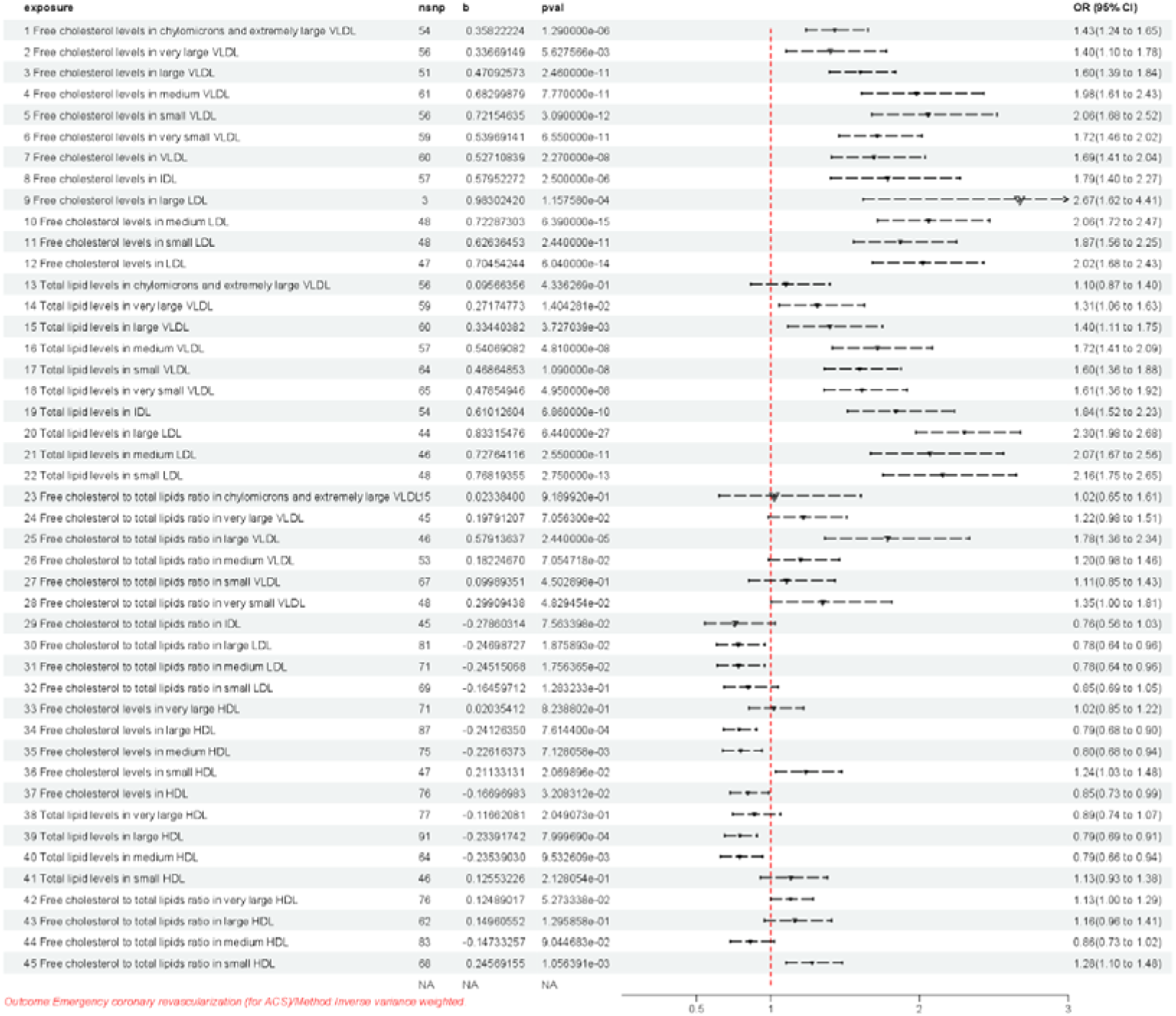
The Mendelian randomization (MR) study results. Outcome: Emergency coronary revascularization (for ACS)/Method: Inverse variance weighted (IVW) *Spontaneous FC efflux drives pro-atherosclerotic effects in small HDL (lines 36,45 of Figure 3). *In HDL, the pro-atherosclerotic effects can be attenuated or reversed through many mechanisms: CE synthesis, increased particle radius, RCT and alternative pathways. *In VLDL/LDL, the pro-atherosclerotic effects can be attenuated or reversed through many mechanisms: CE transportation, bidirectional exchange of FC between HDL and other lipoproteins/cells, RCT and alternative pathways.

This mechanistic framework is further substantiated by:

1. Replacing FC levels (lines 1-12 of Figure 3) with the FC to total lipids ratio (lines 23-32 of Figure 3) enhances anti-atherosclerotic effects.
2. FC-to-total lipids ratio in VLDL (lines 23-28 of Figure 3) vs. LDL (lines 30-32 of Figure 3) subfractions, the rate of FC spontaneous diffusion from larger particles (VLDL) to HDL is lower than that in small particles (LDL), leading to decreased anti-atherosclerotic effects. It indicates that the loss of bidirectional spontaneous diffusion of FC reduces the anti-atherosclerotic effect.
3. FC-to-total lipids ratio in LDL (lines 30-31 of Figure 3) vs. HDL (lines 42-44 of Figure 3) Subfractions, the former has the abilities not only to promote the synthesis and transportation of CE, but also to accept CE due to lower total lipid (mainly CE) levels, leading to anti-atherosclerotic effects. The latter enhances spontaneous FC efflux driving pro-atherosclerotic effects.
4. Comparison of total lipid levels in HDL (lines 38-41 of Figure 3) vs. VLDL/LDL (lines 13-22 of Figure 3) subfractions suggests that enhancing CETP activity exerts an anti-atherosclerotic effect, whereas inhibiting it promotes atherosclerosis.
5. Comparison of FC levels in HDL (lines 34,35,37 of Figure 3) vs. VLDL/LDL (lines 1-12 of Figure 3) subfractions, HDL has the ability to synthesize CE and can disperse and transfer CE to a greater variety and quantity of lipoproteins, thereby promoting the RCT and its alternative pathways. Therefore, higher FC levels in HDL can play an anti-atherosclerotic role.

The causal relationship between lipoprotein subfractions and emergency coronary revascularization (for ACS) demonstrates distinct patterns based on FC composition and particle characteristics. Specifically:

1. Inverse causal relationship was found in:

*Fc to total lipids ratio in:

large LDL P=1.88E-02, OR:0.78, 95% CI:0.64-0.96

medium LDL P=1.76E-02, OR:0.78, 95% CI:0.64-0.96

*FC levels in:

large HDL P=7.61e-04, OR:0.79, 95% CI:0.68-0.90

medium HDL P=7.13e-03, OR:0.80, 95% CI:0.68-0.94

HDL P=3.21e-02, OR:0.85, 95% CI:0.73-0.99

*Total lipid levels in large HDL P=8.00e-04, OR:0.79, 95% CI:0.69-0.91

*Total lipid levels in medium HDL P=9.53e-03, OR:0.79, 95% CI:0.66-0.94

2. Null causal relationship was observed in:

*FC to total lipids ratio in:

very large VLDL P=7.06e-02, OR:1.22,95% CI:0.98-1.51

medium VLDL P=6.61e-02, OR:1.28,95% CI:0.98-1.66

small VLDL P=4.5e-01, OR:1.11, 95% CI:0.85-1.43

very small VLDL P=4.83e-02, OR:1.35, 95% CI:1.00-1.81

IDL P=7.56e-02, OR:0.76, 95% CI:0.56-1.03

small LDL P=1.28e-01, OR:0.85, 95% CI:0.69-1.05

very large HDL P=5.27e-02, OR:1.13,95% CI:1.00-1.29

large HDL P=1.30e-01, OR:1.16,95% CI:0.96-1.41

medium HDL P=9.04e-02, OR:0.86,95% CI:0.73-1.02

*FC levels in very large HDL P=8.24e-01, OR:1.02,95% CI:0.85-1.22

*Total lipid levels in CM and extremely large VLDL P=4.34e-01, OR:1.10,95% CI:0.87-1.40

very large HDL P=2.05e-01, OR:0.89,95% CI:0.74-1.07

small HDL P=2.13e-01, OR:1.13,95% CI:0.93-1.38

3. Positive correlation emerged in:

*FC to total lipids ratio in:

CM and extremely large VLDL P=1.30e-02, OR:1.43,95% CI:1.08-1.89

large VLDL P=2.44e-05, OR:1.78,95%CI:1.36-2.34

small HDL P=1.06e-03, OR:1.28,95% CI:1.10-1.48

*FC levels in:

small HDL P=2.07e-02, OR:1.24,95% CI:1.03-1.48

CM and extremely large VLDL P=1.29e-06, OR:1.43,95% CI:1.24-1.65

very large VLDL P=5.63e-03, OR:1.40, 95% CI:1.10-1.78

large VLDL P=2.46e-11, OR:1.60, 95% CI:1.39-1.84

medium VLDL P=7.77e-11, OR:1.98, 95%CI:1.61-2.43

small VLDL P=3.09e-12, OR:2.06,95% CI:1.68-2.52

very small VLDLP=6.55e-11, OR:1.72, 95% CI:1.46-2.02

VLDL P=2.27e-08, OR:1.69, 95% CI:1.41-2.04

IDL P=2.50e-06, OR:1.79, 95% CI:1.40-2.27

large LDL P=1.16e-04, OR:2.67,95% CI:1.62-4.41

medium LDL P=6.39e-15, OR:2.06,95% CI:1.72-2.47

small LDL P=2.44e-11, OR:1.87,95%CI:1.56-2.25

LDL P=6.04e-14, OR:2.02,95% CI:1.68-2.43

*Total lipid levels in:

very large VLDL P=1.40e-02, OR:1.31,95% CI:1.06-1.63

large VLDL P=3.73e-03, OR:1.40, 95% CI:1.11-1.75

medium VLDL P=4.81e-08, OR:1.72, 95% CI:1.41-2.09

small VLDL P=1.09e-08, OR:1.60,95% CI:1.36-1.88

very small VLDL P=4.95e-08, OR:1.61,95% CI:1.36-1.92

IDL P=6.86e-10, OR:1.84,95% CI:1.52-2.23

large LDL P=6.44e-27, OR:2.30,95% CI:1.98-2.68

medium LDL P=2.55e-11, OR:2.07,95% CI:1.67-2.56

small LDL P=2.75e-13, OR:2.16,95% CI:1.75-2.65

### Sensitivity Analyses

To assess horizontal pleiotropy, we employed the “mr_pleiotropy_test” function in the “TwoSampleMR” package. A significant result (p<0.05) indicated the presence of horizontal pleiotropy in FC levels in (large VLDL [P=4.91e-2], CM and extremely large VLDL[P=0.03]), FC to total lipids ratio in (CM and extremely large VLDL[P=0.02], very large VLDL [P=4.96e-3], medium VLDL [P= 0.04], very large HDL[P=4.77e-2]). We used the“MR-PRESSO” to identify outlier SNPs, which are listed in Table 2. There was no horizontal pleiotropy (p□>□0.05) in the others.

**Table 2:** Outlier SNPs reported by the “MR-PRESSO” test.

|  |  |  |  |  |  |  |
| --- | --- | --- | --- | --- | --- | --- |
| e | Free | Free | Free | Free | Free | Free |
| x | cholesterol | cholesterol to | cholesterol | cholestero | cholesterol | choleste |
| p | levels in | total lipids | to total | l to total | levels in | rol to |
| o | large VLDL | ratio in | lipids ratio | lipids ratio | chylomicron | total |
| s |  | medium | in very | in very | s and | lipids |
| u | id:ebi-a-GC | VLDL \|\| | large VLDL | large HDL | extremely | ratio in |
| r | ST9009287 | id:ebi-a-GCS |  |  | large VLDL | chylomi |
| e | 2 | T90092921 | id:ebi-a-G<br>CST90093<br>021 | id:ebi-a-G<br>CST9009<br>3009 | \|\|<br>id:ebi-a-GC<br>ST9009304<br>4 | crons<br>and<br>extremel<br>y large<br>VLDL \|\|<br>id:ebi-a-GCST9<br>009304<br>5 |
| o | rs10455872 | rs10455872 | rs1045587 | rs112875 | rs10455872 | rs10455 |
| u | rs1047964 | rs11122450 | 2rs115911 | 651 | rs11684306 | 872 |
| t | rs10864726 | rs11591147 | 47 | rs116843 | 4 | rs11773 |
| l | rs11684306 | rs117733303 | rs1177333 | 064 | rs11773330 | 3303 |
| i | 4 | rs1260326 | 03 | rs154695 | 3 | rs12603 |
| e | rs11773330 | rs12740374 | rs1260326 | 4 | rs1883711 | 26 |
| r | 3 | rs2972156 | rs1267983 | rs190712 | rs28601761 | rs18016 |
|  | rs1561748 | rs328 | 4 | 692 | rs3127580 | 89 |
| S | rs4970836 | rs653178 | rs2954038 | rs998584 | rs964184 | rs31275 |
| N | rs73015034 | rs6905288 | rs2972138 |  | rs998584 | 80 |
| P |  | rs73015024 | rs602633 |  |  | rs5112 |
| s |  | rs964184 | rs6905288 |  |  | rs60223 |
|  |  |  | rs7301502 |  |  | 219 |
|  |  |  | 1 |  |  | rs73307 |
|  |  |  | rs7769791 |  |  | 905 |
|  |  |  | 7 |  |  | rs82184 |
|  |  |  | rs964184 |  |  | 0 |

Table 3 shows the “mr_pleiotropy_test” results, after removing outlier SNPs. There was no horizontal pleiotropy (p□>□0.05).

In our study, the inverse-variance weighted (IVW) Cochran’s Q test revealed significant heterogeneity (p < 0.05), prompting the use of the random-effects IVW method. This adjustment did not invalidate the Mendelian randomization (MR) results.

**Table 3:**
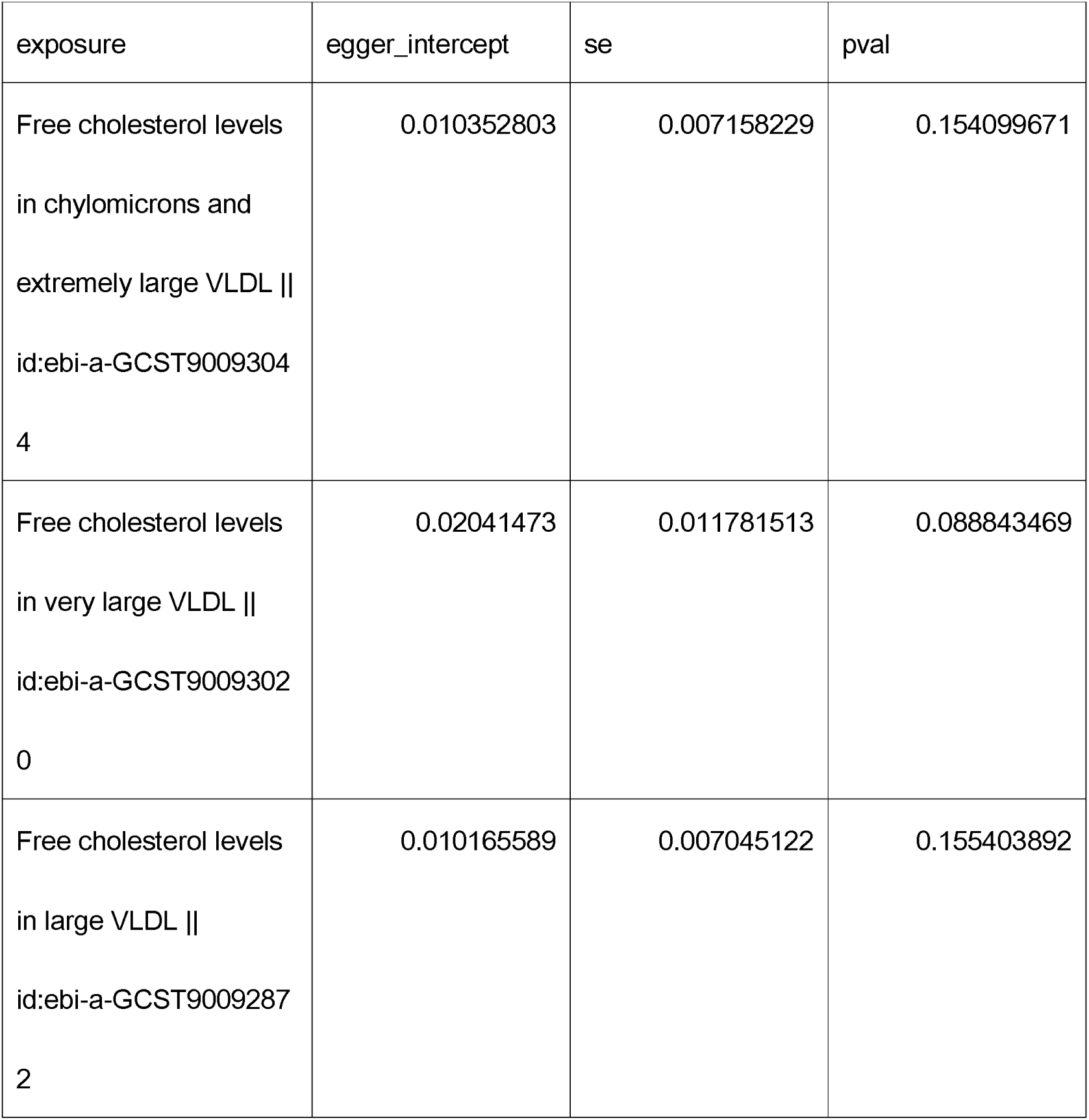
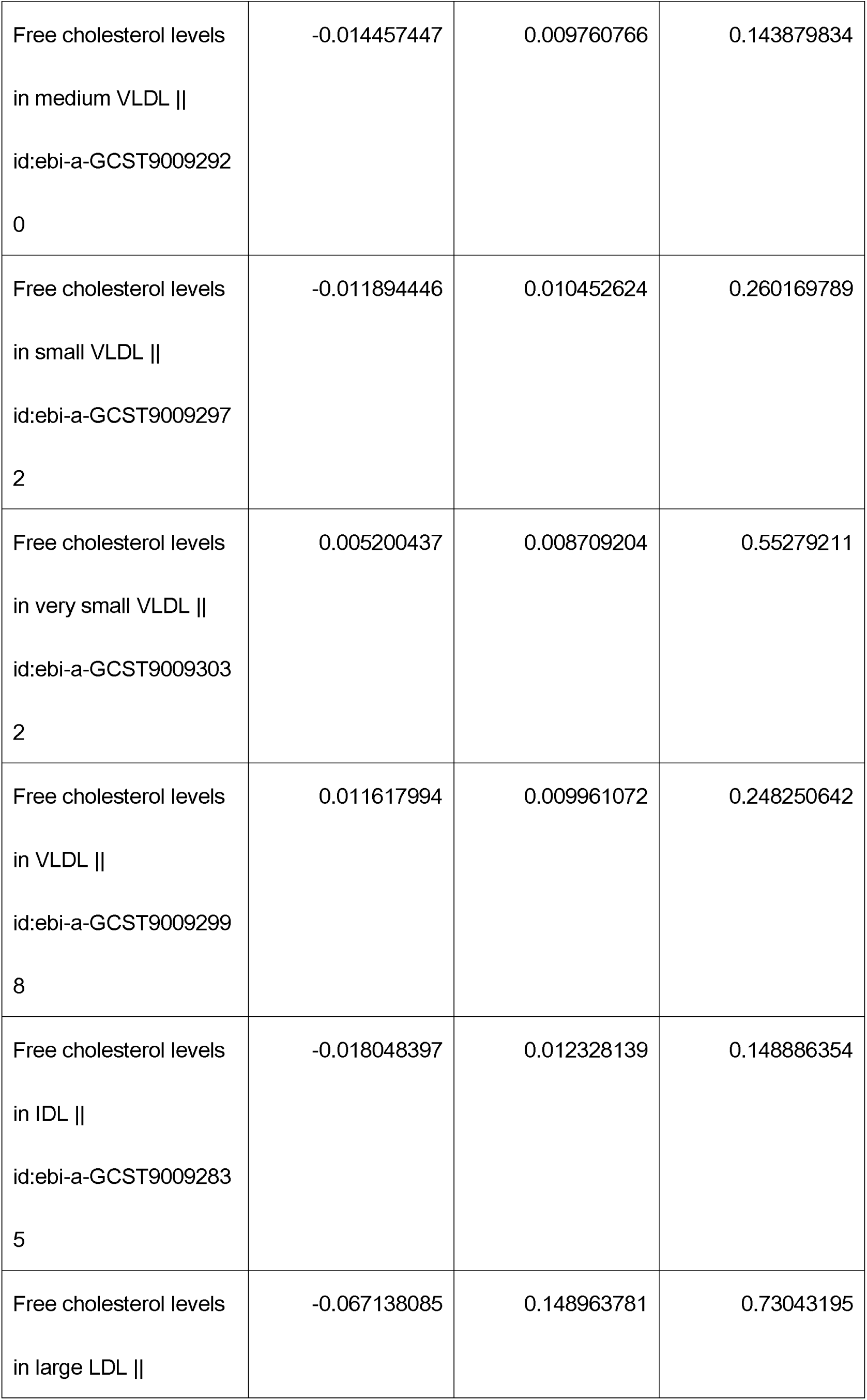
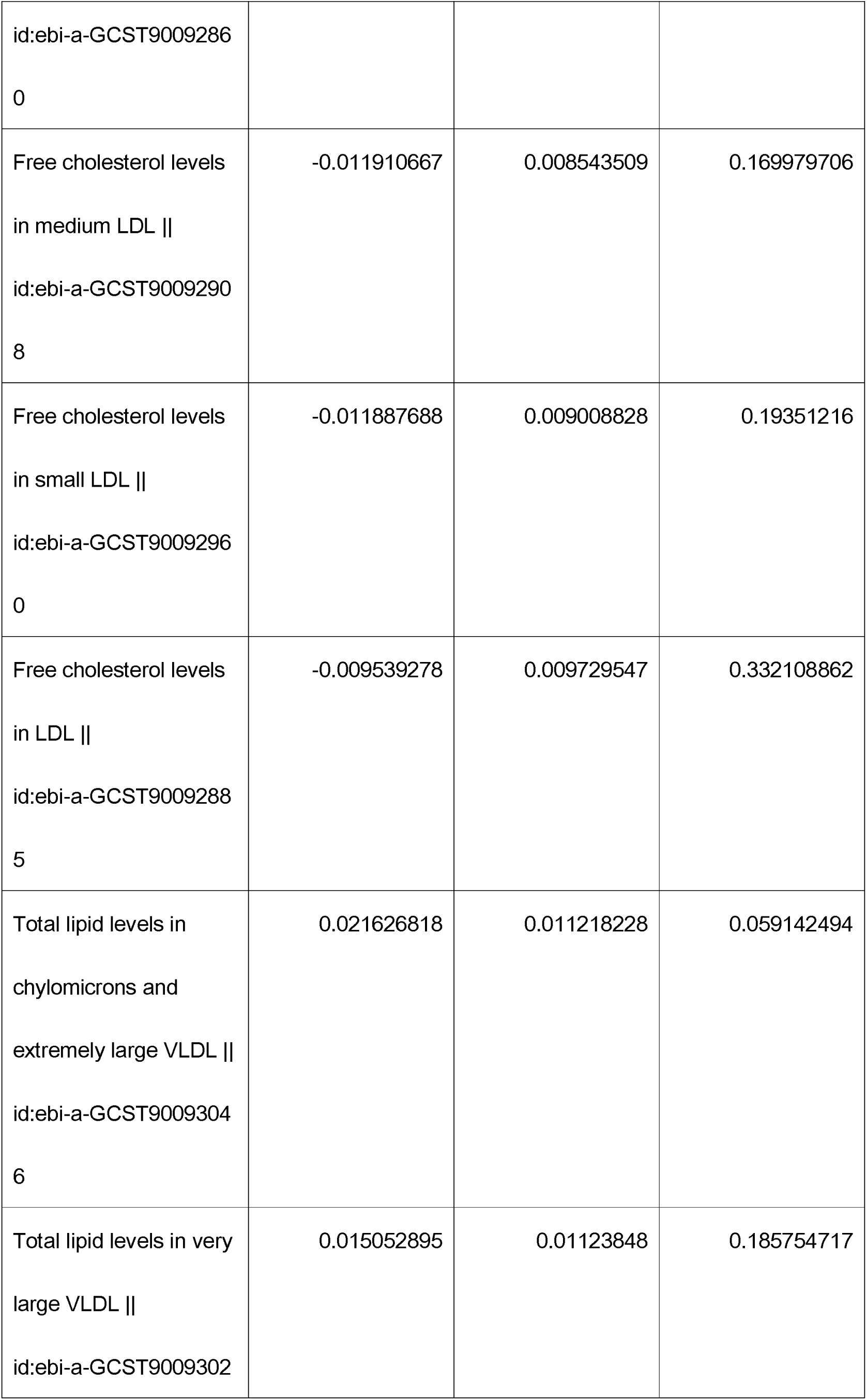
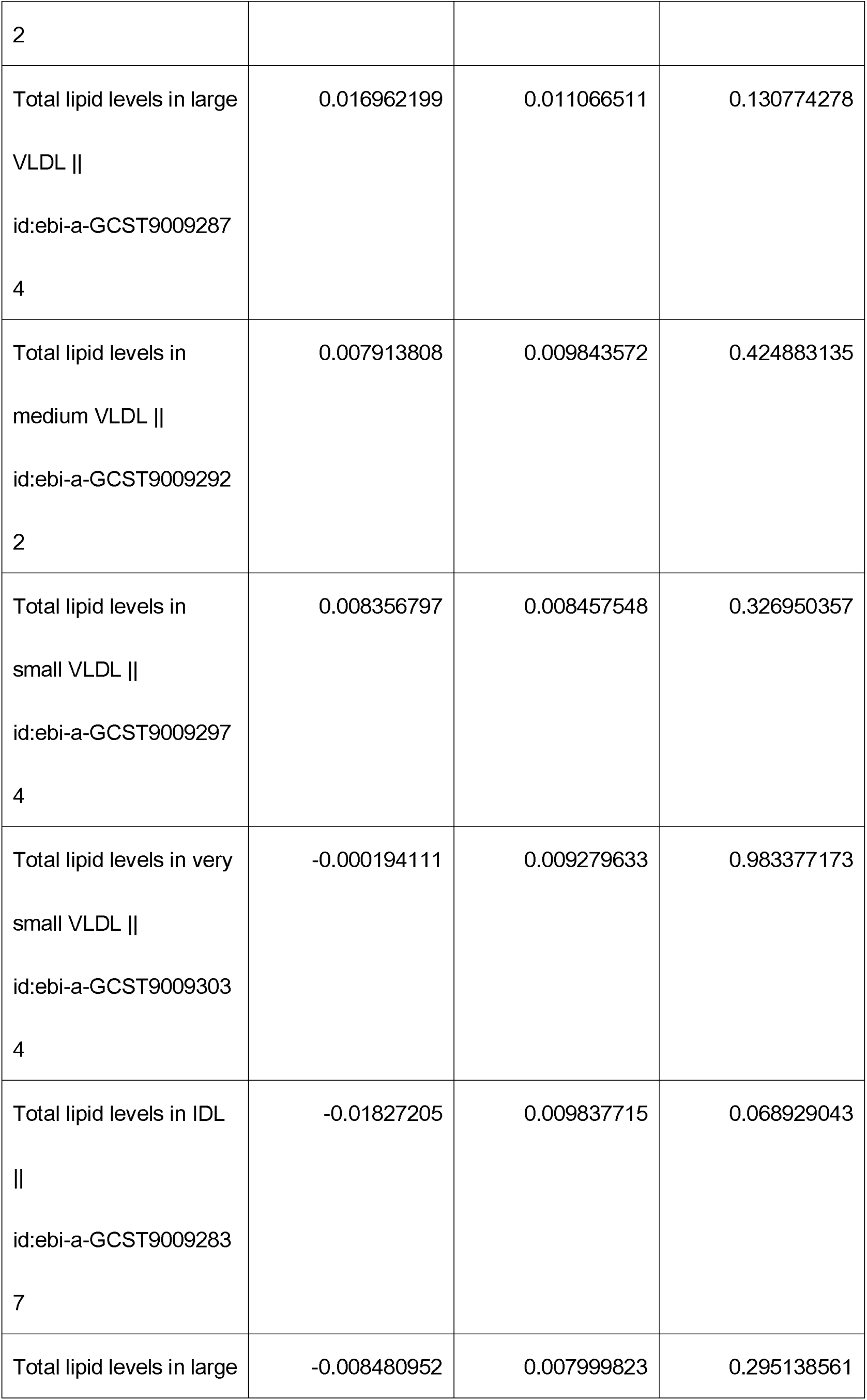
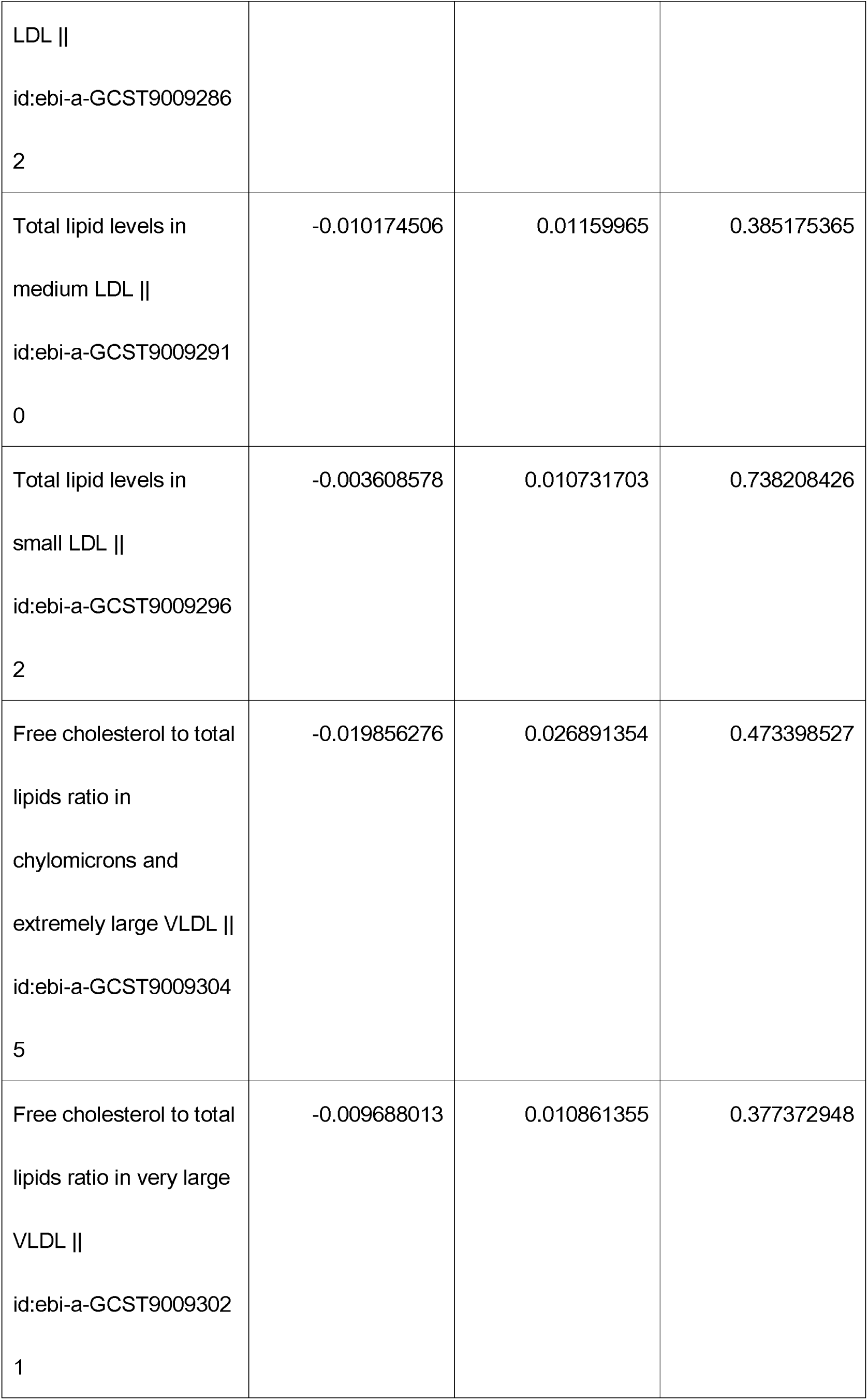
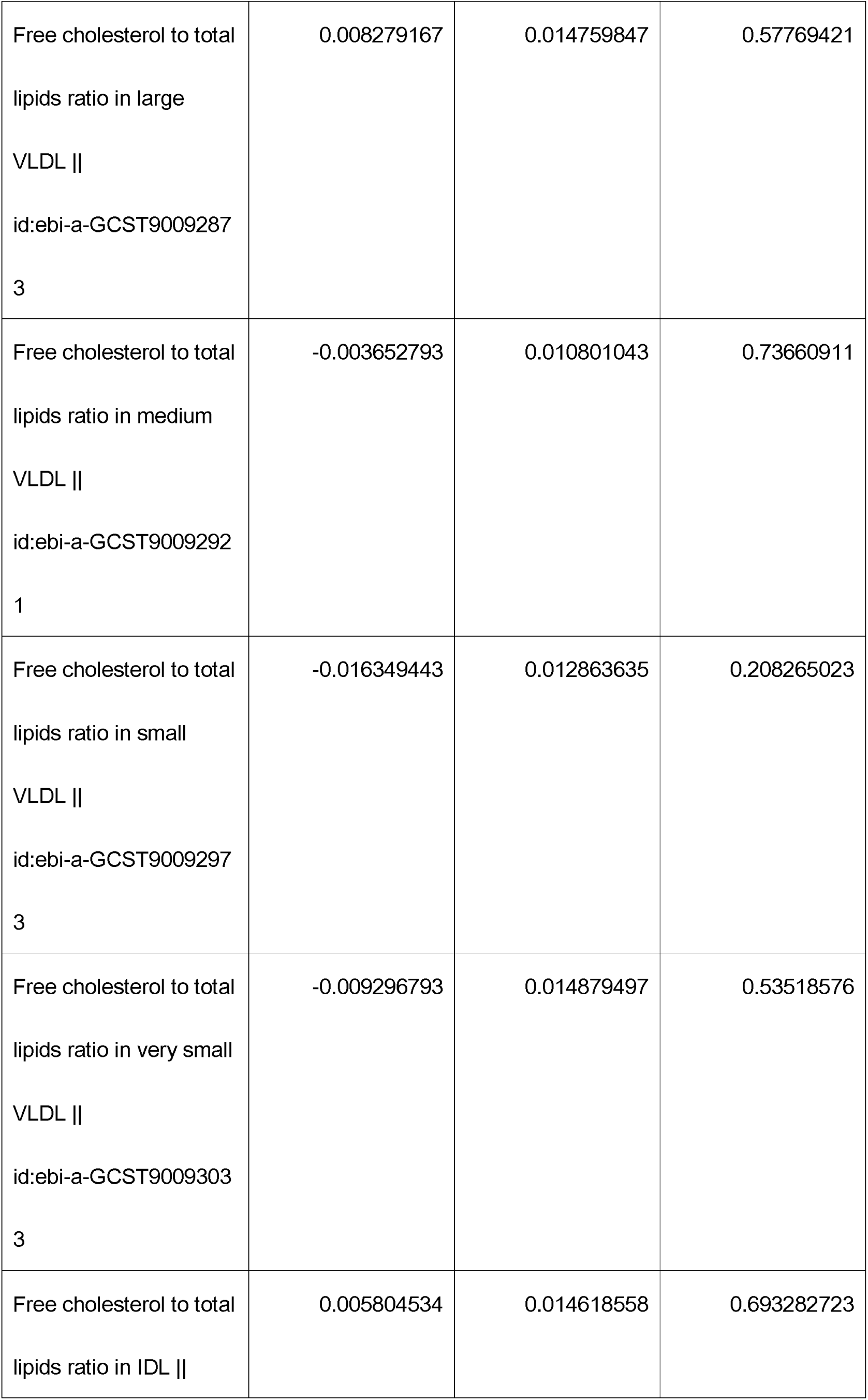
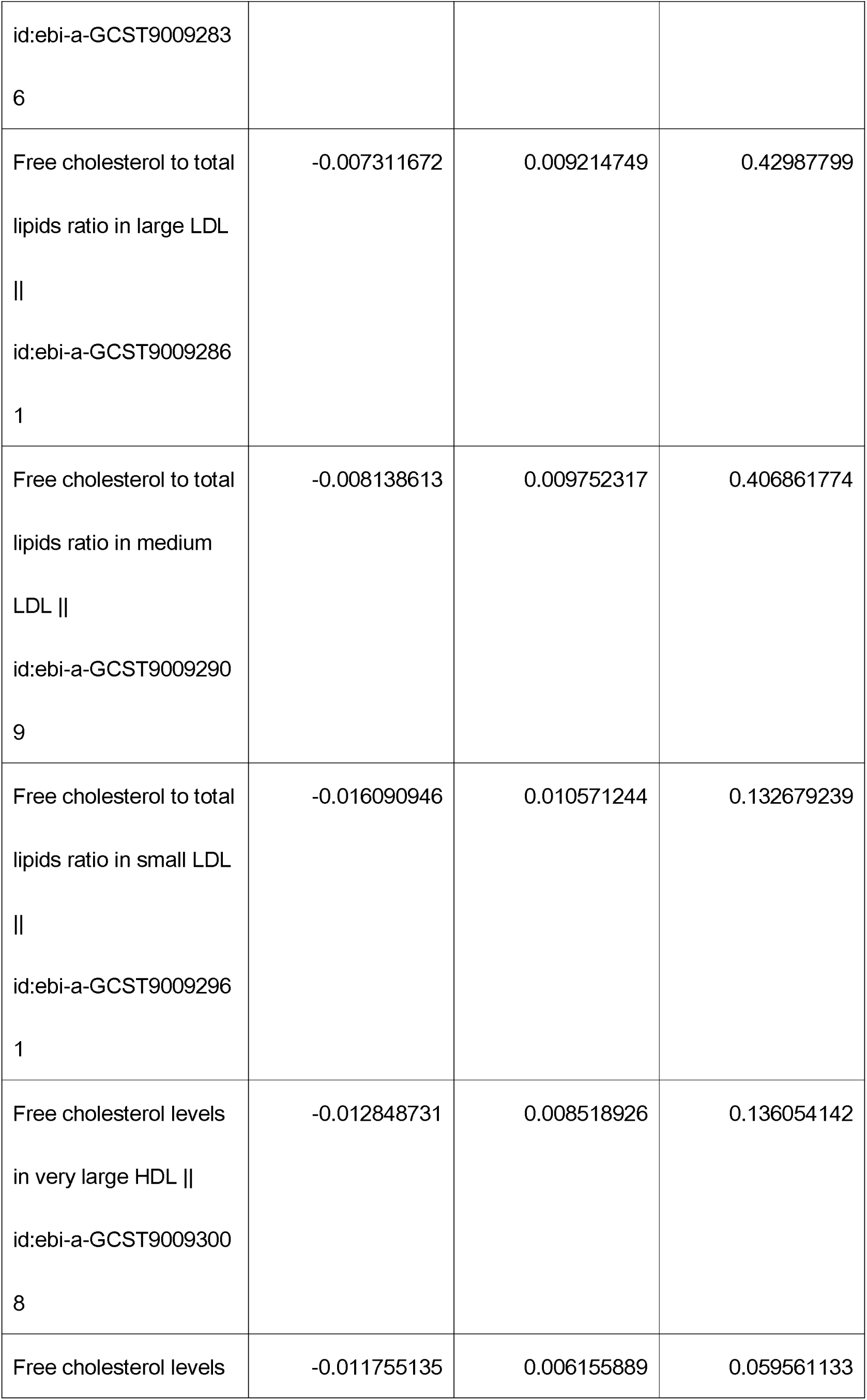
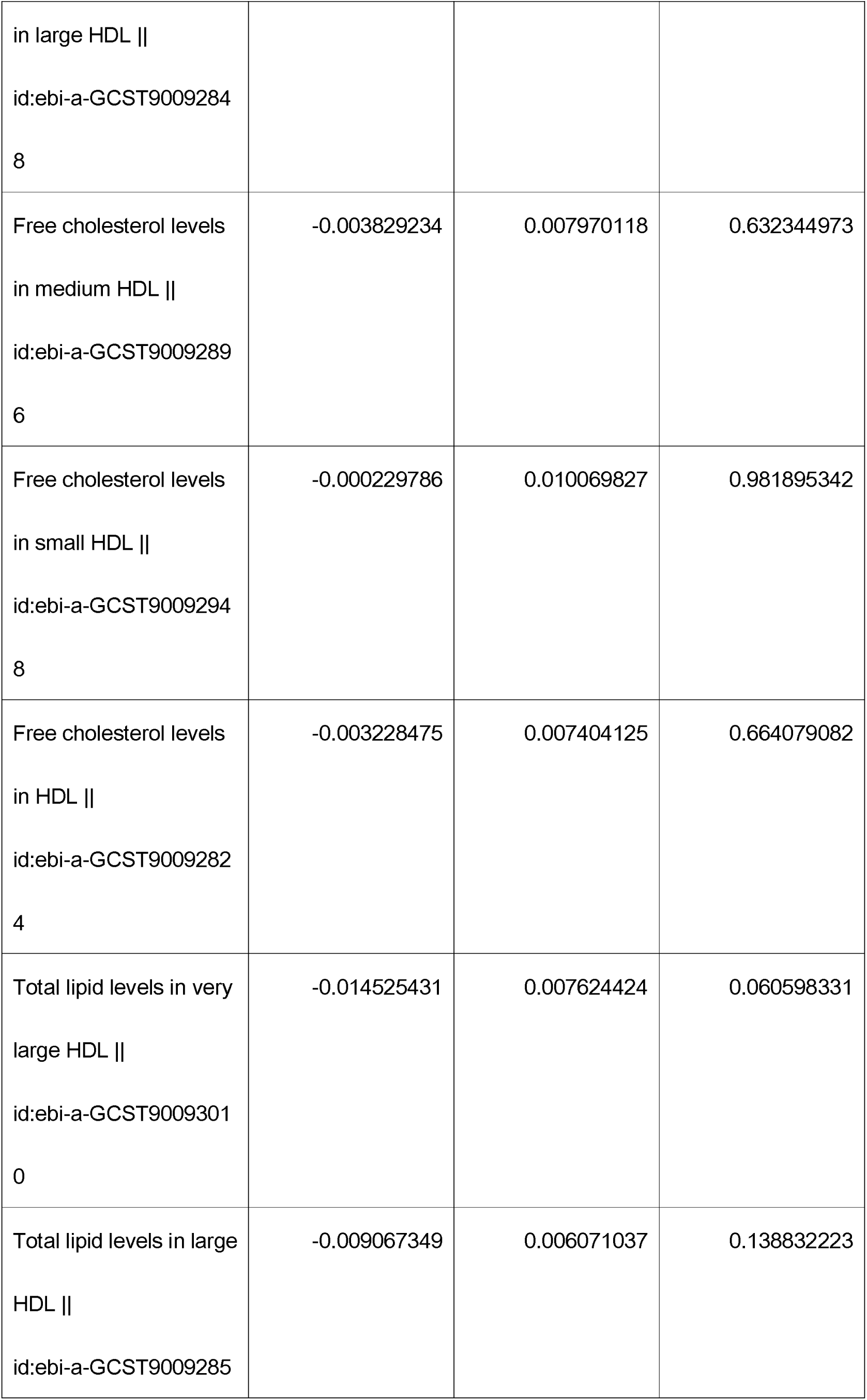
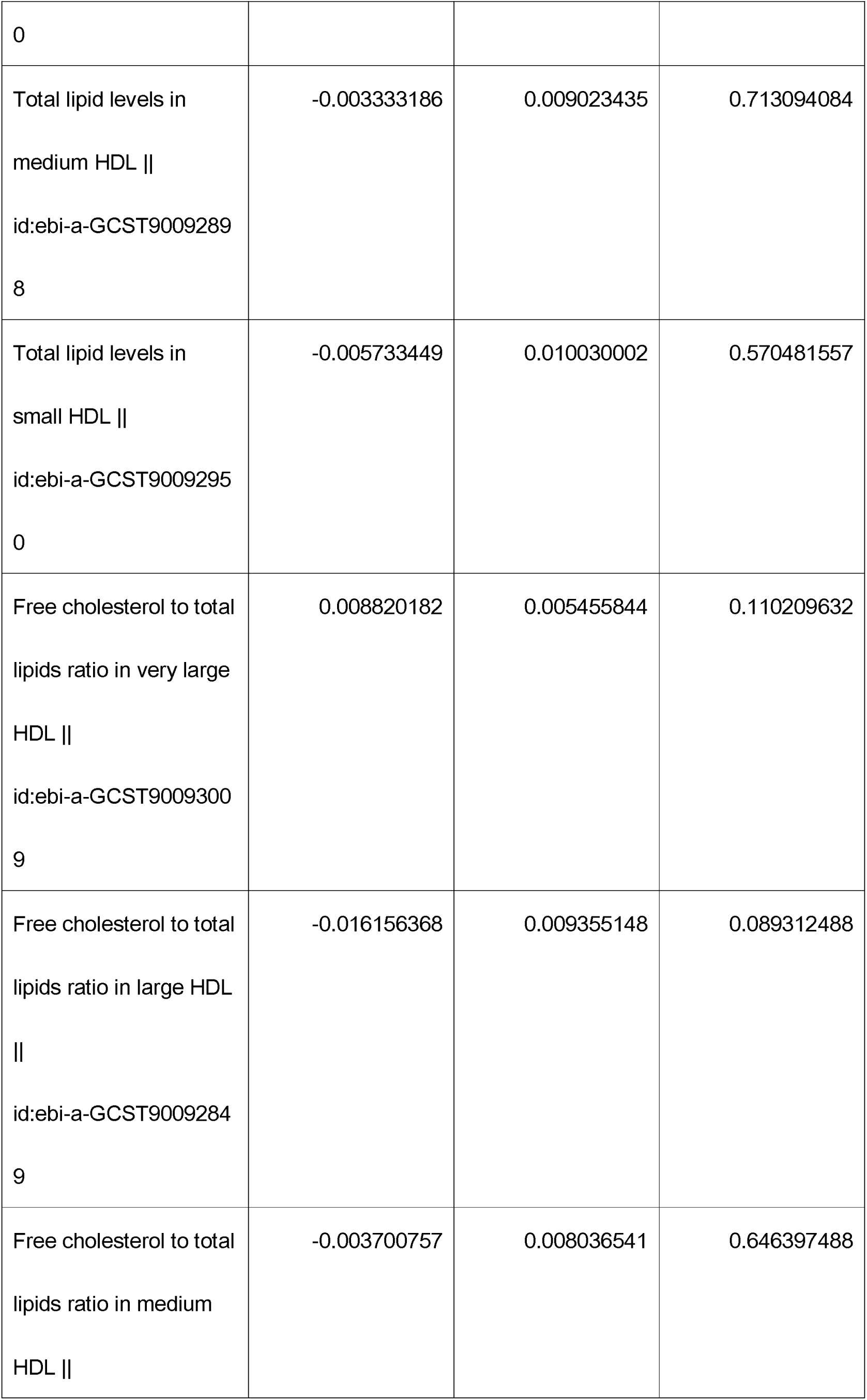
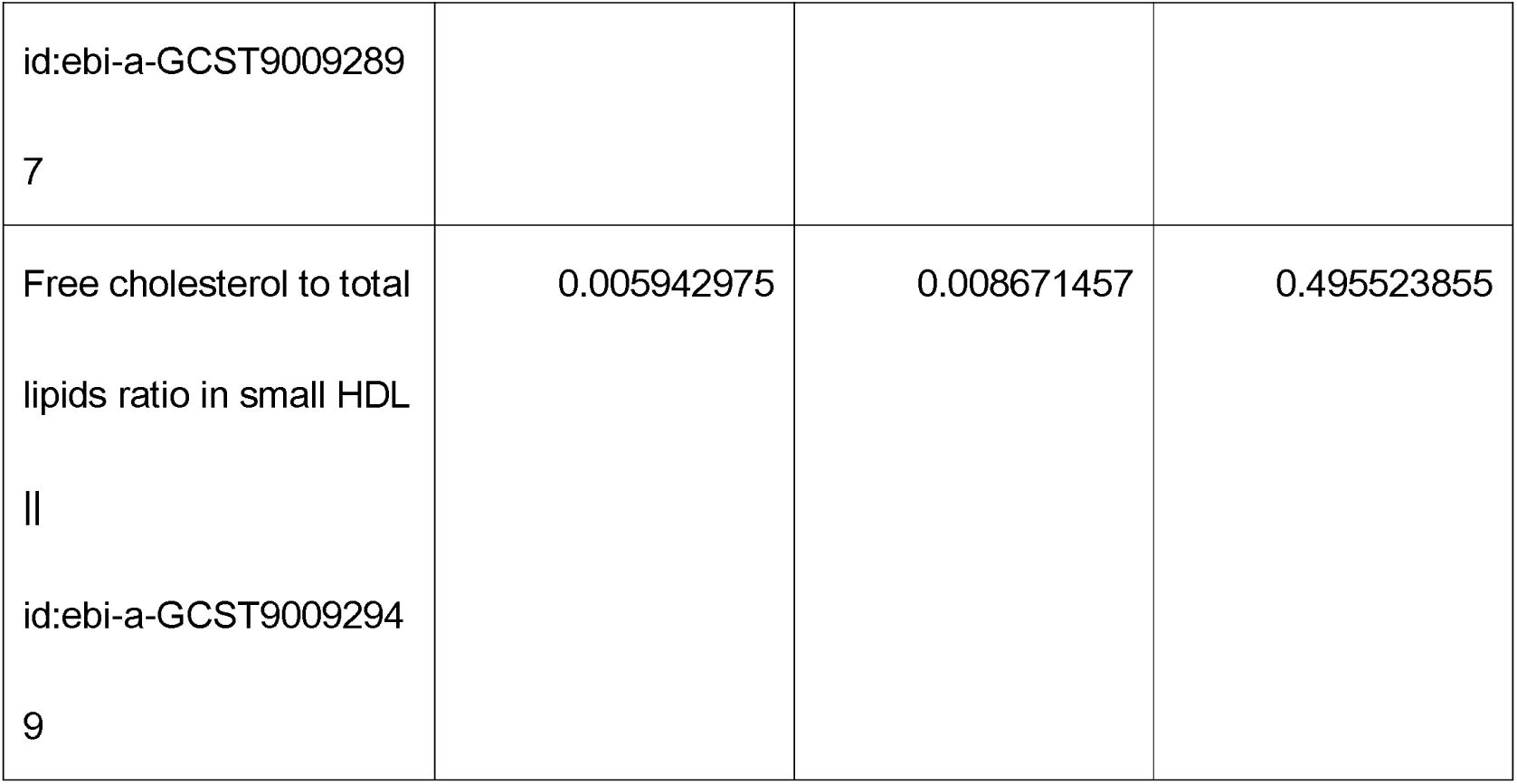
The “mr_pleiotropy_test” results, after removing outlier SNPs.

## Discussion

### Spontaneous FC efflux drives pro-atherosclerotic effects in HDL

The ratio of FC to the total/FC levels in small HDL had positive causal relationship with emergency coronary revascularization (for ACS) (no controls excluded), High-density lipoprotein (HDL)-free cholesterol (FC) transfers to other lipoproteins and cells, the former by a spontaneous mechanism and the latter by both spontaneous and receptor-mediated mechanisms[3]. Thus, during the long plasma halftimes of HDL and LDL, FC can exchange between the two lipoproteins many times thereby achieving dynamic equilibrium[3]. Free cholesterol can diffuse from HDL, causing cytotoxicity, especially in small HDL. This is demonstrated by comparing data from lines 36 of Figure 3 (FC levels in small HDL [P=2.07e-02, OR:1.24,95% CI:1.03-1.48]) with those in lines 45 of Figure 3 (FC to total lipids ratio in small HDL [P=1.06e-03, OR:1.28,95% CI:1.10-1.48]). The minimal alteration in the pro-atherosclerotic effect of small HDL following the substitution of free cholesterol (FC) levels (Figure 3, lines 36) with the FC-to-total lipids ratio (Figure 3, lines 45) suggests that FC diffusion from HDL particles may constitute a predominant pathogenic mechanism. This process likely mediates cytotoxicity and subsequent pro-atherogenic effects through cholesterol efflux from HDL to peripheral tissues. Spontaneous FC efflux from small HDL exerts pro-atherosclerotic effects, which are only weakly influenced by changes in lipid levels or lipid ratios.

As with other lipids[19][27], the spontaneous transfer rate follows Kelvin’s law and increases with decreasing particle radius. For example, the halftimes for free cholesterol transfer from HDL (D□=□∼10□nm) and LDL (D□=□∼22□nm) to other lipid surfaces are 5 and 45□min respectively[19][28].

In HDL, the pro-atherosclerotic effects can be attenuated or reversed through many mechanisms: CE synthesis, increased particle radius, RCT and alternative pathways.

Comparative analysis of the causal relationship between HDL particles across varying sizes and emergency coronary revascularization (for ACS), with larger HDL particles demonstrating attenuated atherosclerotic-promoting effects. The proatherogenic effects associated with spontaneous FC efflux diminish as particle radius increases. This observation further implies that free cholesterol (FC) diffusion from HDL particles may constitute a predominant pathogenic mechanism underlying these effects. This process likely mediates cytotoxicity and subsequent pro-atherogenic effects through cholesterol efflux from HDL to peripheral tissues.

When replacing FC levels (lines 34-35 of Figure 3) with the FC to total lipids ratio (lines 43-44 of Figure 3), we observed a shift in the causal relationship with emergency coronary revascularization (for ACS) from negative to non-significant. This suggests that a higher ratio of FC to total lipids in HDL reflects increased spontaneous FC efflux from HDL particles, which appears to exert stronger pro-atherosclerotic effects than absolute FC levels alone.

Supplying FC to HDL can promote the synthesis of CE and reduce the harmful effects of the spontaneous diffusion of FC, promoting the RCT and its alternative pathways.

The higher FC levels promote CE synthesis, facilitating CE to transfer from HDL to a broader range of lipoproteins (e.g., CM, VLDL, IDL, and LDL), enabling cholesterol to be recognized and processed by various receptors. Which serves as an alternative pathway for the transport cholesterol to the liver[1]. This mechanism enhances the liver’s capacity to uptake, process, and excrete cholesterol, thereby promoting RCT. Consequently, FC levels in large and medium HDL exhibit a negative causal relationship with emergency coronary revascularization (for ACS).

But the concentrations of CE are the highest in very large HDL particles, The elevated FC content in these very large HDL particles further enhances CE production, leading to excessive formation of cholesterol-rich lipoprotein particles. The accumulation of CE-enriched lipoproteins impairs receptor-mediated clearance, reducing cellular uptake and promoting atherosclerotic plaque formation. Consequently, the pro-atherogenic effects of these particles are enhanced. Notably, the causal relationship between FC levels in HDL subfractions and emergency coronary revascularization (for ACS) shifts from negative (FC levels in medium and large HDL had negative causal relationship with emergency coronary revascularization, observed in lines34,35 of Figure 3) to no (FC levels in very large HDL had no causal relationship with it, observed in lines33 of Figure 3).

In VLDL/LDL, the pro-atherosclerotic effects can be mitigated or reversed through many mechanisms: CE transportation, bidirectional exchange of FC between HDL and other lipoproteins/cells, RCT and alternative pathways.

This mechanistic framework is further substantiated by replacing FC levels (lines 1-12 of Figure 3) with the FC to total lipids ratio (lines 23-32 of Figure 3). Overall, their anti-atherosclerotic effect is enhanced.

The most remarkable finding is the negative causal relationship between the FC to total lipids ratio in (large LDL [P=1.88E-02, OR:0.78, 95%CI:0.64-0.96], medium LDL [P=1.76E-02, OR:0.78, 95%CI:0.64-0.96]) (lines 30,31 of Figure 3) and emergency coronary revascularization (for ACS). which involves diverse underlying mechanisms: (1) the bidirectional exchange of FC between HDL and other lipoproteins/cells, (2) the RCT and alternative pathways.

FC dynamically exchanges between HDL and LDL, maintaining equilibrium. This bidirectional exchange prevents a rapid rise in FC concentration while enabling its rapid replenishment. When FC levels increase, excess FC transfers to LDL, thereby avoiding uncontrolled CE production and exerting anti-atherosclerotic effects.

FC bidirectional exchange between HDL and other lipoproteins/cells confers several anti-atherosclerotic benefits:

1. Dynamic equilibrium of FC: The exchange between HDL and other lipoproteins/cells prevents excessive FC accumulation in HDL, avoiding overproduction of cholesteryl esters (CE).
2. Reduced concentration fluctuations: Dispersing FC across more particles stabilizes its concentration, enhancing HDL’s capacity for further FC uptake and expanding its storage and transport potential.
3. Diversified cholesterol sources: HDL can acquire FC from a broader range of sources, including VLDL, LDL, and peripheral cells.
4. Enhanced RCT: The process facilitates more efficient RCT and its alternative pathway, the key atheroprotective mechanism.
5. Prevention of CE accumulation: By maintaining FC equilibrium, excessive CE buildup is avoided.

Meanwhile, as CE is continuously synthesized in HDL, FC is replenished from LDL and other lipoproteins or cells, promoting RCT. HDL binds to SR-BI, facilitating cholesterol delivery to the liver.

Additionally, the decreased total lipids in VLDL and LDL promote CETP to transfer CE from HDL to ApoB-containing lipoproteins (e.g., chylomicrons, VLDL, and LDL) in exchange for TG to HDL. These lipoproteins are then recognized and processed by a broader range of receptors—including LDL receptors, LRP-1, VLDL receptors, and other hepatic receptors—thereby expanding cholesterol clearance pathways. This enhances hepatic uptake, processing, and excretion of cholesterol, increasing its metabolic clearance for cellular use and further promoting RCT. As a result, the ratio of FC to total lipids in large LDL and medium LDL exhibits a negative causal relationship with emergency coronary revascularization (for ACS) (lines 30,31 of Figure 3).

Notably, as VLDL particle size increases, the rate of FC transfer to HDL declines, thereby diminishing these beneficial effects. This is demonstrated by data from lines 23-29 of Figure 3 (FC to total lipids ratio in CM and extremely large VLDL, very large VLDL, large VLDL, medium VLDL, and small VLDL, very small VLDL, IDL, small LDL had no or positive causal relationship with emergency coronary revascularization [for ACS]).

This finding suggests that bidirectional free cholesterol (FC) diffusion plays an irreplaceable role in the dynamic supply of cholesterol sources required for reverse cholesterol transport (RCT). Loss of this mechanism (e.g., in VLDL subfractions) may trigger proatherogenic effects by disrupting cholesterol clearance.

CE continuously accumulates in HDL through ongoing synthesis. Through repeated bidirectional FC exchange among HDL, LDL, other lipoproteins/cells, a new dynamic equilibrium is maintained. However, if LDL/VLDL accepts CE repeatedly from HDL, excessive CE can be transferred to these lipoproteins, their total lipid content increases, leading to an overabundance of CE-rich particles.

Elevated levels of CE in lipoproteins disrupt ligand-receptor interactions—particularly with LDL receptors (LDLR)—resulting in prolonged circulatory retention and increased endothelial deposition of atherogenic lipids. This process promotes pro-atherosclerotic effects, especially when LDL carries higher total lipids. This demonstrates that total lipid levels in very large VLDL, large VLDL, medium VLDL, small VLDL, very small VLDL, IDL, large LDL, medium LDL, small LDL had a positive causal relationship with emergency coronary revascularization (for ACS).

### Comparison of Total Lipid Levels in HDL vs. VLDL/LDL Subfractions

Enhancing CETP activity exerts an anti-atherosclerotic effect, whereas inhibiting it promotes atherosclerosis.

Comparative analysis of MR results (Figure 3, lines 38-41 vs. lines 13-22) demonstrates that elevated total lipid levels in HDL — primarily CE — enhance CETP-mediated CE transfer from HDL to a broader spectrum of ApoB-containing lipoproteins (VLDL/LDL). Conversely, high total lipid levels in VLDL/LDL suppress this lipid redistribution. Mechanistically, HDL-enriched lipids promote RCT and its alternative pathways, thereby exerting anti-atherosclerotic effects, whereas excessive lipids in VLDL/LDL drive pro-atherogenic outcomes. These findings collectively suggest that augmenting CETP activity may attenuate atherosclerosis, while CETP inhibition could exacerbate disease progression.

### Comparison of FC Levels in HDL vs. VLDL/LDL Subfractions

HDL has the ability to synthesize CE and can disperse and transfer CE to a greater variety and quantity of lipoproteins, thereby promoting the RCT and its alternative pathways. Therefore, higher FC levels in HDL can play an anti-atherosclerotic role (lines 34,35,37 of Figure 3). However, VLDL/LDL do not have these functions (LCAT is also active on LDL, but the efficiency is relatively low), the higher FC levels in VLDL/LDL have a pro-atherosclerotic effect, as the MR results lines 1-12 of Figure 3. This shows that in the context where FC can freely diffuse bidirectionally and achieve a dynamic balance between different lipoproteins and cells, whether different lipoproteins and cells have the ability to synthesize CE (synthesize CE can also reduce cytotoxicity caused by the free diffusion of FC) and transport it out (cells can synthesize CE but cannot transport it out, only HDL can) determines their ability to have anti-atherosclerotic effects.

### FC-to-Total Lipids Ratio in VLDL/LDL vs. HDL Subfractions

A higher ratio of FC to total lipids in VLDL/LDL (Figure 3, lines 23–32) not only has the higher FC levels to promote the synthesis and transportation of CE, but also improves CE acceptance due to lower total lipid (mainly CE) levels, leading to anti-atherosclerotic effects. In contrast, a higher FC-to-total lipids ratio in HDL (Figure 3, lines 42–45) increases FC spontaneous diffusion, exacerbating pro-atherogenic effects, furthermore, the lower total lipid level does not exert a favorable protective effect to promote the synthesis of CE and transport it to more lipoproteins, thereby reducing the anti-atherosclerotic effect. This indicates that only when the spontaneous bidirectional diffusion of FC is closely connected to the favorable protective effect of effective CE synthesis and transportation to more lipoproteins can the anti-atherosclerotic effect be maximally exerted.

In conclusion, enhancing HDL’s capacity to accept cholesterol from peripheral tissues, synthesize CE, and redistribute them to a broader range of lipoproteins—thereby increasing receptor-mediated uptake and utilization by target cells — promotes anti-atherogenic effects.

## Conclusions

While spontaneously diffused FC from HDL promotes atherosclerosis, its bidirectional exchange with lipoproteins/cells—together with RCT and alternative pathways—can mitigate or reverse the pro-atherosclerotic effect. When this mechanism is impaired, the pro-atherosclerotic effect will increase. Alternatively, the excessive production of CE can also lose these benefits and re-generate the pro-atherosclerotic effect.

## Data Availability

All relevant data are within the manuscript and its Supporting Information files.

## Acknowledgements

We sincerely appreciate the IEU Open GWAS project and the FinnGen consortium for providing the data essential to our Mendelian randomization study. We are also deeply grateful to the researchers who shared these resources and to the participants whose contributions made this work possible.

## Footnote

### Nonstandard Abbreviations and Acronyms

ACS: acute coronary syndromes
ApoER2: apolipoprotein E receptor 2
AS: atherosclerosis
CE: cholesterol esters
CETP: Cholesteryl Ester Transfer Protein
CI: confidence interval
CM: chylomicrons
FC: free cholesterol
LCAT: Lecithin: Cholesterol Acyltransferase
LDLR: low-density lipoprotein receptor
MR: Mendelian Randomization
RCT: reverse cholesterol transport
SNPs: single nucleotide polymorphisms
SR-B1: Class B Scavenger Receptor B1
TG: triglycerides
VLDL: Very Low-Density Lipoproteins

