## Supplemental Table S2 for "Mendelian Randomization Study Reveals That Combined Spontaneous Free Cholesterol Diffusion and Reverse Cholesterol Transport Pathways Shift from Pro-Atherogenic to Anti-Atherogenic Effects"

### Supplemental Material

Table S2: The Mendelian randomization (MR) study results. Outcome: Emergency coronary revascularization (for ACS) (no controls excluded) || id:finn-b-I9_REVACS_EXNONE

| exposure | method | nsnp | b | se | pval | lo_ci | up_ci | or | or_lci95 | or_uci95 |
|---|---|---|---|---|---|---|---|---|---|---|
| Free cholesterol levels in chylomicrons and extremely large VLDL \|\| id:ebi-a-GCST90093044 | MR Egger | 54 | 0.18997471 9 | 0.13747301 | 0.17290878 6 | -0.07947238 | 0.45942181 8 | 1.20921902 7 | 0.92360353 | 1.58315836 7 |
| Free cholesterol levels in chylomicrons and extremely large VLDL \|\| id:eb | Weighted median | 54 | 0.32483234 3 | 0.09724641 6 | 0.00083687 5 | 0.13422936 7 | 0.51543531 9 | 1.38379862 3 | 1.14365510 6 | 1.67436722 6 |

| | | | | | | | | | | |
|---|---|---|---|---|---|---|---|---|---|---|
| i-a-GCST900093044 | | | | | | | | | | |
| Free cholesterol levels in chylomicrons and extremely large VLDL \|\| id:ebi-a-GCST90093044 | Inverse variance weighted | 54 | 0.3582222245 | 0.0740027 23 | 1.29E-06 | 0.2131769 09 | 0.5032675 81 | 1.4307835 69 | 1.2376035 75 | 1.6541174 12 |
| Free cholesterol levels in chylomicrons and extremely large VLDL \|\| id:ebi-a-GCST90093044 | Simple mode | 54 | 0.21508157 | 0.2063660 12 | 0.3020343 67 | -0.1893958 13 | 0.6195589 53 | 1.2399630 37 | 0.8274589 23 | 1.8581083 48 |

| | | | | | | | | | | |
|---|---|---|---|---|---|---|---|---|---|---|
| Free cholesterol levels in chylomicrons and extremely large VLDL \|\| id:ebi-a-GCST90093044 | Weighted mode | 54 | 0.304619561 | 0.114978652 | 0.010604549 | 0.079261404 | 0.529977718 | 1.356108988 | 1.082487251 | 1.698894454 |
| Free cholesterol levels in very large VLDL \|\| id:ebi-a-GCST90093020 | MR Egger | 56 | 0.073867631 | 0.193066008 | 0.703515997 | -0.304541745 | 0.452277007 | 1.076664279 | 0.737461242 | 1.571887312 |
| Free cholesterol levels in very large VLDL | Weighted median | 56 | 0.495601797 | 0.079713363 | 5.06E-10 | 0.339363607 | 0.651839988 | 1.641485784 | 1.404053776 | 1.919068646 |

| | | | | | | | | | | |
|---|---|---|---|---|---|---|---|---|---|---|
| \|\| id:ebi-a-GCST90093020 | | | | | | | | | | |
| Free cholesterol levels in very large VLDL \|\| id:ebi-a-GCST90093020 | Inverse variance weighted | 56 | 0.336691492 | 0.121605133 | 0.005627566 | 0.098345431 | 0.575037553 | 1.400306991 | 1.103343848 | 1.777197265 |
| Free cholesterol levels in very large VLDL \|\| id:ebi-a-GCST90093020 | Simple mode | 56 | 0.197397391 | 0.169558063 | 0.249373261 | -0.134936412 | 0.529731195 | 1.218228058 | 0.873771471 | 1.698475688 |
| Free cholesterol levels in very large | Weighted mode | 56 | 0.440575529 | 0.07552429 | 2.98E-07 | 0.292547921 | 0.588603138 | 1.553601104 | 1.339836941 | 1.801470251 |

| | | | | | | | | | | |
|---|---|---|---|---|---|---|---|---|---|---|
| VLDL \|\| id:ebi-a-GCST90093020 | | | | | | | | | | |
| Free cholesterol levels in large VLDL \|\| id:ebi-a-GCST90092872 | MR Egger | 51 | 0.333189053 | 0.118248589 | 0.00695534 | 0.101421819 | 0.564956287 | 1.39541108 | 1.106743388 | 1.759370873 |
| Free cholesterol levels in large VLDL \|\| id:ebi-a-GCST90092872 | Weighted median | 51 | 0.490445822 | 0.081756988 | 1.99E-09 | 0.330202125 | 0.650689518 | 1.633044104 | 1.391249307 | 1.916862084 |
| Free cholesterol levels in large VLDL \|\| | Inverse variance weighted | 51 | 0.470925725 | 0.070542126 | 2.46E-11 | 0.332663159 | 0.609188292 | 1.601476034 | 1.394677435 | 1.838938111 |

| | | | | | | | | | | |
|---|---|---|---|---|---|---|---|---|---|---|
| id:ebi-a-GCST90092872 | | | | | | | | | | |
| Free cholesterol levels in large VLDL \|\| id:ebi-a-GCST90092872 | Simple mode | 51 | 0.182165206 | 0.158722307 | 0.256555293 | -0.128930515 | 0.493260927 | 1.199812393 | 0.879035043 | 1.637647772 |
| Free cholesterol levels in large VLDL \|\| id:ebi-a-GCST90092872 | Weighted mode | 51 | 0.435990259 | 0.07586831 | 5.40E-07 | 0.287288371 | 0.584692147 | 1.54649373 | 1.332808502 | 1.794438477 |
| Free cholesterol levels in medium VLDL \|\| id:eb | MR Egger | 61 | 0.918943864 | 0.190221822 | 1.00E-05 | 0.546109092 | 1.291778635 | 2.506641637 | 1.726522193 | 3.639253708 |

| | | | | | | | | | | |
|---|---|---|---|---|---|---|---|---|---|---|
| i-a-GCST900092920 | | | | | | | | | | |
| Free cholesterol levels in medium VLDL \|\| id:ebi-a-GCST900092920 | Weighted median | 61 | 0.755510664 | 0.09233678 | 2.79E-16 | 0.574530576 | 0.936490752 | 2.128698295 | 1.776296495 | 2.551013553 |
| Free cholesterol levels in medium VLDL \|\| id:ebi-a-GCST900092920 | Inverse variance weighted | 61 | 0.682998791 | 0.104997175 | 7.77E-11 | 0.477204329 | 0.888793253 | 1.979805863 | 1.611562699 | 2.432192839 |
| Free cholesterol levels in medium VLDL \|\| | Simple mode | 61 | 0.655609294 | 0.192765785 | 0.001199508 | 0.277788357 | 1.033430232 | 1.926315853 | 1.320206755 | 2.810690639 |

| | | | | | | | | | | |
|---|---|---|---|---|---|---|---|---|---|---|
| id:ebi-a-GCST90092920 | | | | | | | | | | |
| Free cholesterol levels in medium VLDL \|\| id:ebi-a-GCST90092920 | Weighted mode | 61 | 1.026764666 | 0.0929957244 | 4.40E-16 | 0.844493048 | 1.209036285 | 2.792018096 | 2.32679794 | 3.350254404 |
| Free cholesterol levels in small VLDL \|\| id:ebi-a-GCST90092972 | MR Egger | 56 | 0.903500384 | 0.190304802 | 1.56E-05 | 0.530502972 | 1.276497796 | 2.468227752 | 1.699787038 | 3.58406559 |
| Free cholesterol levels in small VLDL \|\| id:eb | Weighted median | 56 | 0.994331144 | 0.085593016 | 3.38E-31 | 0.826568832 | 1.162093456 | 2.702915876 | 2.285463463 | 3.196618257 |

| | | | | | | | | | | |
|---|---|---|---|---|---|---|---|---|---|---|
| i-a-GCST900929 72 | | | | | | | | | | |
| Free cholesterol levels in small VLDL \|\| id:ebi-a-GCST900929 72 | Inverse variance weighted | 56 | 0.7215463 49 | 0.1034682 31 | 3.09E-12 | 0.5187486 16 | 0.9243440 82 | 2.0576125 4 | 1.6799241 04 | 2.5202146 65 |
| Free cholesterol levels in small VLDL \|\| id:ebi-a-GCST900929 72 | Simple mode | 56 | 1.0262025 43 | 0.1699660 52 | 1.40E-07 | 0.6930690 82 | 1.3593360 04 | 2.7904490 79 | 1.9998438 09 | 3.8936071 05 |
| Free cholesterol levels in small VLDL \|\| id:ebi-a-GCST9 | Weighted mode | 56 | 1.0606551 16 | 0.0938130 74 | 5.73E-16 | 0.8767814 91 | 1.2445287 4 | 2.8882625 15 | 2.4031526 78 | 3.4712985 29 |

| | | | | | | | | | | |
|---|---|---|---|---|---|---|---|---|---|---|
| 0092972 | | | | | | | | | | |
| Free cholesterol levels in very small VLDL \|\| id:ebi-a-GCST90093032 | MR Egger | 59 | 0.4740494244 | 0.1378076099 | 0.0010949277 | 0.20394651 | 0.7441523338 | 1.6064863844 | 1.2262325611 | 2.1046566411 |
| Free cholesterol levels in very small VLDL \|\| id:ebi-a-GCST90093032 | Weighted median | 59 | 0.2611541922 | 0.0937931044 | 0.0053632722 | 0.0773197099 | 0.4449886755 | 1.2984278588 | 1.0803874311 | 1.5604725244 |
| Free cholesterol levels in very small VLDL \|\| id:ebi-a- | Inverse variance weighted | 59 | 0.5396914055 | 0.0826407344 | 6.55E-11 | 0.3777155666 | 0.7016672444 | 1.7154773933 | 1.4589479099 | 2.0171129266 |

| | | | | | | | | | | |
|---|---|---|---|---|---|---|---|---|---|---|
| GCST900930 32 | | | | | | | | | | |
| Free cholesterol levels in very small VLDL \|\| id:ebi-a-GCST900930 32 | Simple mode | 59 | 0.968449443 | 0.292181424 | 0.001586106 | 0.395773851 | 1.541125035 | 2.633857345 | 1.485533327 | 4.669841049 |
| Free cholesterol levels in very small VLDL \|\| id:ebi-a-GCST900930 32 | Weighted mode | 59 | 0.190695931 | 0.078201222 | 0.017830187 | 0.037421537 | 0.343970326 | 1.210091445 | 1.038130539 | 1.410536779 |
| Free cholesterol levels in VLDL \|\| id:ebi-a-GCST9 | MR Egger | 60 | 0.357752736 | 0.172980064 | 0.043092825 | 0.018711811 | 0.696793661 | 1.430111961 | 1.018887974 | 2.007306273 |

| | | | | | | | | | | |
|---|---|---|---|---|---|---|---|---|---|---|
| 0092998 | | | | | | | | | | |
| Free cholesterol levels in VLDL \|\| id:ebi-a-GCST90092998 | Weighted median | 60 | 0.5692401066 | 0.0794485788 | 7.79E-13 | 0.4135208933 | 0.7249593199 | 1.7669238666 | 1.51213248 | 2.0646471077 |
| Free cholesterol levels in VLDL \|\| id:ebi-a-GCST90092998 | Inverse variance weighted | 60 | 0.52710839 | 0.0942988099 | 2.27E-08 | 0.3422827224 | 0.7119340566 | 1.6940267555 | 1.4081583622 | 2.0379289199 |
| Free cholesterol levels in VLDL \|\| id:ebi-a-GCST90092998 | Simple mode | 60 | 0.3427660455 | 0.16371407 | 0.0405945677 | 0.0218864688 | 0.6636456222 | 1.4088391199 | 1.0221277344 | 1.9418587299 |
| Free cholesterol stero | Weighted | 60 | 0.5340001599 | 0.0955723336 | 6.19E-07 | 0.3466783811 | 0.7213219377 | 1.7057419188 | 1.4143617677 | 2.0571508377 |

| | | | | | | | | | | |
|---|---|---|---|---|---|---|---|---|---|---|
| l levels in VLDL \|\| id:ebi-a-GCST90092998 | mode | | | | | | | | | |
| Free cholesterol levels in IDL \|\| id:ebi-a-GCST90092835 | MR Egger | 57 | 0.852654913 | 0.222837594 | 0.000334617 | 0.415893229 | 1.289416597 | 2.345866664 | 1.515724025 | 3.630667796 |
| Free cholesterol levels in IDL \|\| id:ebi-a-GCST90092835 | Weighted median | 57 | 0.606044849 | 0.102665491 | 3.57E-09 | 0.404820487 | 0.80726921 | 1.83316659 | 1.49903338 | 2.241777796 |
| Free cholesterol levels in IDL \|\| | Inverse variance wei | 57 | 0.579522722 | 0.123097582 | 2.50E-06 | 0.338251462 | 0.820793983 | 1.785186198 | 1.402493133 | 2.272303292 |

| | | | | | | | | | | |
|---|---|---|---|---|---|---|---|---|---|---|
| id:ebi-a-GCST900092835 | ghted | | | | | | | | | |
| Free cholesterol levels in IDL \|\| id:ebi-a-GCST900092835 | Simple mode | 57 | 0.587375998 | 0.1885102 | 0.002891379 | 0.217896006 | 0.956855991 | 1.799260952 | 1.243457748 | 2.603498171 |
| Free cholesterol levels in IDL \|\| id:ebi-a-GCST900092835 | Weighted mode | 57 | 0.773099716 | 0.131452794 | 2.37E-07 | 0.51545224 | 1.030747192 | 2.166471303 | 1.674395559 | 2.803159552 |
| Free cholesterol levels in large LDL \|\| id:ebi-a-GCST9 | MR Egger | 3 | 1.975358313 | 2.216476229 | 0.536578609 | -2.368935096 | 6.319651723 | 7.209202344 | 0.093580327 | 555.3795327 |

| | | | | | | | | | | |
|---|---|---|---|---|---|---|---|---|---|---|
| 0092860 | | | | | | | | | | |
| Free cholesterol levels in large LDL \|\| id:ebi-a-GCST90092860 | Weighted median | 3 | 0.9571426 68 | 0.2952410 56 | 0.0011873 47 | 0.3784701 99 | 1.5358151 37 | 2.60424464 | 1.4600492 95 | 4.6451103 87 |
| Free cholesterol levels in large LDL \|\| id:ebi-a-GCST90092860 | Inverse variance weighted | 3 | 0.9830242 01 | 0.2550037 59 | 0.0001157 58 | 0.4832168 34 | 1.4828315 68 | 2.6725262 89 | 1.6212814 15 | 4.4054022 32 |
| Free cholesterol levels in large LDL \|\| id:ebi-a-GCST90092860 | Simple mode | 3 | 0.8625072 92 | 0.3554476 08 | 0.1360247 91 | 0.16582998 | 1.5591846 04 | 2.3690932 62 | 1.1803723 98 | 4.7549425 05 |

| | | | | | | | | | | |
|---|---|---|---|---|---|---|---|---|---|---|
| Free cholesterol levels in large LDL \|\| id:ebi-a-GCST90092860 | Weighted mode | 3 | 0.882110008 | 0.344570834 | 0.124680718 | 0.206751174 | 1.557468843 | 2.415992096 | 1.229676558 | 4.746791152 |
| Free cholesterol levels in medium LDL \|\| id:ebi-a-GCST90092908 | MR Egger | 48 | 0.900975057 | 0.157318995 | 7.41E-07 | 0.592629827 | 1.209320287 | 2.462002534 | 1.808738837 | 3.35120602 |
| Free cholesterol levels in medium LDL \|\| id:ebi-a-GCST90092908 | Weighted median | 48 | 0.94703382 | 0.101866192 | 1.45E-20 | 0.747376084 | 1.146691556 | 2.578051342 | 2.111452468 | 3.14776147 |

| | | | | | | | | | | |
|---|---|---|---|---|---|---|---|---|---|---|
| Free cholesterol levels in medium LDL \|\| id:ebi-a-GCST90092908 | Inverse variance weighted | 48 | 0.722873033 | 0.092724324 | 6.39E-15 | 0.541133358 | 0.904612708 | 2.060344152 | 1.717952815 | 2.470974748 |
| Free cholesterol levels in medium LDL \|\| id:ebi-a-GCST90092908 | Simple mode | 48 | 0.627637277 | 0.219723813 | 0.006360315 | 0.196978603 | 1.058295951 | 1.873179542 | 1.217717985 | 2.881456659 |
| Free cholesterol levels in medium LDL \|\| id:ebi-a-GCST90092908 | Weighted mode | 48 | 0.949618171 | 0.118922609 | 2.68E-10 | 0.716529858 | 1.182706484 | 2.584722548 | 2.047316391 | 3.263194043 |

| | | | | | | | | | | |
|---|---|---|---|---|---|---|---|---|---|---|
| Free cholesterol levels in small LDL \|\| id:ebi-a-GCST90092960 | MR Egger | 48 | 0.791810631 | 0.156152048 | 6.93E-06 | 0.485752617 | 1.097868646 | 2.207389579 | 1.62539785 | 2.997769901 |
| Free cholesterol levels in small LDL \|\| id:ebi-a-GCST90092960 | Weighted median | 48 | 0.793453285 | 0.099822837 | 1.89E-15 | 0.597800525 | 0.989106044 | 2.211018534 | 1.8181155 | 2.688829703 |
| Free cholesterol levels in small LDL \|\| id:ebi-a-GCST90092960 | Inverse variance weighted | 48 | 0.626364526 | 0.093808354 | 2.44E-11 | 0.442500153 | 0.810228899 | 1.870796968 | 1.556594081 | 2.24842259 |

| | | | | | | | | | | |
|---|---|---|---|---|---|---|---|---|---|---|
| Free cholesterol levels in small LDL \|\| id:ebi-a-GCST90092960 | Simple mode | 48 | 0.5629077155 | 0.2143432566 | 0.0116184088 | 0.1427949333 | 0.9830204977 | 1.7557703655 | 1.1534932344 | 2.67251639 |
| Free cholesterol levels in small LDL \|\| id:ebi-a-GCST90092960 | Weighted mode | 48 | 0.8116893488 | 0.1068953088 | 1.04E-09 | 0.6021745444 | 1.0212041511 | 2.2517086944 | 1.8260853899 | 2.7765361211 |
| Free cholesterol levels in LDL \|\| id:ebi-a-GCST90092885 | MR Egger | 47 | 0.8380221112 | 0.1653774244 | 7.35E-06 | 0.51388236 | 1.1621618633 | 2.3117899899 | 1.6717690222 | 3.196836935 |
| Free cholesterol stero | Weighted | 47 | 0.8722686744 | 0.0993562911 | 1.65E-18 | 0.6775303445 | 1.0670070044 | 2.3923321244 | 1.96900895 | 2.906666825 |

| | | | | | | | | | | |
|---|---|---|---|---|---|---|---|---|---|---|
| l levels in LDL \|\| id:ebi-a-GCST90092885 | median | | | | | | | | | |
| Free cholesterol levels in LDL \|\| id:ebi-a-GCST90092885 | Inverse variance weighted | 47 | 0.704542442 | 0.093848905 | 6.04E-14 | 0.520598588 | 0.888486295 | 2.022920869 | 1.683034793 | 2.431446372 |
| Free cholesterol levels in LDL \|\| id:ebi-a-GCST90092885 | Simple mode | 47 | 0.73557752 | 0.212535438 | 0.00117275 | 0.319008062 | 1.152146978 | 2.086686747 | 1.375762416 | 3.164980764 |
| Free cholesterol levels in LDL \|\| | Weighted mode | 47 | 0.943967099 | 0.111094202 | 5.53E-11 | 0.726222463 | 1.161711734 | 2.570157289 | 2.067256701 | 3.195398271 |

| | | | | | | | | | | |
|---|---|---|---|---|---|---|---|---|---|---|
| id:ebi-a-GCST900092885 | | | | | | | | | | |
| Total lipid levels in chylomicrons and extremely large VLDL \|\| id:ebi-a-GCST90093046 | MR Egger | 56 | -0.181400929 | 0.186760046 | 0.335727293 | -0.547450619 | 0.184648761 | 0.834100876 | 0.578422552 | 1.202795897 |
| Total lipid levels in chylomicrons and extremely large VLDL \|\| id:ebi-a-GCST90093046 | Weighted median | 56 | 0.342928889 | 0.092987082 | 0.000226095 | 0.160674209 | 0.52518357 | 1.409068559 | 1.174302329 | 1.690769195 |
| Total lipid level | Inverse | 56 | 0.095663557 | 0.122175616 | 0.433626871 | -0.14 | 0.335127764 | 1.100388784 | 0.866060381 | 1.398119003 |

| | | | | | | | | | | |
|---|---|---|---|---|---|---|---|---|---|---|
| s in chylomicrons and extremely large VLDL \|\| id:ebi-a-GCST90093046 | variance weighted | | | | | 3800649 | | | | |
| Total lipid levels in chylomicrons and extremely large VLDL \|\| id:ebi-a-GCST90093046 | Simple mode | 56 | 0.176246998 | 0.202789973 | 0.388563395 | -0.221221348 | 0.573715345 | 1.192732625 | 0.801539241 | 1.774848993 |
| Total lipid levels in chylomicrons and extremely large | Weighted mode | 56 | 0.466357926 | 0.090641629 | 3.69E-06 | 0.288700333 | 0.644015519 | 1.594177495 | 1.334691706 | 1.904111545 |

| | | | | | | | | | | |
|---|---|---|---|---|---|---|---|---|---|---|
| VLDL \|\| id:ebi-a-GCST90093046 | | | | | | | | | | |
| Total lipid levels in very large VLDL \|\| id:ebi-a-GCST90093022 | MR Egger | 59 | 0.081302446 | 0.179701455 | 0.652676029 | -0.270912406 | 0.433517298 | 1.084698909 | 0.7626833 | 1.542674036 |
| Total lipid levels in very large VLDL \|\| id:ebi-a-GCST90093022 | Weighted median | 59 | 0.478404685 | 0.077249812 | 5.90E-10 | 0.326995052 | 0.629814317 | 1.613498309 | 1.386794616 | 1.877261971 |
| Total lipid levels in very large VLDL \|\| id:ebi-a-GCST9 | Inverse variance weighted | 59 | 0.271747727 | 0.110638973 | 0.014042812 | 0.054895339 | 0.488600114 | 1.312255912 | 1.056430042 | 1.630032762 |

| | | | | | | | | | | |
|---|---|---|---|---|---|---|---|---|---|---|
| 0093022 | | | | | | | | | | |
| Total lipid levels in very large VLDL \|\| id:ebi-a-GCST90093022 | Simple mode | 59 | 0.116259862 | 0.177503231 | 0.515074298 | -0.23164647 | 0.464166194 | 1.123287734 | 0.793226503 | 1.590687311 |
| Total lipid levels in very large VLDL \|\| id:ebi-a-GCST90093022 | Weighted mode | 59 | 0.470091261 | 0.082779528 | 4.58E-07 | 0.307843386 | 0.632339136 | 1.600140217 | 1.360487901 | 1.882007706 |
| Total lipid levels in large VLDL \|\| id:ebi-a-GCST90092874 | MR Egger | 60 | 0.112398931 | 0.18432282 | 0.544380052 | -0.248873795 | 0.473671658 | 1.118959159 | 0.779678366 | 1.605879622 |
| Total lipid levels in | Weighted | 60 | 0.486380093 | 0.078745134 | 6.55E-10 | 0.332039629 | 0.640720556 | 1.626418068 | 1.393808083 | 1.897847893 |

| | | | | | | | | | | |
|---|---|---|---|---|---|---|---|---|---|---|
| large VLDL \|\| id:ebi-a-GCST90092874 | median | | | | | | | | | |
| Total lipid levels in large VLDL \|\| id:ebi-a-GCST90092874 | Inverse variance weighted | 60 | 0.334403822 | 0.115296333 | 0.003727039 | 0.108423011 | 0.560384634 | 1.397107213 | 1.114519099 | 1.751345998 |
| Total lipid levels in large VLDL \|\| id:ebi-a-GCST90092874 | Simple mode | 60 | 0.237787743 | 0.188759824 | 0.212724689 | -0.132181512 | 0.607756998 | 1.268439928 | 0.876181943 | 1.836307933 |
| Total lipid levels in large VLDL \|\| id:ebi-a-GCST90092874 | Weighted mode | 60 | 0.460823844 | 0.074133434 | 5.65E-08 | 0.315522313 | 0.606125376 | 1.585379553 | 1.370975202 | 1.833314216 |

| | | | | | | | | | | |
|---|---|---|---|---|---|---|---|---|---|---|
| Total lipid levels in medium VLDL \|\| id:ebi-a-GCST90092922 | MR Egger | 57 | 0.4240631333 | 0.1758417833 | 0.0192475199 | 0.0794132339 | 0.7687130277 | 1.5281580688 | 1.0826516233 | 2.15698848 |
| Total lipid levels in medium VLDL \|\| id:ebi-a-GCST90092922 | Weighted median | 57 | 0.6073914977 | 0.08847294 | 6.64E-12 | 0.4339845334 | 0.78079846 | 1.8356368844 | 1.5433949988 | 2.18321478 |
| Total lipid levels in medium VLDL \|\| id:ebi-a-GCST90092922 | Inverse variance weighted | 57 | 0.5406908199 | 0.0990631377 | 4.81E-08 | 0.3465270711 | 0.7348545677 | 1.7171927222 | 1.4141477766 | 2.0851787177 |
| Total lipid levels in mediu | Simple mode | 57 | 0.3671878922 | 0.1877965022 | 0.0555517533 | -0.0008932511 | 0.7352690355 | 1.4436691488 | 0.9991071488 | 2.0860431377 |

| | | | | | | | | | | |
|---|---|---|---|---|---|---|---|---|---|---|
| m VLDL \|\| id:ebi-a-GCST90092922 | | | | | | | | | | |
| Total lipid levels in medium VLDL \|\| id:ebi-a-GCST90092922 | Weighted mode | 57 | 0.559997494 | 0.108054863 | 3.10E-06 | 0.348209963 | 0.771785026 | 1.750668114 | 1.416529637 | 2.163624936 |
| Total lipid levels in small VLDL \|\| id:ebi-a-GCST90092974 | MR Egger | 64 | 0.347665677 | 0.147363528 | 0.021476888 | 0.058833163 | 0.636498192 | 1.415758851 | 1.060598278 | 1.889851383 |
| Total lipid levels in small VLDL \|\| id:ebi-a-GCST9 | Weighted median | 64 | 0.517211057 | 0.076355363 | 1.25E-11 | 0.367554545 | 0.666867569 | 1.677343106 | 1.44419857 | 1.948125384 |

| | | | | | | | | | | |
|---|---|---|---|---|---|---|---|---|---|---|
| 0092974 | | | | | | | | | | |
| Total lipid levels in small VLDL \|\| id:ebi-a-GCST90092974 | Inverse variance weighted | 64 | 0.4686648535 | 0.0819851999 | 1.09E-08 | 0.3079575446 | 0.6293395244 | 1.5978333318 | 1.3606432223 | 1.8763708733 |
| Total lipid levels in small VLDL \|\| id:ebi-a-GCST90092974 | Simple mode | 64 | 0.28829497 | 0.1520219544 | 0.0624940966 | -0.0096680599 | 0.5862579999 | 1.33415078 | 0.9903785266 | 1.7972505033 |
| Total lipid levels in small VLDL \|\| id:ebi-a-GCST90092974 | Weighted mode | 64 | 0.4751329144 | 0.0854209771 | 5.80E-07 | 0.3077078112 | 0.6425580177 | 1.60822794 | 1.3603034666 | 1.90133832 |
| Total lipid levels in very small | MR Egger | 65 | 0.4809396667 | 0.1444982833 | 0.0014624877 | 0.1977230322 | 0.7641563011 | 1.6175936877 | 1.2186248277 | 2.1471820355 |

| | | | | | | | | | | |
|---|---|---|---|---|---|---|---|---|---|---|
| VLDL \|\| id:ebi-a-GCST90093034 | | | | | | | | | | |
| Total lipid levels in very small VLDL \|\| id:ebi-a-GCST90093034 | Weighted median | 65 | 0.24232162 | 0.090175776 | 0.007205074 | 0.0655771 | 0.419066141 | 1.274203937 | 1.06777506 | 1.520540921 |
| Total lipid levels in very small VLDL \|\| id:ebi-a-GCST90093034 | Inverse variance weighted | 65 | 0.478549462 | 0.087755266 | 4.95E-08 | 0.30654914 | 0.650549784 | 1.613731924 | 1.358728234 | 1.916594252 |
| Total lipid levels in very small VLDL \|\| id:ebi-a-GCST9 | Simple mode | 65 | 0.679542888 | 0.323751843 | 0.039770418 | 0.044989276 | 1.3140965 | 1.972975655 | 1.046016642 | 3.721387193 |

| | | | | | | | | | | |
|---|---|---|---|---|---|---|---|---|---|---|
| 0093034 | | | | | | | | | | |
| Total lipid levels in very small VLDL \|\| id:ebi-a-GCST90093034 | Weighted mode | 65 | 0.183820193 | 0.071661024 | 0.012668208 | 0.043364585 | 0.324275801 | 1.201799712 | 1.044318569 | 1.383028696 |
| Total lipid levels in IDL \|\| id:ebi-a-GCST90092837 | MR Egger | 54 | 0.876657324 | 0.173035133 | 5.46E-06 | 0.537508463 | 1.215806185 | 2.402854305 | 1.71173669 | 3.373012242 |
| Total lipid levels in IDL \|\| id:ebi-a-GCST90092837 | Weighted median | 54 | 0.524577397 | 0.10493638 | 5.76E-07 | 0.318902092 | 0.730252703 | 1.689744607 | 1.375616635 | 2.075605052 |
| Total lipid levels in IDL \|\| id:eb | Inverse variance wei | 54 | 0.610126044 | 0.098897456 | 6.86E-10 | 0.416287031 | 0.803965057 | 1.840663389 | 1.516321038 | 2.234382843 |

| | | | | | | | | | | |
|---|---|---|---|---|---|---|---|---|---|---|
| i-a-GCST900092837 | ghted | | | | | | | | | |
| Total lipid levels in IDL \|\| id:ebi-a-GCST90092837 | Simple mode | 54 | 0.5473853233 | 0.2247269977 | 0.0182553555 | 0.10692041 | 0.9878502237 | 1.7287270411 | 1.11284568 | 2.6854551699 |
| Total lipid levels in IDL \|\| id:ebi-a-GCST90092837 | Weighted mode | 54 | 0.57479049 | 0.2027563222 | 0.0064747888 | 0.1773880999 | 0.97219288 | 1.7767582399 | 1.19409443 | 2.6437355033 |
| Total lipid levels in large LDL \|\| id:ebi-a-GCST90092862 | MR Egger | 44 | 0.9565169449 | 0.1397804422 | 2.44E-08 | 0.6825472833 | 1.2304866155 | 2.6026156244 | 1.9789121666 | 3.4228947622 |
| Total lipid levels in large LDL | Weighted median | 44 | 0.9691370933 | 0.0963821811 | 8.72E-24 | 0.7802280199 | 1.1580461677 | 2.63566914 | 2.1819697399 | 3.1837067644 |

| | | | | | | | | | | |
|---|---|---|---|---|---|---|---|---|---|---|
| \|\| id:ebi-a-GCST90092862 | | | | | | | | | | |
| Total lipid levels in large LDL \|\| id:ebi-a-GCST90092862 | Inverse variance weighted | 44 | 0.833154761 | 0.077558517 | 6.44E-27 | 0.681140067 | 0.985169456 | 2.300565038 | 1.976129369 | 2.678265694 |
| Total lipid levels in large LDL \|\| id:ebi-a-GCST90092862 | Simple mode | 44 | 1.002422887 | 0.223670275 | 5.41E-05 | 0.564029148 | 1.440816627 | 2.724875904 | 1.757740448 | 4.224143958 |
| Total lipid levels in large LDL \|\| id:ebi-a-GCST90092862 | Weighted mode | 44 | 1.057268487 | 0.119147583 | 2.85E-11 | 0.823739224 | 1.290797749 | 2.878497587 | 2.279005638 | 3.635685764 |

| | | | | | | | | | | |
|---|---|---|---|---|---|---|---|---|---|---|
| Total lipid levels in medium LDL \|\| id:ebi-a-GCST90092910 | MR Egger | 46 | 0.8830773344 | 0.20823725 | 0.00011261 | 0.474932323 | 1.291222344 | 2.418330276 | 1.607905375 | 3.637229787 |
| Total lipid levels in medium LDL \|\| id:ebi-a-GCST90092910 | Weighted median | 46 | 0.935901933 | 0.101148139 | 2.19E-20 | 0.737651582 | 1.134152285 | 2.549511911 | 2.091019156 | 3.108537273 |
| Total lipid levels in medium LDL \|\| id:ebi-a-GCST90092910 | Inverse variance weighted | 46 | 0.727641155 | 0.109081286 | 2.55E-11 | 0.513841835 | 0.941440475 | 2.070191582 | 1.671701274 | 2.563671665 |
| Total lipid levels in medium LDL \|\| id:eb | Simple mode | 46 | 0.742935903 | 0.230798692 | 0.002388629 | 0.290570466 | 1.19530134 | 2.10209802 | 1.337190092 | 3.304553415 |

| | | | | | | | | | | |
|---|---|---|---|---|---|---|---|---|---|---|
| i-a-GCST900929 10 | | | | | | | | | | |
| Total lipid levels in medium LDL \|\| id:ebi-a-GCST90092910 | Weighted mode | 46 | 1.053643041 | 0.1074207033 | 9.49E-13 | 0.8430984633 | 1.264187619 | 2.8680806 45 | 2.3235552 84 | 3.5402155 65 |
| Total lipid levels in small LDL \|\| id:ebi-a-GCST90092962 | MR Egger | 48 | 0.81980028 | 0.1866076 79 | 6.51E-05 | 0.4540492 29 | 1.18555133 | 2.2700464 18 | 1.5746755 16 | 3.2724905 46 |
| Total lipid levels in small LDL \|\| id:ebi-a-GCST90092962 | Weighted median | 48 | 0.9250576 44 | 0.0969328 14 | 1.38E-21 | 0.7350693 28 | 1.11504596 | 2.5220136 36 | 2.08562658 | 3.0497083 42 |
| Total lipid levels in | Inverse var | 48 | 0.7681935 54 | 0.10514319 | 2.75E-13 | 0.5621129 03 | 0.9742742 06 | 2.1558682 75 | 1.7543754 11 | 2.6492437 08 |

| | | | | | | | | | | |
|---|---|---|---|---|---|---|---|---|---|---|
| small LDL \|\| id:ebi-a-GCST90092962 | iance weighted | | | | | | | | | |
| Total lipid levels in small LDL \|\| id:ebi-a-GCST90092962 | Simple mode | 48 | 0.637376831 | 0.205089667 | 0.003195289 | 0.235401083 | 1.039352579 | 1.891512608 | 1.265416203 | 2.827385912 |
| Total lipid levels in small LDL \|\| id:ebi-a-GCST90092962 | Weighted mode | 48 | 0.965241109 | 0.112407217 | 3.44E-11 | 0.744922964 | 1.185559253 | 2.625420592 | 2.10627917 | 3.272516475 |
| Free cholesterol to total lipids ratio in chylomicrons | MR Egger | 15 | 0.298081853 | 0.439364317 | 0.509390704 | -0.563072208 | 1.159235914 | 1.347272062 | 0.569456884 | 3.187496825 |

| | | | | | | | | | | |
|---|---|---|---|---|---|---|---|---|---|---|
| and extremely large VLDL \|\| id:ebi-a-GCST90093045 | | | | | | | | | | |
| Free cholesterol to total lipids ratio in chylomicrons and extremely large VLDL \|\| id:ebi-a-GCST90093045 | Weighted median | 15 | 0.130405711 | 0.119688643 | 0.275915287 | -0.10418403 | 0.364995451 | 1.139290512 | 0.90105946 | 1.440507456 |
| Free cholesterol to total lipids ratio in chylo | Inverse variance weighted | 15 | 0.023384003 | 0.229923314 | 0.918992033 | -0.427265692 | 0.474033699 | 1.023659553 | 0.652290221 | 1.606461122 |

| | | | | | | | | | | |
|---|---|---|---|---|---|---|---|---|---|---|
| microns and extremely large VLDL \|\| id:ebi-a-GCST90093045 | | | | | | | | | | |
| Free cholesterol to total lipids ratio in chylomicrons and extremely large VLDL \|\| id:ebi-a-GCST90093045 | Simple mode | 15 | 0.466107851 | 0.435260157 | 0.302341928 | -0.387002057 | 1.319217759 | 1.593778881 | 0.679089698 | 3.740494265 |
| Free cholesterol to total lipids ratio | Weighted mode | 15 | 0.181970965 | 0.11176934 | 0.125792723 | -0.037096941 | 0.401038871 | 1.199579363 | 0.96358272 | 1.493375316 |

| | | | | | | | | | | |
|---|---|---|---|---|---|---|---|---|---|---|
| in chylomicrons and extremely large VLDL \|\| id:ebi-a-GCST90093045 | | | | | | | | | | |
| Free cholesterol to total lipids ratio in very large VLDL \|\| id:ebi-a-GCST90093021 | MR Egger | 45 | 0.375151507 | 0.226976962 | 0.105652188 | -0.06972334 | 0.820026353 | 1.455211872 | 0.932651812 | 2.270559673 |
| Free cholesterol to total lipids ratio in very large | Weighted median | 45 | 0.166296153 | 0.098711768 | 0.092053432 | -0.027178912 | 0.359771218 | 1.180922784 | 0.973187111 | 1.433001532 |

| | | | | | | | | | | |
|---|---|---|---|---|---|---|---|---|---|---|
| VLDL \|\| id:ebi-a-GCST90093021 | | | | | | | | | | |
| Free cholesterol to total lipids ratio in very large VLDL \|\| id:ebi-a-GCST90093021 | Inverse variance weighted | 45 | 0.1979120688 | 0.1094477144 | 0.0705629977 | -0.0166054552 | 0.4124295888 | 1.2188552133 | 0.9835316588 | 1.5104831822 |
| Free cholesterol to total lipids ratio in very large VLDL \|\| id:ebi-a-GCST90093021 | Simple mode | 45 | 0.1466205844 | 0.2096934933 | 0.4880984011 | -0.2643786633 | 0.55761983 | 1.1579145488 | 0.7676827922 | 1.7465105588 |

| | | | | | | | | | | |
|---|---|---|---|---|---|---|---|---|---|---|
| Free cholesterol to total lipids ratio in very large VLDL \|\| id:ebi-a-GCST90093021 | Weighted mode | 45 | 0.215224606 | 0.098518775 | 0.03428899 | 0.022127807 | 0.408321405 | 1.240140409 | 1.022374443 | 1.50429057 |
| Free cholesterol to total lipids ratio in large VLDL \|\| id:ebi-a-GCST90092873 | MR Egger | 46 | 0.475131135 | 0.231313557 | 0.045940946 | 0.021756563 | 0.928505707 | 1.608225079 | 1.021994963 | 2.530724707 |
| Free cholesterol to total lipids ratio in | Weighted median | 46 | 0.206358293 | 0.079317628 | 0.009277107 | 0.050895743 | 0.361820843 | 1.229193537 | 1.052213187 | 1.43594166 |

| | | | | | | | | | | |
|---|---|---|---|---|---|---|---|---|---|---|
| large VLDL \|\| id:ebi-a-GCST90092873 | | | | | | | | | | |
| Free cholesterol to total lipids ratio in large VLDL \|\| id:ebi-a-GCST90092873 | Inverse variance weighted | 46 | 0.579136371 | 0.1372409033 | 2.44E-05 | 0.3101442011 | 0.84812854 | 1.7844966621 | 1.3636217366 | 2.3352723911 |
| Free cholesterol to total lipids ratio in large VLDL \|\| id:ebi-a-GCST90092873 | Simple mode | 46 | 0.09605426 | 0.2977838677 | 0.7485196552 | -0.4876021188 | 0.6797106399 | 1.1008187933 | 0.6140971622 | 1.9733066511 |

| | | | | | | | | | | |
|---|---|---|---|---|---|---|---|---|---|---|
| Free cholesterol to total lipids ratio in large VLDL \|\| id:ebi-a-GCST90092873 | Weighted mode | 46 | 0.217932462 | 0.071244731 | 0.003734538 | 0.07829279 | 0.357572135 | 1.243503082 | 1.081439247 | 1.429853705 |
| Free cholesterol to total lipids ratio in medium VLDL \|\| id:ebi-a-GCST90092921 | MR Egger | 53 | 0.249095173 | 0.222270659 | 0.267671978 | -0.186555318 | 0.684745664 | 1.282864121 | 0.829812657 | 1.983267355 |
| Free cholesterol to total lipids ratio in | Weighted median | 53 | 0.128123203 | 0.103154445 | 0.214217311 | -0.074059509 | 0.330305915 | 1.136693038 | 0.928616431 | 1.391393712 |

| | | | | | | | | | | |
|---|---|---|---|---|---|---|---|---|---|---|
| medium VLDL \|\| id:ebi-a-GCST90092921 | | | | | | | | | | |
| Free cholesterol to total lipids ratio in medium VLDL \|\| id:ebi-a-GCST90092921 | Inverse variance weighted | 53 | 0.1822466996 | 0.10077891 | 0.0705471777 | -0.0152799688 | 0.3797733599 | 1.19991017 | 0.9848361788 | 1.4619532133 |
| Free cholesterol to total lipids ratio in medium VLDL \|\| id:ebi-a-GCST9 | Simple mode | 53 | 0.08623162 | 0.2312039122 | 0.7106884411 | -0.3669280466 | 0.5393912877 | 1.0900587799 | 0.6928594977 | 1.7149626244 |

| | | | | | | | | | | |
|---|---|---|---|---|---|---|---|---|---|---|
| 0092921 | | | | | | | | | | |
| Free cholesterol to total lipids ratio in medium VLDL \|\| id:ebi-a-GCST90092921 | Weighted mode | 53 | 0.184280762 | 0.112350125 | 0.106992211 | -0.035925482 | 0.404487006 | 1.202353351 | 0.964712179 | 1.498533565 |
| Free cholesterol to total lipids ratio in small VLDL \|\| id:ebi-a-GCST90092973 | MR Egger | 67 | 0.337411847 | 0.228627417 | 0.144821563 | -0.11069789 | 0.785521583 | 1.401316072 | 0.89520916 | 2.193550762 |
| Free cholesterol to total lipids | Weighted median | 67 | -0.016195303 | 0.108644106 | 0.881500385 | -0.22913775 | 0.196747144 | 0.983935136 | 0.795218984 | 1.217436166 |

| | | | | | | | | | | |
|---|---|---|---|---|---|---|---|---|---|---|
| ratio in small VLDL \|\| id:ebi-a-GCST90092973 | | | | | | | | | | |
| Free cholesterol to total lipids ratio in small VLDL \|\| id:ebi-a-GCST90092973 | Inverse variance weighted | 67 | 0.09989351 | 0.132321205 | 0.450289795 | -0.159456053 | 0.359243072 | 1.105053234 | 0.852607436 | 1.432244899 |
| Free cholesterol to total lipids ratio in small VLDL \|\| id:ebi-a-GCST90092973 | Simple mode | 67 | -0.163635097 | 0.300591456 | 0.588016577 | -0.75279435 | 0.425524156 | 0.849051787 | 0.471048438 | 1.530392374 |

| | | | | | | | | | | |
|---|---|---|---|---|---|---|---|---|---|---|
| Free cholesterol to total lipids ratio in small VLDL \|\| id:ebi-a-GCST90092973 | Weighted mode | 67 | -0.401800151 | 0.10094801 | 0.000173883 | -0.599658251 | -0.203942052 | 0.669114454 | 0.548999224 | 0.815509627 |
| Free cholesterol to total lipids ratio in very small VLDL \|\| id:ebi-a-GCST90093033 | MR Egger | 48 | 0.438435214 | 0.270141698 | 0.111427584 | -0.091042514 | 0.967912942 | 1.550279464 | 0.912978896 | 2.632444659 |
| Free cholesterol to total lipids ratio in | Weighted median | 48 | 0.088536749 | 0.115399479 | 0.442950923 | -0.137646229 | 0.314719727 | 1.092574403 | 0.871406916 | 1.369875319 |

| | | | | | | | | | | |
|---|---|---|---|---|---|---|---|---|---|---|
| very small VLDL \|\| id:ebi-a-GCST90093033 | | | | | | | | | | |
| Free cholesterol to total lipids ratio in very small VLDL \|\| id:ebi-a-GCST90093033 | Inverse variance weighted | 48 | 0.2990943 79 | 0.1514580 21 | 0.0482945 38 | 0.0022366 57 | 0.5959521 | 1.3486369 | 1.00223916 | 1.8147579 54 |
| Free cholesterol to total lipids ratio in very small VLDL \|\| id:ebi-a-GCST9 | Simple mode | 48 | 0.18287043 | 0.4867307 88 | 0.70882174 | -0.7711219 14 | 1.1368627 75 | 1.2006588 29 | 0.4624938 99 | 3.1169743 59 |

| | | | | | | | | | | |
|---|---|---|---|---|---|---|---|---|---|---|
| 0093033 | | | | | | | | | | |
| Free cholesterol to total lipids ratio in very small VLDL \|\| id:ebi-a-GCST90093033 | Weighted mode | 48 | -0.101423723 | 0.16180106 | 0.533795917 | -0.4185538 | 0.215706355 | 0.903550097 | 0.657997728 | 1.240737989 |
| Free cholesterol to total lipids ratio in IDL \|\| id:ebi-a-GCST90092836 | MR Egger | 45 | -0.36843558 | 0.276146083 | 0.189159727 | -0.909681902 | 0.172810743 | 0.691815775 | 0.402652287 | 1.188641125 |
| Free cholesterol to total lipids ratio | Weighted median | 45 | -0.461606693 | 0.106126302 | 1.36E-05 | -0.669614245 | -0.253599141 | 0.630270181 | 0.51190601 | 0.776002807 |

| | | | | | | | | | | |
|---|---|---|---|---|---|---|---|---|---|---|
| in IDL \|\| id:ebi-a-GCST90092836 | | | | | | | | | | |
| Free cholesterol to total lipids ratio in IDL \|\| id:ebi-a-GCST90092836 | Inverse variance weighted | 45 | −0.278603138 | 0.156818107 | 0.075633983 | −0.585966629 | 0.028760353 | 0.756840205 | 0.556567608 | 1.029177925 |
| Free cholesterol to total lipids ratio in IDL \|\| id:ebi-a-GCST90092836 | Simple mode | 45 | −0.363257446 | 0.215388791 | 0.098774772 | −0.785419475 | 0.058904584 | 0.695407381 | 0.455928411 | 1.06067403 |
| Free cholesterol to | Weighted | 45 | −0.405164128 | 0.09847801 | 0.000167536 | −0.598181027 | −0.212147229 | 0.66686735 | 0.549810818 | 0.808845604 |

| | | | | | | | | | | |
|---|---|---|---|---|---|---|---|---|---|---|
| total lipids ratio in IDL \|\| id:ebi-a-GCST90092836 | mode | | | | | | | | | |
| Free cholesterol to total lipids ratio in large LDL \|\| id:ebi-a-GCST90092861 | MR Egger | 81 | -0.139165208 | 0.171929815 | 0.420698924 | -0.476147645 | 0.197817229 | 0.870084272 | 0.621171763 | 1.218739623 |
| Free cholesterol to total lipids ratio in large LDL \|\| id:ebi-a-GCST9 | Weighted median | 81 | -0.458912309 | 0.085119925 | 6.99E-08 | -0.625747362 | -0.292077256 | 0.63197066 | 0.534861544 | 0.746710846 |

| | | | | | | | | | | |
|---|---|---|---|---|---|---|---|---|---|---|
| 0092861 | | | | | | | | | | |
| Free cholesterol to total lipids ratio in large LDL \|\| id:ebi-a-GCST90092861 | Inverse variance weighted | 81 | -0.246987267 | 0.105088118 | 0.018758926 | -0.452959978 | -0.041014557 | 0.78115064 | 0.635743577 | 0.959815158 |
| Free cholesterol to total lipids ratio in large LDL \|\| id:ebi-a-GCST90092861 | Simple mode | 81 | -0.472737974 | 0.205762976 | 0.024204261 | -0.876033407 | -0.069442542 | 0.623293369 | 0.416431454 | 0.932913736 |
| Free cholesterol to total lipids ratio | Weighted mode | 81 | -0.441919943 | 0.098673984 | 2.47E-05 | -0.635320952 | -0.248518934 | 0.642801094 | 0.529765432 | 0.779955093 |

| | | | | | | | | | | |
|---|---|---|---|---|---|---|---|---|---|---|
| in large LDL \|\| id:ebi-a-GCST900092861 | | | | | | | | | | |
| Free cholesterol to total lipids ratio in medium LDL \|\| id:ebi-a-GCST90092909 | MR Egger | 71 | -0.135674523 | 0.167071144 | 0.419540665 | -0.463133965 | 0.191784919 | 0.873126769 | 0.629308321 | 1.211409937 |
| Free cholesterol to total lipids ratio in medium LDL \|\| id:ebi-a-GCST90092909 | Weighted median | 71 | -0.451723535 | 0.080076529 | 1.69E-08 | -0.608673531 | -0.294773539 | 0.636530124 | 0.544072085 | 0.744700214 |

| | | | | | | | | | | |
|---|---|---|---|---|---|---|---|---|---|---|
| Free cholesterol to total lipids ratio in medium LDL \|\| id:ebi-a-GCST90092909 | Inverse variance weighted | 71 | -0.245150678 | 0.1032347744 | 0.0175636511 | -0.4474908344 | -0.0428105221 | 0.7825866111 | 0.6392300755 | 0.9580929111 |
| Free cholesterol to total lipids ratio in medium LDL \|\| id:ebi-a-GCST90092909 | Simple mode | 71 | -0.1689302288 | 0.1874021944 | 0.3704501099 | -0.5362385299 | 0.1983780722 | 0.8445678288 | 0.5849443711 | 1.2194233377 |
| Free cholesterol to total lipids ratio in mediu | Weighted mode | 71 | -0.4227387442 | 0.0815231211 | 2.00E-06 | -0.5825240599 | -0.2629534255 | 0.6552498 | 0.5584869322 | 0.7687777033 |

<table>
<tr><td>m LDL || id:ebi-a-GCST90092909</td><td></td><td></td><td></td><td></td><td></td><td></td><td></td><td></td><td></td><td></td></tr>
<tr><td>Free cholesterol to total lipids ratio in small LDL || id:ebi-a-GCST90092961</td><td>MR Egger</td><td>69</td><td>0.035967789</td><td>0.169864905</td><td>0.832949952</td><td>-0.296967424</td><td>0.368903001</td><td>1.036622455</td><td>0.743068218</td><td>1.446147323</td></tr>
<tr><td>Free cholesterol to total lipids ratio in small LDL || id:ebi-a-GCST90092961</td><td>Weighted median</td><td>69</td><td>-0.377980176</td><td>0.077287118</td><td>1.01E-06</td><td>-0.529462927</td><td>-0.226497424</td><td>0.685244085</td><td>0.588921178</td><td>0.797321396</td></tr>
<tr><td>Free cholestero</td><td>Inverse</td><td>69</td><td>-0.164</td><td>0.108234378</td><td>0.128323313</td><td>-0.376</td><td>0.047542264</td><td>0.848235375</td><td>0.686096839</td><td>1.048690522</td></tr>
</table>

| | | | | | | | | | | |
|---|---|---|---|---|---|---|---|---|---|---|
| l to total lipids ratio in small LDL \|\| id:ebi-a-GCST90092961 | variance weighted | | 597117 | | | 736497 | | | | |
| Free cholesterol to total lipids ratio in small LDL \|\| id:ebi-a-GCST90092961 | Simple mode | 69 | −0.19378937 | 0.16541328 | 0.245467782 | −0.517999398 | 0.130420658 | 0.823831409 | 0.595711137 | 1.139307541 |
| Free cholesterol to total lipids ratio in small LDL \|\| id:eb | Weighted mode | 69 | −0.363002737 | 0.071018545 | 2.80E−06 | −0.502199087 | −0.223806388 | 0.695584529 | 0.605198312 | 0.799469906 |

| | | | | | | | | | | |
|---|---|---|---|---|---|---|---|---|---|---|
| i-a-GCST90092961 | | | | | | | | | | |
| Free cholesterol levels in very large HDL \|\| id:ebi-a-GCST90093008 | MR Egger | 71 | 0.1868505543 | 0.14283026 | 0.1951467911 | -0.0930967677 | 0.4667978522 | 1.2054471109 | 0.9111053311 | 1.5948789669 |
| Free cholesterol levels in very large HDL \|\| id:ebi-a-GCST90093008 | Weighted median | 71 | 0.09646919 | 0.0663928444 | 0.1462221344 | -0.0336607855 | 0.2265991655 | 1.1012756511 | 0.9668994366 | 1.2543269899 |
| Free cholesterol levels in very large HDL \|\| | Inverse variance weighted | 71 | 0.0203541211 | 0.0914557033 | 0.8238801877 | -0.1588990577 | 0.1996072988 | 1.0205626799 | 0.8530824688 | 1.2209232055 |

| | | | | | | | | | | |
|---|---|---|---|---|---|---|---|---|---|---|
| id:ebi-a-GCST90093008 | | | | | | | | | | |
| Free cholesterol levels in very large HDL \|\| id:ebi-a-GCST90093008 | Simple mode | 71 | -0.128361472 | 0.211360086 | 0.545609074 | -0.542627241 | 0.285904297 | 0.879535395 | 0.581219242 | 1.330965072 |
| Free cholesterol levels in very large HDL \|\| id:ebi-a-GCST90093008 | Weighted mode | 71 | 0.055703625 | 0.060284989 | 0.358658057 | -0.062454954 | 0.173862204 | 1.057284285 | 0.939455381 | 1.189891592 |
| Free cholesterol levels in large HDL \|\| | MR Egger | 87 | -0.083664781 | 0.108600972 | 0.443206311 | -0.296522686 | 0.129193123 | 0.919739518 | 0.743398763 | 1.13790986 |

| | | | | | | | | | | |
|---|---|---|---|---|---|---|---|---|---|---|
| id:ebi-a-GCST90092848 | | | | | | | | | | |
| Free cholesterol levels in large HDL \|\| id:ebi-a-GCST90092848 | Weighted median | 87 | -0.108507776 | 0.0769138922 | 0.1583125777 | -0.2592590004 | 0.0422434522 | 0.8971719188 | 0.7716231444 | 1.0431484044 |
| Free cholesterol levels in large HDL \|\| id:ebi-a-GCST90092848 | Inverse variance weighted | 87 | -0.2412635033 | 0.0716671633 | 0.00076144 | -0.3817311444 | -0.1007958633 | 0.7856345822 | 0.6826785711 | 0.9041175788 |
| Free cholesterol levels in large HDL \|\| id:ebi-a- | Simple mode | 87 | -0.0324299775 | 0.2213023 | 0.8838368866 | -0.4661824844 | 0.4013225344 | 0.9680902388 | 0.6273927844 | 1.4937989911 |

| | | | | | | | | | | |
|---|---|---|---|---|---|---|---|---|---|---|
| GCST900092848 | | | | | | | | | | |
| Free cholesterol levels in large HDL \|\| id:ebi-a-GCST90092848 | Weighted mode | 87 | 0.0130659055 | 0.0537585833 | 0.8085471444 | -0.0923009188 | 0.1184327288 | 1.0131516337 | 0.9118307222 | 1.1257311411 |
| Free cholesterol levels in medium HDL \|\| id:ebi-a-GCST90092896 | MR Egger | 75 | -0.1684181449 | 0.1469168955 | 0.25539421 | -0.4563752644 | 0.1195389666 | 0.8450004245 | 0.6335760344 | 1.1269771577 |
| Free cholesterol levels in medium HDL \|\| id:ebi-a-GCST9 | Weighted median | 75 | -0.1787721644 | 0.0824835288 | 0.0302068011 | -0.34043988 | -0.0171044449 | 0.8362964166 | 0.7114572988 | 0.9830410022 |

<table>
<tr><td>0092896</td><td></td><td></td><td></td><td></td><td></td><td></td><td></td><td></td><td></td><td></td></tr>
<tr><td>Free cholesterol levels in medium HDL || id:ebi-a-GCST90092896</td><td>Inverse variance weighted</td><td>75</td><td>-0.226163729</td><td>0.084050691</td><td>0.007128058</td><td>-0.390903084</td><td>-0.061424375</td><td>0.797587503</td><td>0.676445711</td><td>0.940424063</td></tr>
<tr><td>Free cholesterol levels in medium HDL || id:ebi-a-GCST90092896</td><td>Simple mode</td><td>75</td><td>0.000676727</td><td>0.211189561</td><td>0.997451919</td><td>-0.413254812</td><td>0.414608266</td><td>1.000676956</td><td>0.661493705</td><td>1.513777626</td></tr>
<tr><td>Free cholesterol levels in medium HDL || id:ebi-a-GCST90092896</td><td>Weighted mode</td><td>75</td><td>-0.045391619</td><td>0.079434479</td><td>0.569436598</td><td>-0.201083198</td><td>0.110299961</td><td>0.955623169</td><td>0.817844385</td><td>1.116612961</td></tr>
</table>

| | | | | | | | | | | |
|---|---|---|---|---|---|---|---|---|---|---|
| Free cholesterol levels in small HDL \|\| id:ebi-a-GCST90092948 | MR Egger | 47 | 0.2151641966 | 0.1916848066 | 0.2676078811 | -0.1605380233 | 0.5908664166 | 1.2400654944 | 0.8516854399 | 1.8055520988 |
| Free cholesterol levels in small HDL \|\| id:ebi-a-GCST90092948 | Weighted median | 47 | 0.3223694233 | 0.0965862844 | 0.0008449577 | 0.1330603077 | 0.5116785399 | 1.3803946311 | 1.1423188866 | 1.6680887988 |
| Free cholesterol levels in small HDL \|\| id:ebi-a-GCST90092948 | Inverse variance weighted | 47 | 0.2113313133 | 0.0913497444 | 0.0206989644 | 0.0322858155 | 0.3903768111 | 1.2353215666 | 1.0328126577 | 1.4775374422 |

| | | | | | | | | | | |
|---|---|---|---|---|---|---|---|---|---|---|
| Free cholesterol levels in small HDL \|\| id:ebi-a-GCST90092948 | Simple mode | 47 | 0.2064452688 | 0.2215842899 | 0.35636713 | -0.2278599388 | 0.6407504774 | 1.2293004511 | 0.7962357774 | 1.8979046744 |
| Free cholesterol levels in small HDL \|\| id:ebi-a-GCST90092948 | Weighted mode | 47 | 0.3650269422 | 0.1262044711 | 0.0058229966 | 0.1176661799 | 0.6123877044 | 1.4405528188 | 1.1248685444 | 1.8448310555 |
| Free cholesterol levels in HDL \|\| id:ebi-a-GCST90092824 | MR Egger | 76 | -0.1212752556 | 0.1308309699 | 0.3569600644 | -0.3777039555 | 0.1351534422 | 0.8857901077 | 0.68543339 | 1.1447124188 |
| Free cholesterol | Weighted | 76 | -0.05747979 | 0.0760316663 | 0.4496503544 | -0.20650185 | 0.0915422699 | 0.9441409711 | 0.8134247566 | 1.0958630977 |

| | | | | | | | | | | |
|---|---|---|---|---|---|---|---|---|---|---|
| l levels in HDL \|\| id:ebi-a-GCST90092824 | median | | | | | | | | | |
| Free cholesterol levels in HDL \|\| id:ebi-a-GCST90092824 | Inverse variance weighted | 76 | -0.16696983 | 0.0779004833 | 0.03208312 | -0.3196547 77 | -0.0142848 84 | 0.8462251 41 | 0.7263997 64 | 0.9858166 61 |
| Free cholesterol levels in HDL \|\| id:ebi-a-GCST90092824 | Simple mode | 76 | 0.0613223 02 | 0.19315933 | 0.7517691 16 | -0.3172699 85 | 0.4399145 89 | 1.0632415 44 | 0.7281341 43 | 1.5525746 07 |
| Free cholesterol levels in HDL \|\| | Weighted mode | 76 | 0.0004003 89 | 0.0636246 57 | 0.9949956 61 | -0.1243039 39 | 0.1251047 18 | 1.00040047 | 0.8831113 88 | 1.13326712 |

| | | | | | | | | | | |
|---|---|---|---|---|---|---|---|---|---|---|
| id:ebi-a-GCST90092824 | | | | | | | | | | |
| Total lipid levels in very large HDL \|\| id:ebi-a-GCST90093010 | MR Egger | 77 | 0.0812092566 | 0.13770618 | 0.5571451889 | -0.1886948588 | 0.3511133699 | 1.0845978311 | 0.8280391388 | 1.4206483755 |
| Total lipid levels in very large HDL \|\| id:ebi-a-GCST90093010 | Weighted median | 77 | 0.1054098799 | 0.0724449077 | 0.1456595599 | -0.0365821399 | 0.2474018988 | 1.1111659611 | 0.9640789022 | 1.2806937177 |
| Total lipid levels in very large HDL \|\| id:ebi-a-GCST90093010 | Inverse variance weighted | 77 | -0.1166208111 | 0.0919944555 | 0.20490728 | -0.2969299433 | 0.0636883222 | 0.8899225788 | 0.7430960699 | 1.0657601733 |

| | | | | | | | | | | |
|---|---|---|---|---|---|---|---|---|---|---|
| Total lipid levels in very large HDL \|\| id:ebi-a-GCST90093010 | Simple mode | 77 | -0.2435499558 | 0.23154057 | 0.2961934445 | -0.6973694755 | 0.2102695559 | 0.7838403166 | 0.4978933033 | 1.2340106544 |
| Total lipid levels in very large HDL \|\| id:ebi-a-GCST90093010 | Weighted mode | 77 | 0.0328911557 | 0.0545167033 | 0.5480905155 | -0.07396158 | 0.1397438944 | 1.0334380551 | 0.9287073744 | 1.1499792455 |
| Total lipid levels in large HDL \|\| id:ebi-a-GCST90092850 | MR Egger | 91 | -0.1093257677 | 0.1084472444 | 0.3161389666 | -0.3218823644 | 0.1032308331 | 0.89643834 | 0.7247834466 | 1.1087473133 |
| Total lipid levels in large HDL | Weighted median | 91 | -0.1694414077 | 0.0723849977 | 0.0192406177 | -0.3113160011 | -0.0275668133 | 0.8441362144 | 0.7324823744 | 0.9728096844 |

| | | | | | | | | | | |
|---|---|---|---|---|---|---|---|---|---|---|
| \|\| id:ebi-a-GCST90092850 | | | | | | | | | | |
| Total lipid levels in large HDL \|\| id:ebi-a-GCST90092850 | Inverse variance weighted | 91 | -0.233917418 | 0.06976767 | 0.000799969 | -0.37066205 | -0.097172785 | 0.791427171 | 0.690277181 | 0.907399208 |
| Total lipid levels in large HDL \|\| id:ebi-a-GCST90092850 | Simple mode | 91 | -0.061283793 | 0.213959418 | 0.775209269 | -0.480644253 | 0.358076667 | 0.940556279 | 0.618384867 | 1.430575294 |
| Total lipid levels in large HDL \|\| id:ebi-a-GCST90092850 | Weighted mode | 91 | 0.008817168 | 0.059232892 | 0.882000335 | -0.1072793 | 0.124913637 | 1.008856154 | 0.89827475 | 1.133050595 |

| | | | | | | | | | | |
|---|---|---|---|---|---|---|---|---|---|---|
| Total lipid levels in medium HDL \|\| id:ebi-a-GCST90092898 | MR Egger | 64 | -0.183951353 | 0.166586408 | 0.273756294 | -0.510460714 | 0.142558007 | 0.831976276 | 0.600218986 | 1.153219974 |
| Total lipid levels in medium HDL \|\| id:ebi-a-GCST90092898 | Weighted median | 64 | -0.186717245 | 0.090885065 | 0.039933835 | -0.364851972 | -0.008582518 | 0.829678299 | 0.694299419 | 0.991454206 |
| Total lipid levels in medium HDL \|\| id:ebi-a-GCST90092898 | Inverse variance weighted | 64 | -0.2353903 | 0.090802322 | 0.009532609 | -0.413362851 | -0.057417749 | 0.79026235 | 0.661422242 | 0.944199548 |
| Total lipid levels in medium HDL \|\| id:eb | Simple mode | 64 | -0.332799155 | 0.243839027 | 0.177160862 | -0.810723648 | 0.145125338 | 0.716914168 | 0.444536262 | 1.156184475 |

| | | | | | | | | | | |
|---|---|---|---|---|---|---|---|---|---|---|
| i-a-GCST900092898 | | | | | | | | | | |
| Total lipid levels in medium HDL \|\| id:ebi-a-GCST900092898 | Weighted mode | 64 | −0.1404044 29 | 0.0892545 16 | 0.1207095 29 | −0.31534328 | 0.0345344 21 | 0.8690067 13 | 0.7295383 96 | 1.0351376 58 |
| Total lipid levels in small HDL \|\| id:ebi-a-GCST900092950 | MR Egger | 46 | 0.2127879 | 0.1833201 53 | 0.2520047 86 | −0.1465196 | 0.5720954 | 1.23712223 | 0.8637088 03 | 1.7719761 63 |
| Total lipid levels in small HDL \|\| id:ebi-a-GCST900092950 | Weighted median | 46 | 0.2237945 15 | 0.0991439 33 | 0.0239912 79 | 0.0294724 06 | 0.4181166 24 | 1.2508139 69 | 1.0299110 16 | 1.5190978 27 |
| Total lipid levels in | Inverse var | 46 | 0.1255322 55 | 0.1007572 26 | 0.2128053 86 | −0.07195190 9 | 0.3230164 19 | 1.1337517 38 | 0.9305756 47 | 1.38128803 |

| | | | | | | | | | | |
|---|---|---|---|---|---|---|---|---|---|---|
| small HDL \|\| id:ebi-a-GCST90092950 | iance weighted | | | | | | | | | |
| Total lipid levels in small HDL \|\| id:ebi-a-GCST90092950 | Simple mode | 46 | 0.1052841 99 | 0.2186715 15 | 0.6325142 94 | -0.32331197 | 0.5338803 67 | 1.1110263 18 | 0.7237480 32 | 1.7055375 97 |
| Total lipid levels in small HDL \|\| id:ebi-a-GCST90092950 | Weighted mode | 46 | 0.1976988 22 | 0.1275295 52 | 0.1280939 31 | -0.0522591 | 0.4476567 44 | 1.2185953 24 | 0.9490829 27 | 1.5646415 31 |
| Free cholesterol to total lipids ratio in very large HDL | MR Egger | 76 | 0.0129961 13 | 0.0941260 97 | 0.8905590 87 | -0.1714910 37 | 0.1974832 62 | 1.0130809 29 | 0.8424078 19 | 1.2183326 73 |

| | | | | | | | | | | |
|---|---|---|---|---|---|---|---|---|---|---|
| \|\| id:ebi-a-GCST900930 09 | | | | | | | | | | |
| Free cholesterol to total lipids ratio in very large HDL \|\| id:ebi-a-GCST900930 09 | Weighted median | 76 | -0.07176467 9 | 0.07766688 1 | 0.35548308 6 | -0.22399176 5 | 0.08046240 7 | 0.93074989 5 | 0.79932171 6 | 1.08378810 3 |
| Free cholesterol to total lipids ratio in very large HDL \|\| id:ebi-a-GCST900930 09 | Inverse variance weighted | 76 | 0.12489017 | 0.06447282 3 | 0.05273337 7 | -0.00147656 3 | 0.25125690 3 | 1.13302400 6 | 0.99852452 7 | 1.28564032 7 |

| | | | | | | | | | | |
|---|---|---|---|---|---|---|---|---|---|---|
| Free cholesterol to total lipids ratio in very large HDL \|\| id:ebi-a-GCST90093009 | Simple mode | 76 | 0.1626683 51 | 0.2306422 83 | 0.4828181 56 | -0.2893905 23 | 0.6147272 25 | 1.1766463 91 | 0.7487197 56 | 1.8491521 28 |
| Free cholesterol to total lipids ratio in very large HDL \|\| id:ebi-a-GCST90093009 | Weighted mode | 76 | -0.0077816 2 | 0.0599042 19 | 0.8969924 98 | -0.1251938 89 | 0.1096306 49 | 0.9922485 79 | 0.8823258 13 | 1.1158658 48 |
| Free cholesterol to total lipids ratio | MR Egger | 62 | 0.3531773 43 | 0.1527417 05 | 0.0242121 44 | 0.0538036 01 | 0.6525510 86 | 1.4235835 84 | 1.0552773 27 | 1.9204337 76 |

| | | | | | | | | | | |
|---|---|---|---|---|---|---|---|---|---|---|
| in large HDL \|\| id:ebi-a-GCST90092849 | | | | | | | | | | |
| Free cholesterol to total lipids ratio in large HDL \|\| id:ebi-a-GCST90092849 | Weighted median | 62 | 0.11369277 1 | 0.07003423 8 | 0.10450639 7 | -0.02357433 5 | 0.25095987 8 | 1.12040785 1 | 0.97670136 9 | 1.28525851 6 |
| Free cholesterol to total lipids ratio in large HDL \|\| id:ebi-a-GCST90092849 | Inverse variance weighted | 62 | 0.14960552 3 | 0.09870148 8 | 0.12958579 9 | -0.04384939 2 | 0.34306043 9 | 1.16137601 7 | 0.95709809 3 | 1.40925393 3 |

| | | | | | | | | | | |
|---|---|---|---|---|---|---|---|---|---|---|
| Free cholesterol to total lipids ratio in large HDL \|\| id:ebi-a-GCST90092849 | Simple mode | 62 | 0.0323339555 | 0.1887006088 | 0.8645152822 | -0.3375192377 | 0.4021871466 | 1.0328623777 | 0.7135382488 | 1.4950911088 |
| Free cholesterol to total lipids ratio in large HDL \|\| id:ebi-a-GCST90092849 | Weighted mode | 62 | 0.1120243566 | 0.0621883255 | 0.0765868888 | -0.00986476 | 0.2339134733 | 1.1185401044 | 0.9901837377 | 1.2635351577 |
| Free cholesterol to total lipids ratio in mediu | MR Egger | 83 | -0.0963260911 | 0.1411215922 | 0.4968228377 | -0.3729244111 | 0.1802722288 | 0.9081678222 | 0.68871729 | 1.1975433244 |

| | | | | | | | | | | |
|---|---|---|---|---|---|---|---|---|---|---|
| m HDL \|\| id:ebi-a-GCST90092897 | | | | | | | | | | |
| Free cholesterol to total lipids ratio in medium HDL \|\| id:ebi-a-GCST90092897 | Weighted median | 83 | -0.162838525 | 0.071999355 | 0.02371802 | -0.303957261 | -0.021719789 | 0.849728387 | 0.737892403 | 0.978514387 |
| Free cholesterol to total lipids ratio in medium HDL \|\| id:ebi-a-GCST90092897 | Inverse variance weighted | 83 | -0.147332574 | 0.087022224 | 0.09044683 | -0.317896132 | 0.023230985 | 0.863006916 | 0.727678367 | 1.023502926 |
| Free cholestero | Simple | 83 | -0.038 | 0.205524694 | 0.851636598 | -0.441 | 0.36426748 | 0.962173087 | 0.643142268 | 1.43945919 |

| | | | | | | | | | | |
|---|---|---|---|---|---|---|---|---|---|---|
| l to total lipids ratio in medium HDL \|\| id:ebi-a-GCST90092897 | mode | | 560921 | | | 389322 | | | | |
| Free cholesterol to total lipids ratio in medium HDL \|\| id:ebi-a-GCST90092897 | Weighted mode | 83 | -0.020842933 | 0.065396024 | 0.750750282 | -0.149019139 | 0.107333274 | 0.97937278 | 0.861552625 | 1.113305229 |
| Free cholesterol to total lipids ratio in small HDL \|\| id:eb | MR Egger | 68 | 0.190957126 | 0.109776719 | 0.086607879 | -0.024205244 | 0.406119497 | 1.210407556 | 0.976085354 | 1.500981904 |

| | | | | | | | | | | |
|---|---|---|---|---|---|---|---|---|---|---|
| i-a-GCST90092949 | | | | | | | | | | |
| Free cholesterol to total lipids ratio in small HDL \|\| id:ebi-a-GCST90092949 | Weighted median | 68 | 0.100851275 | 0.056112642 | 0.072287939 | -0.009129503 | 0.210832054 | 1.106112123 | 0.990912044 | 1.234704974 |
| Free cholesterol to total lipids ratio in small HDL \|\| id:ebi-a-GCST90092949 | Inverse variance weighted | 68 | 0.245691552 | 0.075018921 | 0.001056391 | 0.098654466 | 0.392728639 | 1.278505161 | 1.103684873 | 1.481016444 |
| Free cholesterol to total lipid | Simple mode | 68 | 0.261605404 | 0.193185764 | 0.180233558 | -0.117038693 | 0.640249502 | 1.299013856 | 0.889550773 | 1.896954114 |

| | | | | | | | | | | |
|---|---|---|---|---|---|---|---|---|---|---|
| s ratio in small HDL \|\| id:ebi-a-GCST90092949 | | | | | | | | | | |
| Free cholesterol to total lipids ratio in small HDL \|\| id:ebi-a-GCST90092949 | Weighted mode | 68 | 0.085349635 | 0.044749564 | 0.060773351 | -0.002359511 | 0.173058781 | 1.089097787 | 0.99764327 | 1.18893599 |
