## Supplemental Table S1 for "Mendelian Randomization Study Reveals That Combined Spontaneous Free Cholesterol Diffusion and Reverse Cholesterol Transport Pathways Shift from Pro-Atherogenic to Anti-Atherogenic Effects"

### Supplemental Material

Table S1: The selected SNPs

| exposure | chr.exposure | pos.exposure | beta.exposure | se.exposure | pval.exposure | SNP | effect_allele.exposure | other_allele.exposure | eaf.exposure | R2 | F |
|---|---|---|---|---|---|---|---|---|---|---|---|
| Free cholesterol levels in chylomicrons and extremely large VLDL \|\| id:ebi-a-GCST90093044 | 1 | 230302838 | -0.0400333 | 0.00411896 | 2.50E-22 | rs59484402 | CT | C | 0.611688 | 0.00082017 | 94.46264146 |
| | 1 | 63060037 | -0.0817947 | 0.00415351 | 2.50E-86 | rs79439217 | TAAGAA | T | 0.352737 | 0.003358546 | 387.8039777 |
| | 1 | 27284913 | 0.0959345 | 0.0131824 | 3.40E-13 | rs79598313 | T | C | 0.023201 | 0.000459995 | 52.96062652 |
| | 2 | 21231524 | -0.0973343 | 0.00489291 | 4.70E-88 | rs676210 | A | G | 0.205766 | 0.003426882 | 395.7216555 |
| | 2 | 27730940 | -0.0879509 | 0.00405566 | 2.80E-104 | rs1260326 | C | T | 0.603982 | 0.004069855 | 470.2728963 |
| | 2 | 165528876 | -0.0504882 | 0.00406452 | 2.00E-35 | rs13389219 | T | C | 0.392532 | 0.001338973 | 154.2956555 |
| | 2 | 227077377 | 0.0398786 | 0.00425361 | 6.90E-21 | rs2943635 | T | C | 0.679832 | 0.000763177 | 87.89353311 |
| | 3 | 135956305 | 0.0361897 | 0.00470025 | 1.40E-14 | rs684773 | C | A | 0.767075 | 0.000514869 | 59.28167992 |
| | 4 | 100508556 | -0.0394449 | 0.00716273 | 3.70E-08 | rs2035816 | G | A | 0.084301 | 0.000263453 | 30.32613122 |

| | | | | | | | | | | | |
|---|---|---|---|---|---|---|---|---|---|---|---|
| | 4 | 87976359 | 0.0337612 | 0.00407524 | 1.20E-16 | rs1471251 | T | A | 0.397121 | 0.000596023 | 68.6312361 |
| | 4 | 171563776 | -0.0261163 | 0.00467165 | 2.30E-08 | rs4491981 | A | C | 0.241218 | 0.000271492 | 31.25180846 |
| | 4 | 3443931 | -0.0311763 | 0.00411102 | 3.40E-14 | rs13108218 | G | A | 0.615355 | 0.000499489 | 57.50987411 |
| | 5 | 55857025 | 0.0541497 | 0.00506168 | 1.00E-26 | rs11429307 | GT | G | 0.191517 | 0.00099349 | 114.4445698 |
| | 5 | 156443066 | 0.0371819 | 0.00414312 | 2.90E-19 | rs4704834 | G | A | 0.644028 | 0.000699354 | 80.53795259 |
| | 6 | 31303922 | 0.0250666 | 0.00411061 | 1.10E-09 | rs28752924 | C | T | 0.445537 | 0.000323021 | 37.1852551 |
| | 6 | 43757896 | 0.0261954 | 0.00396715 | 4.00E-11 | rs998584 | A | C | 0.482313 | 0.000378722 | 43.59987839 |
| | 6 | 161010118 | -0.238256 | 0.0073451 | 1.00E-200 | rs10455872 | G | A | 0.078987 | 0.009060083 | 1052.167103 |
| | 6 | 160922870 | -0.360562 | 0.0146992 | 7.19E-133 | rs117733303 | G | A | 0.0185 | 0.005201162 | 601.6791504 |
| | 6 | 32587213 | 0.0356552 | 0.00408767 | 2.70E-18 | rs28383314 | C | T | 0.622964 | 0.000660693 | 76.08281114 |
| | 6 | 160093887 | -0.0896924 | 0.0157385 | 1.20E-08 | rs184089815 | A | G | 0.016433 | 0.000282134 | 32.47708678 |

| | | | | | | | | | | | |
|---|---|---|---|---|---|---|---|---|---|---|---|
| | 6 | 160578069 | -0.114945 | 0.0171682 | 2.20E-11 | rs146534110 | T | G | 0.013508 | 0.00389362 | 44.82528853 |
| | 6 | 127451665 | 0.0291451 | 0.00399693 | 3.10E-13 | rs1936806 | T | C | 0.448453 | 0.00461817 | 53.17046608 |
| | 6 | 139834012 | -0.0335938 | 0.00409902 | 2.50E-16 | rs632057 | G | T | 0.627778 | 0.00583307 | 67.16617897 |
| | 6 | 161080457 | 0.0779734 | 0.0139233 | 2.10E-08 | rs117857195 | T | G | 0.02424 | 0.00272448 | 31.36180387 |
| | 6 | 31850308 | 0.0699569 | 0.00958166 | 2.90E-13 | rs74434374 | A | C | 0.044813 | 0.00462989 | 53.30550387 |
| | 6 | 160710851 | -0.0478405 | 0.00544866 | 1.60E-18 | rs3127580 | T | C | 0.156213 | 0.00669443 | 77.09113777 |
| | 6 | 161017363 | -0.117389 | 0.0110319 | 1.90E-26 | rs73596816 | A | G | 0.033593 | 0.00982923 | 113.22612 |
| | 7 | 130432481 | 0.0282933 | 0.00436565 | 9.10E-11 | rs6971365 | C | T | 0.292826 | 0.00364841 | 42.00121288 |
| | 7 | 25992323 | -0.0421371 | 0.00543037 | 8.50E-15 | rs10642257 | AAGGCC | A | 0.160158 | 0.00522921 | 60.20920534 |
| | 7 | 73020337 | -0.113316 | 0.00591249 | 7.20E-82 | rs3812316 | G | C | 0.129282 | 0.03181634 | 367.3111276 |
| | 7 | 73123473 | 0.0642246 | 0.00921698 | 3.20E-12 | rs72555385 | G | A | 0.049304 | 0.0042173 | 48.55321342 |

| | | | | | | | | | | | |
|---|---|---|---|---|---|---|---|---|---|---|---|
| | 8 | 8363683 | -0.035007 | 0.00401606 | 2.90E-18 | rs2980755 | G | A | 0.458034 | 0.00659805 | 75.98044913 |
| | 8 | 19819724 | -0.190011 | 0.00658411 | 3.90E-183 | rs328 | G | C | 0.100307 | 0.007184959 | 832.8288931 |
| | 8 | 18239697 | 0.030459 | 0.00516246 | 3.60E-09 | rs13266992 | C | T | 0.180568 | 0.00302398 | 34.8105059 |
| | 8 | 126500031 | -0.0789352 | 0.00406506 | 5.40E-84 | rs28601761 | G | C | 0.420159 | 0.003265724 | 377.0508807 |
| | 8 | 126631799 | 0.0335268 | 0.00487264 | 6.00E-12 | rs3860847 | A | G | 0.212508 | 0.00411216 | 47.34215333 |
| | 8 | 19733244 | -0.0650994 | 0.0048367 | 2.70E-41 | rs1561748 | C | G | 0.267445 | 0.01571684 | 181.1540811 |
| | 10 | 65160321 | -0.0371764 | 0.00397428 | 8.40E-21 | rs10761752 | C | T | 0.474418 | 0.00759767 | 87.50043147 |
| | 10 | 94796504 | -0.0250439 | 0.00408932 | 9.10E-10 | rs835273 | T | A | 0.38487 | 0.00325801 | 37.50543278 |
| | 11 | 61575158 | 0.0680185 | 0.00417325 | 1.00E-59 | rs174553 | G | A | 0.342558 | 0.02303014 | 265.6426162 |
| | 11 | 64021821 | 0.0413441 | 0.00702154 | 3.90E-09 | rs2302263 | T | C | 0.08812 | 0.00301179 | 34.6700756 |
| | 11 | 116389017 | 0.0246765 | 0.00433344 | 1.20E-08 | rs883863 | A | G | 0.308012 | 0.0028169 | 32.42604639 |

| | | | | | | | | | | | |
|---|---|---|---|---|---|---|---|---|---|---|---|
| | 11 | 117156893 | -0.264372 | 0.020191 | 3.60E-39 | rs1047964 | T | G | 0.010073 | 0.001487515 | 171.43824433 |
| | 11 | 116648917 | -0.211165 | 0.00585107 | 1.00E-200 | rs964184 | C | G | 0.86726 | 0.01119124 | 1302.464029 |
| | 11 | 68562328 | 0.0471092 | 0.00796655 | 3.40E-09 | rs2229738 | T | C | 0.066036 | 0.00303761 | 34.96739974 |
| | 11 | 116916060 | 0.278788 | 0.020095 | 9.20E-44 | rs144018203 | C | G | 0.010653 | 0.001669702 | 192.47068814 |
| | 12 | 109874794 | 0.023603 | 0.00431761 | 4.60E-08 | rs10850137 | G | C | 0.305274 | 0.00259614 | 29.88409052 |
| | 12 | 57781893 | -0.0281538 | 0.00464574 | 1.40E-09 | rs2122982 | A | G | 0.238672 | 0.0031902 | 36.72456778 |
| | 12 | 124409502 | -0.0255866 | 0.00425902 | 1.90E-09 | rs7133378 | A | G | 0.319789 | 0.00313518 | 36.09094494 |
| | 13 | 95231825 | 0.029735 | 0.00415429 | 8.20E-13 | rs2275647 | A | G | 0.351884 | 0.00444981 | 51.2312223 |
| | 13 | 114544024 | 0.0292314 | 0.004339 | 1.60E-11 | rs7140110 | C | T | 0.299365 | 0.00394222 | 45.3850046 |
| | 13 | 114623821 | -0.0363526 | 0.00642395 | 1.50E-08 | rs41284486 | A | G | 0.111972 | 0.00278188 | 32.02278532 |
| | 15 | 44027885 | 0.140793 | 0.0125285 | 2.70E-29 | rs139974673 | C | T | 0.025912 | 0.01096176 | 126.2863536 |

| | | | | | | | | | | | |
|---|---|---|---|---|---|---|---|---|---|---|---|
| | 16 | 81534790 | －0.0306254 | 0.0043293 | 1.50E-12 | rs2925979 | C | T | 0.698949 | 0.000434642 | 50.04038346 |
| | 16 | 15172118 | －0.0288494 | 0.00436453 | 3.80E-11 | rs11644601 | C | T | 0.296695 | 0.00379513 | 43.69090664 |
| | 16 | 56993886 | －0.0453797 | 0.0042473 | 1.20E-26 | rs821840 | G | A | 0.324526 | 0.00990967 | 114.1536541 |
| | 16 | 72219756 | －0.0354393 | 0.00531771 | 2.70E-11 | rs34417180 | A | C | 0.166852 | 0.00385786 | 44.41333888 |
| | 17 | 64210580 | －0.110233 | 0.011549 | 1.40E-21 | rs1801689 | C | A | 0.030526 | 0.00791013 | 91.10179699 |
| | 17 | 17434141 | 0.0479249 | 0.00822965 | 5.80E-09 | rs9889584 | A | G | 0.062203 | 0.00294594 | 33.91190229 |
| | 17 | 41926126 | 0.110419 | 0.0113762 | 2.80E-22 | rs72836561 | T | C | 0.031482 | 0.00817957 | 94.20753374 |
| | 19 | 19393890 | －0.0912192 | 0.007651 | 9.00E-33 | rs200210321 | AG | A | 0.073166 | 0.01233652 | 142.1440112 |
| | 19 | 45401783 | 0.0437396 | 0.00429314 | 2.20E-24 | rs56290633 | C | T | 0.539526 | 0.00901157 | 103.7987119 |
| | 19 | 8439538 | －0.0543676 | 0.00862293 | 2.90E-10 | rs57192995 | C | G | 0.056297 | 0.00345313 | 39.75237267 |
| | 19 | 8429323 | －0.22754 | 0.0142387 | 1.70E-57 | rs116843064 | A | G | 0.019851 | 0.02214138 | 255.3684734 |

| | | | | | | | | | | | |
|---|---|---|---|---|---|---|---|---|---|---|---|
| | 19 | 45413233 | 0.200631 | 0.00732303 | 3.00E-165 | rs1065853 | T | G | 0.080581 | 0.006480128 | 750.5971332 |
| | 19 | 45470298 | 0.0264341 | 0.00398589 | 3.30E-11 | rs11878597 | C | T | 0.45019 | 0.000382037 | 43.98158651 |
| | 20 | 39179822 | 0.0723621 | 0.0116732 | 5.70E-10 | rs1883711 | C | G | 0.031207 | 0.000333803 | 38.42686222 |
| exposure | chr.exposure | pos.exposure | beta.exposure | se.exposure | pval.exposure | SNP | effect_allele.exposure | other_allele.exposure | eaf.exposure | R2 | F |
| Free cholesterol levels in very large VLDL \|\| id:ebi-a-GCST90093020 | 1 | 63060037 | -0.0773121 | 0.00417601 | 1.60E-76 | rs79439217 | TAAGAA | T | 0.352737 | 0.002969429 | 342.739637 |
| | 1 | 109821797 | 0.0313108 | 0.00475944 | 4.70E-11 | rs4970836 | A | G | 0.773326 | 0.000375929 | 43.27818789 |
| | 1 | 27284913 | 0.105856 | 0.0132538 | 1.40E-15 | rs79598313 | T | C | 0.023201 | 0.00055399 | 63.78852871 |
| | 1 | 219618564 | 0.0297583 | 0.00544017 | 4.50E-08 | rs61838776 | A | C | 0.161109 | 0.000259939 | 29.92152645 |
| | 1 | 230295789 | -0.0434221 | 0.00408618 | 2.20E-26 | rs10127775 | T | A | 0.605599 | 0.000980287 | 112.9221429 |
| | 2 | 21231524 | -0.109088 | 0.00491924 | 5.90E-109 | rs676210 | A | G | 0.205766 | 0.004254991 | 491.7568165 |
| | 2 | 21530659 | -0.028571 | 0.00407384 | 2.30E-12 | rs60960031 | A | G | 0.402852 | 0.000427218 | 49.18530837 |

| | | | | | | | | | | | |
|---|---|---|---|---|---|---|---|---|---|---|---|
| | 2 | 27730940 | －0.0848325 | 0.00407749 | 3.90E-96 | rs1260326 | C | T | 0.603982 | 0.003747148 | 432.8437737 |
| | 2 | 227093585 | 0.0362694 | 0.0041758 | 3.80E-18 | rs2943640 | C | A | 0.651848 | 0.00655103 | 75.43863382 |
| | 2 | 219699999 | 0.0596522 | 0.0103284 | 7.70E-09 | rs78058190 | A | G | 0.050108 | 0.0028977 | 33.35640858 |
| | 2 | 165528876 | －0.0496128 | 0.00408639 | 6.40E-34 | rs13389219 | T | C | 0.392532 | 0.001279218 | 147.4009569 |
| | 3 | 52838654 | －0.0366552 | 0.0061202 | 2.10E-09 | rs9968117 | T | C | 0.120935 | 0.003116 | 35.87008536 |
| | 3 | 135956305 | 0.038106 | 0.00472605 | 7.40E-16 | rs684773 | C | A | 0.767075 | 0.00564596 | 65.01035755 |
| | 4 | 3443931 | －0.0331868 | 0.00413333 | 9.80E-16 | rs13108218 | G | A | 0.615355 | 0.00559861 | 64.46486063 |
| | 4 | 87976359 | 0.0330816 | 0.00409736 | 6.80E-16 | rs1471251 | T | A | 0.397121 | 0.00566124 | 65.18643312 |
| | 4 | 171563776 | －0.0264175 | 0.00469701 | 1.90E-08 | rs4491981 | A | C | 0.241218 | 0.00274798 | 31.63245617 |
| | 5 | 55857025 | 0.054694 | 0.00508817 | 6.00E-27 | rs11429307 | GT | G | 0.191517 | 0.01003028 | 115.5443194 |
| | 5 | 74656539 | 0.0272855 | 0.00407601 | 2.20E-11 | rs12916 | C | T | 0.400341 | 0.00389239 | 44.81112052 |

| | | | | | | | | | | | |
|---|---|---|---|---|---|---|---|---|---|---|---|
| | 5 | 156443066 | 0.041279 | 0.0041648 | 3.70E-23 | rs4704834 | G | A | 0.644028 | 0.00852888 | 98.23414882 |
| | 6 | 32586831 | 0.0429031 | 0.00429708 | 1.80E-23 | rs13191810 | T | C | 0.312309 | 0.0086546 | 99.6834181 |
| | 6 | 139834012 | -0.0354259 | 0.00412417 | 8.70E-18 | rs632057 | G | T | 0.627778 | 0.00640742 | 73.78381718 |
| | 6 | 160922870 | -0.303219 | 0.0147894 | 2.10E-93 | rs117733303 | G | A | 0.0185 | 0.0036393325 | 420.3433337 |
| | 6 | 161017363 | -0.09959 | 0.0110996 | 2.90E-19 | rs73596816 | A | G | 0.033593 | 0.00699046 | 80.50248017 |
| | 6 | 32622150 | 0.0406403 | 0.00696295 | 5.30E-09 | rs35932914 | A | G | 0.090022 | 0.00295932 | 34.06588763 |
| | 6 | 160545046 | 0.0632313 | 0.00983458 | 1.30E-10 | rs111909590 | T | C | 0.043584 | 0.00359078 | 41.3375791 |
| | 6 | 161010118 | -0.17945 | 0.00739018 | 3.00E-130 | rs10455872 | G | A | 0.078987 | 0.05097416 | 589.6161148 |
| | 6 | 31236679 | 0.0253858 | 0.00454777 | 2.40E-08 | rs1130580 | C | T | 0.289865 | 0.00270682 | 31.15854546 |
| | 6 | 43757896 | 0.03035 | 0.00399149 | 2.90E-14 | rs998584 | A | C | 0.482313 | 0.00502136 | 57.81489644 |
| | 6 | 127455029 | 0.0327353 | 0.00400025 | 2.80E-16 | rs2186037 | G | A | 0.518794 | 0.00581565 | 66.96545673 |

| | | | | | | | | | | | |
|---|---|---|---|---|---|---|---|---|---|---|---|
| | 7 | 25983400 | -0.0252702 | 0.00417107 | 1.40E-09 | rs56001710 | T | A | 0.580583 | 0.00318843 | 36.7041232 |
| | 7 | 130433594 | 0.0330281 | 0.00430962 | 1.80E-14 | rs11976955 | G | C | 0.314211 | 0.00510106 | 58.7329315 |
| | 7 | 73037366 | -0.0888518 | 0.00497186 | 2.00E-71 | rs55747707 | A | G | 0.203642 | 0.02767475 | 319.3648617 |
| | 8 | 126500031 | -0.0847069 | 0.00408776 | 2.20E-95 | rs28601761 | G | C | 0.420159 | 0.03717423 | 429.397241 |
| | 8 | 126631799 | 0.0324854 | 0.00489985 | 3.40E-11 | rs3860847 | A | G | 0.212508 | 0.00381801 | 43.95449751 |
| | 8 | 19733244 | -0.0632265 | 0.00486371 | 1.20E-38 | rs1561748 | C | G | 0.267445 | 0.01466285 | 168.9878152 |
| | 8 | 10649767 | 0.0305646 | 0.00412079 | 1.20E-13 | rs752446 | A | G | 0.375244 | 0.00477817 | 55.01345485 |
| | 8 | 19819724 | -0.190667 | 0.00662088 | 2.30E-182 | rs328 | G | C | 0.100307 | 0.07154739 | 829.3008029 |
| | 8 | 59402570 | -0.0269545 | 0.00426816 | 2.70E-10 | rs7018333 | G | A | 0.671955 | 0.00346436 | 39.88169703 |
| | 10 | 94797973 | -0.0278472 | 0.00407599 | 8.40E-12 | rs2478236 | A | G | 0.403791 | 0.00405428 | 46.67553457 |
| | 10 | 64882300 | -0.0324044 | 0.00402409 | 8.10E-16 | rs10761716 | G | C | 0.44151 | 0.00563145 | 64.84329133 |

| | | | | | | | | | | | |
|---|---|---|---|---|---|---|---|---|---|---|---|
| | 11 | 116648917 | -0.208041 | 0.00588363 | 1.00E-200 | rs964184 | C | G | 0.86726 | 0.010747487 | 1250.257908 |
| | 11 | 116916060 | 0.269087 | 0.0202068 | 1.90E-40 | rs144018203 | C | G | 0.010653 | 0.001538559 | 177.3302393 |
| | 11 | 117156893 | -0.2478 | 0.0203034 | 2.90E-34 | rs1047964 | T | G | 0.010073 | 0.001292694 | 148.9558331 |
| | 11 | 61575158 | 0.0434367 | 0.00419647 | 4.20E-25 | rs174553 | G | A | 0.342558 | 0.000930109 | 107.1365985 |
| | 11 | 116389017 | 0.024607 | 0.00435755 | 1.60E-08 | rs883863 | A | G | 0.308012 | 0.000277016 | 31.8878349 |
| | 12 | 57781893 | -0.0316651 | 0.00467047 | 1.20E-11 | rs2122982 | A | G | 0.238672 | 0.000399264 | 45.96560774 |
| | 12 | 124409502 | -0.0317793 | 0.00428169 | 1.20E-13 | rs7133378 | A | G | 0.319789 | 0.000478457 | 55.08719574 |
| | 13 | 114544024 | 0.0301484 | 0.00436257 | 4.80E-12 | rs7140110 | C | T | 0.299365 | 0.000414817 | 47.75690491 |
| | 13 | 95231825 | 0.0242318 | 0.00417685 | 6.60E-09 | rs2275647 | A | G | 0.351884 | 0.000292374 | 33.65627487 |
| | 15 | 44027885 | 0.127425 | 0.0125975 | 4.70E-24 | rs139974673 | C | T | 0.025912 | 0.000888275 | 102.3135144 |
| | 16 | 81520932 | -0.0224951 | 0.00404416 | 2.70E-08 | rs2927320 | G | A | 0.56231 | 0.000268779 | 30.93938309 |

| | | | | | | | | | | | |
|---|---|---|---|---|---|---|---|---|---|---|---|
| | 16 | 56993886 | -0.0759173 | 0.00426887 | 9.40E-71 | rs821840 | G | A | 0.324526 | 0.002740668 | 316.2627927 |
| | 16 | 72108093 | 0.0381546 | 0.00509902 | 7.30E-14 | rs2000999 | A | G | 0.188613 | 0.000486297 | 55.99030476 |
| | 17 | 17431208 | 0.0559479 | 0.00872364 | 1.40E-10 | rs8078513 | A | C | 0.057349 | 0.000357281 | 41.13055426 |
| | 17 | 41926126 | 0.115656 | 0.0114369 | 4.90E-24 | rs72836561 | T | C | 0.031482 | 0.000887823 | 102.2614958 |
| | 17 | 64210580 | -0.0840626 | 0.0116106 | 4.50E-13 | rs1801689 | C | A | 0.030526 | 0.000455293 | 52.41900371 |
| | 19 | 8423432 | -0.0550856 | 0.00873807 | 2.90E-10 | rs371786442 | AT | A | 0.055899 | 0.000345214 | 39.74088934 |
| | 19 | 49263477 | -0.0229452 | 0.00418476 | 4.20E-08 | rs2081194 | C | G | 0.601344 | 0.000261169 | 30.06318821 |
| | 19 | 11209722 | -0.0462223 | 0.0060487 | 2.10E-14 | rs73015034 | A | C | 0.126031 | 0.000507167 | 58.39443571 |
| | 19 | 19379549 | -0.117996 | 0.00761217 | 3.40E-54 | rs58542926 | T | C | 0.074399 | 0.002083552 | 240.2758185 |
| | 19 | 45430280 | 0.0861813 | 0.0042931 | 1.20E-89 | rs5112 | G | C | 0.533735 | 0.003489463 | 402.9735347 |
| | 19 | 8429323 | -0.22562 | 0.0143202 | 6.30E-56 | rs116843064 | A | G | 0.019851 | 0.002152355 | 248.227261 |

| | | | | | | | | | | | |
|---|---|---|---|---|---|---|---|---|---|---|---|
| | 19 | 45424514 | 0.0539584 | 0.00526722 | 1.30E-24 | rs157592 | C | A | 0.185291 | 0.00091107 | 104.9415954 |
| | 20 | 39179822 | 0.08911 | 0.0117345 | 3.10E-14 | rs1883711 | C | G | 0.031207 | 0.00050084 | 57.66551316 |
| | 20 | 44545048 | -0.0283009 | 0.00461415 | 8.60E-10 | rs4810479 | T | C | 0.750217 | 0.00326789 | 37.61919764 |
| exposure | chr.exposure | pos.exposure | beta.exposure | se.exposure | pval.exposure | SNP | effect_allele.exposure | other_allele.exposure | eaf.exposure | R2 | F |
| Free cholesterol levels in large VLDL \|\| id:ebi-a-GCST90092872 | 1 | 63060037 | -0.0739371 | 0.00417038 | 2.50E-70 | rs79439217 | TAAGAA | T | 0.352737 | 0.00272384 | 314.3156598 |
| | 1 | 109821797 | 0.0419608 | 0.00475302 | 1.10E-18 | rs4970836 | A | G | 0.773326 | 0.00676779 | 77.93646225 |
| | 1 | 27021913 | 0.107766 | 0.0133718 | 7.70E-16 | rs114165349 | C | G | 0.022955 | 0.00564068 | 64.94956314 |
| | 1 | 230296153 | -0.0454758 | 0.00407939 | 7.30E-29 | rs10864726 | T | C | 0.605471 | 0.01078684 | 124.2689687 |
| | 2 | 21530659 | -0.0300758 | 0.00406799 | 1.40E-13 | rs60960031 | A | G | 0.402852 | 0.00474746 | 54.65967881 |
| | 2 | 21231524 | -0.111283 | 0.00491218 | 1.30E-113 | rs676210 | A | G | 0.205766 | 0.04439859 | 513.2176187 |
| | 2 | 165528876 | -0.0486965 | 0.00408053 | 7.90E-33 | rs13389219 | T | C | 0.392532 | 0.01235998 | 142.414693 |

| | | | | | | | | | | | |
|---|---|---|---|---|---|---|---|---|---|---|---|
| | 2 | 219699999 | 0.057752 7 | 0.0103135 | 2.10 E-08 | rs78058190 | A | G | 0.050108 | 0.0002724 | 31.35630586 |
| | 2 | 227093585 | 0.036224 4 | 0.00416981 | 3.70 E-18 | rs2943640 | C | A | 0.651848 | 0.00655357 | 75.46790966 |
| | 2 | 27730940 | -0.088654 8 | 0.00407163 | 4.10 E-105 | rs1260326 | C | T | 0.603982 | 0.00410275 | 474.0895144 |
| | 3 | 52838654 | -0.038257 9 | 0.00611257 | 3.90 E-10 | rs9968117 | T | C | 0.120935 | 0.00340282 | 39.17301659 |
| | 3 | 135956305 | 0.034606 1 | 0.00472016 | 2.30 E-13 | rs684773 | C | A | 0.767075 | 0.00466855 | 53.75072528 |
| | 4 | 87976359 | 0.031328 | 0.00409199 | 1.90 E-14 | rs1471251 | T | A | 0.397121 | 0.00509058 | 58.61228669 |
| | 4 | 171563776 | -0.026057 7 | 0.00469084 | 2.80 E-08 | rs4491981 | A | C | 0.241218 | 0.00268069 | 30.85768719 |
| | 4 | 3443931 | -0.036677 5 | 0.00412791 | 6.40 E-19 | rs13108218 | G | A | 0.615355 | 0.00685542 | 78.94624247 |
| | 5 | 55857025 | 0.056017 2 | 0.00508184 | 3.00 E-28 | rs11429307 | GT | G | 0.191517 | 0.01054715 | 121.5047543 |
| | 5 | 74656539 | 0.026434 6 | 0.00407094 | 8.40 E-11 | rs12916 | C | T | 0.400341 | 0.0036626 | 42.16465349 |
| | 5 | 156443066 | 0.039753 3 | 0.00415963 | 1.20 E-21 | rs4704834 | G | A | 0.644028 | 0.00793022 | 91.3333571 |

| | | | | | | | | | | | |
|---|---|---|---|---|---|---|---|---|---|---|---|
| | 6 | 31308524 | 0.025272 | 0.00430096 | 4.20E-09 | rs9295981 | A | G | 0.342231 | 0.00299924 | 34.5255711 |
| | 6 | 139834012 | -0.0356225 | 0.00412035 | 5.40E-18 | rs632057 | G | T | 0.627778 | 0.00649069 | 74.74343033 |
| | 6 | 160545046 | 0.0632557 | 0.00982547 | 1.20E-10 | rs111909590 | T | C | 0.043584 | 0.00360022 | 41.44623801 |
| | 6 | 160922870 | -0.261643 | 0.0147757 | 3.70E-70 | rs117733303 | G | A | 0.0185 | 0.02717271 | 313.5556011 |
| | 6 | 161010118 | -0.145977 | 0.00738333 | 5.30E-87 | rs10455872 | G | A | 0.078987 | 0.03385199 | 390.8919811 |
| | 6 | 32586831 | 0.0444045 | 0.00429309 | 4.50E-25 | rs13191810 | T | C | 0.312309 | 0.00928759 | 106.9809462 |
| | 6 | 43757896 | 0.0332497 | 0.00398779 | 7.60E-17 | rs998584 | A | C | 0.482313 | 0.00603728 | 69.51897414 |
| | 6 | 127455029 | 0.0338221 | 0.00399654 | 2.60E-17 | rs2186037 | G | A | 0.518794 | 0.0062195 | 71.61850693 |
| | 6 | 161017363 | -0.0835401 | 0.0110893 | 4.90E-14 | rs73596816 | A | G | 0.033593 | 0.00492902 | 56.75108495 |
| | 7 | 130433594 | 0.0342256 | 0.00430398 | 1.80E-15 | rs11976955 | G | C | 0.314211 | 0.00549181 | 63.2345022 |
| | 7 | 73037366 | -0.0934329 | 0.00496536 | 5.50E-79 | rs55747707 | A | G | 0.203642 | 0.03067303 | 354.07122 |

| | | | | | | | | | | | |
|---|---|---|---|---|---|---|---|---|---|---|---|
| | 7 | 25983400 | -0.0236301 | 0.00416561 | 1.40E-08 | rs56001710 | T | A | 0.580583 | 0.00279541 | 32.17854159 |
| | 8 | 19733244 | -0.0631318 | 0.00485732 | 1.30E-38 | rs1561748 | C | G | 0.267445 | 0.01465745 | 168.9255591 |
| | 8 | 59398461 | -0.0265847 | 0.00422631 | 3.20E-10 | rs10504255 | A | G | 0.662958 | 0.00343704 | 39.56701441 |
| | 8 | 126631799 | 0.0316521 | 0.00489342 | 9.90E-11 | rs3860847 | A | G | 0.212508 | 0.00363425 | 41.83815508 |
| | 8 | 19819724 | -0.189059 | 0.00661218 | 8.30E-180 | rs328 | G | C | 0.100307 | 0.00705380 9 | 817.518953 |
| | 8 | 11493678 | 0.0306981 | 0.00411412 | 8.50E-14 | rs13265201 | G | C | 0.396261 | 0.00483561 | 55.6751679 |
| | 8 | 126500031 | -0.0847211 | 0.00408239 | 1.20E-95 | rs28601761 | G | C | 0.420159 | 0.03728417 | 430.672004 |
| | 10 | 64882300 | -0.032902 | 0.00401873 | 2.70E-16 | rs10761716 | G | C | 0.44151 | 0.00582112 | 67.02848302 |
| | 10 | 94797973 | -0.0280076 | 0.00407057 | 6.00E-12 | rs2478236 | A | G | 0.403791 | 0.00411202 | 47.34060326 |
| | 11 | 116714293 | 0.271927 | 0.0210618 | 3.90E-38 | rs141469619 | G | A | 0.010141 | 0.01446364 | 166.6886949 |
| | 11 | 116389017 | 0.0242468 | 0.00435188 | 2.50E-08 | rs883863 | A | G | 0.308012 | 0.00269669 | 31.04184241 |

| | | | | | | | | | | | |
|---|---|---|---|---|---|---|---|---|---|---|---|
| | 11 | 63862882 | 0.049015 | 0.00875364 | 2.20E-08 | rs11231694 | C | G | 0.055796 | 0.000272367 | 31.35256684 |
| | 11 | 116648917 | -0.210004 | 0.00587597 | 1.00E-200 | rs964184 | C | G | 0.86726 | 0.010977283 | 1277.286866 |
| | 11 | 117156893 | -0.239585 | 0.0202769 | 3.20E-32 | rs1047964 | T | G | 0.010073 | 0.001211664 | 139.607446 |
| | 11 | 61575158 | 0.0402225 | 0.00419101 | 8.20E-22 | rs174553 | G | A | 0.342558 | 0.000799735 | 92.10711488 |
| | 12 | 4384844 | -0.0846451 | 0.0155122 | 4.90E-08 | rs76895963 | G | T | 0.020556 | 0.000258665 | 29.77483107 |
| | 12 | 124409502 | -0.0341432 | 0.00427614 | 1.40E-15 | rs7133378 | A | G | 0.319789 | 0.000553677 | 63.75247732 |
| | 12 | 57766392 | -0.0323232 | 0.00466162 | 4.10E-12 | rs4760254 | C | G | 0.239066 | 0.000417605 | 48.07811307 |
| | 12 | 124645301 | 0.0269884 | 0.00493503 | 4.50E-08 | rs146628494 | G | T | 0.210989 | 0.000259809 | 29.90660638 |
| | 13 | 114544024 | 0.0291479 | 0.00435712 | 2.20E-11 | rs7140110 | C | T | 0.299365 | 0.000388722 | 44.75153661 |
| | 15 | 43726625 | 0.1275 | 0.0126226 | 5.50E-24 | rs150844304 | C | A | 0.025671 | 0.000885789 | 102.0270153 |
| | 16 | 56993886 | -0.0829416 | 0.00426298 | 2.60E-84 | rs821840 | G | A | 0.324526 | 0.003278571 | 378.5390431 |

| | | | | | | | | | | | |
|---|---|---|---|---|---|---|---|---|---|---|---|
| | 16 | 72108093 | 0.0392489 | 0.00509199 | 1.30E-14 | rs2000999 | A | G | 0.188613 | 0.00515998 | 59.41175034 |
| | 16 | 81520932 | -0.0234739 | 0.00403858 | 6.20E-09 | rs2927320 | G | A | 0.56231 | 0.00293348 | 33.78357245 |
| | 17 | 64210580 | -0.073369 | 0.0115956 | 2.50E-10 | rs1801689 | C | A | 0.030526 | 0.00347761 | 40.03420427 |
| | 17 | 17431208 | 0.0578478 | 0.00871237 | 3.10E-11 | rs8078513 | A | C | 0.057349 | 0.00382937 | 44.08527199 |
| | 17 | 41926126 | 0.122125 | 0.0114221 | 1.10E-26 | rs72836561 | T | C | 0.031482 | 0.00992381 | 114.3167028 |
| | 19 | 19379549 | -0.12166 | 0.00760249 | 1.20E-57 | rs58542926 | T | C | 0.074399 | 0.02220298 | 256.080413 |
| | 19 | 45424514 | 0.0584875 | 0.00526052 | 1.00E-28 | rs157592 | C | A | 0.185291 | 0.01072989 | 123.612166 |
| | 19 | 8423432 | -0.0548049 | 0.00872696 | 3.40E-10 | rs371786442 | AT | A | 0.055899 | 0.00342576 | 39.43712626 |
| | 19 | 8429323 | -0.224744 | 0.014302 | 1.20E-55 | rs116843064 | A | G | 0.019851 | 0.02141137 | 246.9307169 |
| | 19 | 11209722 | -0.0494628 | 0.00604101 | 2.70E-16 | rs73015034 | A | C | 0.126031 | 0.00582208 | 67.03949556 |
| | 19 | 45430280 | 0.0789414 | 0.00428764 | 1.10E-75 | rs5112 | G | C | 0.533735 | 0.02936894 | 338.9732857 |

| | | | | | | | | | | | |
|---|---|---|---|---|---|---|---|---|---|---|---|
| | 20 | 39174140 | -0.0276877 | 0.00437832 | 2.60E-10 | rs1883715 | T | C | 0.31262 | 0.000347377 | 39.98998249 |
| | 20 | 44551855 | 0.0581815 | 0.00500623 | 3.20E-31 | rs6073958 | C | T | 0.198731 | 0.00117228 | 135.0643345 |
| exposure | chr.exposure | pos.exposure | beta.exposure | se.exposure | pval.exposure | SNP | effect_allele.exposure | other_allele.exposure | eaf.exposure | R2 | F |
| Free cholesterol levels in medium VLDL \|\| id:ebi-a-GCST90092920 | 1 | 109817590 | -0.0987901 | 0.00496275 | 3.60E-88 | rs12740374 | T | G | 0.221064 | 0.003431483 | 396.2547729 |
| | 1 | 55526428 | 0.0853178 | 0.0115464 | 1.50E-13 | rs11206517 | G | T | 0.033012 | 0.000474212 | 54.59822848 |
| | 1 | 25713234 | 0.0240683 | 0.00412526 | 5.40E-09 | rs1293261 | T | A | 0.525176 | 0.000295701 | 34.03930099 |
| | 1 | 62999675 | 0.058785 | 0.00431115 | 2.50E-42 | rs7534572 | G | C | 0.64747 | 0.001613015 | 185.9256164 |
| | 1 | 234852760 | 0.0295578 | 0.00412553 | 7.80E-13 | rs553427 | T | C | 0.517037 | 0.000445845 | 51.3306975 |
| | 1 | 55518467 | -0.0285325 | 0.00430765 | 3.50E-11 | rs2495477 | G | A | 0.393802 | 0.000381089 | 43.87238918 |
| | 1 | 55505647 | -0.281851 | 0.0157171 | 6.50E-72 | rs11591147 | T | G | 0.017457 | 0.002786603 | 321.5783331 |
| | 2 | 165539661 | -0.0247858 | 0.00421368 | 4.00E-09 | rs6717858 | C | T | 0.401534 | 0.000300569 | 34.599931 |

| | | | | | | | | | | | |
|---|---|---|---|---|---|---|---|---|---|---|---|
| | 2 | 134688669 | 0.0269278 | 0.00490782 | 4.10E-08 | rs1900349 | C | T | 0.770721 | 0.00261519 | 30.10349683 |
| | 2 | 44072576 | -0.0446814 | 0.00441705 | 4.70E-24 | rs4299376 | T | G | 0.676277 | 0.00888373 | 102.3249088 |
| | 2 | 21141815 | 0.0241957 | 0.00420616 | 8.80E-09 | rs4533439 | C | T | 0.411038 | 0.00287457 | 33.09003729 |
| | 2 | 21212348 | -0.0287325 | 0.00413198 | 3.60E-12 | rs1320143 | A | G | 0.536438 | 0.00419992 | 48.35293733 |
| | 2 | 27598097 | -0.0480513 | 0.00424189 | 9.60E-30 | rs4665972 | C | T | 0.604609 | 0.01113782 | 128.3169214 |
| | 2 | 21232195 | 0.0947314 | 0.00412592 | 1.20E-116 | rs693 | A | G | 0.520889 | 0.04559887 | 527.1555191 |
| | 3 | 136581611 | -0.0322188 | 0.00506534 | 2.00E-10 | rs71630059 | A | G | 0.210845 | 0.00351432 | 40.45702751 |
| | 3 | 52841220 | -0.0365131 | 0.00650232 | 2.00E-08 | rs13059141 | A | G | 0.11318 | 0.00273927 | 31.53212126 |
| | 4 | 3443931 | -0.0275152 | 0.00427109 | 1.20E-10 | rs13108218 | G | A | 0.615355 | 0.00360499 | 41.50117132 |
| | 4 | 87976359 | 0.0231128 | 0.00423392 | 4.80E-08 | rs1471251 | T | A | 0.397121 | 0.00258881 | 29.79972658 |
| | 5 | 74656539 | 0.0558332 | 0.00421119 | 4.00E-40 | rs12916 | C | T | 0.400341 | 0.01525124 | 175.7793357 |

| | | | | | | | | | | | |
|---|---|---|---|---|---|---|---|---|---|---|---|
| | 5 | 122832289 | 0.0251391 | 0.00438825 | 1.00E-08 | rs7704651 | A | G | 0.331143 | 0.00285092 | 32.81777857 |
| | 5 | 156443066 | 0.0487454 | 0.00430293 | 9.50E-30 | rs4704834 | G | A | 0.644028 | 0.00113903 | 128.3308785 |
| | 5 | 55861464 | 0.0288813 | 0.00523114 | 3.40E-08 | rs28650790 | T | C | 0.192419 | 0.002648 | 30.48128289 |
| | 5 | 74472939 | 0.0334919 | 0.0042978 | 6.60E-15 | rs7707394 | A | G | 0.357075 | 0.00527413 | 60.72669154 |
| | 6 | 32586376 | 0.0473724 | 0.00450851 | 8.00E-26 | rs11755689 | G | A | 0.334143 | 0.00958431 | 110.4020539 |
| | 6 | 160751531 | -0.174714 | 0.0163241 | 9.90E-27 | rs9295128 | T | G | 0.016821 | 0.00994392 | 114.5484911 |
| | 6 | 32434716 | 0.0498035 | 0.00644227 | 1.10E-14 | rs28377109 | A | C | 0.116096 | 0.0051905 | 59.7632678 |
| | 6 | 160514283 | 0.0558001 | 0.00779006 | 7.90E-13 | rs80254170 | G | A | 0.075568 | 0.00445643 | 51.30749382 |
| | 6 | 160986915 | -0.042604 | 0.00506146 | 3.90E-17 | rs6938647 | C | A | 0.78233 | 0.00615282 | 70.85028252 |
| | 6 | 16134247 | -0.0233501 | 0.00415324 | 1.90E-08 | rs6920309 | T | C | 0.456849 | 0.00274585 | 31.6079177 |
| | 6 | 160569068 | 0.0385574 | 0.00583331 | 3.80E-11 | rs62440901 | T | C | 0.149233 | 0.00379501 | 43.68957506 |

| | | | | | | | | | | | |
|---|---|---|---|---|---|---|---|---|---|---|---|
| | 7 | 21513359 | 0.0231812 | 0.00424867 | 4.90E-08 | rs1989985 | G | C | 0.38286 | 0.00025861 | 29.76859105 |
| | 7 | 73037366 | -0.0389351 | 0.00513777 | 3.50E-14 | rs55747707 | A | G | 0.203642 | 0.00049878 | 57.42826877 |
| | 7 | 44584551 | 0.0346365 | 0.00532792 | 8.00E-11 | rs10260606 | C | G | 0.182652 | 0.00367101 | 42.2615151 |
| | 8 | 19830921 | -0.0822462 | 0.00625218 | 1.60E-39 | rs10096633 | T | C | 0.123907 | 0.01501443 | 173.04058505 |
| | 8 | 9184691 | 0.0382212 | 0.00686085 | 2.50E-08 | rs6601299 | C | T | 0.899281 | 0.00269605 | 31.03453481 |
| | 8 | 126506694 | -0.0717052 | 0.00428353 | 6.70E-63 | rs112875651 | A | G | 0.392379 | 0.02429037 | 280.2141935 |
| | 8 | 19773155 | 0.0962244 | 0.0155717 | 6.40E-10 | rs144503444 | C | T | 0.018253 | 0.00331701 | 38.18482632 |
| | 8 | 59385919 | -0.0285984 | 0.00437206 | 6.10E-11 | rs2326077 | T | C | 0.663229 | 0.00371657 | 42.78619815 |
| | 8 | 116658583 | -0.0295557 | 0.00461088 | 1.50E-10 | rs2737245 | T | G | 0.278602 | 0.00356905 | 41.0872725 |
| | 9 | 136149711 | 0.052719 | 0.00533667 | 5.20E-23 | rs115478735 | T | A | 0.183304 | 0.00847263 | 97.5856778 |
| | 11 | 116714293 | 0.128527 | 0.0217965 | 3.70E-09 | rs141469619 | G | A | 0.010141 | 0.00302048 | 34.77023852 |

| | | | | | | | | | | | |
|---|---|---|---|---|---|---|---|---|---|---|---|
| | 11 | 61557803 | -0.0305861 | 0.00431397 | 1.30E-12 | rs102275 | C | T | 0.34979 | 0.000436613 | 50.26739962 |
| | 11 | 126260684 | 0.053997 | 0.0074666 | 4.80E-13 | rs12804228 | G | A | 0.08472 | 0.000454244 | 52.29810393 |
| | 11 | 116648917 | -0.113818 | 0.00608094 | 3.60E-78 | rs964184 | C | G | 0.86726 | 0.00303496 | 350.3264399 |
| | 12 | 124852564 | -0.0241003 | 0.00420272 | 9.80E-09 | rs11057601 | G | A | 0.591494 | 0.00285662 | 32.88337887 |
| | 12 | 9172646 | 0.0252171 | 0.00420628 | 2.00E-09 | rs10842724 | C | T | 0.412903 | 0.00312213 | 35.94068711 |
| | 13 | 114551993 | 0.0297503 | 0.00439533 | 1.30E-11 | rs6602909 | C | T | 0.328942 | 0.00397942 | 45.81336339 |
| | 14 | 24874026 | 0.0228717 | 0.0041403 | 3.30E-08 | rs72694391 | C | T | 0.482108 | 0.00265101 | 30.51586767 |
| | 15 | 58678720 | -0.0418848 | 0.00434917 | 5.90E-22 | rs261290 | C | T | 0.654645 | 0.00805272 | 92.74544238 |
| | 15 | 58725839 | 0.0292775 | 0.00455345 | 1.30E-10 | rs633695 | G | A | 0.292354 | 0.00359107 | 41.34083912 |
| | 16 | 72217113 | -0.0274976 | 0.00489046 | 1.90E-08 | rs7202323 | G | T | 0.229928 | 0.0027464 | 31.61423014 |
| | 16 | 56989590 | -0.0664348 | 0.00441005 | 2.80E-51 | rs247616 | T | C | 0.324699 | 0.01968073 | 226.9324957 |

| | | | | | | | | | | | |
|---|---|---|---|---|---|---|---|---|---|---|---|
| | 16 | 72101525 | 0.0475696 | 0.00531184 | 3.40E-19 | rs34042070 | G | C | 0.186978 | 0.000696401 | 80.19762027 |
| | 17 | 67191270 | －0.0432996 | 0.00615622 | 2.00E-12 | rs72631343 | G | C | 0.129021 | 0.00042968 | 49.46886637 |
| | 17 | 67081278 | 0.128662 | 0.0138751 | 1.80E-20 | rs77542162 | G | A | 0.022666 | 0.000746615 | 85.98462565 |
| | 17 | 45766771 | 0.0295281 | 0.00415679 | 1.20E-12 | rs56325564 | A | G | 0.482691 | 0.000438285 | 50.46000499 |
| | 19 | 11197261 | －0.143655 | 0.00634374 | 1.60E-113 | rs12151108 | A | G | 0.119677 | 0.004436213 | 512.7943074 |
| | 19 | 45039971 | －0.102871 | 0.0135182 | 2.70E-14 | rs74747585 | C | T | 0.025173 | 0.000502947 | 57.90827666 |
| | 19 | 45215081 | －0.0851771 | 0.0146694 | 6.40E-09 | rs111278137 | A | G | 0.021614 | 0.000292878 | 33.71424589 |
| | 19 | 11227480 | 0.034068 | 0.00420352 | 5.30E-16 | rs2738447 | C | A | 0.59318 | 0.000570442 | 65.68398501 |
| | 19 | 19379549 | －0.127732 | 0.007857 | 2.00E-59 | rs58542926 | T | C | 0.074399 | 0.002291302 | 264.2885637 |
| | 19 | 45215711 | －0.0427102 | 0.00607383 | 2.00E-12 | rs9973286 | G | C | 0.134761 | 0.000429481 | 49.44591272 |
| | 19 | 45401868 | 0.117379 | 0.0105867 | 1.40E-28 | rs118170342 | C | T | 0.040768 | 0.001067059 | 122.9283493 |

| | | | | | | | | | | | |
|---|---|---|---|---|---|---|---|---|---|---|---|
| | 19 | 45413233 | -0.315192 | 0.00760185 | 1.00E-200 | rs1065853 | T | G | 0.080581 | 0.014718539 | 1719.112241 |
| | 20 | 39179822 | 0.109082 | 0.0121189 | 2.20E-19 | rs1883711 | C | G | 0.031207 | 0.00703504 | 81.01626784 |
| | 22 | 50868669 | 0.0259907 | 0.00425271 | 9.90E-10 | rs9616847 | T | A | 0.388272 | 0.00324456 | 37.35053856 |

| exposure | chr.exposure | pos.exposure | beta.exposure | se.exposure | pval.exposure | SNP | effect_allele.exposure | other_allele.exposure | eaf.exposure | R2 | F |
|---|---|---|---|---|---|---|---|---|---|---|---|
| Free cholesterol levels in small VLDL \|\| id:ebi-a-GCST90092972 | 1 | 55505647 | -0.294517 | 0.0157191 | 2.50E-78 | rs11591147 | T | G | 0.017457 | 0.03041132 | 351.0409933 |
| | 1 | 55518467 | -0.0294232 | 0.0043082 | 8.50E-12 | rs2495477 | G | A | 0.393802 | 0.0040514 | 46.64236329 |
| | 1 | 55526428 | 0.0894919 | 0.0115479 | 9.20E-15 | rs11206517 | G | T | 0.033012 | 0.00521588 | 60.05565125 |
| | 1 | 25713234 | 0.0231466 | 0.00412579 | 2.00E-08 | rs1293261 | T | A | 0.525176 | 0.00273422 | 31.47404981 |
| | 1 | 62999675 | 0.0525596 | 0.0043117 | 3.50E-34 | rs7534572 | G | C | 0.64747 | 0.01289553 | 148.5933875 |
| | 1 | 234852760 | 0.02984 | 0.00412606 | 4.80E-13 | rs553427 | T | C | 0.517037 | 0.00454278 | 52.30208616 |
| | 1 | 109817590 | -0.107157 | 0.00496338 | 2.20E-103 | rs12740374 | T | G | 0.221064 | 0.04033882 | 466.0993449 |

| | | | | | | | | | | | |
|---|---|---|---|---|---|---|---|---|---|---|---|
| | 2 | 21232195 | 0.0983249 | 0.00412622 | 1.70E-125 | rs693 | A | G | 0.520889 | 0.004909952 | 567.8252746 |
| | 2 | 44072576 | -0.0433896 | 0.00441737 | 9.00E-23 | rs4299376 | T | G | 0.676277 | 0.00837669 | 96.47975468 |
| | 2 | 63268355 | 0.030635 | 0.00545153 | 1.90E-08 | rs12620918 | C | T | 0.173985 | 0.00274329 | 31.57849361 |
| | 2 | 27598097 | -0.047851 | 0.00424219 | 1.70E-29 | rs4665972 | C | T | 0.604609 | 0.0110437 | 127.2313856 |
| | 2 | 165539661 | -0.0234697 | 0.00421399 | 2.60E-08 | rs6717858 | C | T | 0.401534 | 0.00269466 | 31.0184811 |
| | 2 | 20927997 | -0.0556305 | 0.00834192 | 2.60E-11 | rs6729843 | C | T | 0.065781 | 0.00386295 | 44.47196691 |
| | 3 | 136581611 | -0.0317518 | 0.00506597 | 3.70E-10 | rs71630059 | A | G | 0.210845 | 0.00341237 | 39.28293477 |
| | 4 | 3443931 | -0.0270402 | 0.00427186 | 2.50E-10 | rs13108218 | G | A | 0.615355 | 0.00348038 | 40.06620685 |
| | 5 | 74472939 | 0.033203 | 0.00429874 | 1.10E-14 | rs7707394 | A | G | 0.357075 | 0.00518131 | 59.65745826 |
| | 5 | 122832289 | 0.0248685 | 0.0043892 | 1.50E-08 | rs7704651 | A | G | 0.331143 | 0.00278869 | 32.10117229 |
| | 5 | 55861464 | 0.0290048 | 0.00523228 | 3.00E-08 | rs28650790 | T | C | 0.192419 | 0.00266953 | 30.7291289 |

| | | | | | | | | | | | |
|---|---|---|---|---|---|---|---|---|---|---|---|
| | 5 | 74656539 | 0.0542951 | 0.00421211 | 5.10E-38 | rs12916 | C | T | 0.400341 | 0.001441743 | 166.1553457 |
| | 5 | 156443066 | 0.0475694 | 0.00430386 | 2.10E-28 | rs4704834 | G | A | 0.644028 | 0.001060403 | 122.1607044 |
| | 6 | 16134247 | −0.0238797 | 0.00415444 | 9.00E-09 | rs6920309 | T | C | 0.456849 | 0.000287012 | 33.03887137 |
| | 6 | 32434716 | 0.0506034 | 0.00644412 | 4.10E-15 | rs28377109 | A | C | 0.116096 | 0.00053554 | 61.66299426 |
| | 6 | 160751531 | −0.130945 | 0.0163288 | 1.10E-15 | rs9295128 | T | G | 0.016821 | 0.000558495 | 64.30751874 |
| | 6 | 30598588 | 0.0545362 | 0.00999667 | 4.90E-08 | rs34193381 | A | G | 0.045941 | 0.000258547 | 29.76127193 |
| | 6 | 160543148 | 0.0691981 | 0.00811326 | 1.50E-17 | rs12208357 | T | C | 0.07 | 0.000631707 | 72.74279852 |
| | 6 | 32586376 | 0.0473496 | 0.00450981 | 8.70E-26 | rs11755689 | G | A | 0.334143 | 0.000956958 | 110.2322294 |
| | 7 | 44584551 | 0.0344065 | 0.00532882 | 1.10E-10 | rs10260606 | C | G | 0.182652 | 0.000362121 | 41.68802721 |
| | 7 | 73037366 | −0.0359056 | 0.00513864 | 2.80E-12 | rs55747707 | A | G | 0.203642 | 0.000424069 | 48.82254819 |
| | 7 | 21513359 | 0.0239442 | 0.00424939 | 1.80E-08 | rs1989985 | G | C | 0.38286 | 0.000275817 | 31.74972237 |

| | | | | | | | | | | | |
|---|---|---|---|---|---|---|---|---|---|---|---|
| | 8 | 19830921 | －0.071868 | 0.00625343 | 1.40E-30 | rs10096633 | T | C | 0.123907 | 0.001146381 | 132.0769359 |
| | 8 | 19773155 | 0.0947444 | 0.0155748 | 1.20E-09 | rs144503444 | C | T | 0.018253 | 0.000321451 | 37.00450465 |
| | 8 | 59385919 | －0.0264191 | 0.00437294 | 1.50E-09 | rs2326077 | T | C | 0.663229 | 0.000317062 | 36.49904092 |
| | 8 | 126506694 | －0.0726765 | 0.00428439 | 1.50E-64 | rs112875651 | A | G | 0.392379 | 0.002494124 | 287.7414757 |
| | 8 | 116658583 | －0.0300069 | 0.0046118 | 7.70E-11 | rs2737245 | T | G | 0.278602 | 0.000367734 | 42.33443659 |
| | 9 | 136149711 | 0.0605995 | 0.00533751 | 7.10E-30 | rs115478735 | T | A | 0.183304 | 0.001118838 | 128.9000633 |
| | 11 | 61565908 | －0.0289892 | 0.00429168 | 1.40E-11 | rs174541 | C | T | 0.357464 | 0.000396313 | 45.62577442 |
| | 11 | 126260684 | 0.0572142 | 0.00746784 | 1.80E-14 | rs12804228 | G | A | 0.08472 | 0.000509787 | 58.69621581 |
| | 11 | 116648917 | －0.103607 | 0.00608195 | 4.50E-65 | rs964184 | C | G | 0.86726 | 0.002515309 | 290.1916813 |
| | 12 | 124419728 | －0.0249561 | 0.00436044 | 1.00E-08 | rs11057397 | T | C | 0.337502 | 0.000284552 | 32.75557721 |
| | 12 | 9172646 | 0.0253635 | 0.00420696 | 1.70E-09 | rs10842724 | C | T | 0.412903 | 0.000315745 | 36.34745883 |

| | | | | | | | | | | | |
|---|---|---|---|---|---|---|---|---|---|---|---|
| | 13 | 114551993 | 0.0294062 | 0.00439606 | 2.20E-11 | rs6602909 | C | T | 0.328942 | 0.00388664 | 44.74484862 |
| | 14 | 24874026 | 0.0248084 | 0.00414089 | 2.10E-09 | rs72694391 | C | T | 0.482108 | 0.00311794 | 35.89240635 |
| | 15 | 58725839 | 0.0425953 | 0.00455382 | 8.50E-21 | rs633695 | G | A | 0.292354 | 0.00759686 | 87.4911439 |
| | 15 | 58678720 | -0.0595658 | 0.00434952 | 1.10E-42 | rs261290 | C | T | 0.654645 | 0.01627034 | 187.5442468 |
| | 16 | 72101525 | 0.0469697 | 0.00531222 | 9.40E-19 | rs34042070 | G | C | 0.186978 | 0.00678861 | 78.17644543 |
| | 16 | 56989590 | -0.0789067 | 0.00441037 | 1.40E-71 | rs247616 | T | C | 0.324699 | 0.02773729 | 320.0885679 |
| | 16 | 72217113 | -0.0277544 | 0.00489081 | 1.40E-08 | rs7202323 | G | T | 0.229928 | 0.00279752 | 32.20286827 |
| | 17 | 67191270 | -0.0443831 | 0.00615701 | 5.70E-13 | rs72631343 | G | C | 0.129021 | 0.00451328 | 51.96225624 |
| | 17 | 7080316 | -0.0290477 | 0.0052473 | 3.10E-08 | rs55714927 | T | C | 0.190994 | 0.00266213 | 30.64390904 |
| | 17 | 45766771 | 0.0317396 | 0.00415732 | 2.30E-14 | rs56325564 | A | G | 0.482691 | 0.00506231 | 58.28656229 |
| | 17 | 67081278 | 0.136706 | 0.0138768 | 6.80E-23 | rs77542162 | G | A | 0.022666 | 0.00842603 | 97.04852664 |

| | | | | | | | | | | | |
|---|---|---|---|---|---|---|---|---|---|---|---|
| | 19 | 11197261 | −0.147705 | 0.00634394 | 6.61E-120 | rs12151108 | A | G | 0.119677 | 0.004688392 | 542.081658 |
| | 19 | 19379549 | −0.112646 | 0.00785725 | 1.30E-46 | rs58542926 | T | C | 0.074399 | 0.001782822 | 205.533614 |
| | 19 | 44543760 | −0.0882283 | 0.0125956 | 2.50E-12 | rs62115478 | G | A | 0.029334 | 0.00426173 | 49.06485891 |
| | 19 | 45039971 | −0.107047 | 0.0135187 | 2.40E-15 | rs74747585 | C | T | 0.025173 | 0.00544547 | 62.70058498 |
| | 19 | 45337918 | 0.0812086 | 0.0113053 | 6.80E-13 | rs147711004 | A | G | 0.036038 | 0.00448165 | 51.59793447 |
| | 19 | 11227480 | 0.0354692 | 0.00420365 | 3.20E-17 | rs2738447 | C | A | 0.59318 | 0.00618264 | 71.19379448 |
| | 19 | 45401868 | 0.120665 | 0.010587 | 4.30E-30 | rs118170342 | C | T | 0.040768 | 0.01127508 | 129.9000328 |
| | 19 | 45413233 | −0.334733 | 0.00760209 | 1.00E-200 | rs1065853 | T | G | 0.080581 | 0.016567921 | 1938.757533 |
| | 20 | 39179822 | 0.11158 | 0.0121205 | 3.40E-20 | rs1883711 | C | G | 0.031207 | 0.00735876 | 84.74695343 |
| | 22 | 50868669 | 0.025289 | 0.00425344 | 2.80E-09 | rs9616847 | T | A | 0.388272 | 0.00307073 | 35.34883811 |
| exposure | chr.exposure | pos.exposure | beta.exposure | se.exposure | pval.exposure | SNP | effect_allele.exposure | other_allele.exposure | eaf.exposure | R2 | F |

| | | | | | | | | | | | |
|---|---|---|---|---|---|---|---|---|---|---|---|
| Free cholesterol levels in very small VLDL \|\| id:ebi-a-GCST90093032 | 1 | 55505647 | -0.294853 | 0.0153928 | 8.80E-82 | rs11591147 | T | G | 0.017457 | 0.003178235 | 366.9173946 |
| | 1 | 25713234 | 0.0227195 | 0.00404013 | 1.90E-08 | rs1293261 | T | A | 0.525176 | 0.00274714 | 31.62272649 |
| | 1 | 234873275 | 0.0258261 | 0.00407273 | 2.30E-10 | rs822928 | C | A | 0.52964 | 0.00349291 | 40.2104441 |
| | 1 | 55518467 | -0.0255312 | 0.00421876 | 1.40E-09 | rs2495477 | G | A | 0.393802 | 0.00318147 | 36.62395726 |
| | 1 | 55526428 | 0.081761 | 0.0113081 | 4.80E-13 | rs11206517 | G | T | 0.033012 | 0.00454055 | 52.27638656 |
| | 1 | 62999675 | 0.0557894 | 0.00422219 | 7.30E-40 | rs7534572 | G | C | 0.64747 | 0.01514824 | 174.5903711 |
| | 1 | 109817590 | -0.0851144 | 0.00486034 | 1.20E-68 | rs12740374 | T | G | 0.221064 | 0.02657722 | 306.6657218 |
| | 2 | 27839369 | 0.0260652 | 0.00456918 | 1.20E-08 | rs4666000 | C | T | 0.270514 | 0.00282693 | 32.54153548 |
| | 2 | 63208460 | 0.0238248 | 0.00432324 | 3.60E-08 | rs2539980 | C | T | 0.682109 | 0.00263826 | 30.36912565 |
| | 2 | 20354039 | -0.0229436 | 0.00404623 | 1.40E-08 | rs11096640 | A | C | 0.483356 | 0.00279314 | 32.15247772 |
| | 2 | 21232195 | 0.0863359 | 0.00404076 | 2.80E-101 | rs693 | A | G | 0.520889 | 0.03951208 | 456.5088285 |

| | | | | | | | | | | | |
|---|---|---|---|---|---|---|---|---|---|---|---|
| | 2 | 21272778 | -0.0913798 | 0.00997738 | 5.30E-20 | rs60403635 | C | T | 0.043024 | 0.00728356 | 83.88027246 |
| | 2 | 44063098 | -0.0759417 | 0.00814649 | 1.10E-20 | rs115445558 | C | G | 0.06589 | 0.00754544 | 86.89844891 |
| | 3 | 135800409 | -0.0420716 | 0.00482435 | 2.80E-18 | rs34330586 | T | A | 0.231198 | 0.00660399 | 76.04889259 |
| | 3 | 12327431 | -0.0254044 | 0.00462744 | 4.00E-08 | rs13076933 | G | T | 0.259653 | 0.00261827 | 30.13899345 |
| | 4 | 87976359 | 0.0277562 | 0.00414642 | 2.20E-11 | rs1471251 | T | A | 0.397121 | 0.00389222 | 44.80906558 |
| | 4 | 69340991 | 0.0305913 | 0.00471544 | 8.70E-11 | rs4860948 | A | T | 0.244338 | 0.00365582 | 42.08660288 |
| | 5 | 74656539 | 0.051826 | 0.00412444 | 3.30E-36 | rs12916 | C | T | 0.400341 | 0.01370132 | 157.8911739 |
| | 5 | 74472939 | 0.0338238 | 0.00420927 | 9.30E-16 | rs7707394 | A | G | 0.357075 | 0.00560764 | 64.56894337 |
| | 5 | 156443066 | 0.0444457 | 0.00421429 | 5.30E-26 | rs4704834 | G | A | 0.644028 | 0.0096557 | 111.2251802 |
| | 6 | 32601288 | 0.0496998 | 0.00523825 | 2.40E-21 | rs72848251 | A | G | 0.194675 | 0.0078161 | 90.01798581 |
| | 6 | 160543148 | 0.0696482 | 0.00794632 | 1.90E-18 | rs12208357 | T | C | 0.07 | 0.00667099 | 76.82103354 |

| | | | | | | | | | | | |
|---|---|---|---|---|---|---|---|---|---|---|---|
| | 6 | 160575366 | 0.0384587 | 0.0053697 | 7.90E-13 | rs11753995 | A | G | 0.171039 | 0.000445542 | 51.29577629 |
| | 6 | 31436032 | 0.0244955 | 0.0043013 | 1.20E-08 | rs2523675 | A | G | 0.342119 | 0.000281737 | 32.43139289 |
| | 7 | 44584551 | 0.0319955 | 0.00521769 | 8.70E-10 | rs10260606 | C | G | 0.182652 | 0.000326642 | 37.60223937 |
| | 7 | 1074134 | -0.0374399 | 0.00483777 | 1.00E-14 | rs869412 | C | T | 0.226812 | 0.00052017 | 59.89235339 |
| | 7 | 21607283 | 0.0275548 | 0.00488269 | 1.70E-08 | rs55649657 | G | C | 0.219849 | 0.000276661 | 31.84701077 |
| | 8 | 126506694 | -0.0735777 | 0.00419505 | 7.20E-69 | rs112875651 | A | G | 0.392379 | 0.002665946 | 307.617179 |
| | 8 | 19830921 | -0.0567327 | 0.00612304 | 1.90E-20 | rs10096633 | T | C | 0.123907 | 0.000745421 | 85.84700829 |
| | 8 | 59370320 | -0.0274887 | 0.0042988 | 1.60E-10 | rs4738679 | A | G | 0.666712 | 0.000355183 | 40.88898925 |
| | 8 | 116658583 | -0.035617 | 0.00451564 | 3.10E-15 | rs2737245 | T | G | 0.278602 | 0.000540299 | 62.21118905 |
| | 9 | 136149711 | 0.0549926 | 0.00522666 | 6.90E-26 | rs115478735 | T | A | 0.183304 | 0.000961026 | 110.7012333 |
| | 11 | 5701074 | 0.0496033 | 0.00787057 | 2.90E-10 | rs11601507 | A | C | 0.069168 | 0.000345026 | 39.71926661 |

| | | | | | | | | | | | |
|---|---|---|---|---|---|---|---|---|---|---|---|
| | 11 | 116648917 | －0.0753614 | 0.00595508 | 1.10E-36 | rs964184 | C | G | 0.86726 | 0.001389669 | 160.1456578 |
| | 11 | 126260684 | 0.043584 | 0.00731207 | 2.50E-09 | rs12804228 | G | A | 0.08472 | 0.00308626 | 35.52760597 |
| | 11 | 61557826 | －0.0574151 | 0.00424653 | 1.20E-41 | rs102274 | C | T | 0.343868 | 0.001585941 | 182.8000501 |
| | 12 | 121424406 | －0.0401141 | 0.00416754 | 6.20E-22 | rs2393776 | A | G | 0.620298 | 0.00080441 | 92.64607059 |
| | 12 | 124854903 | －0.0243496 | 0.00413808 | 4.00E-09 | rs3782257 | A | G | 0.610818 | 0.00300779 | 34.62408857 |
| | 13 | 32976654 | －0.0261338 | 0.00427182 | 9.50E-10 | rs61946991 | C | A | 0.451649 | 0.0032511 | 37.42585143 |
| | 13 | 114551993 | 0.0268767 | 0.00430455 | 4.30E-10 | rs6602909 | C | T | 0.328942 | 0.00338643 | 38.98421793 |
| | 15 | 59243880 | －0.0702042 | 0.0118592 | 3.20E-09 | rs146407081 | A | G | 0.031687 | 0.00304422 | 35.04353067 |
| | 15 | 58725839 | 0.142399 | 0.00445023 | 1.00E-200 | rs633695 | G | A | 0.292354 | 0.008818501 | 1023.861982 |
| | 15 | 58620695 | 0.0407019 | 0.00420764 | 3.90E-22 | rs261274 | C | A | 0.639963 | 0.00812441 | 93.57173196 |
| | 15 | 58678720 | －0.190747 | 0.00425058 | 1.00E-200 | rs261290 | C | T | 0.654645 | 0.017197989 | 2013.777555 |

| | | | | | | | | | | | |
|---|---|---|---|---|---|---|---|---|---|---|---|
| | 15 | 58854753 | -0.0825191 | 0.0151075 | 4.70E-08 | rs117911817 | A | G | 0.0182 | 0.00259181 | 29.83432469 |
| | 16 | 72144174 | -0.0454499 | 0.00521682 | 3.00E-18 | rs9302635 | C | T | 0.182459 | 0.00659114 | 75.9008452 |
| | 16 | 56993886 | -0.0830792 | 0.00431709 | 1.60E-82 | rs821840 | G | A | 0.324526 | 0.003207744 | 370.3350854 |
| | 17 | 45763073 | 0.0289581 | 0.00405403 | 9.10E-13 | rs4794048 | C | A | 0.480798 | 0.00443166 | 51.02213917 |
| | 17 | 64195431 | 0.0729222 | 0.0114215 | 1.70E-10 | rs75003668 | G | A | 0.033354 | 0.00354089 | 40.76295778 |
| | 17 | 67081278 | 0.166821 | 0.0135867 | 1.20E-34 | rs77542162 | G | A | 0.022666 | 0.0130827 | 150.7529851 |
| | 17 | 67191270 | -0.0440509 | 0.00602829 | 2.70E-13 | rs72631343 | G | C | 0.129021 | 0.0046378 | 53.39661802 |
| | 18 | 47109955 | 0.115466 | 0.0176545 | 6.10E-11 | rs77960347 | G | A | 0.013234 | 0.0037156 | 42.77498345 |
| | 19 | 45401868 | 0.0954162 | 0.0103713 | 3.60E-20 | rs118170342 | C | T | 0.040768 | 0.00734939 | 84.63895483 |
| | 19 | 11227480 | 0.0327071 | 0.00411799 | 2.00E-15 | rs2738447 | C | A | 0.59318 | 0.00547858 | 63.0820727 |
| | 19 | 49236102 | 0.0306609 | 0.00405299 | 3.90E-14 | rs439523 | C | T | 0.52969 | 0.00497045 | 57.22834483 |

| | | | | | | | | | | | |
|---|---|---|---|---|---|---|---|---|---|---|---|
| | 19 | 45039971 | −0.0796745 | 0.0132432 | 1.80E-09 | rs74747585 | C | T | 0.025173 | 0.00314419 | 36.19472407 |
| | 19 | 45215989 | −0.0335592 | 0.00598204 | 2.00E-08 | rs58530605 | GT | G | 0.134829 | 0.002734 | 31.47146977 |
| | 19 | 11197598 | −0.146794 | 0.0062147 | 2.40E-123 | rs73015024 | T | G | 0.119667 | 0.04824679 | 557.9158814 |
| | 19 | 19388500 | −0.0960123 | 0.00765173 | 4.10E-36 | rs8107974 | T | A | 0.075552 | 0.0136626 | 157.4442938 |
| | 19 | 45413233 | −0.250461 | 0.00744717 | 1.00E-200 | rs1065853 | T | G | 0.080581 | 0.09732908 | 1131.071703 |
| | 20 | 34415578 | −0.0338104 | 0.00605405 | 2.30E-08 | rs112103450 | C | T | 0.129471 | 0.00270946 | 31.18897219 |
| | 20 | 44545460 | 0.0399047 | 0.00466811 | 1.20E-17 | rs4812975 | A | G | 0.750337 | 0.00634575 | 73.0732444 |
| | 20 | 39179822 | 0.106358 | 0.0118687 | 3.20E-19 | rs1883711 | C | G | 0.031207 | 0.00697306 | 80.30202026 |
| | 22 | 50868669 | 0.0259956 | 0.00416495 | 4.30E-10 | rs9616847 | T | A | 0.388272 | 0.00338396 | 38.95583877 |
| exposure | chr.exposure | pos.exposure | beta.exposure | se.exposure | pval.exposure | SNP | effect_allele.exposure | other_allele.exposure | eaf.exposure | R2 | F |
| Free choles | 1 | 63085997 | −0.077638 | 0.00435066 | 3.20E-71 | rs12239736 | A | T | 0.341234 | 0.02759503 | 318.4423652 |

| | | | | | | | | | | | |
|---|---|---|---|---|---|---|---|---|---|---|---|
| terol levels in VLDL \|\| id: ebi-a-GCST90092998 | 1 | 230295789 | -0.0337841 | 0.00418675 | 7.10E-16 | rs10127775 | T | A | 0.605599 | 0.0056548 | 65.11230986 |
| | 1 | 234872431 | 0.0248758 | 0.00410735 | 1.40E-09 | rs613334 | A | G | 0.503583 | 0.00318629 | 36.67947756 |
| | 1 | 55491853 | -0.0383877 | 0.00549651 | 2.90E-12 | rs200730299 | C | A | 0.194501 | 0.00423661 | 48.77559678 |
| | 1 | 109817590 | -0.0689646 | 0.00492077 | 1.30E-44 | rs12740374 | T | G | 0.221064 | 0.01703877 | 196.4168596 |
| | 1 | 27021913 | 0.0905172 | 0.0137194 | 4.20E-11 | rs114165349 | C | G | 0.022955 | 0.00378112 | 43.52957611 |
| | 2 | 27730940 | -0.0758241 | 0.00417839 | 1.40E-73 | rs1260326 | C | T | 0.603982 | 0.02853305 | 329.297955 |
| | 2 | 21198900 | -0.108594 | 0.00486422 | 2.10E-110 | rs11902417 | A | G | 0.227855 | 0.00431221 | 498.3996114 |
| | 2 | 21530659 | -0.0409284 | 0.00417466 | 1.10E-22 | rs60960031 | A | G | 0.402852 | 0.00834521 | 96.11690225 |
| | 2 | 165528876 | -0.0400262 | 0.00418752 | 1.20E-21 | rs13389219 | T | C | 0.392532 | 0.00793273 | 91.36237611 |
| | 2 | 44072576 | -0.0345972 | 0.0043789 | 2.80E-15 | rs4299376 | T | G | 0.676277 | 0.00542137 | 62.42295187 |
| | 3 | 135798658 | -0.0377822 | 0.00489561 | 1.20E-14 | rs34894639 | T | C | 0.231578 | 0.00517284 | 59.55981103 |

| | | | | | | | | | | | |
|---|---|---|---|---|---|---|---|---|---|---|---|
| | 3 | 52841220 | -0.0380093 | 0.00644849 | 3.80E-09 | rs13059141 | A | G | 0.11318 | 0.000301804 | 34.74210543 |
| | 4 | 87976359 | 0.0317382 | 0.00419807 | 4.00E-14 | rs1471251 | T | A | 0.397121 | 0.00496412 | 57.15545491 |
| | 4 | 3443931 | -0.0319673 | 0.00423493 | 4.40E-14 | rs13108218 | G | A | 0.615355 | 0.00494877 | 56.97861566 |
| | 5 | 55857025 | 0.0453126 | 0.00521252 | 3.50E-18 | rs11429307 | GT | G | 0.191517 | 0.00656221 | 75.56748151 |
| | 5 | 74472939 | 0.0234187 | 0.0042615 | 3.90E-08 | rs7707394 | A | G | 0.357075 | 0.00262349 | 30.19903276 |
| | 5 | 156443066 | 0.0492678 | 0.00426659 | 7.60E-31 | rs4704834 | G | A | 0.644028 | 0.01157322 | 133.3389312 |
| | 5 | 74656539 | 0.0443723 | 0.00417562 | 2.20E-26 | rs12916 | C | T | 0.400341 | 0.00980277 | 112.9209168 |
| | 6 | 32586376 | 0.04833 | 0.00446833 | 2.90E-27 | rs11755689 | G | A | 0.334143 | 0.01015534 | 116.9864503 |
| | 6 | 127455029 | 0.0278143 | 0.0041017 | 1.20E-11 | rs2186037 | G | A | 0.518794 | 0.00399418 | 45.98338479 |
| | 6 | 32434716 | 0.0424421 | 0.00638485 | 3.00E-11 | rs28377109 | A | C | 0.116096 | 0.00383812 | 44.18600968 |
| | 6 | 43757896 | 0.0223295 | 0.00409272 | 4.90E-08 | rs998584 | A | C | 0.482313 | 0.00258591 | 29.76640468 |

| | | | | | | | | | | | |
|---|---|---|---|---|---|---|---|---|---|---|---|
| | 6 | 139834012 | -0.0310194 | 0.00422876 | 2.20E-13 | rs632057 | G | T | 0.627778 | 0.00467337 | 53.80630425 |
| | 6 | 160575366 | 0.0359496 | 0.00543209 | 3.60E-11 | rs11753995 | A | G | 0.171039 | 0.00380436 | 43.79723199 |
| | 6 | 161111700 | -0.281953 | 0.0172465 | 4.50E-60 | rs186696265 | T | C | 0.014352 | 0.02317057 | 267.2661432 |
| | 6 | 160543148 | 0.0676294 | 0.00803864 | 4.00E-17 | rs12208357 | T | C | 0.07 | 0.00614655 | 70.77801226 |
| | 7 | 44584551 | 0.0297373 | 0.00528292 | 1.80E-08 | rs10260606 | C | G | 0.182652 | 0.0027525 | 31.68454336 |
| | 7 | 73037366 | -0.068521 | 0.00509438 | 3.10E-41 | rs55747707 | A | G | 0.203642 | 0.0156955 | 180.9077465 |
| | 7 | 130433594 | 0.0276597 | 0.00441581 | 3.80E-10 | rs11976955 | G | C | 0.314211 | 0.00340815 | 39.2343648 |
| | 8 | 126506694 | -0.0882199 | 0.00424441 | 5.90E-96 | rs112875651 | A | G | 0.392379 | 0.03739933 | 432.0071319 |
| | 8 | 116657911 | -0.0293345 | 0.00455779 | 1.20E-10 | rs2721961 | G | T | 0.28083 | 0.0035982 | 41.42296771 |
| | 8 | 19733244 | -0.0457316 | 0.00498732 | 4.70E-20 | rs1561748 | C | G | 0.267445 | 0.00730086 | 84.07962683 |
| | 8 | 19819724 | -0.149116 | 0.00678914 | 6.40E-107 | rs328 | G | C | 0.100307 | 0.04174408 | 482.4045709 |

| | | | | | | | | | | | |
|---|---|---|---|---|---|---|---|---|---|---|---|
| | 8 | 59398461 | −0.0300281 | 0.00433941 | 4.50E-12 | rs10504255 | A | G | 0.662958 | 0.00415917 | 47.88359936 |
| | 9 | 136155000 | 0.0348395 | 0.00530333 | 5.10E-11 | rs635634 | T | C | 0.183543 | 0.00374866 | 43.15578051 |
| | 10 | 94797973 | −0.0281756 | 0.00417587 | 1.50E-11 | rs2478236 | A | G | 0.403791 | 0.00395433 | 45.52446581 |
| | 11 | 116714293 | 0.222165 | 0.0216108 | 8.70E-25 | rs141469619 | G | A | 0.010141 | 0.00917495 | 105.6822488 |
| | 11 | 117156893 | −0.200137 | 0.0208055 | 6.60E-22 | rs1047964 | T | G | 0.010073 | 0.00803419 | 92.53177836 |
| | 11 | 116648917 | −0.175486 | 0.00602913 | 3.00E-186 | rs964184 | C | G | 0.86726 | 0.07307737 | 847.1651965 |
| | 12 | 124419728 | −0.0290445 | 0.00432278 | 1.80E-11 | rs11057397 | T | C | 0.337502 | 0.00392124 | 45.14337187 |
| | 12 | 9172646 | 0.0236107 | 0.00417062 | 1.50E-08 | rs10842724 | C | T | 0.412903 | 0.00278412 | 32.04859061 |
| | 12 | 57781893 | −0.0297129 | 0.00478512 | 5.30E-10 | rs2122982 | A | G | 0.238672 | 0.00334928 | 38.55643472 |
| | 13 | 114551993 | 0.0318366 | 0.00435846 | 2.80E-13 | rs6602909 | C | T | 0.328942 | 0.00463424 | 53.35556032 |
| | 15 | 58678720 | −0.048548 | 0.00431291 | 2.20E-29 | rs261290 | C | T | 0.654645 | 0.01099807 | 126.7051735 |

| | | | | | | | | | | | |
|---|---|---|---|---|---|---|---|---|---|---|---|
| | 15 | 58725839 | 0.0330184 | 0.00451549 | 2.60E-13 | rs633695 | G | A | 0.292354 | 0.00464401 | 53.46809754 |
| | 15 | 44027885 | 0.103299 | 0.0129025 | 1.20E-15 | rs139974673 | C | T | 0.025912 | 0.00556667 | 64.09688905 |
| | 16 | 56993886 | −0.0800543 | 0.00437241 | 7.00E-75 | rs821840 | G | A | 0.324526 | 0.02904404 | 335.2123598 |
| | 16 | 72217113 | −0.0270963 | 0.00484943 | 2.30E-08 | rs7202323 | G | T | 0.229928 | 0.00271215 | 31.21986687 |
| | 16 | 72108093 | 0.0460302 | 0.0052227 | 1.20E-18 | rs2000999 | A | G | 0.188613 | 0.0067452 | 77.67620803 |
| | 17 | 17431208 | 0.0526184 | 0.0089361 | 3.90E-09 | rs8078513 | A | C | 0.057349 | 0.0030119 | 34.67142474 |
| | 17 | 67191270 | −0.0376709 | 0.00610483 | 6.80E-10 | rs72631343 | G | C | 0.129021 | 0.00330761 | 38.07652402 |
| | 17 | 41840849 | 0.0765032 | 0.0122377 | 4.10E-10 | rs77697917 | T | C | 0.028723 | 0.00339473 | 39.07977459 |
| | 17 | 67081278 | 0.0941753 | 0.0137592 | 7.70E-12 | rs77542162 | G | A | 0.022666 | 0.00406915 | 46.84681937 |
| | 19 | 45360488 | 0.120677 | 0.0202514 | 2.50E-09 | rs375972689 | G | T | 0.010543 | 0.00308459 | 35.5084209 |
| | 19 | 45425460 | 0.0835519 | 0.00429987 | 8.10E-94 | rs157595 | G | A | 0.617355 | 0.03655303 | 422.1954884 |

| | | | | | | | | | | | |
|---|---|---|---|---|---|---|---|---|---|---|---|
| | 19 | 49263477 | −0.0269071 | 0.00428567 | 3.40E-10 | rs2081194 | C | G | 0.601344 | 0.00342405 | 39.41747347 |
| | 19 | 45345787 | −0.0593179 | 0.00853086 | 3.60E-12 | rs111371860 | T | A | 0.063586 | 0.00419949 | 48.34798268 |
| | 19 | 11202306 | −0.10947 | 0.00628816 | 7.10E-68 | rs6511720 | T | G | 0.119788 | 0.02626597 | 303.0648167 |
| | 19 | 19379549 | −0.13003 | 0.00779572 | 1.80E-62 | rs58542926 | T | C | 0.074399 | 0.02411674 | 278.2063979 |
| | 19 | 11227480 | 0.0273941 | 0.00417074 | 5.10E-11 | rs2738447 | C | A | 0.59318 | 0.00374729 | 43.14001498 |
| | 19 | 45239536 | −0.0714207 | 0.00721503 | 4.20E-23 | rs62117162 | A | C | 0.088161 | 0.00850736 | 97.98607797 |
| | 20 | 39179822 | 0.109732 | 0.0120186 | 6.80E-20 | rs1883711 | C | G | 0.031207 | 0.00723831 | 83.35876646 |
| | 22 | 50868669 | 0.0258627 | 0.00421756 | 8.70E-10 | rs9616847 | T | A | 0.388272 | 0.00326645 | 37.60257943 |
| exposure | chr.exposure | pos.exposure | beta.exposure | se.exposure | pval.exposure | SNP | effect_allele.exposure | other_allele.exposure | eaf.exposure | R2 | F |
| Free cholesterol levels | 1 | 220970028 | 0.0372462 | 0.00437657 | 1.70E-17 | rs2642438 | G | A | 0.70385 | 0.00628949 | 72.42502206 |
| | 1 | 55521313 | 0.0358474 | 0.00418241 | 1.00E-17 | rs472495 | T | G | 0.649121 | 0.00637937 | 73.46061631 |

| | | | | | | | | | | | |
|---|---|---|---|---|---|---|---|---|---|---|---|
| in IDL \|\| id: ebi-a-GCST90092835 | 1 | 234851165 | 0.0285091 | 0.00399705 | 9.90E-13 | rs496654 | C | A | 0.51724 | 0.00441864 | 50.8721749 |
| | 1 | 25788425 | -0.0231734 | 0.00402315 | 8.40E-09 | rs35346083 | A | C | 0.44513 | 0.00288214 | 33.17718376 |
| | 1 | 55505647 | -0.326041 | 0.0152253 | 9.79E-102 | rs11591147 | T | G | 0.017457 | 0.003968973 | 458.5695021 |
| | 1 | 55676374 | 0.0747721 | 0.0102613 | 3.20E-13 | rs67276129 | G | T | 0.039899 | 0.00461176 | 53.09661548 |
| | 1 | 109817590 | -0.0998514 | 0.00480746 | 8.10E-96 | rs12740374 | T | G | 0.221064 | 0.003734604 | 431.3893412 |
| | 2 | 227105921 | -0.0276494 | 0.00416948 | 3.30E-11 | rs2943650 | T | C | 0.644754 | 0.00381974 | 43.97441648 |
| | 2 | 44072576 | -0.0466363 | 0.00428113 | 1.20E-27 | rs4299376 | T | G | 0.676277 | 0.01030093 | 118.6653148 |
| | 2 | 21288321 | 0.097882 | 0.00518946 | 2.40E-79 | rs562338 | G | A | 0.819616 | 0.03081864 | 355.7572702 |
| | 5 | 74625487 | 0.0572245 | 0.0041275 | 1.00E-43 | rs7703051 | A | C | 0.374854 | 0.01667469 | 192.2127985 |
| | 5 | 156397673 | 0.0255569 | 0.00414134 | 6.80E-10 | rs6882345 | A | G | 0.632836 | 0.00330814 | 38.08263808 |
| | 5 | 52106278 | -0.0582177 | 0.0100456 | 6.80E-09 | rs1499279 | G | T | 0.041623 | 0.00291759 | 33.58541952 |

| | | | | | | | | | | | |
|---|---|---|---|---|---|---|---|---|---|---|---|
| | 5 | 74472939 | 0.0413579 | 0.00416474 | 3.10E-23 | rs7707394 | A | G | 0.357075 | 0.00856173 | 98.61287528 |
| | 5 | 122924864 | 0.0240631 | 0.00402603 | 2.30E-09 | rs1895233 | T | G | 0.544776 | 0.00310318 | 35.72247909 |
| | 6 | 161103805 | 0.0497733 | 0.0072726 | 7.70E-12 | rs374071816 | A | T | 0.092249 | 0.00406845 | 46.83881731 |
| | 6 | 32601807 | 0.0455951 | 0.00602087 | 3.70E-14 | rs147841808 | G | C | 0.132301 | 0.00498074 | 57.34694632 |
| | 6 | 26267755 | 0.0288873 | 0.00400112 | 5.20E-13 | rs7773004 | G | A | 0.497283 | 0.00452738 | 52.12465604 |
| | 6 | 160564476 | 0.0306638 | 0.00513435 | 2.30E-09 | rs662138 | G | C | 0.185876 | 0.00309841 | 35.66756478 |
| | 7 | 100216773 | -0.0277048 | 0.00473413 | 4.90E-09 | rs10953298 | T | C | 0.235563 | 0.00297504 | 34.24696831 |
| | 7 | 1081442 | -0.0269358 | 0.0040708 | 3.70E-11 | rs10282584 | T | C | 0.410979 | 0.00380301 | 43.78170578 |
| | 7 | 25992323 | 0.0321389 | 0.00546571 | 4.10E-09 | rs10642257 | AAGGCC | A | 0.160158 | 0.00300352 | 34.57492914 |
| | 7 | 72878645 | 0.0543271 | 0.00602564 | 2.00E-19 | rs12056034 | G | A | 0.125693 | 0.0070585 | 81.28663311 |
| | 7 | 44584551 | 0.0292598 | 0.00516118 | 1.40E-08 | rs10260606 | C | G | 0.182652 | 0.002792 | 32.13935432 |

| | | | | | | | | | | | |
|---|---|---|---|---|---|---|---|---|---|---|---|
| | 8 | 19820916 | 0.076917 | 0.00664318 | 5.30E-31 | rs117199990 | T | C | 0.100233 | 0.001163535 | 134.0556388 |
| | 8 | 9187242 | 0.0495572 | 0.00664589 | 8.90E-14 | rs1461729 | G | A | 0.899237 | 0.000482937 | 55.6031859 |
| | 8 | 19733244 | 0.0352613 | 0.00487666 | 4.80E-13 | rs1561748 | C | G | 0.267445 | 0.000454095 | 52.28103014 |
| | 8 | 126506694 | −0.0347058 | 0.00415024 | 6.10E-17 | rs112875651 | A | G | 0.392379 | 0.000607277 | 69.92784558 |
| | 9 | 136149711 | 0.0649358 | 0.00517054 | 3.60E-36 | rs115478735 | T | A | 0.183304 | 0.001368656 | 157.7208245 |
| | 9 | 107661742 | −0.027145 | 0.00453852 | 2.20E-09 | rs2740488 | C | A | 0.265313 | 0.000310748 | 35.77203419 |
| | 10 | 64934548 | 0.0322187 | 0.00400228 | 8.30E-16 | rs10822145 | T | C | 0.476857 | 0.000562794 | 64.80276564 |
| | 10 | 113921825 | −0.036182 | 0.00445229 | 4.40E-16 | rs2792735 | A | G | 0.72034 | 0.000573537 | 66.04055315 |
| | 11 | 126244955 | 0.0420702 | 0.00651112 | 1.00E-10 | rs4307732 | A | G | 0.105933 | 0.000362638 | 41.74747436 |
| | 11 | 116648917 | 0.0447461 | 0.00589009 | 3.00E-14 | rs964184 | C | G | 0.86726 | 0.000501235 | 57.71104901 |
| | 11 | 61557803 | −0.0904323 | 0.00417858 | 7.29E-104 | rs102275 | C | T | 0.34979 | 0.00405339 | 468.3625512 |

| | | | | | | | | | | | |
|---|---|---|---|---|---|---|---|---|---|---|---|
| | 12 | 112007756 | 0.0303093 | 0.004004 | 3.70E-14 | rs653178 | T | C | 0.516036 | 0.000497668 | 57.30019863 |
| | 12 | 121413027 | 0.0284828 | 0.00419143 | 1.10E-11 | rs148608463 | A | G | 0.353374 | 0.000401106 | 46.17781484 |
| | 13 | 32976654 | -0.0238749 | 0.0042262 | 1.60E-08 | rs61946991 | C | A | 0.451649 | 0.00027724 | 31.91357693 |
| | 15 | 44052248 | -0.0474221 | 0.00798935 | 2.90E-09 | rs117519814 | A | G | 0.067423 | 0.000306054 | 35.23149902 |
| | 15 | 58678720 | -0.0802471 | 0.00421214 | 6.40E-81 | rs261290 | C | T | 0.654645 | 0.003143968 | 362.9489553 |
| | 15 | 58725839 | 0.0605145 | 0.00440999 | 7.50E-43 | rs633695 | G | A | 0.292354 | 0.001633527 | 188.2938527 |
| | 16 | 72079657 | -0.0315131 | 0.00507648 | 5.40E-10 | rs77303550 | T | C | 0.192705 | 0.000334737 | 38.53446631 |
| | 16 | 15054534 | 0.030812 | 0.00481082 | 1.50E-10 | rs56015824 | A | T | 0.654366 | 0.000356319 | 41.01984502 |
| | 17 | 41926126 | -0.095145 | 0.0114461 | 9.40E-17 | rs72836561 | T | C | 0.031482 | 0.000600052 | 69.09542963 |
| | 17 | 64210580 | 0.0862571 | 0.01162 | 1.10E-13 | rs1801689 | C | A | 0.030526 | 0.000478588 | 55.10232222 |
| | 17 | 67081278 | 0.156166 | 0.0134429 | 3.40E-31 | rs77542162 | G | A | 0.022666 | 0.001171307 | 134.952051 |

| | | | | | | | | | | | |
|---|---|---|---|---|---|---|---|---|---|---|---|
| | 17 | 67191270 | −0.0361044 | 0.00596449 | 1.40E-09 | rs72631343 | G | C | 0.129021 | 0.00318294 | 36.64089615 |
| | 17 | 45763073 | 0.0322512 | 0.00401112 | 9.00E-16 | rs4794048 | C | A | 0.480798 | 0.00561447 | 64.64767337 |
| | 18 | 47109955 | 0.125466 | 0.0174656 | 6.80E-13 | rs77960347 | G | A | 0.013234 | 0.00448211 | 51.60330743 |
| | 19 | 45254168 | 0.0567827 | 0.00598951 | 2.50E-21 | rs8103315 | A | C | 0.135788 | 0.00780374 | 89.87562886 |
| | 19 | 11229066 | −0.0788182 | 0.0125557 | 3.40E-10 | rs79915079 | A | G | 0.025969 | 0.00342306 | 39.40611466 |
| | 19 | 45173951 | −0.127062 | 0.00925792 | 7.20E-43 | rs28807203 | C | A | 0.049207 | 0.01634131 | 188.3635987 |
| | 19 | 45215081 | −0.115332 | 0.0141891 | 4.40E-16 | rs111278137 | A | G | 0.021614 | 0.00573764 | 66.06666736 |
| | 19 | 45412955 | 0.137465 | 0.0121922 | 1.70E-29 | rs1081105 | C | A | 0.027621 | 0.01103401 | 127.119615 |
| | 19 | 19379549 | −0.070486 | 0.00759974 | 1.80E-20 | rs58542926 | T | C | 0.074399 | 0.00746924 | 86.02025275 |
| | 19 | 45357939 | 0.050351 | 0.00404129 | 1.20E-35 | rs2927468 | G | A | 0.523287 | 0.01347049 | 155.2274848 |
| | 19 | 11147526 | −0.144082 | 0.0191911 | 6.00E-14 | rs73013176 | C | T | 0.011281 | 0.00489553 | 56.36534532 |

| | | | | | | | | | | | |
|---|---|---|---|---|---|---|---|---|---|---|---|
| | 19 | 11197261 | -0.151725 | 0.00613603 | 5.50E-135 | rs12151108 | A | G | 0.119677 | 0.005284826 | 611.4089561 |
| | 19 | 45413233 | -0.493832 | 0.00735294 | 1.00E-200 | rs1065853 | T | G | 0.080581 | 0.037716556 | 4510.543392 |
| | 20 | 17844684 | -0.0274304 | 0.00422119 | 8.10E-11 | rs2618566 | T | G | 0.660294 | 0.00366799 | 42.22667955 |
| | 20 | 44570192 | -0.0386323 | 0.00534313 | 4.80E-13 | rs6073966 | T | C | 0.169019 | 0.00454051 | 52.27597006 |
| exposure | chr.exposure | pos.exposure | beta.exposure | se.exposure | pval.exposure | SNP | effect_allele.exposure | other_allele.exposure | eaf.exposure | R2 | F |
| Free cholesterol levels in large LDL \|\| id:ebi-a-GCST90092860 | 1 | 109817590 | -0.114164 | 0.00487891 | 4.30E-121 | rs12740374 | T | G | 0.221064 | 0.00473526 1 | 547.5265615 |
| | 1 | 220970028 | 0.0310718 | 0.00444161 | 2.60E-12 | rs2642438 | G | A | 0.703885 | 0.00425069 | 48.93779908 |
| | 1 | 234852760 | 0.0319903 | 0.00405583 | 3.10E-15 | rs553427 | T | C | 0.517037 | 0.005403 | 62.2113451 |
| | 1 | 55505647 | -0.323486 | 0.0154516 | 2.50E-97 | rs11591147 | T | G | 0.017457 | 0.03794074 | 438.2849098 |
| | 1 | 55521313 | 0.0373957 | 0.00424456 | 1.20E-18 | rs472495 | T | G | 0.649121 | 0.00674028 | 77.61942701 |
| | 1 | 55676374 | 0.0765397 | 0.0104138 | 2.00E-13 | rs67276129 | G | T | 0.039899 | 0.00469185 | 54.01911917 |

| | | | | | | | | | | | |
|---|---|---|---|---|---|---|---|---|---|---|---|
| | 2 | 44072576 | -0.0487562 | 0.00434446 | 3.20E-29 | rs4299376 | T | G | 0.676277 | 0.001093215 | 125.9448885 |
| | 2 | 21288321 | 0.102512 | 0.00526622 | 2.10E-84 | rs562338 | G | A | 0.819616 | 0.003281832 | 378.9168074 |
| | 2 | 227105921 | -0.024578 | 0.00423116 | 6.30E-09 | rs2943650 | T | C | 0.644754 | 0.00293116 | 33.7416748 |
| | 5 | 156397673 | 0.0255975 | 0.00420319 | 1.10E-09 | rs6882345 | A | G | 0.632836 | 0.00322173 | 37.0876667 |
| | 5 | 74472939 | 0.0402918 | 0.00422694 | 1.50E-21 | rs7707394 | A | G | 0.357075 | 0.00788916 | 90.86018673 |
| | 5 | 122924864 | 0.0251394 | 0.00408615 | 7.60E-10 | rs1895233 | T | G | 0.544776 | 0.00328799 | 37.85067757 |
| | 5 | 52088271 | -0.0658814 | 0.01023 | 1.20E-10 | rs115018790 | A | G | 0.041229 | 0.00360255 | 41.47313104 |
| | 5 | 74651084 | 0.0559306 | 0.00410686 | 3.10E-42 | rs3846662 | G | A | 0.424111 | 0.01609061 | 185.4691204 |
| | 6 | 161108536 | 0.0291694 | 0.00444468 | 5.30E-11 | rs6935921 | T | C | 0.700123 | 0.00374114 | 43.06916446 |
| | 6 | 26123502 | -0.0641466 | 0.00780594 | 2.10E-16 | rs115740542 | C | T | 0.073601 | 0.00586455 | 67.5288446 |
| | 6 | 32608770 | 0.0439386 | 0.00583653 | 5.10E-14 | rs4452681 | T | C | 0.14263 | 0.00492223 | 56.67290022 |

| | | | | | | | | | | | |
|---|---|---|---|---|---|---|---|---|---|---|---|
| | 6 | 160490174 | −0.0248675 | 0.00419957 | 3.20E-09 | rs2065397 | A | G | 0.369923 | 0.00304589 | 35.06283729 |
| | 6 | 3921059 | 0.0246048 | 0.00450377 | 4.70E-08 | rs12198895 | T | G | 0.282199 | 0.00259279 | 29.8455594 |
| | 6 | 31269219 | −0.0492496 | 0.00892743 | 3.50E-08 | rs35192021 | T | C | 0.0635 | 0.00264381 | 30.43301362 |
| | 6 | 34729158 | −0.0413406 | 0.00697354 | 3.10E-09 | rs114863007 | A | G | 0.094525 | 0.00305286 | 35.14304679 |
| | 7 | 44584551 | 0.0286859 | 0.00523855 | 4.40E-08 | rs10260606 | C | G | 0.182652 | 0.00260492 | 29.98521904 |
| | 7 | 100216773 | −0.0281904 | 0.0048051 | 4.40E-09 | rs10953298 | T | C | 0.235563 | 0.00298993 | 34.41835171 |
| | 7 | 25992323 | 0.0364503 | 0.00554764 | 5.00E-11 | rs10642257 | AAGGCC | A | 0.160158 | 0.00374986 | 43.16960996 |
| | 7 | 72878645 | 0.0343869 | 0.00611597 | 1.90E-08 | rs12056034 | G | A | 0.125693 | 0.00274618 | 31.61169702 |
| | 8 | 19855006 | 0.0690266 | 0.00675438 | 1.60E-24 | rs17411113 | G | C | 0.09988 | 0.00906694 | 104.4370115 |
| | 8 | 126506694 | −0.0336399 | 0.00421261 | 1.40E-15 | rs112875651 | A | G | 0.392379 | 0.00553808 | 63.76749193 |
| | 8 | 9187242 | 0.0603964 | 0.00674577 | 3.50E-19 | rs1461729 | G | A | 0.899237 | 0.00696064 | 80.15885381 |

| | | | | | | | | | | | |
|---|---|---|---|---|---|---|---|---|---|---|---|
| | 9 | 107661742 | －0.0339704 | 0.00460594 | 1.60E-13 | rs2740488 | C | A | 0.265313 | 0.000472446 | 54.39477958 |
| | 9 | 136149711 | 0.0634442 | 0.00524734 | 1.20E-33 | rs115478735 | T | A | 0.183304 | 0.001268664 | 146.1833443 |
| | 10 | 64934548 | 0.0276246 | 0.00406223 | 1.00E-11 | rs10822145 | T | C | 0.476857 | 0.000401681 | 46.24400405 |
| | 10 | 113921825 | －0.0295853 | 0.00451897 | 5.90E-11 | rs2792735 | A | G | 0.72034 | 0.000372309 | 42.86131497 |
| | 11 | 61557803 | －0.0689191 | 0.00424146 | 2.30E-59 | rs102275 | C | T | 0.34979 | 0.0022289 | 264.0224738 |
| | 11 | 126244955 | 0.0483323 | 0.0066091 | 2.60E-13 | rs4307732 | A | G | 0.105933 | 0.000464316 | 53.45835406 |
| | 12 | 9084375 | 0.0274294 | 0.00486864 | 1.80E-08 | rs1805776 | C | T | 0.22603 | 0.000275734 | 31.74020609 |
| | 12 | 112007756 | 0.0289256 | 0.0040641 | 1.10E-12 | rs653178 | T | C | 0.516036 | 0.000439985 | 50.65571314 |
| | 13 | 32976654 | －0.0239897 | 0.00428934 | 2.20E-08 | rs61946991 | C | A | 0.451649 | 0.000271734 | 31.27959644 |
| | 14 | 24871926 | 0.023496 | 0.00409687 | 9.70E-09 | rs11621792 | T | C | 0.453813 | 0.000285727 | 32.8909148 |
| | 15 | 44052248 | －0.0477823 | 0.00811128 | 3.80E-09 | rs117519814 | A | G | 0.067423 | 0.000301451 | 34.70146156 |

| | | | | | | | | | | | |
|---|---|---|---|---|---|---|---|---|---|---|---|
| | 15 | 58678512 | 0.0354775 | 0.00445108 | 1.60E-15 | rs10468017 | T | C | 0.295937 | 0.000551732 | 63.52836377 |
| | 15 | 58725839 | 0.0284406 | 0.00447729 | 2.10E-10 | rs633695 | G | A | 0.292354 | 0.0003505 | 40.34962573 |
| | 16 | 72101525 | 0.0338453 | 0.00522456 | 9.30E-11 | rs34042070 | G | C | 0.186978 | 0.000364529 | 41.96524338 |
| | 17 | 64195431 | 0.0688522 | 0.0114697 | 1.90E-09 | rs75003668 | G | A | 0.033354 | 0.000313032 | 36.03495451 |
| | 17 | 7080316 | -0.0315275 | 0.00515927 | 9.90E-10 | rs55714927 | T | C | 0.190994 | 0.00032438 | 37.34177934 |
| | 17 | 45763073 | 0.032865 | 0.00407112 | 6.90E-16 | rs4794048 | C | A | 0.480798 | 0.000565961 | 65.16762866 |
| | 17 | 67191270 | -0.035257 | 0.00605371 | 5.70E-09 | rs72631343 | G | C | 0.129021 | 0.000294654 | 33.9187578 |
| | 17 | 67081278 | 0.132786 | 0.013644 | 2.20E-22 | rs77542162 | G | A | 0.022666 | 0.00082235 | 94.71387563 |
| | 17 | 41926126 | -0.0788612 | 0.0116173 | 1.10E-11 | rs72836561 | T | C | 0.031482 | 0.000400253 | 46.07961118 |
| | 19 | 45215081 | -0.138732 | 0.0143913 | 5.40E-22 | rs111278137 | A | G | 0.021614 | 0.000806855 | 92.92780815 |
| | 19 | 45357939 | 0.0589523 | 0.0040989 | 6.70E-47 | rs2927468 | G | A | 0.523287 | 0.001794236 | 206.8518043 |

| | | | | | | | | | | | |
|---|---|---|---|---|---|---|---|---|---|---|---|
| | 19 | 45413233 | -0.551825 | 0.00745775 | 1.00E-200 | rs1065853 | T | G | 0.080581 | 0.0454145 | 5474.942403 |
| | 19 | 11227480 | 0.0300688 | 0.00412384 | 3.10E-13 | rs2738447 | C | A | 0.59318 | 0.00461765 | 53.16441403 |
| | 19 | 45173951 | -0.142484 | 0.00938988 | 5.20E-52 | rs28807203 | C | A | 0.049207 | 0.0199681 | 230.252612 |
| | 19 | 45254168 | 0.0655819 | 0.00607489 | 3.60E-27 | rs8103315 | A | C | 0.135788 | 0.01011682 | 116.5423054 |
| | 19 | 45412955 | 0.157562 | 0.012366 | 3.50E-37 | rs1081105 | C | A | 0.027621 | 0.01408721 | 162.3442599 |
| | 19 | 11197261 | -0.150123 | 0.00622349 | 1.50E-128 | rs12151108 | A | G | 0.119677 | 0.05030704 | 581.8605719 |
| | 19 | 19379549 | -0.0786471 | 0.00770806 | 1.90E-24 | rs58542926 | T | C | 0.074399 | 0.00903806 | 104.1040693 |
| | 20 | 39179822 | 0.0780362 | 0.0119183 | 5.80E-11 | rs1883711 | C | G | 0.031207 | 0.00372387 | 42.8702536 |
| | 20 | 43042364 | -0.0682823 | 0.0118478 | 8.20E-09 | rs1800961 | T | C | 0.0302 | 0.00288541 | 33.21492731 |
| | 20 | 17844684 | -0.0268914 | 0.00428415 | 3.50E-10 | rs2618566 | T | G | 0.660294 | 0.00342248 | 39.39943022 |
| exposure | chr.exposure | pos.exposure | beta.exposure | se.exposure | pval.exposure | SNP | effect_allele.exposure | other_allele.exposure | eaf.exposure | R2 | F |

| | | | | | | | | | | | |
|---|---|---|---|---|---|---|---|---|---|---|---|
| Free cholesterol levels in medium LDL \|\| id:ebi-a-GCST90092908 | 1 | 55505647 | -0.309686 | 0.0157146 | 1.90E-86 | rs11591147 | T | G | 0.017457 | 0.00336303 | 388.355031 |
| | 1 | 55520408 | 0.0340255 | 0.00432058 | 3.40E-15 | rs553741 | C | G | 0.645268 | 0.00538621 | 62.01794651 |
| | 1 | 109817590 | -0.123837 | 0.00496195 | 1.80E-137 | rs12740374 | T | G | 0.221064 | 0.05383249 | 622.8572586 |
| | 1 | 55526428 | 0.0968757 | 0.0115445 | 4.80E-17 | rs11206517 | G | T | 0.033012 | 0.00611514 | 70.41608954 |
| | 1 | 221032324 | 0.0240553 | 0.00440952 | 4.90E-08 | rs2784259 | T | A | 0.673603 | 0.00258535 | 29.75989358 |
| | 1 | 234852760 | 0.0318296 | 0.00412486 | 1.20E-14 | rs553427 | T | C | 0.517037 | 0.00517144 | 59.54377821 |
| | 2 | 27752463 | -0.0273422 | 0.00430292 | 2.10E-10 | rs11127048 | A | G | 0.614116 | 0.00350736 | 40.37688587 |
| | 2 | 21149771 | -0.0255157 | 0.00435609 | 4.70E-09 | rs140798831 | C | T | 0.656814 | 0.00298047 | 34.30944864 |
| | 2 | 21288321 | 0.103925 | 0.00535524 | 6.80E-84 | rs562338 | G | A | 0.819616 | 0.03261789 | 376.5950791 |
| | 2 | 44072576 | -0.0457193 | 0.0044179 | 4.20E-25 | rs4299376 | T | G | 0.676277 | 0.00929728 | 107.0926917 |
| | 4 | 3452345 | 0.0257149 | 0.00443757 | 6.80E-09 | rs59950280 | A | G | 0.331493 | 0.00291706 | 33.57930462 |

| | | | | | | | | | | | |
|---|---|---|---|---|---|---|---|---|---|---|---|
| | 5 | 131732774 | -0.0301714 | 0.00549416 | 4.00E-08 | rs17689595 | A | G | 0.172909 | 0.00261979 | 30.15648866 |
| | 5 | 156397673 | 0.0281589 | 0.00427404 | 4.40E-11 | rs6882345 | A | G | 0.632836 | 0.00377036 | 43.40568943 |
| | 5 | 52083226 | -0.0766476 | 0.0103898 | 1.60E-13 | rs115421711 | G | A | 0.041298 | 0.00472683 | 54.42208779 |
| | 5 | 74472939 | 0.0363054 | 0.00429819 | 3.00E-17 | rs7707394 | A | G | 0.357075 | 0.00619576 | 71.34502354 |
| | 5 | 74651084 | 0.0523238 | 0.00417608 | 5.20E-36 | rs3846662 | G | A | 0.424111 | 0.01362265 | 156.98327756 |
| | 5 | 122924864 | 0.0242706 | 0.00415503 | 5.20E-09 | rs1895233 | T | G | 0.544776 | 0.00296399 | 34.11969579 |
| | 6 | 26123502 | -0.0670521 | 0.0079376 | 3.00E-17 | rs115740542 | C | T | 0.073601 | 0.00619683 | 71.35736443 |
| | 6 | 16134247 | -0.0229574 | 0.00415437 | 3.30E-08 | rs6920309 | T | C | 0.456849 | 0.00265285 | 30.53708008 |
| | 6 | 160567928 | -0.0329764 | 0.0051886 | 2.10E-10 | rs3798167 | T | G | 0.200093 | 0.0035087 | 40.39229733 |
| | 6 | 34729158 | -0.040208 | 0.00709116 | 1.40E-08 | rs114863007 | A | G | 0.094525 | 0.00279294 | 32.15013683 |
| | 6 | 32608770 | 0.0448685 | 0.00593496 | 4.00E-14 | rs4452681 | T | C | 0.14263 | 0.00496392 | 57.15312236 |

| | | | | | | | | | | | |
|---|---|---|---|---|---|---|---|---|---|---|---|
| | 7 | 100216773 | -0.0285301 | 0.00488641 | 5.30E-09 | rs10953298 | T | C | 0.235563 | 0.00296136 | 34.08939015 |
| | 7 | 25991826 | 0.0344549 | 0.00564719 | 1.10E-09 | rs4722551 | C | T | 0.1581 | 0.00323362 | 37.22456576 |
| | 8 | 126506694 | -0.0423361 | 0.00428415 | 5.00E-23 | rs112875651 | A | G | 0.392379 | 0.00847845 | 97.65282521 |
| | 8 | 19855006 | 0.0399911 | 0.00686909 | 5.80E-09 | rs17411113 | G | C | 0.09988 | 0.00294437 | 33.89383721 |
| | 8 | 9187242 | 0.0577885 | 0.00686033 | 3.70E-17 | rs1461729 | G | A | 0.899237 | 0.00616194 | 70.95537212 |
| | 9 | 107661742 | -0.0284768 | 0.00468344 | 1.20E-09 | rs2740488 | C | A | 0.265313 | 0.00321148 | 36.96961271 |
| | 9 | 136149711 | 0.0644691 | 0.00533564 | 1.30E-33 | rs115478735 | T | A | 0.183304 | 0.01266987 | 145.9898384 |
| | 11 | 61557803 | -0.0529629 | 0.00431343 | 1.20E-34 | rs102275 | C | T | 0.34979 | 0.01308344 | 150.7615026 |
| | 11 | 126228000 | 0.066381 | 0.0079268 | 5.60E-17 | rs59379014 | T | C | 0.072864 | 0.00609004 | 70.12683396 |
| | 12 | 9084375 | 0.0278552 | 0.00495072 | 1.80E-08 | rs1805776 | C | T | 0.22603 | 0.0027501 | 31.6568928 |
| | 12 | 111904371 | 0.0258922 | 0.00413287 | 3.70E-10 | rs4766578 | A | T | 0.503234 | 0.00340941 | 39.24884391 |

| | | | | | | | | | | | |
|---|---|---|---|---|---|---|---|---|---|---|---|
| | 14 | 24871926 | 0.0283451 | 0.00416545 | 1.00E-11 | rs11621792 | T | C | 0.453813 | 0.00402207 | 46.30464807 |
| | 16 | 72101525 | 0.0380449 | 0.00531218 | 8.00E-13 | rs34042070 | G | C | 0.186978 | 0.00445499 | 51.29083642 |
| | 17 | 67191270 | -0.0351031 | 0.00615544 | 1.20E-08 | rs72631343 | G | C | 0.129021 | 0.00282516 | 32.52109725 |
| | 17 | 67081278 | 0.114046 | 0.0138733 | 2.00E-16 | rs77542162 | G | A | 0.022666 | 0.00586865 | 67.57608264 |
| | 17 | 45763073 | 0.034329 | 0.00413954 | 1.10E-16 | rs4794048 | C | A | 0.480798 | 0.00597243 | 68.77183154 |
| | 17 | 7080316 | -0.0407305 | 0.00524597 | 8.20E-15 | rs55714927 | T | C | 0.190994 | 0.00523544 | 60.28098358 |
| | 19 | 11197261 | -0.143948 | 0.00632922 | 1.70E-114 | rs12151108 | A | G | 0.119677 | 0.04474617 | 517.2533824 |
| | 19 | 11227480 | 0.0311392 | 0.0041939 | 1.10E-13 | rs2738447 | C | A | 0.59318 | 0.0047881 | 55.12786995 |
| | 19 | 45215081 | -0.148803 | 0.0146358 | 2.80E-24 | rs111278137 | A | G | 0.021614 | 0.00897415 | 103.3672338 |
| | 19 | 45412955 | 0.166983 | 0.0125761 | 3.10E-40 | rs1081105 | C | A | 0.027621 | 0.01529609 | 176.2970344 |
| | 19 | 45413233 | -0.570466 | 0.00758445 | 1.00E-200 | rs1065853 | T | G | 0.080581 | 0.04685571 | 5657.229507 |

| | | | | | | | | | | | |
|---|---|---|---|---|---|---|---|---|---|---|---|
| | 19 | 19379549 | −0.0849664 | 0.00783901 | 2.30E-27 | rs58542926 | T | C | 0.074399 | 0.01019815 | 117.4801388 |
| | 19 | 45358353 | 0.0611708 | 0.00416882 | 9.50E-49 | rs10410835 | C | T | 0.523103 | 0.01867424 | 215.3051835 |
| | 19 | 45173951 | −0.148865 | 0.0095494 | 8.70E-55 | rs28807203 | C | A | 0.049207 | 0.02107218 | 243.0107498 |
| | 19 | 45254168 | 0.0674583 | 0.00617809 | 9.40E-28 | rs8103315 | A | C | 0.135788 | 0.01034916 | 119.2215533 |
| | 20 | 39678289 | 0.026325 | 0.00413125 | 1.90E-10 | rs6016505 | T | C | 0.549538 | 0.00352706 | 40.60376059 |
| | 20 | 17844684 | −0.0243187 | 0.00435587 | 2.40E-08 | rs2618566 | T | G | 0.660294 | 0.00270773 | 31.16903403 |
| | 20 | 44534651 | 0.0516455 | 0.00502349 | 8.60E-25 | rs6065904 | A | G | 0.214782 | 0.00917588 | 105.6930282 |
| exposure | chr.exposure | pos.exposure | beta.exposure | se.exposure | pval.exposure | SNP | effect_allele.exposure | other_allele.exposure | eaf.exposure | R2 | F |
| Free cholesterol levels in small LDL | 1 | 55505647 | −0.329559 | 0.0156991 | 7.70E-98 | rs11591147 | T | G | 0.017457 | 0.03814605 | 440.6657464 |
| | 1 | 55676374 | 0.0765255 | 0.0105806 | 4.70E-13 | rs67276129 | G | T | 0.039899 | 0.00454346 | 52.30993878 |
| | 1 | 109817590 | −0.135763 | 0.00495705 | 3.80E-165 | rs12740374 | T | G | 0.221064 | 0.00647509 | 750.0819344 |

| | | | | | | | | | | | |
|---|---|---|---|---|---|---|---|---|---|---|---|
| \|\| id: ebi-a-GCST90092960 | 1 | 55521313 | 0.0387768 | 0.00431255 | 2.40E-19 | rs472495 | T | G | 0.649121 | 0.000702042 | 80.84777999 |
| | 1 | 25818398 | -0.0231786 | 0.00414508 | 2.20E-08 | rs34791230 | A | G | 0.444008 | 0.000271633 | 31.26806602 |
| | 1 | 234852760 | 0.0310183 | 0.0041208 | 5.20E-14 | rs553427 | T | C | 0.517037 | 0.000492098 | 56.65853784 |
| | 2 | 44072576 | -0.0463295 | 0.00441367 | 8.90E-26 | rs4299376 | T | G | 0.676277 | 0.000956517 | 110.1813175 |
| | 2 | 21149771 | -0.0275329 | 0.00435193 | 2.50E-10 | rs140798831 | C | T | 0.656814 | 0.000347682 | 40.02511282 |
| | 2 | 21289432 | 0.113576 | 0.0053484 | 4.50E-100 | rs581411 | A | G | 0.819411 | 0.003903189 | 450.9390346 |
| | 5 | 74651084 | 0.05385 | 0.00417276 | 4.20E-38 | rs3846662 | G | A | 0.424111 | 0.001445071 | 166.5394225 |
| | 5 | 52088271 | -0.0804559 | 0.0103942 | 9.90E-15 | rs115018790 | A | G | 0.041229 | 0.000520356 | 59.91369565 |
| | 5 | 131732774 | -0.0323058 | 0.00548979 | 4.00E-09 | rs17689595 | A | G | 0.172909 | 0.000300823 | 34.62916255 |
| | 5 | 74472939 | 0.0377658 | 0.00429477 | 1.50E-18 | rs7707394 | A | G | 0.357075 | 0.000671457 | 77.32323376 |
| | 5 | 156397673 | 0.0289882 | 0.00427064 | 1.10E-11 | rs6882345 | A | G | 0.632836 | 0.000400198 | 46.07326857 |

| | | | | | | | | | | | |
|---|---|---|---|---|---|---|---|---|---|---|---|
| | 6 | 160564476 | 0.0295794 | 0.00529354 | 2.30E-08 | rs662138 | G | C | 0.185876 | 0.00271245 | 31.22329806 |
| | 6 | 32608770 | 0.0432531 | 0.00592967 | 3.00E-13 | rs4452681 | T | C | 0.14263 | 0.00462131 | 53.20664586 |
| | 6 | 34729158 | -0.0391311 | 0.00708483 | 3.30E-08 | rs114863007 | A | G | 0.094525 | 0.00265001 | 30.50546837 |
| | 6 | 161108536 | 0.0295663 | 0.00451561 | 5.80E-11 | rs6935921 | T | C | 0.700123 | 0.00372385 | 42.87000741 |
| | 6 | 16134247 | -0.0246942 | 0.00415066 | 2.70E-09 | rs6920309 | T | C | 0.456849 | 0.00307478 | 35.3955003 |
| | 6 | 26123502 | -0.0707991 | 0.00793052 | 4.40E-19 | rs115740542 | C | T | 0.073601 | 0.00692061 | 79.69748137 |
| | 7 | 25991826 | 0.0413224 | 0.00564277 | 2.40E-13 | rs4722551 | C | T | 0.1581 | 0.00465776 | 53.62642301 |
| | 7 | 100216773 | -0.0286191 | 0.00488258 | 4.60E-09 | rs10953298 | T | C | 0.235563 | 0.00298453 | 34.35624262 |
| | 8 | 9206069 | 0.0238558 | 0.00424324 | 1.90E-08 | rs10102352 | G | A | 0.605685 | 0.00274578 | 31.60714216 |
| | 8 | 19855006 | 0.0531641 | 0.00686301 | 9.40E-15 | rs17411113 | G | C | 0.09988 | 0.00521163 | 60.00675216 |
| | 8 | 126506694 | -0.0296805 | 0.00428036 | 4.10E-12 | rs112875651 | A | G | 0.392379 | 0.00417631 | 48.08108774 |

| | | | | | | | | | | | |
|---|---|---|---|---|---|---|---|---|---|---|---|
| | 8 | 9187595 | 0.0347113 | 0.00514873 | 1.60E-11 | rs7004769 | G | A | 0.799213 | 0.00394787 | 45.45000911 |
| | 9 | 136149711 | 0.0554033 | 0.00533176 | 2.70E-25 | rs115478735 | T | A | 0.183304 | 0.00937379 | 107.9748315 |
| | 11 | 61557803 | -0.063162 | 0.00430954 | 1.20E-48 | rs102275 | C | T | 0.34979 | 0.01863085 | 214.8040012 |
| | 11 | 126232385 | 0.0488945 | 0.00604698 | 6.20E-16 | rs3967200 | T | C | 0.135609 | 0.00567792 | 65.37856841 |
| | 11 | 68562328 | -0.0467457 | 0.00827102 | 1.60E-08 | rs2229738 | T | C | 0.066036 | 0.00277484 | 31.94167199 |
| | 12 | 9084375 | 0.0273772 | 0.00494678 | 3.10E-08 | rs1805776 | C | T | 0.22603 | 0.00266079 | 30.62847101 |
| | 12 | 111904371 | 0.0269464 | 0.00412958 | 6.80E-11 | rs4766578 | A | T | 0.503234 | 0.00369847 | 42.57769917 |
| | 13 | 32976650 | -0.0250384 | 0.00435669 | 9.10E-09 | rs61946987 | T | A | 0.451474 | 0.00286925 | 33.0287609 |
| | 14 | 24871926 | 0.0268682 | 0.00416216 | 1.10E-10 | rs11621792 | T | C | 0.453813 | 0.00361972 | 41.67082079 |
| | 14 | 70924249 | -0.0237423 | 0.00416555 | 1.20E-08 | rs3751521 | G | A | 0.438796 | 0.00282209 | 32.48578199 |
| | 15 | 58723939 | -0.0457734 | 0.00498696 | 4.40E-20 | rs2070895 | A | G | 0.218679 | 0.00731525 | 84.24556125 |

| | | | | | | | | | | | |
|---|---|---|---|---|---|---|---|---|---|---|---|
| | 15 | 58680954 | 0.0391789 | 0.00422953 | 2.00E-20 | rs2043085 | C | T | 0.612685 | 0.00745056 | 85.80502254 |
| | 16 | 72101525 | 0.037223 | 0.00530799 | 2.30E-12 | rs34042070 | G | C | 0.186978 | 0.00427139 | 49.17620437 |
| | 17 | 45763073 | 0.0347836 | 0.00413618 | 4.10E-17 | rs4794048 | C | A | 0.480798 | 0.00614152 | 70.72006451 |
| | 17 | 7080316 | −0.0362902 | 0.00524172 | 4.40E-12 | rs55714927 | T | C | 0.190994 | 0.00416335 | 47.931778 |
| | 17 | 64195431 | 0.0650602 | 0.011653 | 2.40E-08 | rs75003668 | G | A | 0.033354 | 0.00270789 | 31.17078556 |
| | 17 | 67081278 | 0.121145 | 0.0138621 | 2.30E-18 | rs77542162 | G | A | 0.022666 | 0.0066322 | 76.37397346 |
| | 19 | 11197261 | −0.151202 | 0.00631443 | 1.00E-126 | rs12151108 | A | G | 0.119677 | 0.04957708 | 573.3756132 |
| | 19 | 45413233 | −0.661954 | 0.00756672 | 1.00E-200 | rs1065853 | T | G | 0.080581 | 0.62355007 | 7653.018213 |
| | 19 | 19379549 | −0.0740868 | 0.00782069 | 2.70E-21 | rs58542926 | T | C | 0.074399 | 0.00779194 | 89.73956979 |
| | 19 | 45215081 | −0.159437 | 0.0146016 | 9.30E-28 | rs111278137 | A | G | 0.021614 | 0.01034952 | 119.2256674 |
| | 19 | 45254168 | 0.0763296 | 0.00616365 | 3.20E-35 | rs8103315 | A | C | 0.135788 | 0.01330835 | 153.3566052 |

| | | | | | | | | | | | |
|---|---|---|---|---|---|---|---|---|---|---|---|
| | 19 | 45358353 | 0.0692962 | 0.00415908 | 2.50E-62 | rs10410835 | C | T | 0.523103 | 0.002406415 | 277.5982627 |
| | 19 | 11228745 | 0.0351058 | 0.00418965 | 5.30E-17 | rs2569550 | C | T | 0.593326 | 0.00609719 | 70.20931134 |
| | 19 | 45820144 | -0.0778385 | 0.0138747 | 2.00E-08 | rs17875609 | T | C | 0.023904 | 0.00273411 | 31.47271186 |
| | 19 | 45173951 | -0.170561 | 0.00952709 | 1.10E-71 | rs28807203 | C | A | 0.049207 | 0.02777306 | 320.5024722 |
| | 19 | 45412955 | 0.180729 | 0.0125467 | 4.80E-47 | rs1081105 | C | A | 0.027621 | 0.01799728 | 207.4861517 |
| | 20 | 17844684 | -0.0293445 | 0.00435259 | 1.60E-11 | rs2618566 | T | G | 0.660294 | 0.00394802 | 45.45172396 |
| | 20 | 39755154 | 0.0235497 | 0.0041144 | 1.00E-08 | rs6072279 | A | G | 0.474349 | 0.00284595 | 32.76047483 |
| exposure | chr.exposure | pos.exposure | beta.exposure | se.exposure | pval.exposure | SNP | effect_allele.exposure | other_allele.exposure | eaf.exposure | R2 | F |
| Free cholesterol levels in LDL \|\| id: | 1 | 55505647 | -0.324014 | 0.0155667 | 3.20E-96 | rs11591147 | T | G | 0.017457 | 0.03750551 | 433.2383488 |
| | 1 | 55521313 | 0.0375637 | 0.00427619 | 1.60E-18 | rs472495 | T | G | 0.649121 | 0.00670076 | 77.16408168 |
| | 1 | 55676374 | 0.0771585 | 0.0104914 | 1.90E-13 | rs67276129 | G | T | 0.039899 | 0.00469774 | 54.08702744 |

| | | | | | | | | | | | |
|---|---|---|---|---|---|---|---|---|---|---|---|
| ebi-a-GCST90092885 | 1 | 234852760 | 0.0321631 | 0.00408605 | 3.50E-15 | rs553427 | T | C | 0.517037 | 0.00538105 | 61.95850085 |
| | 1 | 109817590 | -0.1202 | 0.00491526 | 4.50E-132 | rs12740374 | T | G | 0.221064 | 0.00516968 | 598.0099318 |
| | 1 | 220970028 | 0.0273623 | 0.00447471 | 9.70E-10 | rs2642438 | G | A | 0.70385 | 0.00324808 | 37.39106745 |
| | 2 | 44072576 | -0.0481814 | 0.00437672 | 3.50E-28 | rs4299376 | T | G | 0.676277 | 0.01051954 | 121.186364 |
| | 2 | 21288321 | 0.105086 | 0.00530533 | 2.60E-87 | rs562338 | G | A | 0.819616 | 0.03397656 | 392.3352857 |
| | 5 | 52088271 | -0.0709669 | 0.010306 | 5.70E-12 | rs115018790 | A | G | 0.041229 | 0.00411856 | 47.41588331 |
| | 5 | 74472939 | 0.0394231 | 0.00425832 | 2.10E-20 | rs7707394 | A | G | 0.357075 | 0.00744208 | 85.70721972 |
| | 5 | 156397673 | 0.0268745 | 0.00423439 | 2.20E-10 | rs6882345 | A | G | 0.632836 | 0.00349897 | 40.28019159 |
| | 5 | 74651084 | 0.0553584 | 0.00413735 | 7.90E-41 | rs3846662 | G | A | 0.424111 | 0.01553245 | 179.0255381 |
| | 5 | 122924864 | 0.0248818 | 0.00411648 | 1.50E-09 | rs1895233 | T | G | 0.544776 | 0.0031737 | 36.53457189 |
| | 6 | 34729158 | -0.0412998 | 0.00702524 | 4.10E-09 | rs114863007 | A | G | 0.094525 | 0.00300217 | 34.55938605 |

| | | | | | | | | | | | |
|---|---|---|---|---|---|---|---|---|---|---|---|
| | 6 | 160567928 | －0.0324745 | 0.00514037 | 2.70E-10 | rs3798167 | T | G | 0.200093 | 0.00346688 | 39.91063502 |
| | 6 | 26123502 | －0.0663069 | 0.00786381 | 3.40E-17 | rs115740542 | C | T | 0.073601 | 0.00617413 | 71.09578879 |
| | 6 | 32608770 | 0.0445377 | 0.0058798 | 3.60E-14 | rs4452681 | T | C | 0.14263 | 0.00498318 | 57.3750287 |
| | 7 | 25991826 | 0.036766 | 0.00559466 | 5.00E-11 | rs4722551 | C | T | 0.1581 | 0.00375124 | 43.18548694 |
| | 7 | 100216773 | －0.0286315 | 0.00484095 | 3.30E-09 | rs10953298 | T | C | 0.235563 | 0.0030387 | 34.97997226 |
| | 8 | 126506694 | －0.0358116 | 0.0042441 | 3.20E-17 | rs112875651 | A | G | 0.392379 | 0.00618301 | 71.19814617 |
| | 8 | 9187242 | 0.0588617 | 0.00679618 | 4.70E-18 | rs1461729 | G | A | 0.899237 | 0.00651397 | 75.01157918 |
| | 8 | 19855006 | 0.0604958 | 0.00680486 | 6.10E-19 | rs1741113 | G | C | 0.09988 | 0.00686288 | 79.03228008 |
| | 9 | 136149711 | 0.0635309 | 0.0052863 | 2.90E-33 | rs115478735 | T | A | 0.183304 | 0.01253471 | 144.4304805 |
| | 9 | 107661742 | －0.031373 | 0.00464013 | 1.40E-11 | rs2740488 | C | A | 0.265313 | 0.00397074 | 45.71347698 |
| | 10 | 64934548 | 0.0238623 | 0.00409243 | 5.50E-09 | rs10822145 | T | C | 0.476857 | 0.00295343 | 33.9980913 |

| | | | | | | | | | | | |
|---|---|---|---|---|---|---|---|---|---|---|---|
| | 11 | 61557803 | -0.0648547 | 0.0042731 | 5.00E-52 | rs102275 | C | T | 0.34979 | 0.001997657 | 230.3505476 |
| | 11 | 126244955 | 0.0512831 | 0.0066584 | 1.30E-14 | rs4307732 | A | G | 0.105933 | 0.000515202 | 59.32001182 |
| | 12 | 111904371 | 0.0282182 | 0.00409458 | 5.50E-12 | rs4766578 | A | T | 0.503234 | 0.000412528 | 47.49329982 |
| | 12 | 9084375 | 0.0278404 | 0.00490486 | 1.40E-08 | rs1805776 | C | T | 0.22603 | 0.000279878 | 32.21737575 |
| | 13 | 32976654 | -0.0239543 | 0.00432109 | 3.00E-08 | rs61946991 | C | A | 0.451649 | 0.000266967 | 30.73072435 |
| | 14 | 24871926 | 0.0253677 | 0.00412714 | 7.90E-10 | rs11621792 | T | C | 0.453813 | 0.000328181 | 37.77950263 |
| | 15 | 44052248 | -0.0457782 | 0.00817183 | 2.10E-08 | rs117519814 | A | G | 0.067423 | 0.000272617 | 31.38132163 |
| | 16 | 72101525 | 0.0356378 | 0.00526327 | 1.30E-11 | rs34042070 | G | C | 0.186978 | 0.000398226 | 45.84615581 |
| | 17 | 41926126 | -0.0691002 | 0.0117034 | 3.50E-09 | rs72836561 | T | C | 0.031482 | 0.000302828 | 34.85996637 |
| | 17 | 67191270 | -0.0354174 | 0.00609858 | 6.30E-09 | rs72631343 | G | C | 0.129021 | 0.000292982 | 33.72627328 |
| | 17 | 45763073 | 0.0383779 | 0.0041013 | 1.60E-16 | rs4794048 | C | A | 0.480798 | 0.000591152 | 68.07006755 |

| | | | | | | | | | | | |
|---|---|---|---|---|---|---|---|---|---|---|---|
| | 17 | 67081278 | 0.127918 | 0.0137452 | 1.30E-20 | rs77542162 | G | A | 0.022666 | 0.00752016 | 86.60712991 |
| | 17 | 7080316 | -0.0347493 | 0.00519751 | 2.30E-11 | rs55714927 | T | C | 0.190994 | 0.00388262 | 44.69859848 |
| | 17 | 64195431 | 0.0661348 | 0.0115547 | 1.00E-08 | rs75003668 | G | A | 0.033354 | 0.00284585 | 32.75934374 |
| | 19 | 11197261 | -0.150077 | 0.00626849 | 1.10E-126 | rs12151108 | A | G | 0.119677 | 0.00495606 | 573.1850533 |
| | 19 | 45254168 | 0.0679259 | 0.00611881 | 1.20E-28 | rs8103315 | A | C | 0.135788 | 0.01069707 | 123.2336522 |
| | 19 | 45357939 | 0.0612756 | 0.00412853 | 7.80E-50 | rs2927468 | G | A | 0.523287 | 0.01910497 | 220.2808318 |
| | 19 | 45413233 | -0.575069 | 0.00751167 | 1.00E-200 | rs1065853 | T | G | 0.080581 | 0.04846035 | 5860.833252 |
| | 19 | 19379549 | -0.0806667 | 0.00776379 | 2.80E-25 | rs58542926 | T | C | 0.074399 | 0.00937187 | 107.9526914 |
| | 19 | 45173951 | -0.148846 | 0.00945777 | 8.30E-56 | rs28807203 | C | A | 0.049207 | 0.02147612 | 247.6790607 |
| | 19 | 45215081 | -0.145173 | 0.0144954 | 1.30E-23 | rs111278137 | A | G | 0.021614 | 0.00870814 | 100.3006499 |
| | 19 | 11228745 | 0.0312146 | 0.00415917 | 6.10E-14 | rs2569550 | C | T | 0.593326 | 0.00489195 | 56.32415133 |

| | | | | | | | | | | | |
|---|---|---|---|---|---|---|---|---|---|---|---|
| | 19 | 45412955 | 0.164097 | 0.0124554 | 1.20E-39 | rs1081105 | C | A | 0.027621 | 0.0015059 97 | 173.571484 |
| | 20 | 39179822 | 0.0791582 | 0.0120064 | 4.30E-11 | rs1883711 | C | G | 0.031207 | 0.00377568 | 43.46689885 |
| | 20 | 17844684 | -0.0267168 | 0.00431585 | 6.00E-10 | rs2618566 | T | G | 0.660294 | 0.00332877 | 38.32027763 |
| exposure | chr.exposure | pos.exposure | beta.exposure | se.exposure | pval.exposure | SNP | effect_allele.exposure | other_allele.exposure | eaf.exposure | R2 | F |
| Total lipid levels in chylomicrons and extremely large VLDL \|\| id:ebi-a-GCST90093046 | 1 | 230302838 | -0.0384907 | 0.00412223 | 9.90E-21 | rs59484402 | CT | C | 0.611688 | 0.00757026 | 87.18457039 |
| | 1 | 27284913 | 0.0981809 | 0.0131929 | 9.90E-14 | rs79598313 | T | C | 0.023201 | 0.00481013 | 55.38165488 |
| | 1 | 63060037 | -0.0750931 | 0.00415681 | 6.00E-73 | rs79439217 | TAAGAA | T | 0.352737 | 0.0282776 | 326.3413987 |
| | 1 | 149866868 | -0.0794082 | 0.0141495 | 2.00E-08 | rs74910819 | G | T | 0.029891 | 0.00273604 | 31.4949533 |
| | 2 | 21231524 | -0.0833392 | 0.00489666 | 5.90E-65 | rs676210 | A | G | 0.205766 | 0.02510728 | 289.661788 |
| | 2 | 27730940 | -0.0830804 | 0.00405877 | 4.00E-93 | rs1260326 | C | T | 0.603982 | 0.03627626 | 418.9871722 |
| | 2 | 227077377 | 0.0394207 | 0.00425687 | 2.00E-20 | rs2943635 | T | C | 0.679832 | 0.00744624 | 85.75517569 |

| | | | | | | | | | | | |
|---|---|---|---|---|---|---|---|---|---|---|---|
| | 2 | 165528876 | -0.0500596 | 0.00406764 | 8.30E-35 | rs13389219 | T | C | 0.392532 | 0.00131435 | 151.4545007 |
| | 3 | 135956305 | 0.0354796 | 0.00470267 | 4.50E-14 | rs684773 | C | A | 0.767075 | 0.00494363 | 56.91947346 |
| | 4 | 3443931 | -0.0273969 | 0.0041134 | 2.70E-11 | rs13108218 | G | A | 0.615355 | 0.00385324 | 44.36019167 |
| | 4 | 87976359 | 0.0322078 | 0.00407761 | 2.80E-15 | rs1471251 | T | A | 0.397121 | 0.00541836 | 62.38830728 |
| | 5 | 156443066 | 0.035845 | 0.00414512 | 5.30E-18 | rs4704834 | G | A | 0.644028 | 0.00649372 | 74.77826876 |
| | 5 | 55857025 | 0.0522913 | 0.00506412 | 5.40E-25 | rs11429307 | GT | G | 0.191517 | 0.00925638 | 106.6211478 |
| | 6 | 139834012 | -0.0317655 | 0.00410003 | 9.40E-15 | rs632057 | G | T | 0.627778 | 0.00521319 | 60.02466912 |
| | 6 | 160578069 | -0.130338 | 0.0171725 | 3.20E-14 | rs146534110 | T | G | 0.013508 | 0.00500323 | 57.6059718 |
| | 6 | 160711566 | 0.0260609 | 0.00456155 | 1.10E-08 | rs12179053 | T | C | 0.252677 | 0.00283546 | 32.63971761 |
| | 6 | 161080457 | 0.0857829 | 0.0139268 | 7.30E-10 | rs117857195 | T | G | 0.02424 | 0.0032957 | 37.93946678 |
| | 6 | 32623528 | 0.0349862 | 0.00585751 | 2.30E-09 | rs73732214 | A | G | 0.140436 | 0.00309903 | 35.67466667 |

| | | | | | | | | | | | |
|---|---|---|---|---|---|---|---|---|---|---|---|
| | 6 | 43757896 | 0.0256583 | 0.00396813 | 1.00E-10 | rs998584 | A | C | 0.482313 | 0.000363177 | 41.80963984 |
| | 6 | 160710851 | −0.0587003 | 0.00545001 | 4.70E-27 | rs3127580 | T | C | 0.156213 | 0.001007027 | 116.0054774 |
| | 6 | 160922870 | −0.389421 | 0.0147029 | 1.40E-154 | rs117733303 | G | A | 0.0185 | 0.006058792 | 701.4959707 |
| | 6 | 160093887 | −0.0975343 | 0.0157424 | 5.80E-10 | rs184089815 | A | G | 0.016433 | 0.000333442 | 38.38533266 |
| | 6 | 161017363 | −0.133032 | 0.0110346 | 1.80E-33 | rs73596816 | A | G | 0.033593 | 0.001261373 | 145.3421243 |
| | 6 | 127451665 | 0.0296246 | 0.00399792 | 1.30E-13 | rs1936806 | T | C | 0.448453 | 0.000476894 | 54.9071932 |
| | 6 | 161010118 | −0.261294 | 0.00734692 | 1.00E-200 | rs10455872 | G | A | 0.078987 | 0.010871601 | 1264.854893 |
| | 6 | 32586831 | 0.0350026 | 0.00427193 | 2.50E-16 | rs13191810 | T | C | 0.312309 | 0.000583031 | 67.13435287 |
| | 7 | 25983400 | −0.0320645 | 0.00415093 | 1.10E-14 | rs56001710 | T | A | 0.580583 | 0.000518234 | 59.66924975 |
| | 7 | 130432481 | 0.0285762 | 0.00436835 | 6.10E-11 | rs6971365 | C | T | 0.292826 | 0.000371711 | 42.79239078 |
| | 7 | 73123473 | 0.0586758 | 0.00922268 | 2.00E-10 | rs72555385 | G | A | 0.049304 | 0.000351596 | 40.47587176 |

| | | | | | | | | | | | |
|---|---|---|---|---|---|---|---|---|---|---|---|
| | 7 | 73020337 | −0.104757 | 0.00591615 | 3.70E-70 | rs3812316 | G | C | 0.129282 | 0.002717057 | 313.5307955 |
| | 8 | 10649767 | 0.0346621 | 0.00410171 | 2.90E-17 | rs752446 | A | G | 0.375244 | 0.000620159 | 71.41218001 |
| | 8 | 19733244 | −0.0599132 | 0.00484119 | 3.50E-35 | rs1561748 | C | G | 0.267445 | 0.001329094 | 153.1557229 |
| | 8 | 126500031 | −0.0741421 | 0.00406883 | 3.50E-74 | rs28601761 | G | C | 0.420159 | 0.002876948 | 332.0344363 |
| | 8 | 126631799 | 0.031717 | 0.00487717 | 7.90E-11 | rs3860847 | A | G | 0.212508 | 0.000367351 | 42.29031006 |
| | 8 | 19819724 | −0.178378 | 0.00659022 | 2.40E-161 | rs328 | G | C | 0.100307 | 0.006325857 | 732.6139892 |
| | 10 | 65160321 | −0.0372663 | 0.00397658 | 7.20E-21 | rs10761752 | C | T | 0.474418 | 0.000762561 | 87.82245153 |
| | 10 | 94796504 | −0.0252752 | 0.00409168 | 6.50E-10 | rs835273 | T | A | 0.38487 | 0.000331463 | 38.15736105 |
| | 11 | 116916060 | 0.269484 | 0.0201086 | 5.90E-41 | rs144018203 | C | G | 0.010653 | 0.00155818 | 179.5952111 |
| | 11 | 64021821 | 0.0412711 | 0.00702628 | 4.30E-09 | rs2302263 | T | C | 0.08812 | 0.000299712 | 34.50115512 |
| | 11 | 68562328 | 0.0475875 | 0.00797192 | 2.40E-09 | rs2229738 | T | C | 0.066036 | 0.000309541 | 35.6329984 |

| | | | | | | | | | | | |
|---|---|---|---|---|---|---|---|---|---|---|---|
| | 11 | 116648917 | -0.193744 | 0.00585502 | 1.00E-200 | rs964184 | C | G | 0.86726 | 0.009424965 | 1094.944762 |
| | 11 | 117156893 | -0.249809 | 0.0202046 | 4.10E-35 | rs1047964 | T | G | 0.010073 | 0.001326575 | 152.8650139 |
| | 11 | 61575158 | 0.0664937 | 0.00417607 | 4.40E-57 | rs174553 | G | A | 0.342558 | 0.002198175 | 253.5233192 |
| | 11 | 116389017 | 0.0249366 | 0.00433636 | 8.90E-09 | rs883863 | A | G | 0.308012 | 0.000287271 | 33.06863518 |
| | 11 | 27631005 | 0.0260454 | 0.00475668 | 4.40E-08 | rs141078087 | AT | A | 0.227747 | 0.000260456 | 29.98103657 |
| | 12 | 110027795 | 0.0238421 | 0.00414715 | 9.00E-09 | rs11067376 | G | A | 0.358037 | 0.000287116 | 33.05079992 |
| | 12 | 57781893 | -0.025952 | 0.00464822 | 2.40E-08 | rs2122982 | A | G | 0.238672 | 0.000270797 | 31.17172006 |
| | 12 | 124409502 | -0.0236755 | 0.0042613 | 2.80E-08 | rs7133378 | A | G | 0.319789 | 0.000268158 | 30.8678621 |
| | 13 | 114623821 | -0.0359554 | 0.00642732 | 2.20E-08 | rs41284486 | A | G | 0.111972 | 0.000271859 | 31.29398397 |
| | 13 | 95231825 | 0.0293788 | 0.00415647 | 1.60E-12 | rs2275647 | A | G | 0.351884 | 0.000433933 | 49.95871477 |
| | 13 | 114544024 | 0.0283276 | 0.00434128 | 6.80E-11 | rs7140110 | C | T | 0.299365 | 0.000369842 | 42.57713358 |

| | | | | | | | | | | | |
|---|---|---|---|---|---|---|---|---|---|---|---|
| | 15 | 58723479 | −0.040217 | 0.00481462 | 6.60E-17 | rs1077834 | C | T | 0.218646 | 0.000605933 | 69.77305895 |
| | 15 | 58671765 | 0.0296918 | 0.0042122 | 1.80E-12 | rs1601935 | T | G | 0.654638 | 0.000431579 | 49.68754483 |
| | 15 | 44027885 | 0.126954 | 0.0125345 | 4.10E-24 | rs139974673 | C | T | 0.025912 | 0.00890603 | 102.5820092 |
| | 16 | 15172118 | −0.0294895 | 0.00436709 | 1.50E-11 | rs11644601 | C | T | 0.296695 | 0.00396069 | 45.59770515 |
| | 16 | 57004889 | −0.0311075 | 0.00401133 | 8.80E-15 | rs7205804 | A | G | 0.440369 | 0.00522299 | 60.13757186 |
| | 16 | 81534790 | −0.0294896 | 0.00433185 | 9.90E-12 | rs2925979 | C | T | 0.698949 | 0.0040254 | 46.34292019 |
| | 17 | 17432456 | 0.0461545 | 0.00822167 | 2.00E-08 | rs16961845 | T | C | 0.062316 | 0.00273767 | 31.51377799 |
| | 17 | 41926126 | 0.106939 | 0.011382 | 5.70E-21 | rs72836561 | T | C | 0.031482 | 0.00766469 | 88.27292694 |
| | 17 | 64210580 | −0.126517 | 0.0115549 | 6.70E-28 | rs1801689 | C | A | 0.030526 | 0.01040653 | 119.8830657 |
| | 19 | 19393890 | −0.0945998 | 0.00765721 | 4.60E-35 | rs200210321 | AG | A | 0.073166 | 0.01324513 | 152.6271424 |
| | 19 | 45401783 | 0.0422777 | 0.00429663 | 7.60E-23 | rs56290633 | C | T | 0.539526 | 0.00840609 | 96.81869952 |

| | | | | | | | | | | | |
|---|---|---|---|---|---|---|---|---|---|---|---|
| | 19 | 8429323 | -0.20664 | 0.0142502 | 1.20E-47 | rs116843064 | A | G | 0.019851 | 0.0182384 | 210.2709706 |
| | 19 | 8439538 | -0.0503297 | 0.00862994 | 5.50E-09 | rs57192995 | C | G | 0.056297 | 0.00295459 | 34.01148413 |
| | 19 | 45413233 | 0.166217 | 0.00732898 | 7.10E-114 | rs1065853 | T | G | 0.080581 | 0.04449587 | 514.3470561 |
| | 19 | 45470298 | 0.0238447 | 0.00398912 | 2.30E-09 | rs11878597 | C | T | 0.45019 | 0.00310375 | 35.72909173 |
| exposure | chr.exposure | pos.exposure | beta.exposure | se.exposure | pval.exposure | SNP | effect_allele.exposure | other_allele.exposure | eaf.exposure | R2 | F |
| Total lipid levels in very large VLDL \|\| id:ebi-a-GCST90093022 | 1 | 27284913 | 0.107112 | 0.0131708 | 4.20E-16 | rs79598313 | T | C | 0.023201 | 0.00574374 | 66.13698602 |
| | 1 | 63060037 | -0.0746659 | 0.00414985 | 2.20E-72 | rs79439217 | TAAGAA | T | 0.352737 | 0.02805126 | 323.7220358 |
| | 1 | 109615481 | -0.0329224 | 0.00586711 | 2.00E-08 | rs72986845 | C | A | 0.135554 | 0.00273559 | 31.48974741 |
| | 1 | 149866868 | -0.0783609 | 0.0141258 | 2.90E-08 | rs74910819 | G | T | 0.029891 | 0.00267331 | 30.77266982 |
| | 1 | 230302838 | -0.0450444 | 0.00411533 | 7.00E-28 | rs59484402 | CT | C | 0.611688 | 0.01039951 | 119.8021833 |
| | 1 | 219618564 | 0.0302961 | 0.00540609 | 2.10E-08 | rs61838776 | A | C | 0.161109 | 0.00272823 | 31.4050407 |

| | | | | | | | | | | | |
|---|---|---|---|---|---|---|---|---|---|---|---|
| | 2 | 227104111 | 0.0395435 | 0.00414948 | 1.60E-21 | rs2972140 | C | T | 0.650637 | 0.00078852 | 90.81452449 |
| | 2 | 21231524 | -0.0985067 | 0.00488817 | 2.60E-90 | rs676210 | A | G | 0.205766 | 0.003516428 | 406.0985047 |
| | 2 | 165528876 | -0.0506342 | 0.00406058 | 1.10E-35 | rs13389219 | T | C | 0.392532 | 0.001349329 | 155.4906272 |
| | 2 | 21530659 | -0.0243413 | 0.00404811 | 1.80E-09 | rs60960031 | A | G | 0.402852 | 0.000314079 | 36.15558393 |
| | 2 | 27730940 | -0.0908106 | 0.00405173 | 3.00E-111 | rs1260326 | C | T | 0.603982 | 0.004346034 | 502.3247174 |
| | 2 | 219699999 | 0.0641502 | 0.0102631 | 4.10E-10 | rs78058190 | A | G | 0.050108 | 0.000339378 | 39.06891593 |
| | 3 | 52838654 | -0.0366885 | 0.00608215 | 1.60E-09 | rs9968117 | T | C | 0.120935 | 0.000316083 | 36.38631804 |
| | 3 | 135956305 | 0.0353714 | 0.00469666 | 5.00E-14 | rs684773 | C | A | 0.767075 | 0.000492612 | 56.71771259 |
| | 4 | 171563776 | -0.0266458 | 0.00466759 | 1.10E-08 | rs4491981 | A | C | 0.241218 | 0.000283101 | 32.58851557 |
| | 4 | 3443931 | -0.0336918 | 0.00410745 | 2.40E-16 | rs13108218 | G | A | 0.615355 | 0.000584309 | 67.2815998 |
| | 4 | 87976359 | 0.0315794 | 0.0040717 | 8.80E-15 | rs1471251 | T | A | 0.397121 | 0.000522422 | 60.15180403 |

| | | | | | | | | | | | |
|---|---|---|---|---|---|---|---|---|---|---|---|
| | 5 | 156443066 | 0.036477 | 0.00413911 | 1.20E-18 | rs4704834 | G | A | 0.644028 | 0.00067441 | 77.66346129 |
| | 5 | 55857025 | 0.0570556 | 0.00505678 | 1.60E-29 | rs11429307 | GT | G | 0.191517 | 0.01104996 | 127.3036591 |
| | 6 | 161017363 | -0.106367 | 0.0110283 | 5.20E-22 | rs73596816 | A | G | 0.033593 | 0.00807678 | 93.02274054 |
| | 6 | 127455029 | 0.0332724 | 0.00397455 | 5.70E-17 | rs2186037 | G | A | 0.518794 | 0.00608584 | 70.07849715 |
| | 6 | 160710851 | -0.0305509 | 0.00544688 | 2.00E-08 | rs3127580 | T | C | 0.156213 | 0.00273292 | 31.45899575 |
| | 6 | 32586831 | 0.0414982 | 0.00426947 | 2.50E-22 | rs13191810 | T | C | 0.312309 | 0.00820251 | 94.47198458 |
| | 6 | 139834012 | -0.0354873 | 0.00409768 | 4.70E-18 | rs632057 | G | T | 0.627778 | 0.00651298 | 75.00017697 |
| | 6 | 160922870 | -0.310277 | 0.0146944 | 5.80E-99 | rs117733303 | G | A | 0.0185 | 0.03859302 | 445.8491103 |
| | 6 | 43757896 | 0.0323467 | 0.00396585 | 3.50E-16 | rs998584 | A | C | 0.482313 | 0.00577735 | 66.5242284 |
| | 6 | 160545046 | 0.0590403 | 0.0097714 | 1.50E-09 | rs111909590 | T | C | 0.043584 | 0.00317131 | 36.50698605 |
| | 6 | 31236679 | 0.0251223 | 0.00451855 | 2.70E-08 | rs1130580 | C | T | 0.289865 | 0.00268532 | 30.91100054 |

| | | | | | | | | | | | |
|---|---|---|---|---|---|---|---|---|---|---|---|
| | 6 | 32623528 | 0.0353373 | 0.00585415 | 1.60E-09 | rs73732214 | A | G | 0.140436 | 0.00031651 5 | 36.4360666 |
| | 6 | 161010118 | −0.189654 | 0.0073427 | 4.20E-147 | rs10455872 | G | A | 0.078987 | 0.005763611 | 667.1214377 |
| | 7 | 25983400 | −0.0279925 | 0.00414486 | 1.40E-11 | rs56001710 | T | A | 0.580583 | 0.00396172 | 45.6095822 |
| | 7 | 73020337 | −0.114006 | 0.0059075 | 5.50E-83 | rs3812316 | G | C | 0.129282 | 0.003225799 | 372.4263566 |
| | 7 | 73123473 | 0.0693554 | 0.0092092 | 5.00E-14 | rs72555385 | G | A | 0.049304 | 0.00492601 | 56.71647151 |
| | 7 | 130433594 | 0.0341088 | 0.00428254 | 1.70E-15 | rs11976955 | G | C | 0.314211 | 0.00550913 | 63.43405542 |
| | 8 | 10649767 | 0.0320778 | 0.00409447 | 4.70E-15 | rs752446 | A | G | 0.375244 | 0.00533058 | 61.37707902 |
| | 8 | 22882293 | −0.0236684 | 0.0042712 | 3.00E-08 | rs59296513 | A | G | 0.320415 | 0.00266756 | 30.70650848 |
| | 8 | 19733244 | −0.0640336 | 0.00483265 | 4.50E-40 | rs1561748 | C | G | 0.267445 | 0.01523266 | 175.5648639 |
| | 8 | 19819724 | −0.190391 | 0.00657859 | 3.60E-184 | rs328 | G | C | 0.100307 | 0.00722554 1 | 837.5671606 |
| | 8 | 126631799 | 0.0347222 | 0.00486856 | 6.20E-12 | rs3860847 | A | G | 0.212508 | 0.00410565 | 47.26720417 |

| | | | | | | | | | | | |
|---|---|---|---|---|---|---|---|---|---|---|---|
| | 8 | 126500031 | -0.0826333 | 0.00406166 | 5.20E-92 | rs28601761 | G | C | 0.420159 | 0.00358374 | 413.9000836 |
| | 8 | 59402570 | -0.0257451 | 0.0042409 | 1.30E-09 | rs7018333 | G | A | 0.671955 | 0.00320133 | 36.85238118 |
| | 10 | 64882300 | -0.0352811 | 0.00399904 | 1.10E-18 | rs10761716 | G | C | 0.44151 | 0.00675883 | 77.83325452 |
| | 10 | 94797973 | -0.0273838 | 0.00405062 | 1.40E-11 | rs2478236 | A | G | 0.403791 | 0.00396976 | 45.70217568 |
| | 11 | 64021821 | 0.0406463 | 0.00701635 | 6.90E-09 | rs2302263 | T | C | 0.08812 | 0.00291532 | 33.55923087 |
| | 11 | 116916060 | 0.269353 | 0.0200802 | 5.00E-41 | rs144018203 | C | G | 0.010653 | 0.01561067 | 179.9285243 |
| | 11 | 61575158 | 0.0514948 | 0.00417017 | 5.00E-35 | rs174553 | G | A | 0.342558 | 0.01323234 | 152.4795791 |
| | 11 | 117156893 | -0.250225 | 0.0201761 | 2.50E-35 | rs1047964 | T | G | 0.010073 | 0.01334749 | 153.8081699 |
| | 11 | 116648917 | -0.207732 | 0.00584675 | 1.00E-200 | rs964184 | C | G | 0.86726 | 0.10850069 | 1262.322167 |
| | 11 | 116389017 | 0.0251832 | 0.00433024 | 6.00E-09 | rs883863 | A | G | 0.308012 | 0.00293807 | 33.82130389 |
| | 12 | 57766392 | -0.0303751 | 0.00463832 | 5.80E-11 | rs4760254 | C | G | 0.239066 | 0.00372516 | 42.88510424 |

| | | | | | | | | | | | |
|---|---|---|---|---|---|---|---|---|---|---|---|
| | 12 | 4384844 | -0.0845171 | 0.0154346 | 4.40E-08 | rs76895963 | G | T | 0.020556 | 0.00260482 | 29.98408969 |
| | 12 | 124409502 | -0.0316639 | 0.00425477 | 9.90E-14 | rs7133378 | A | G | 0.319789 | 0.00481017 | 55.38205749 |
| | 13 | 114544024 | 0.0280994 | 0.00433549 | 9.10E-11 | rs7140110 | C | T | 0.299365 | 0.00364881 | 42.00588779 |
| | 13 | 95231825 | 0.0235752 | 0.00415093 | 1.40E-08 | rs2275647 | A | G | 0.351884 | 0.00280215 | 32.25614069 |
| | 15 | 43726625 | 0.130748 | 0.0125599 | 2.20E-25 | rs150844304 | C | A | 0.025671 | 0.00940766 | 108.3652901 |
| | 16 | 81534790 | -0.0339691 | 0.00432508 | 4.00E-15 | rs2925979 | C | T | 0.698949 | 0.00535722 | 61.68395954 |
| | 16 | 72096227 | 0.042193 | 0.00654969 | 1.20E-10 | rs34682685 | A | G | 0.103441 | 0.00360475 | 41.49844022 |
| | 16 | 56993886 | -0.061208 | 0.00424315 | 3.60E-47 | rs821840 | G | A | 0.324526 | 0.01804877 | 208.0808248 |
| | 17 | 41926126 | 0.119593 | 0.0113657 | 6.80E-26 | rs72836561 | T | C | 0.031482 | 0.00961157 | 110.7163082 |
| | 17 | 17434141 | 0.0522963 | 0.0082221 | 2.00E-10 | rs9889584 | A | G | 0.062203 | 0.00351412 | 40.45468929 |
| | 17 | 17538289 | 0.0491186 | 0.00862889 | 1.30E-08 | rs75634664 | G | C | 0.060201 | 0.00281483 | 32.40220361 |

| | | | | | | | | | | | |
|---|---|---|---|---|---|---|---|---|---|---|---|
| | 17 | 64210580 | －0.100552 | 0.0115384 | 2.90E-18 | rs1801689 | C | A | 0.030526 | 0.00659472 | 75.94211669 |
| | 19 | 8423432 | －0.053786 | 0.00868467 | 5.90E-10 | rs371786442 | AT | A | 0.055899 | 0.00333181 | 38.35520479 |
| | 19 | 19380513 | －0.112857 | 0.0181006 | 4.50E-10 | rs187429064 | G | A | 0.013453 | 0.00337689 | 38.87438336 |
| | 19 | 45416178 | 0.0442987 | 0.0046738 | 2.60E-21 | rs483082 | T | G | 0.236539 | 0.00780001 | 89.83264082 |
| | 19 | 19379549 | －0.11249 | 0.00756565 | 5.30E-50 | rs58542926 | T | C | 0.074399 | 0.01917319 | 221.0689729 |
| | 19 | 8429323 | －0.226767 | 0.0142327 | 3.80E-57 | rs116843064 | A | G | 0.019851 | 0.02201004 | 253.8502357 |
| | 19 | 45430280 | 0.0811629 | 0.00426686 | 1.10E-80 | rs5112 | G | C | 0.533735 | 0.03134207 | 361.8185076 |
| | 20 | 44551855 | 0.0455572 | 0.00498211 | 5.80E-20 | rs6073958 | C | T | 0.198731 | 0.00726518 | 83.66850434 |
| exposure | chr.exposure | pos.exposure | beta.exposure | se.exposure | pval.exposure | SNP | effect_allele.exposure | other_allele.exposure | eaf.exposure | R2 | F |
| Total lipid levels in lar | 1 | 63060037 | －0.075277 | 0.00416745 | 6.20E-73 | rs79439217 | TAAGAA | T | 0.352737 | 0.02827137 | 326.2693384 |
| | 1 | 109822166 | 0.0356592 | 0.00473829 | 5.20E-14 | rs599839 | A | G | 0.77208 | 0.00491902 | 56.63597335 |

| | | | | | | | | | | | |
|---|---|---|---|---|---|---|---|---|---|---|---|
| ge VLDL \|\| id: ebi-a-GCST90092874 | 1 | 27284913 | 0.105989 | 0.0132266 | 1.10E-15 | rs79598313 | T | C | 0.023201 | 0.00557668 | 64.21220774 |
| | 1 | 230296153 | -0.0433014 | 0.00407652 | 2.40E-26 | rs10864726 | T | C | 0.605471 | 0.00979471 | 112.8280737 |
| | 2 | 27730940 | -0.0891439 | 0.00406859 | 2.10E-106 | rs1260326 | C | T | 0.603982 | 0.0415413 | 480.0515302 |
| | 2 | 227026864 | -0.0378532 | 0.00428098 | 9.40E-19 | rs13423088 | A | T | 0.317018 | 0.00678917 | 78.18280601 |
| | 2 | 21530659 | -0.0279987 | 0.00406495 | 5.70E-12 | rs60960031 | A | G | 0.402852 | 0.00412078 | 47.44142994 |
| | 2 | 21231524 | -0.107935 | 0.00490851 | 3.60E-107 | rs676210 | A | G | 0.205766 | 0.0418405 | 483.5236086 |
| | 2 | 165528876 | -0.0482706 | 0.00407748 | 2.50E-32 | rs13389219 | T | C | 0.392532 | 0.01216314 | 140.1438898 |
| | 3 | 135956305 | 0.0327062 | 0.00471655 | 4.10E-12 | rs684773 | C | A | 0.767075 | 0.0041766 | 48.08434928 |
| | 3 | 52838654 | -0.0390033 | 0.0061079 | 1.70E-10 | rs9968117 | T | C | 0.120935 | 0.00354207 | 40.77662931 |
| | 4 | 171563776 | -0.0259742 | 0.00468723 | 3.00E-08 | rs4491981 | A | C | 0.241218 | 0.00266765 | 30.70748749 |
| | 4 | 3443931 | -0.0366147 | 0.00412473 | 6.90E-19 | rs13108218 | G | A | 0.615355 | 0.00684251 | 78.79748601 |

| | | | | | | | | | | | |
|---|---|---|---|---|---|---|---|---|---|---|---|
| | 4 | 87976359 | 0.0296749 | 0.00408883 | 3.90E-13 | rs1471251 | T | A | 0.397121 | 0.00457482 | 52.67115863 |
| | 4 | 15099148 | 0.0231543 | 0.00422919 | 4.40E-08 | rs1823949 | C | T | 0.66351 | 0.00260392 | 29.97377004 |
| | 5 | 55857025 | 0.0552631 | 0.00507805 | 1.40E-27 | rs11429307 | GT | G | 0.191517 | 0.01028069 | 118.4319808 |
| | 5 | 74656539 | 0.0239994 | 0.0040679 | 3.60E-09 | rs12916 | C | T | 0.400341 | 0.00302358 | 34.80588527 |
| | 5 | 156443066 | 0.0381645 | 0.00415652 | 4.20E-20 | rs4704834 | G | A | 0.644028 | 0.00732039 | 84.30471358 |
| | 6 | 127455029 | 0.0339381 | 0.0039928 | 1.90E-17 | rs2186037 | G | A | 0.518794 | 0.00627393 | 72.24576409 |
| | 6 | 139834012 | -0.0350701 | 0.0041165 | 1.60E-17 | rs632057 | G | T | 0.627778 | 0.00630284 | 72.57887319 |
| | 6 | 160545046 | 0.0593932 | 0.00981628 | 1.40E-09 | rs11909590 | T | C | 0.043584 | 0.00318005 | 36.60766485 |
| | 6 | 160922870 | -0.282919 | 0.0147619 | 7.20E-82 | rs117733303 | G | A | 0.0185 | 0.0318162 | 367.3095018 |
| | 6 | 161010118 | -0.162068 | 0.00737642 | 5.50E-107 | rs10455872 | G | A | 0.078987 | 0.0417713 | 482.7204798 |
| | 6 | 31236679 | 0.0255383 | 0.0045393 | 1.80E-08 | rs1130580 | C | T | 0.289865 | 0.00274966 | 31.65181722 |

| | | | | | | | | | | | |
|---|---|---|---|---|---|---|---|---|---|---|---|
| | 6 | 32586831 | 0.0433674 | 0.00428908 | 4.90E-24 | rs13191810 | T | C | 0.312309 | 0.00887576 | 102.2329588 |
| | 6 | 43757896 | 0.0330401 | 0.00398406 | 1.10E-16 | rs998584 | A | C | 0.482313 | 0.00597261 | 68.77386281 |
| | 6 | 32623528 | 0.0345693 | 0.00588103 | 4.10E-09 | rs73732214 | A | G | 0.140436 | 0.00300149 | 34.55149423 |
| | 6 | 161017363 | -0.0938829 | 0.0110789 | 2.40E-17 | rs73596816 | A | G | 0.033593 | 0.00623593 | 71.8078891 |
| | 7 | 130433594 | 0.0341869 | 0.00430057 | 1.90E-15 | rs11976955 | G | C | 0.314211 | 0.00548809 | 63.19167301 |
| | 7 | 73037956 | -0.112694 | 0.00583873 | 5.30E-83 | rs34060476 | G | A | 0.13482 | 0.00322665 | 372.5265714 |
| | 7 | 73123473 | 0.0703452 | 0.00924797 | 2.80E-14 | rs72555385 | G | A | 0.049304 | 0.00502517 | 57.85868586 |
| | 7 | 25983400 | -0.0256605 | 0.00416231 | 7.10E-10 | rs56001710 | T | A | 0.580583 | 0.00330149 | 38.00614636 |
| | 8 | 59398461 | -0.0266898 | 0.0042232 | 2.60E-10 | rs10504255 | A | G | 0.662958 | 0.00346936 | 39.9392397 |
| | 8 | 19733244 | -0.0625253 | 0.00485375 | 5.70E-38 | rs1561748 | C | G | 0.267445 | 0.01439871 | 165.9392847 |
| | 8 | 19819724 | -0.189738 | 0.00660732 | 2.40E-181 | rs328 | G | C | 0.100307 | 0.007114587 | 824.613438 |

| | | | | | | | | | | | |
|---|---|---|---|---|---|---|---|---|---|---|---|
| | 8 | 126500031 | -0.0805155 | 0.00407939 | 1.00E-86 | rs28601761 | G | C | 0.420159 | 0.003373599 | 389.5480072 |
| | 8 | 11493678 | 0.0324251 | 0.0041111 | 3.10E-15 | rs13265201 | G | C | 0.396261 | 0.000540262 | 62.20696582 |
| | 8 | 22882293 | -0.0241227 | 0.00428985 | 1.90E-08 | rs59296513 | A | G | 0.320415 | 0.00274689 | 31.61986929 |
| | 8 | 126631799 | 0.0315239 | 0.00488982 | 1.10E-10 | rs3860847 | A | G | 0.212508 | 0.00361019 | 41.56105752 |
| | 10 | 94796504 | -0.0270884 | 0.00410251 | 4.00E-11 | rs835273 | T | A | 0.38487 | 0.003787 | 43.59732703 |
| | 10 | 64882300 | -0.0353155 | 0.00401567 | 1.40E-18 | rs10761716 | G | C | 0.44151 | 0.00671607 | 77.34052912 |
| | 11 | 116389017 | 0.024986 | 0.00434872 | 9.20E-09 | rs883863 | A | G | 0.308012 | 0.00286773 | 33.01133077 |
| | 11 | 116648917 | -0.208614 | 0.0058717 | 1.00E-200 | rs964184 | C | G | 0.86726 | 0.01084961 | 1262.268186 |
| | 11 | 116714293 | 0.274257 | 0.0210465 | 8.20E-39 | rs141469619 | G | A | 0.010141 | 0.01473357 | 169.8040824 |
| | 11 | 117156893 | -0.2349 | 0.0202622 | 4.50E-31 | rs1047964 | T | G | 0.010073 | 0.01166483 | 134.3956742 |
| | 11 | 61575158 | 0.038254 | 0.00418797 | 6.60E-20 | rs174553 | G | A | 0.342558 | 0.00724477 | 83.43322585 |

| | | | | | | | | | | | |
|---|---|---|---|---|---|---|---|---|---|---|---|
| | 11 | 63862882 | 0.0486564 | 0.00874729 | 2.70E-08 | rs11231694 | C | G | 0.055796 | 0.00268787 | 30.94035901 |
| | 12 | 124409502 | -0.0328966 | 0.00427276 | 1.40E-14 | rs7133378 | A | G | 0.319789 | 0.00514818 | 59.27580792 |
| | 12 | 124645325 | 0.0270402 | 0.0049555 | 4.90E-08 | rs113087348 | C | T | 0.208741 | 0.00258658 | 29.77400729 |
| | 12 | 4384844 | -0.0846566 | 0.0154999 | 4.70E-08 | rs76895963 | G | T | 0.020556 | 0.00259146 | 29.83020956 |
| | 12 | 57766392 | -0.0299128 | 0.00465794 | 1.30E-10 | rs4760254 | C | G | 0.239066 | 0.00358231 | 41.24001427 |
| | 13 | 114544024 | 0.0282774 | 0.00435371 | 8.30E-11 | rs7140110 | C | T | 0.299365 | 0.00366432 | 42.18445164 |
| | 15 | 58723479 | -0.0331193 | 0.00482852 | 6.90E-12 | rs1077834 | C | T | 0.218646 | 0.00408648 | 47.04643083 |
| | 15 | 43726625 | 0.126321 | 0.0126117 | 1.30E-23 | rs150844304 | C | A | 0.025671 | 0.00871 | 100.3220272 |
| | 15 | 58671765 | 0.0264633 | 0.00422435 | 3.70E-10 | rs1601935 | T | G | 0.654638 | 0.00340889 | 39.24286131 |
| | 16 | 81534790 | -0.0322224 | 0.00434306 | 2.00E-14 | rs2925979 | C | T | 0.698949 | 0.00508209 | 58.51440585 |
| | 16 | 56993886 | -0.0565935 | 0.00426079 | 2.90E-40 | rs821840 | G | A | 0.324526 | 0.01530665 | 176.4189771 |

| | | | | | | | | | | | |
|---|---|---|---|---|---|---|---|---|---|---|---|
| | 16 | 72096227 | 0.0441994 | 0.00657692 | 1.80E-11 | rs34682685 | A | G | 0.103441 | 0.00392292 | 45.16271912 |
| | 17 | 41926126 | 0.124675 | 0.011413 | 8.90E-28 | rs72836561 | T | C | 0.031482 | 0.01035861 | 119.3305306 |
| | 17 | 17431208 | 0.0562641 | 0.00870541 | 1.00E-10 | rs8078513 | A | C | 0.057349 | 0.00362843 | 41.77118011 |
| | 17 | 64210580 | -0.0927838 | 0.0115863 | 1.20E-15 | rs1801689 | C | A | 0.030526 | 0.00556936 | 64.12793041 |
| | 19 | 45430280 | 0.0807926 | 0.00428444 | 2.60E-79 | rs5112 | G | C | 0.533735 | 0.03080405 | 355.5883228 |
| | 19 | 8423432 | -0.0550402 | 0.00872044 | 2.80E-10 | rs371786442 | AT | A | 0.055899 | 0.00346039 | 39.83599438 |
| | 19 | 19380513 | -0.1273 | 0.0181752 | 2.50E-12 | rs187429064 | G | A | 0.013453 | 0.00426094 | 49.05585648 |
| | 19 | 8429323 | -0.223225 | 0.0142913 | 5.40E-55 | rs116843064 | A | G | 0.019851 | 0.0211551 | 243.9689987 |
| | 19 | 19379549 | -0.124873 | 0.00759681 | 1.00E-60 | rs58542926 | T | C | 0.074399 | 0.02342333 | 270.1885957 |
| | 19 | 45424514 | 0.0490743 | 0.00525659 | 1.00E-20 | rs157592 | C | A | 0.185291 | 0.0075677 | 87.15503273 |
| | 20 | 44576502 | 0.0465018 | 0.00512778 | 1.20E-19 | rs7679 | C | T | 0.186406 | 0.00714107 | 82.23812728 |

| | | | | | | | | | | | |
|---|---|---|---|---|---|---|---|---|---|---|---|
| | 20 | 39186590 | -0.0247265 | 0.00430921 | 9.60E-09 | rs6102034 | G | C | 0.323224 | 0.00286021 | 32.92474562 |
| exposure | chr.exposure | pos.exposure | beta.exposure | se.exposure | pval.exposure | SNP | effect_allele.exposure | other_allele.exposure | eaf.exposure | R2 | F |
| Total lipid levels in medium VLDL \|\| id:ebi-a-GCST90092922 | 1 | 27021913 | 0.0860533 | 0.0137381 | 3.80E-10 | rs114165349 | C | G | 0.022955 | 0.00340821 | 39.23504587 |
| | 1 | 234872431 | 0.0264832 | 0.00411294 | 1.20E-10 | rs613334 | A | G | 0.503583 | 0.00360141 | 41.45993577 |
| | 1 | 55491853 | -0.0386618 | 0.00550399 | 2.20E-12 | rs200730299 | C | A | 0.194501 | 0.00428564 | 49.34024706 |
| | 1 | 109817590 | -0.0777174 | 0.00492747 | 4.80E-56 | rs12740374 | T | G | 0.221064 | 0.02156966 | 248.7602593 |
| | 1 | 63085997 | -0.0766176 | 0.00435658 | 3.10E-69 | rs12239736 | A | T | 0.341234 | 0.02680357 | 309.2844983 |
| | 1 | 230295789 | -0.0329828 | 0.00419245 | 3.60E-15 | rs10127775 | T | A | 0.605599 | 0.00537525 | 61.89160018 |
| | 2 | 27730940 | -0.0764126 | 0.00418383 | 1.60E-74 | rs1260326 | C | T | 0.603982 | 0.02890131 | 333.5602935 |
| | 2 | 134688669 | 0.0272275 | 0.00487177 | 2.30E-08 | rs1900349 | C | T | 0.770721 | 0.00271342 | 31.23449095 |
| | 2 | 44072576 | -0.0348392 | 0.0043846 | 1.90E-15 | rs4299376 | T | G | 0.676277 | 0.00548316 | 63.13480467 |

| | | | | | | | | | | | |
|---|---|---|---|---|---|---|---|---|---|---|---|
| | 2 | 21530659 | -0.0416204 | 0.00418009 | 2.40E-23 | rs60960031 | A | G | 0.402852 | 0.000860716 | 99.13652391 |
| | 2 | 21198900 | -0.109507 | 0.00487055 | 6.00E-112 | rs11902417 | A | G | 0.227855 | 0.004373376 | 505.4988862 |
| | 2 | 165528876 | -0.0362703 | 0.00419297 | 5.10E-18 | rs13389219 | T | C | 0.392532 | 0.00649784 | 74.8257774 |
| | 3 | 52841220 | -0.04085 | 0.00645753 | 2.50E-10 | rs13059141 | A | G | 0.11318 | 0.0034761 | 40.01692462 |
| | 3 | 136581611 | -0.0345291 | 0.00503045 | 6.70E-12 | rs71630059 | A | G | 0.210845 | 0.00409234 | 47.11392608 |
| | 4 | 3443931 | -0.0361707 | 0.00424065 | 1.50E-17 | rs13108218 | G | A | 0.615355 | 0.00631782 | 72.75141091 |
| | 4 | 87976359 | 0.0271541 | 0.00420374 | 1.10E-10 | rs1471251 | T | A | 0.397121 | 0.00362438 | 41.72454256 |
| | 5 | 74472939 | 0.023669 | 0.00426747 | 2.90E-08 | rs7707394 | A | G | 0.357075 | 0.00267236 | 30.76176982 |
| | 5 | 55857025 | 0.0429047 | 0.00521981 | 2.00E-16 | rs11429307 | GT | G | 0.191517 | 0.0058673 | 67.56049167 |
| | 5 | 74656539 | 0.0445826 | 0.00418147 | 1.50E-26 | rs12916 | C | T | 0.400341 | 0.00986817 | 113.6750795 |
| | 5 | 156443066 | 0.048384 | 0.00427256 | 9.90E-30 | rs4704834 | G | A | 0.644028 | 0.01113105 | 128.2388606 |

| | | | | | | | | | | | |
|---|---|---|---|---|---|---|---|---|---|---|---|
| | 6 | 32434716 | 0.042363 | 0.00639416 | 3.50E-11 | rs28377109 | A | C | 0.116096 | 0.00381271 | 43.89336426 |
| | 6 | 139835498 | -0.0291037 | 0.00416258 | 2.70E-12 | rs17585887 | C | T | 0.590922 | 0.00424599 | 48.88365563 |
| | 6 | 32586376 | 0.0484871 | 0.00447485 | 2.30E-27 | rs11755689 | G | A | 0.334143 | 0.01019167 | 117.4053555 |
| | 6 | 161111700 | -0.261219 | 0.0172717 | 1.10E-51 | rs186696265 | T | C | 0.014352 | 0.01983671 | 228.7345692 |
| | 6 | 127455029 | 0.0277966 | 0.00410768 | 1.30E-11 | rs2186037 | G | A | 0.518794 | 0.0039775 | 45.7912607 |
| | 6 | 43757896 | 0.0228486 | 0.00409869 | 2.50E-08 | rs998584 | A | C | 0.482313 | 0.00269963 | 31.07574131 |
| | 6 | 160575366 | 0.0354252 | 0.00544002 | 7.40E-11 | rs11753995 | A | G | 0.171039 | 0.00368346 | 42.4049037 |
| | 6 | 160543148 | 0.0644448 | 0.00805037 | 1.20E-15 | rs12208357 | T | C | 0.07 | 0.00556538 | 64.08206974 |
| | 7 | 73037366 | -0.0727759 | 0.0051012 | 3.50E-46 | rs55747707 | A | G | 0.203642 | 0.01765451 | 203.5274167 |
| | 7 | 130433594 | 0.0269596 | 0.00442173 | 1.10E-09 | rs11976955 | G | C | 0.314211 | 0.0032292 | 37.1736239 |
| | 8 | 59398461 | -0.0298029 | 0.00434588 | 7.00E-12 | rs10504255 | A | G | 0.662958 | 0.00408486 | 47.02773291 |

| | | | | | | | | | | | |
|---|---|---|---|---|---|---|---|---|---|---|---|
| | 8 | 8279462 | －0.0237066 | 0.0041286 | 9.40E-09 | rs2976940 | G | T | 0.549682 | 0.000286419 | 32.97048835 |
| | 8 | 19773155 | 0.133607 | 0.0154525 | 5.30E-18 | rs144503444 | C | T | 0.018253 | 0.00064919 | 74.75725958 |
| | 8 | 19819724 | －0.14183 | 0.00679925 | 1.20E-96 | rs328 | G | C | 0.100307 | 0.003766759 | 435.1175879 |
| | 8 | 116657911 | －0.0266411 | 0.00456458 | 5.30E-09 | rs2721961 | G | T | 0.28083 | 0.000295915 | 34.06395824 |
| | 8 | 126506694 | －0.0821673 | 0.00425073 | 3.00E-83 | rs112875651 | A | G | 0.392379 | 0.003236352 | 373.6486736 |
| | 8 | 19750044 | 0.0608949 | 0.0100727 | 1.50E-09 | rs59347135 | G | C | 0.045803 | 0.000317486 | 36.5479058 |
| | 9 | 136155000 | 0.033166 | 0.00531054 | 4.20E-10 | rs635634 | T | C | 0.183543 | 0.000338808 | 39.00329308 |
| | 10 | 94797973 | －0.0260431 | 0.00418156 | 4.70E-10 | rs2478236 | A | G | 0.403791 | 0.000336942 | 38.78833555 |
| | 10 | 64882300 | －0.0233036 | 0.00412832 | 1.70E-08 | rs10761716 | G | C | 0.44151 | 0.000276804 | 31.86337535 |
| | 11 | 117156893 | －0.180134 | 0.0208342 | 5.30E-18 | rs1047964 | T | G | 0.010073 | 0.000649155 | 74.75326747 |
| | 11 | 116714293 | 0.20944 | 0.0216407 | 3.70E-22 | rs141469619 | G | A | 0.010141 | 0.000813235 | 93.66322379 |

| | | | | | | | | | | | |
|---|---|---|---|---|---|---|---|---|---|---|---|
| | 11 | 116648917 | −0.174512 | 0.00603745 | 1.00E-183 | rs964184 | C | G | 0.86726 | 0.007207664 | 835.4797916 |
| | 12 | 124419728 | −0.0287179 | 0.0043288 | 3.30E-11 | rs11057397 | T | C | 0.337502 | 0.000382293 | 44.01115523 |
| | 12 | 9172646 | 0.0237543 | 0.00417643 | 1.30E-08 | rs10842724 | C | T | 0.412903 | 0.000281025 | 32.34942108 |
| | 12 | 57650599 | −0.0301932 | 0.00539174 | 2.10E-08 | rs10649122 | ATCTC | A | 0.182469 | 0.000272417 | 31.358325 |
| | 13 | 114547372 | 0.030055 | 0.00426587 | 1.80E-12 | rs6602911 | T | C | 0.360028 | 0.000431145 | 49.63758961 |
| | 15 | 43726625 | 0.0951216 | 0.0129634 | 2.20E-13 | rs150844304 | C | A | 0.025671 | 0.000467638 | 53.8409733 |
| | 16 | 72101525 | 0.0463494 | 0.0052754 | 1.60E-18 | rs34042070 | G | C | 0.186978 | 0.000670315 | 77.19157378 |
| | 16 | 56989590 | −0.054649 | 0.0043798 | 9.90E-36 | rs247616 | T | C | 0.324699 | 0.001351019 | 155.6855598 |
| | 17 | 67082962 | −0.0369894 | 0.00574757 | 1.20E-10 | rs740516 | G | C | 0.151123 | 0.000359768 | 41.41698563 |
| | 17 | 41840849 | 0.0818949 | 0.0122544 | 2.30E-11 | rs77697917 | T | C | 0.028723 | 0.00038793 | 44.66034379 |
| | 17 | 45766771 | 0.0230968 | 0.00412774 | 2.20E-08 | rs56325564 | A | G | 0.482691 | 0.00027199 | 31.30915971 |

| | | | | | | | | | | | |
|---|---|---|---|---|---|---|---|---|---|---|---|
| | 17 | 17431208 | 0.0526735 | 0.00894833 | 3.90E-09 | rs8078513 | A | C | 0.057349 | 0.00300997 | 34.64916895 |
| | 19 | 11202306 | −0.107972 | 0.00629575 | 6.30E-66 | rs6511720 | T | G | 0.119788 | 0.002549244 | 294.1167756 |
| | 19 | 11227480 | 0.0264844 | 0.00417577 | 2.30E-10 | rs2738447 | C | A | 0.59318 | 0.0034942 | 40.22532759 |
| | 19 | 45039971 | −0.0857607 | 0.013429 | 1.70E-10 | rs74747585 | C | T | 0.025173 | 0.00354265 | 40.78324131 |
| | 19 | 45413233 | −0.205892 | 0.00755166 | 1.10E-163 | rs1065853 | T | G | 0.080581 | 0.006417863 | 743.3383514 |
| | 19 | 45445517 | 0.13533 | 0.018226 | 1.10E-13 | rs79429216 | A | G | 0.01273 | 0.0047884 | 55.13125568 |
| | 19 | 45401868 | 0.101673 | 0.0105168 | 4.10E-22 | rs118170342 | C | T | 0.040768 | 0.00811492 | 93.46230744 |
| | 19 | 19379549 | −0.140114 | 0.00780513 | 4.70E-72 | rs58542926 | T | C | 0.074399 | 0.02792421 | 322.2517123 |
| | 20 | 39179822 | 0.102972 | 0.0120348 | 1.20E-17 | rs1883711 | C | G | 0.031207 | 0.00635736 | 73.20706468 |
| | 21 | 40569508 | 0.0232678 | 0.00424519 | 4.20E-08 | rs62222988 | C | T | 0.373873 | 0.00260973 | 30.04061584 |
| | 22 | 50868669 | 0.0256284 | 0.00422324 | 1.30E-09 | rs9616847 | T | A | 0.388272 | 0.00319893 | 36.82509832 |

| exposure | chr.exposure | pos.exposure | beta.exposure | se.exposure | pval.exposure | SNP | effect_allele.exposure | other_allele.exposure | eaf.exposure | R2 | F |
|---|---|---|---|---|---|---|---|---|---|---|---|
| Total lipid levels in small VLDL \|\| id:ebi-a-GCST90092974 | 1 | 27021913 | 0.0972894 | 0.0136554 | 1.00E-12 | rs114165349 | C | G | 0.022955 | 0.00440883 | 50.75918613 |
| | 1 | 63085997 | -0.0787886 | 0.00433037 | 5.70E-74 | rs12239736 | A | T | 0.341234 | 0.002868282 | 331.0314137 |
| | 1 | 230295789 | -0.0385901 | 0.00416722 | 2.00E-20 | rs10127775 | T | A | 0.605599 | 0.00744609 | 85.75341078 |
| | 1 | 55491853 | -0.0334852 | 0.00547087 | 9.30E-10 | rs200730299 | C | A | 0.194501 | 0.0032542 | 37.46152533 |
| | 1 | 234873275 | 0.0255923 | 0.00410414 | 4.50E-10 | rs822928 | C | A | 0.52964 | 0.00337769 | 38.88362575 |
| | 1 | 109818306 | 0.0715617 | 0.0048907 | 1.80E-48 | rs629301 | T | G | 0.778041 | 0.01856969 | 214.0975528 |
| | 2 | 21198900 | -0.122255 | 0.00483968 | 8.59E-141 | rs11902417 | A | G | 0.227855 | 0.05514309 | 638.1053977 |
| | 2 | 27730940 | -0.0878763 | 0.00415731 | 3.60E-99 | rs1260326 | C | T | 0.603982 | 0.03867483 | 446.7979359 |
| | 2 | 227026864 | -0.0263224 | 0.00437433 | 1.80E-09 | rs13423088 | A | T | 0.317018 | 0.00314546 | 36.20931716 |
| | 2 | 21530659 | -0.0419387 | 0.00415359 | 5.70E-24 | rs60960031 | A | G | 0.402852 | 0.00885097 | 101.9471597 |

| | | | | | | | | | | | |
|---|---|---|---|---|---|---|---|---|---|---|---|
| | 2 | 44072576 | －0.0275637 | 0.00435681 | 2.50E-10 | rs4299376 | T | G | 0.676277 | 0.0034768 | 40.02489854 |
| | 2 | 165528624 | －0.0404368 | 0.00416761 | 2.90E-22 | rs1128249 | T | G | 0.392003 | 0.000817367 | 94.13949615 |
| | 3 | 52841220 | －0.037581 | 0.0064198 | 4.80E-09 | rs13059141 | A | G | 0.11318 | 0.00297685 | 34.26779181 |
| | 3 | 135798658 | －0.0377742 | 0.00487382 | 9.20E-15 | rs34894639 | T | C | 0.231578 | 0.00521696 | 60.06811889 |
| | 4 | 3443931 | －0.0372804 | 0.0042157 | 9.30E-19 | rs13108218 | G | A | 0.615355 | 0.00679077 | 78.20133799 |
| | 4 | 87976359 | 0.0320677 | 0.00417901 | 1.70E-14 | rs1471251 | T | A | 0.397121 | 0.00511398 | 58.88182412 |
| | 5 | 74656539 | 0.0364422 | 0.00415723 | 1.90E-18 | rs12916 | C | T | 0.400341 | 0.00667273 | 76.84109086 |
| | 5 | 55857025 | 0.050619 | 0.00518956 | 1.80E-22 | rs11429307 | GT | G | 0.191517 | 0.00826037 | 95.13898195 |
| | 5 | 156443066 | 0.0485023 | 0.00424779 | 3.40E-30 | rs4704834 | G | A | 0.644028 | 0.01131617 | 130.3740158 |
| | 6 | 32586376 | 0.0491342 | 0.00444872 | 2.30E-28 | rs11755689 | G | A | 0.334143 | 0.0105884 | 121.9804093 |
| | 6 | 139834012 | －0.0330222 | 0.00421021 | 4.40E-15 | rs632057 | G | T | 0.627778 | 0.00534275 | 61.51725415 |

| | | | | | | | | | | | |
|---|---|---|---|---|---|---|---|---|---|---|---|
| | 6 | 43757896 | 0.0258949 | 0.00407476 | 2.10E-10 | rs998584 | A | C | 0.482313 | 0.00350804 | 40.38469993 |
| | 6 | 160521885 | 0.0668675 | 0.00778554 | 1.10E-17 | rs146203232 | T | C | 0.074662 | 0.00637129 | 73.36752103 |
| | 6 | 32627812 | -0.0433943 | 0.00653303 | 3.10E-11 | rs9273453 | G | C | 0.127094 | 0.00383232 | 44.11929068 |
| | 6 | 127451665 | 0.0285127 | 0.00410535 | 3.80E-12 | rs1936806 | T | C | 0.448453 | 0.00418974 | 48.23571987 |
| | 6 | 161111700 | -0.229381 | 0.0171708 | 1.10E-40 | rs186696265 | T | C | 0.014352 | 0.01548294 | 178.4539956 |
| | 7 | 130433594 | 0.0290454 | 0.00439576 | 3.90E-11 | rs11976955 | G | C | 0.314211 | 0.0037924 | 43.65954751 |
| | 7 | 73037366 | -0.0855596 | 0.00507125 | 7.30E-64 | rs55747707 | A | G | 0.203642 | 0.0246733 | 284.6426028 |
| | 8 | 11677905 | 0.0284798 | 0.00491142 | 6.70E-09 | rs1293299 | C | A | 0.22709 | 0.00292096 | 33.62421654 |
| | 8 | 19773155 | 0.159429 | 0.0153535 | 2.90E-25 | rs144503444 | C | T | 0.018253 | 0.00936064 | 107.8232231 |
| | 8 | 19830921 | -0.151685 | 0.00616457 | 1.10E-133 | rs10096633 | T | C | 0.123907 | 0.05233515 | 605.4414455 |
| | 8 | 59385919 | -0.0284114 | 0.0043108 | 4.40E-11 | rs2326077 | T | C | 0.663229 | 0.0037731 | 43.43721506 |

| | | | | | | | | | | | |
|---|---|---|---|---|---|---|---|---|---|---|---|
| | 8 | 116657911 | -0.0296788 | 0.00453535 | 6.00E-11 | rs2721961 | G | T | 0.28083 | 0.000371965 | 42.82166119 |
| | 8 | 19750044 | 0.0723881 | 0.0100082 | 4.70E-13 | rs59347135 | G | C | 0.045803 | 0.00454378 | 52.31363003 |
| | 8 | 126506694 | -0.092491 | 0.00422351 | 2.70E-106 | rs112875651 | A | G | 0.392379 | 0.004149908 | 479.5615429 |
| | 8 | 126631799 | 0.0284244 | 0.00499964 | 1.30E-08 | rs3860847 | A | G | 0.212508 | 0.00280786 | 32.32195316 |
| | 9 | 136155000 | 0.0372473 | 0.00527951 | 1.70E-12 | rs635634 | T | C | 0.183543 | 0.00432322 | 49.77311397 |
| | 10 | 93641869 | -0.0228483 | 0.00408327 | 2.20E-08 | rs10881987 | G | A | 0.52745 | 0.00271998 | 31.3100702 |
| | 10 | 94797973 | -0.02739 | 0.00415679 | 4.40E-11 | rs2478236 | A | G | 0.403791 | 0.00377135 | 43.41705359 |
| | 10 | 64882300 | -0.027426 | 0.00410386 | 2.30E-11 | rs10761716 | G | C | 0.44151 | 0.00387939 | 44.66139892 |
| | 11 | 14360435 | 0.0265857 | 0.00483734 | 3.90E-08 | rs2970332 | A | G | 0.767351 | 0.00262399 | 30.20476247 |
| | 11 | 116648917 | -0.197198 | 0.00600116 | 1.00E-200 | rs964184 | C | G | 0.86726 | 0.00929547 | 1079.759547 |
| | 11 | 117156893 | -0.209208 | 0.0207089 | 5.40E-24 | rs1047964 | T | G | 0.010073 | 0.00886034 | 102.0551595 |

| | | | | | | | | | | | |
|---|---|---|---|---|---|---|---|---|---|---|---|
| | 11 | 116714293 | 0.237299 | 0.0215106 | 2.70E-28 | rs141469619 | G | A | 0.010141 | 0.00105638 | 121.6968171 |
| | 12 | 57650599 | -0.0370241 | 0.00535999 | 4.90E-12 | rs10649122 | ATCTC | A | 0.182469 | 0.000414432 | 47.71264733 |
| | 12 | 124419728 | -0.0322725 | 0.00430332 | 6.40E-14 | rs11057397 | T | C | 0.337502 | 0.000488471 | 56.24065726 |
| | 13 | 114547372 | 0.0305287 | 0.00424122 | 6.10E-13 | rs6602911 | T | C | 0.360028 | 0.00045002 | 51.81165599 |
| | 14 | 24874026 | 0.0226902 | 0.00408764 | 2.80E-08 | rs72694391 | C | T | 0.482108 | 0.000267675 | 30.81227885 |
| | 15 | 43726625 | 0.130832 | 0.0128843 | 3.20E-24 | rs150844304 | C | A | 0.025671 | 0.000895179 | 103.1095179 |
| | 15 | 58678720 | -0.0774769 | 0.00429285 | 8.20E-73 | rs261290 | C | T | 0.654645 | 0.002822397 | 325.7208023 |
| | 15 | 58725839 | 0.0559876 | 0.00449448 | 1.30E-35 | rs633695 | G | A | 0.292354 | 0.00134658 | 155.1733908 |
| | 16 | 72108093 | 0.0444412 | 0.00519945 | 1.30E-17 | rs2000999 | A | G | 0.188613 | 0.000634415 | 73.05487372 |
| | 16 | 56993886 | -0.0785299 | 0.00435295 | 9.30E-73 | rs821840 | G | A | 0.324526 | 0.002820128 | 325.4581743 |
| | 17 | 17431208 | 0.0557209 | 0.00889611 | 3.80E-10 | rs8078513 | A | C | 0.057349 | 0.000340785 | 39.23091241 |

| | | | | | | | | | | | |
|---|---|---|---|---|---|---|---|---|---|---|---|
| | 17 | 41926126 | 0.0843714 | 0.011663 | 4.70E-13 | rs72836561 | T | C | 0.031482 | 0.00454533 | 52.33140709 |
| | 17 | 67082962 | −0.0363917 | 0.00571403 | 1.90E-10 | rs740516 | G | C | 0.151123 | 0.00352338 | 40.56132256 |
| | 19 | 8424085 | −0.0704953 | 0.0116669 | 1.50E-09 | rs117777747 | T | C | 0.031641 | 0.0031715 | 36.50915889 |
| | 19 | 11202306 | −0.096284 | 0.00626067 | 2.30E-53 | rs6511720 | T | G | 0.119788 | 0.02051009 | 236.5152102 |
| | 19 | 45239536 | −0.0546322 | 0.00718348 | 2.80E-14 | rs62117162 | A | C | 0.088161 | 0.00502345 | 57.83890438 |
| | 19 | 19379549 | −0.116036 | 0.00776164 | 1.60E-50 | rs58542926 | T | C | 0.074399 | 0.01938333 | 223.4966203 |
| | 19 | 11228745 | 0.0245118 | 0.00415802 | 3.70E-09 | rs2569550 | C | T | 0.593326 | 0.00301883 | 34.75119259 |
| | 19 | 45345787 | −0.0463835 | 0.00849356 | 4.70E-08 | rs111371860 | T | A | 0.063586 | 0.00259076 | 29.82221983 |
| | 19 | 45425460 | 0.0811882 | 0.00428107 | 3.40E-80 | rs157595 | G | A | 0.617355 | 0.03115435 | 359.6446631 |
| | 19 | 49136515 | −0.0320013 | 0.00477806 | 2.10E-11 | rs550455 | C | T | 0.240881 | 0.00389633 | 44.85640834 |
| | 20 | 39174140 | −0.0314216 | 0.00447017 | 2.10E-12 | rs1883715 | T | C | 0.31262 | 0.00429156 | 49.40841983 |

| | | | | | | | | | | | |
|---|---|---|---|---|---|---|---|---|---|---|---|
| | 20 | 39678289 | 0.0257139 | 0.0040791 | 2.90E-10 | rs6016505 | T | C | 0.549538 | 0.000345183 | 39.7374184 |
| | 20 | 44551855 | 0.0487067 | 0.00511124 | 1.60E-21 | rs6073958 | C | T | 0.198731 | 0.000788451 | 90.80657944 |
| | 21 | 40569508 | 0.0239997 | 0.00421055 | 1.30E-08 | rs6222298 | C | T | 0.373873 | 0.000280829 | 32.32682746 |
| | 22 | 50868669 | 0.0251526 | 0.00419915 | 2.10E-09 | rs9616847 | T | A | 0.388272 | 0.000311674 | 35.87859633 |
| exposure | chr.exposure | pos.exposure | beta.exposure | se.exposure | pval.exposure | SNP | effect_allele.exposure | other_allele.exposure | eaf.exposure | R2 | F |
| Total lipid levels in very small VLDL \|\| id:ebi-a-GCST90093034 | 1 | 62999675 | 0.0665954 | 0.00422137 | 4.60E-56 | rs7534572 | G | C | 0.64747 | 0.002157923 | 248.8708764 |
| | 1 | 55505647 | -0.266913 | 0.0153898 | 2.20E-67 | rs11591147 | T | G | 0.017457 | 0.002606948 | 300.7917731 |
| | 1 | 55526428 | 0.0758481 | 0.0113059 | 2.00E-11 | rs11206517 | G | T | 0.033012 | 0.000390933 | 45.00612116 |
| | 1 | 234873275 | 0.0248701 | 0.00407194 | 1.00E-09 | rs822928 | C | A | 0.52964 | 0.000324044 | 37.30308719 |
| | 1 | 109817590 | -0.076007 | 0.0048594 | 3.80E-55 | rs12740374 | T | G | 0.221064 | 0.002121349 | 244.6438689 |
| | 2 | 27598097 | -0.0423815 | 0.00415302 | 1.90E-24 | rs4665972 | C | T | 0.604609 | 0.000904116 | 104.1398755 |

| | | | | | | | | | | | |
|---|---|---|---|---|---|---|---|---|---|---|---|
| | 2 | 169912191 | 0.0224855 | 0.00406557 | 3.20E-08 | rs2389599 | C | G | 0.524801 | 0.00265729 | 30.58825237 |
| | 2 | 20927997 | −0.0459911 | 0.00816657 | 1.80E-08 | rs6729843 | C | T | 0.065781 | 0.00275512 | 31.71470444 |
| | 2 | 21232195 | 0.0854519 | 0.00403948 | 2.50E-99 | rs693 | A | G | 0.520889 | 0.00387346 5 | 447.4916903 |
| | 2 | 44063098 | −0.0721747 | 0.00814391 | 7.80E-19 | rs115445558 | C | G | 0.06589 | 0.00682025 | 78.54101258 |
| | 2 | 20354039 | −0.0249952 | 0.00404495 | 6.40E-10 | rs11096640 | A | C | 0.483356 | 0.00331692 | 38.18381959 |
| | 3 | 135800409 | −0.0442645 | 0.0048237 | 4.50E-20 | rs34330586 | T | A | 0.231198 | 0.00731182 | 84.20599236 |
| | 3 | 170687273 | 0.0358139 | 0.00615306 | 5.90E-09 | rs11925442 | C | T | 0.123632 | 0.00294297 | 33.87765848 |
| | 4 | 69340991 | 0.0322122 | 0.00471452 | 8.30E-12 | rs4860948 | A | T | 0.244338 | 0.00405492 | 46.68294666 |
| | 4 | 87976359 | 0.0308089 | 0.00414562 | 1.10E-13 | rs1471251 | T | A | 0.397121 | 0.00479687 | 55.22883163 |
| | 4 | 100497578 | −0.0254796 | 0.00463921 | 4.00E-08 | rs11947558 | T | C | 0.254543 | 0.00262045 | 30.1640464 |
| | 4 | 38316530 | 0.0224321 | 0.00410714 | 4.70E-08 | rs34670304 | A | G | 0.553264 | 0.00259144 | 29.83000343 |

| | | | | | | | | | | | |
|---|---|---|---|---|---|---|---|---|---|---|---|
| | 5 | 74472939 | 0.0309683 | 0.0042085 | 1.90E-13 | rs7707394 | A | G | 0.357075 | 0.00470293 | 54.14676569 |
| | 5 | 74656539 | 0.0481566 | 0.00412369 | 1.70E-31 | rs12916 | C | T | 0.400341 | 0.01183635 | 136.3741532 |
| | 5 | 156443066 | 0.0459885 | 0.00421352 | 9.80E-28 | rs4704834 | G | A | 0.644028 | 0.01034074 | 119.1244263 |
| | 6 | 32601288 | 0.0493811 | 0.00523688 | 4.10E-21 | rs72848251 | A | G | 0.194675 | 0.00772029 | 88.91370907 |
| | 6 | 160543148 | 0.0713799 | 0.00794424 | 2.60E-19 | rs12208357 | T | C | 0.07 | 0.00701028 | 80.73086593 |
| | 6 | 160922870 | -0.0877251 | 0.0149864 | 4.80E-09 | rs117733303 | G | A | 0.0185 | 0.00297657 | 34.2645913 |
| | 6 | 160575366 | 0.0388075 | 0.0053683 | 4.90E-13 | rs11753995 | A | G | 0.171039 | 0.00453893 | 52.25769251 |
| | 6 | 31436032 | 0.0281413 | 0.00430018 | 6.00E-11 | rs2523675 | A | G | 0.342119 | 0.00372003 | 42.8259982 |
| | 7 | 21607283 | 0.0267759 | 0.00488183 | 4.10E-08 | rs55649657 | G | C | 0.219849 | 0.00261338 | 30.08259545 |
| | 7 | 1082918 | -0.0390347 | 0.00491034 | 1.90E-15 | rs10275712 | A | G | 0.215612 | 0.00548824 | 63.19329713 |
| | 7 | 44584551 | 0.0299488 | 0.00521677 | 9.40E-09 | rs10260606 | C | G | 0.182652 | 0.00286302 | 32.95701913 |

| | | | | | | | | | | | |
|---|---|---|---|---|---|---|---|---|---|---|---|
| | 8 | 19830921 | −0.0724603 | 0.00612089 | 2.50E-32 | rs10096633 | T | C | 0.123907 | 0.00121628 84 | 140.1404664 |
| | 8 | 59370320 | −0.0277336 | 0.00429729 | 1.10E-10 | rs4738679 | A | G | 0.666712 | 0.00036179 92 | 41.65005908 |
| | 8 | 116658583 | −0.0358172 | 0.00451406 | 2.10E-15 | rs2737245 | T | G | 0.278602 | 0.00054676 69 | 62.95657042 |
| | 8 | 126506694 | −0.0811535 | 0.00419358 | 2.00E-83 | rs112875651 | A | G | 0.392379 | 0.00324359 2 | 374.4873029 |
| | 9 | 136149711 | 0.0498105 | 0.00522571 | 1.50E-21 | rs115478735 | T | A | 0.183304 | 0.00078886 2 | 90.85391471 |
| | 10 | 93641869 | −0.0222633 | 0.00405068 | 3.90E-08 | rs10881987 | G | A | 0.52745 | 0.00026242 3 | 30.20756078 |
| | 11 | 117156893 | −0.125747 | 0.0205465 | 9.30E-10 | rs1047964 | T | G | 0.010073 | 0.00032536 5 | 37.45519072 |
| | 11 | 116714293 | 0.130224 | 0.0213418 | 1.00E-09 | rs141469619 | G | A | 0.010141 | 0.00032342 4 | 37.231662 |
| | 11 | 5701074 | 0.0455164 | 0.00786925 | 7.30E-09 | rs11601507 | A | C | 0.069168 | 0.00029062 7 | 33.45504186 |
| | 11 | 61557826 | −0.0429954 | 0.00424581 | 4.20E-24 | rs102274 | C | T | 0.343868 | 0.00089028 4 | 102.5451981 |
| | 11 | 116648917 | −0.103278 | 0.00595408 | 2.10E-67 | rs964184 | C | G | 0.86726 | 0.00260762 4 | 300.8699123 |

| | | | | | | | | | | | |
|---|---|---|---|---|---|---|---|---|---|---|---|
| | 12 | 124850753 | -0.0238965 | 0.00414861 | 8.40E-09 | rs2141247 | A | C | 0.612788 | 0.000288224 | 33.17842959 |
| | 12 | 57650599 | -0.0309376 | 0.00531675 | 5.90E-09 | rs10649122 | ATCTC | A | 0.182469 | 0.000294134 | 33.85892871 |
| | 12 | 121420260 | -0.0421376 | 0.00418249 | 7.10E-24 | rs7979473 | G | A | 0.61301 | 0.00088121 | 101.4991072 |
| | 13 | 32976654 | -0.0256976 | 0.00427106 | 1.80E-09 | rs61946991 | C | A | 0.451649 | 0.000314463 | 36.19980549 |
| | 13 | 114547372 | 0.0270052 | 0.0042065 | 1.40E-10 | rs6602911 | T | C | 0.360028 | 0.000358006 | 41.21406597 |
| | 15 | 58678720 | -0.20526 | 0.00424799 | 1.00E-200 | rs261290 | C | T | 0.654645 | 0.019884359 | 2334.716379 |
| | 15 | 58833993 | 0.0907964 | 0.0120558 | 5.00E-14 | rs6078 | A | G | 0.028853 | 0.000492633 | 56.72018544 |
| | 15 | 58725839 | 0.154077 | 0.00444752 | 1.00E-200 | rs633695 | G | A | 0.292354 | 0.010321117 | 1200.140966 |
| | 15 | 58555798 | -0.0481788 | 0.0045655 | 4.90E-26 | rs16939991 | G | C | 0.26718 | 0.000966736 | 111.3596137 |
| | 15 | 59243880 | -0.0743854 | 0.011852 | 3.50E-10 | rs146407081 | A | G | 0.031687 | 0.000342165 | 39.38987224 |
| | 15 | 44027885 | 0.0849452 | 0.0127082 | 2.30E-11 | rs139974673 | C | T | 0.025912 | 0.000388091 | 44.67885725 |

| | | | | | | | | | | | |
|---|---|---|---|---|---|---|---|---|---|---|---|
| | 16 | 56993886 | −0.0797304 | 0.00431658 | 3.60E-76 | rs821840 | G | A | 0.324526 | 0.002955802 | 341.1620771 |
| | 16 | 72144174 | −0.0443982 | 0.0052162 | 1.70E-17 | rs9302635 | C | T | 0.182459 | 0.000629132 | 72.44604966 |
| | 17 | 45752738 | 0.0271451 | 0.00414198 | 5.60E-11 | rs77142415 | T | G | 0.525048 | 0.00373076 | 42.94962231 |
| | 17 | 64195431 | 0.0657939 | 0.0114199 | 8.30E-09 | rs75003668 | G | A | 0.033354 | 0.00288346 | 33.19243483 |
| | 17 | 67191270 | −0.0426274 | 0.00602742 | 1.50E-12 | rs72631343 | G | C | 0.129021 | 0.00434429 | 50.01580132 |
| | 17 | 67081278 | 0.15577 | 0.0135848 | 1.90E-30 | rs77542162 | G | A | 0.022666 | 0.0114119 | 131.4781529 |
| | 18 | 47173181 | 0.0314222 | 0.00528767 | 2.80E-09 | rs1943977 | G | A | 0.822416 | 0.00306764 | 35.31318933 |
| | 18 | 47109955 | 0.129607 | 0.0176521 | 2.10E-13 | rs77960347 | G | A | 0.013234 | 0.00468224 | 53.9084209 |
| | 19 | 19388500 | −0.0983551 | 0.00765139 | 8.10E-38 | rs8107974 | T | A | 0.075552 | 0.0143378 | 165.2363309 |
| | 19 | 45039971 | −0.0741665 | 0.0132426 | 2.10E-08 | rs74747585 | C | T | 0.025173 | 0.00272485 | 31.36617054 |
| | 19 | 49236102 | 0.0315688 | 0.00405281 | 6.70E-15 | rs439523 | C | T | 0.52969 | 0.00526947 | 60.67308974 |

| | | | | | | | | | | | |
|---|---|---|---|---|---|---|---|---|---|---|---|
| | 19 | 45413233 | −0.211401 | 0.00744684 | 2.80E-177 | rs1065853 | T | G | 0.080581 | 0.0069539557 | 805.8653475 |
| | 19 | 11197598 | −0.136459 | 0.00621442 | 7.19E-107 | rs73015024 | T | G | 0.119667 | 0.004172342 | 482.1649287 |
| | 19 | 45173951 | −0.0577061 | 0.00937614 | 7.50E-10 | rs28807203 | C | A | 0.049207 | 0.00329037 | 37.87806052 |
| | 19 | 45445517 | 0.102575 | 0.017973 | 1.10E-08 | rs79429216 | A | G | 0.01273 | 0.00282951 | 32.57124468 |
| | 19 | 11227480 | 0.0301243 | 0.0041178 | 2.60E-13 | rs2738447 | C | A | 0.59318 | 0.0046483 | 53.51750789 |
| | 19 | 45421650 | 0.113961 | 0.0123091 | 2.10E-20 | rs140480140 | A | G | 0.028202 | 0.00744268 | 85.71411699 |
| | 20 | 34415578 | −0.0343792 | 0.00605326 | 1.40E-08 | rs112103450 | C | T | 0.129471 | 0.0028021 | 32.25561517 |
| | 20 | 44545460 | 0.0352876 | 0.00466751 | 4.00E-14 | rs4812975 | A | G | 0.750337 | 0.00496422 | 57.15657307 |
| | 20 | 39179822 | 0.106381 | 0.0118672 | 3.10E-19 | rs1883711 | C | G | 0.031207 | 0.00697784 | 80.35706499 |
| | 22 | 50868669 | 0.0267646 | 0.00416437 | 1.30E-10 | rs9616847 | T | A | 0.388272 | 0.00358806 | 41.30620998 |
| exposure | chr.exposure | pos.exposure | beta.exposure | se.exposure | pval.exposure | SNP | effect_allele.exposure | other_allele.exposure | eaf.exposure | R2 | F |

| | | | | | | | | | | | |
|---|---|---|---|---|---|---|---|---|---|---|---|
| Total lipid levels in IDL \|\| id:ebi-a-GCST90092837 | 1 | 55505647 | -0.319329 | 0.0152077 | 6.90E-98 | rs11591147 | T | G | 0.017457 | 0.003816643 | 440.9020326 |
| | 1 | 55518467 | -0.0311675 | 0.00416804 | 7.60E-14 | rs2495477 | G | A | 0.393802 | 0.00485648 | 55.91555402 |
| | 1 | 62999675 | 0.0346288 | 0.00417143 | 1.00E-16 | rs7534572 | G | C | 0.64747 | 0.00598463 | 68.9124065 |
| | 1 | 220970028 | 0.0350904 | 0.00437151 | 1.00E-15 | rs2642438 | G | A | 0.70385 | 0.00559581 | 64.43267139 |
| | 1 | 109817590 | -0.0967477 | 0.00480191 | 2.80E-90 | rs12740374 | T | G | 0.221064 | 0.00351493 | 405.9249248 |
| | 1 | 234852760 | 0.0278789 | 0.00399182 | 2.90E-12 | rs553427 | T | C | 0.517037 | 0.0042366 | 48.77551022 |
| | 1 | 55526428 | 0.093435 | 0.0111722 | 6.10E-17 | rs11206517 | G | T | 0.033012 | 0.00607394 | 69.94138513 |
| | 1 | 25713234 | 0.0221322 | 0.00399156 | 2.60E-08 | rs1293261 | T | A | 0.525176 | 0.00269037 | 30.9691569 |
| | 2 | 20371380 | -0.0247743 | 0.00403201 | 8.00E-10 | rs907866 | A | G | 0.443212 | 0.00327952 | 37.75304936 |
| | 2 | 21288321 | 0.106125 | 0.00518245 | 3.40E-93 | rs562338 | G | A | 0.819616 | 0.03630598 | 419.3316253 |
| | 2 | 44072576 | -0.0462788 | 0.00427535 | 2.60E-27 | rs4299376 | T | G | 0.676277 | 0.01017118 | 117.169151 |

| | | | | | | | | | | | |
|---|---|---|---|---|---|---|---|---|---|---|---|
| | 4 | 69340991 | 0.0266982 | 0.00465959 | 1.00E-08 | rs4860948 | A | T | 0.244338 | 0.00285192 | 32.8292535 |
| | 5 | 74651084 | 0.0555508 | 0.00404139 | 5.40E-43 | rs3846662 | G | A | 0.424111 | 0.00163907 7 | 188.934628 |
| | 5 | 156397673 | 0.0294918 | 0.00413618 | 1.00E-12 | rs6882345 | A | G | 0.632836 | 0.00441575 | 50.83890078 |
| | 5 | 74472939 | 0.0404621 | 0.00415955 | 2.30E-22 | rs7707394 | A | G | 0.357075 | 0.00821561 | 94.62297367 |
| | 5 | 122827953 | -0.0240208 | 0.0040206 | 2.30E-09 | rs1500188 | G | A | 0.450404 | 0.00310064 | 35.69321354 |
| | 5 | 52106278 | -0.058007 | 0.0100331 | 7.40E-09 | rs1499279 | G | T | 0.041623 | 0.00290374 | 33.42589011 |
| | 6 | 26267755 | 0.0255165 | 0.00399638 | 1.70E-10 | rs7773004 | G | A | 0.497283 | 0.00354118 | 40.76628214 |
| | 6 | 160567928 | -0.0351621 | 0.00502211 | 2.50E-12 | rs3798167 | T | G | 0.200093 | 0.0042578 | 49.01958385 |
| | 6 | 32601807 | 0.0459272 | 0.00601373 | 2.20E-14 | rs147841808 | G | C | 0.132301 | 0.00506553 | 58.32362907 |
| | 7 | 1062366 | -0.0347141 | 0.00481983 | 5.90E-13 | rs1057558 | C | T | 0.220312 | 0.00450552 | 51.87293624 |
| | 7 | 72878645 | 0.0368929 | 0.00601876 | 8.80E-10 | rs12056034 | G | A | 0.125693 | 0.00326379 | 37.57197197 |

| | | | | | | | | | | | |
|---|---|---|---|---|---|---|---|---|---|---|---|
| | 8 | 126506694 | -0.0473955 | 0.00414547 | 2.90E-30 | rs112875651 | A | G | 0.392379 | 0.001134557 | 130.7131009 |
| | 8 | 19733244 | 0.0274984 | 0.00487106 | 1.60E-08 | rs1561748 | C | G | 0.267445 | 0.000276847 | 31.86840621 |
| | 8 | 9187242 | 0.0509028 | 0.00663825 | 1.70E-14 | rs1461729 | G | A | 0.899237 | 0.000510678 | 58.798817 |
| | 8 | 19819328 | 0.0535571 | 0.00662651 | 6.40E-16 | rs325 | C | T | 0.100507 | 0.000567297 | 65.32160434 |
| | 9 | 107661742 | -0.0275449 | 0.00453314 | 1.20E-09 | rs2740488 | C | A | 0.265313 | 0.000320728 | 36.92126638 |
| | 9 | 136149711 | 0.0605343 | 0.00516441 | 9.90E-32 | rs115478735 | T | A | 0.183304 | 0.001192439 | 137.3896683 |
| | 10 | 113921825 | -0.0358031 | 0.00444701 | 8.20E-16 | rs2792735 | A | G | 0.72034 | 0.000562928 | 64.81828012 |
| | 10 | 64934548 | 0.0289371 | 0.00399753 | 4.50E-13 | rs10822145 | T | C | 0.476857 | 0.000455115 | 52.39851744 |
| | 11 | 75450576 | -0.0319575 | 0.00546529 | 5.00E-09 | rs673335 | C | T | 0.159776 | 0.000297018 | 34.19098595 |
| | 11 | 61557826 | -0.0736677 | 0.00419559 | 5.10E-69 | rs102274 | C | T | 0.343868 | 0.002671769 | 308.2908185 |
| | 11 | 126223122 | 0.0559068 | 0.00863263 | 9.40E-11 | rs112819436 | G | A | 0.056983 | 0.000364315 | 41.94068 |

| | | | | | | | | | | | |
|---|---|---|---|---|---|---|---|---|---|---|---|
| | 12 | 9133256 | 0.0277129 | 0.00479388 | 7.40E-09 | rs80017394 | G | A | 0.224972 | 0.00290306 | 33.41812541 |
| | 12 | 112007756 | 0.0293133 | 0.00399924 | 2.30E-13 | rs653178 | T | C | 0.516036 | 0.00466621 | 53.7238271 |
| | 12 | 121426594 | 0.0328216 | 0.00433001 | 3.50E-14 | rs1169294 | A | G | 0.310092 | 0.0049902 | 57.45583631 |
| | 13 | 32976654 | -0.0260129 | 0.00422121 | 7.20E-10 | rs61946991 | C | A | 0.451649 | 0.00329878 | 37.974851 |
| | 15 | 58725839 | 0.0776142 | 0.00440379 | 1.60E-69 | rs633695 | G | A | 0.292354 | 0.02691852 | 310.6144255 |
| | 15 | 58619349 | 0.022952 | 0.00417005 | 3.70E-08 | rs261276 | C | A | 0.641266 | 0.0026317 | 30.29360778 |
| | 15 | 58678720 | -0.101773 | 0.00420622 | 2.50E-129 | rs261290 | C | T | 0.654645 | 0.05061393 | 585.4281985 |
| | 16 | 72079657 | -0.0328948 | 0.00507043 | 8.70E-11 | rs77303550 | T | C | 0.192705 | 0.00365594 | 42.08791096 |
| | 16 | 15054534 | 0.0288044 | 0.00480509 | 2.00E-09 | rs56015824 | A | T | 0.654366 | 0.00312156 | 35.93412431 |
| | 17 | 41926126 | -0.0866469 | 0.0114326 | 3.50E-14 | rs72836561 | T | C | 0.031482 | 0.00498875 | 57.43921469 |
| | 17 | 67081278 | 0.153322 | 0.0134271 | 3.40E-30 | rs77542162 | G | A | 0.022666 | 0.01131736 | 130.3877998 |

| | | | | | | | | | | | |
|---|---|---|---|---|---|---|---|---|---|---|---|
| | 17 | 45763073 | 0.0305531 | 0.0040064 | 2.40E-14 | rs4794048 | C | A | 0.480798 | 0.000505097 | 58.15598301 |
| | 17 | 67191270 | -0.0375055 | 0.00595746 | 3.10E-10 | rs72631343 | G | C | 0.129021 | 0.000344279 | 39.63328784 |
| | 17 | 64195431 | 0.0763715 | 0.0112873 | 1.30E-11 | rs75003668 | G | A | 0.033354 | 0.00397651 | 45.7799124 |
| | 18 | 47160953 | 0.0371624 | 0.00524431 | 1.40E-12 | rs7241918 | T | G | 0.8229 | 0.00436148 | 50.21381847 |
| | 18 | 47109955 | 0.157599 | 0.017444 | 1.60E-19 | rs77960347 | G | A | 0.013234 | 0.0070876 | 81.62199608 |
| | 19 | 11147526 | -0.144346 | 0.0191666 | 5.00E-14 | rs73013176 | C | T | 0.011281 | 0.00492604 | 56.71681059 |
| | 19 | 11229066 | -0.0786913 | 0.0125397 | 3.50E-10 | rs79915079 | A | G | 0.025969 | 0.00342076 | 39.3796271 |
| | 19 | 45173951 | -0.12612 | 0.00924612 | 2.30E-42 | rs28807203 | C | A | 0.049207 | 0.01614135 | 186.0549913 |
| | 19 | 45357939 | 0.0505526 | 0.00403614 | 5.40E-36 | rs2927468 | G | A | 0.523287 | 0.01361305 | 156.8725666 |
| | 19 | 11197261 | -0.150816 | 0.0061282 | 9.79E-134 | rs12151108 | A | G | 0.119677 | 0.05235304 | 605.64959 |
| | 19 | 45413233 | -0.500732 | 0.00734357 | 1.00E-200 | rs1065853 | T | G | 0.080581 | 0.38831859 | 4649.311745 |

| | | | | | | | | | | | |
|---|---|---|---|---|---|---|---|---|---|---|---|
| | 19 | 45215081 | -0.120624 | 0.014171 | 1.70E-17 | rs111278137 | A | G | 0.021614 | 0.00629196 | 72.45342301 |
| | 19 | 19379549 | -0.0876817 | 0.00759005 | 7.20E-31 | rs58542926 | T | C | 0.074399 | 0.01158292 | 133.4507754 |
| | 19 | 45412955 | 0.14617 | 0.0121767 | 3.40E-33 | rs1081105 | C | A | 0.027621 | 0.01250565 | 144.0952728 |
| | 19 | 45254168 | 0.0552998 | 0.00598187 | 2.40E-20 | rs8103315 | A | C | 0.135788 | 0.00742067 | 85.46053996 |
| | 20 | 17844684 | -0.0261761 | 0.00421608 | 5.30E-10 | rs2618566 | T | G | 0.660294 | 0.00334842 | 38.5464732 |
| | 20 | 39179822 | 0.0872248 | 0.0117289 | 1.00E-13 | rs1883711 | C | G | 0.031207 | 0.00480341 | 55.30416632 |
| exposure | chr.exposure | pos.exposure | beta.exposure | se.exposure | pval.exposure | SNP | effect_allele.exposure | other_allele.exposure | eaf.exposure | R2 | F |
| Total lipid levels in large LDL \|\| id:ebi-a-GCST90 | 1 | 62999675 | 0.0300884 | 0.0042729 | 1.90E-12 | rs7534572 | G | C | 0.64747 | 0.00430684 | 49.5844137 |
| | 1 | 234852760 | 0.032868 | 0.00408893 | 9.10E-16 | rs553427 | T | C | 0.517037 | 0.00561146 | 64.61296303 |
| | 1 | 109817590 | -0.115226 | 0.00491872 | 2.30E-121 | rs12740374 | T | G | 0.221064 | 0.04745952 | 548.7685606 |
| | 1 | 220970028 | 0.0245558 | 0.00447786 | 4.20E-08 | rs2642438 | G | A | 0.70385 | 0.00261244 | 30.0718125 |

| | | | | | | | | | | | |
|---|---|---|---|---|---|---|---|---|---|---|---|
| 092862 | 1 | 55505647 | −0.306902 | 0.0155777 | 2.10E-86 | rs11591147 | T | G | 0.017457 | 0.003361422 | 388.1371548 |
| | 1 | 55520408 | 0.0325459 | 0.00428294 | 3.00E-14 | rs553741 | C | G | 0.645268 | 0.005001514 | 57.74322595 |
| | 1 | 55526428 | 0.0954747 | 0.0114439 | 7.30E-17 | rs11206517 | G | T | 0.033012 | 0.00604447 | 69.60187655 |
| | 2 | 21288321 | 0.109679 | 0.00530807 | 7.50E-95 | rs562338 | G | A | 0.819616 | 0.003696222 | 426.9393 |
| | 2 | 44072576 | −0.046564 | 0.00437899 | 2.10E-26 | rs4299376 | T | G | 0.676277 | 0.00981564 | 113.069405 |
| | 2 | 21149771 | −0.0260617 | 0.00431772 | 1.60E-09 | rs140798831 | C | T | 0.656814 | 0.00316484 | 36.43250089 |
| | 2 | 27752463 | −0.0374173 | 0.00426502 | 1.70E-18 | rs11127048 | A | G | 0.614116 | 0.00668352 | 76.96534142 |
| | 5 | 74472939 | 0.0380042 | 0.00426072 | 4.70E-19 | rs7707394 | A | G | 0.357075 | 0.00690859 | 79.55906173 |
| | 5 | 156397673 | 0.0338245 | 0.00423678 | 1.40E-15 | rs6882345 | A | G | 0.632836 | 0.00553532 | 63.73579345 |
| | 5 | 52106278 | −0.0677024 | 0.0102771 | 4.50E-11 | rs1499279 | G | T | 0.041623 | 0.0037696 | 43.39697084 |
| | 5 | 74651084 | 0.0544745 | 0.00413968 | 1.50E-39 | rs3846662 | G | A | 0.424111 | 0.01502424 | 173.1591381 |

| | | | | | | | | | | | |
|---|---|---|---|---|---|---|---|---|---|---|---|
| | 5 | 122924864 | 0.0246142 | 0.0041881 | 2.30E-09 | rs1895233 | T | G | 0.544776 | 0.00310231 | 35.71251087 |
| | 6 | 26123502 | -0.0595996 | 0.00786842 | 3.60E-14 | rs115740542 | C | T | 0.073601 | 0.00498296 | 57.3725397 |
| | 6 | 160575366 | 0.0340519 | 0.00543464 | 3.70E-10 | rs11753995 | A | G | 0.171039 | 0.00341025 | 39.2584898 |
| | 6 | 32597435 | 0.0486929 | 0.00609144 | 1.30E-15 | rs140145058 | G | A | 0.135332 | 0.00554936 | 63.897489 |
| | 6 | 160543148 | 0.0555684 | 0.00804242 | 4.90E-12 | rs12208357 | T | C | 0.07 | 0.00414662 | 47.7391561 |
| | 7 | 100216773 | -0.0267997 | 0.00484399 | 3.20E-08 | rs10953298 | T | C | 0.235563 | 0.00265907 | 30.60876743 |
| | 8 | 9187242 | 0.0604263 | 0.00680059 | 6.40E-19 | rs1461729 | G | A | 0.899237 | 0.00685573 | 78.9498433 |
| | 8 | 126506694 | -0.0542615 | 0.00424685 | 2.20E-37 | rs112875651 | A | G | 0.392379 | 0.01416533 | 163.2458686 |
| | 9 | 136149711 | 0.0617664 | 0.00528946 | 1.70E-31 | rs115478735 | T | A | 0.183304 | 0.01183478 | 136.3560328 |
| | 9 | 107661742 | -0.03428 | 0.0046429 | 1.50E-13 | rs2740488 | C | A | 0.265313 | 0.00473467 | 54.51241459 |
| | 11 | 126228000 | 0.0591662 | 0.00785774 | 5.10E-14 | rs59379014 | T | C | 0.072864 | 0.00492414 | 56.6949658 |

| | | | | | | | | | | | |
|---|---|---|---|---|---|---|---|---|---|---|---|
| | 11 | 61557803 | −0.0547939 | 0.00427585 | 1.40E-37 | rs102275 | C | T | 0.34979 | 0.001424928 | 164.2146712 |
| | 11 | 116705516 | −0.0347634 | 0.00452014 | 1.50E-14 | rs525028 | A | G | 0.708551 | 0.000513701 | 59.14706122 |
| | 12 | 9084375 | 0.0288349 | 0.00490775 | 4.20E-09 | rs1805776 | C | T | 0.22603 | 0.000299871 | 34.51949933 |
| | 12 | 111904371 | 0.0267467 | 0.0040097 | 6.60E-11 | rs4766578 | A | T | 0.503234 | 0.000370203 | 42.61877255 |
| | 13 | 32947133 | −0.0274247 | 0.00468157 | 4.70E-09 | rs35596121 | CT | C | 0.287848 | 0.000298101 | 34.31572513 |
| | 14 | 24871926 | 0.0278892 | 0.00412958 | 1.40E-11 | rs11621792 | T | C | 0.453813 | 0.000396169 | 45.60923607 |
| | 15 | 58678720 | −0.0518356 | 0.00431054 | 2.60E-33 | rs261290 | C | T | 0.654645 | 0.001254999 | 144.6056804 |
| | 15 | 58725839 | 0.0414788 | 0.00451301 | 3.90E-20 | rs633695 | G | A | 0.292354 | 0.000733489 | 84.47189321 |
| | 16 | 72101525 | 0.0405592 | 0.00526628 | 1.30E-14 | rs34042070 | G | C | 0.186978 | 0.000515157 | 59.31483418 |
| | 17 | 67191270 | −0.0377324 | 0.00610219 | 6.30E-10 | rs72631343 | G | C | 0.129021 | 0.000332128 | 38.23401094 |
| | 17 | 45766771 | 0.0326299 | 0.00412031 | 6.90E-16 | rs56325564 | A | G | 0.482691 | 0.000565988 | 65.17077538 |

| | | | | | | | | | | | |
|---|---|---|---|---|---|---|---|---|---|---|---|
| | 17 | 67081278 | 0.125526 | 0.0137533 | 7.00E-20 | rs77542162 | G | A | 0.022666 | 0.00723322 | 83.30019142 |
| | 17 | 7080316 | -0.036007 | 0.00520059 | 4.40E-12 | rs55714927 | T | C | 0.190994 | 0.00416371 | 47.93592329 |
| | 19 | 45215081 | -0.135191 | 0.0145174 | 1.30E-20 | rs111278137 | A | G | 0.021614 | 0.0075298 | 86.71821445 |
| | 19 | 45413233 | -0.508624 | 0.00752308 | 1.00E-200 | rs1065853 | T | G | 0.080581 | 0.03820138 3 | 4570.827064 |
| | 19 | 11197261 | -0.146575 | 0.0062778 | 1.50E-120 | rs12151108 | A | G | 0.119677 | 0.00471430 4 | 545.0917831 |
| | 19 | 45173951 | -0.131883 | 0.00947213 | 4.60E-44 | rs28807203 | C | A | 0.049207 | 0.01681682 | 193.8540095 |
| | 19 | 45254168 | 0.0585058 | 0.0061281 | 1.30E-21 | rs8103315 | A | C | 0.135788 | 0.00791398 | 91.14620891 |
| | 19 | 19379549 | -0.0972719 | 0.00777558 | 6.60E-36 | rs58542926 | T | C | 0.074399 | 0.01358037 | 156.4954309 |
| | 19 | 45412955 | 0.163673 | 0.0124743 | 2.50E-39 | rs1081105 | C | A | 0.027621 | 0.01493706 | 172.1528311 |
| | 19 | 11227480 | 0.0300469 | 0.00415996 | 5.10E-13 | rs2738447 | C | A | 0.59318 | 0.00453124 | 52.16911702 |
| | 19 | 45358353 | 0.054952 | 0.00413509 | 2.70E-40 | rs10410835 | C | T | 0.523103 | 0.01532229 | 176.5995231 |

| | | | | | | | | | | | |
|---|---|---|---|---|---|---|---|---|---|---|---|
| | 20 | 39179822 | 0.0901554 | 0.0120132 | 6.20E-14 | rs1883711 | C | G | 0.031207 | 0.000489155 | 56.31946579 |
| | 20 | 17844684 | -0.0244783 | 0.00431829 | 1.40E-08 | rs2618566 | T | G | 0.660294 | 0.000279132 | 32.13152583 |
| **exposure** | **chr.exposure** | **pos.exposure** | **beta.exposure** | **se.exposure** | **pval.exposure** | **SNP** | **effect_allele.exposure** | **other_allele.exposure** | **eaf.exposure** | **R2** | **F** |
| Total lipid levels in medium LDL \|\| id:ebi-a-GCST90092910 | 1 | 55505647 | -0.275282 | 0.0157507 | 2.10E-68 | rs11591147 | T | G | 0.017457 | 0.002647264 | 305.4557908 |
| | 1 | 62999675 | 0.037723 | 0.00432036 | 2.50E-18 | rs7534572 | G | C | 0.64747 | 0.00066203 | 76.23686457 |
| | 1 | 55526428 | 0.0890877 | 0.011571 | 1.40E-14 | rs11206517 | G | T | 0.033012 | 0.000514829 | 59.2769922 |
| | 1 | 109817590 | -0.114057 | 0.00497335 | 2.10E-116 | rs12740374 | T | G | 0.221064 | 0.004549442 | 525.9425326 |
| | 1 | 230295789 | -0.0244731 | 0.00423148 | 7.30E-09 | rs10127775 | T | A | 0.605599 | 0.000290576 | 33.44920783 |
| | 1 | 55518467 | -0.0289783 | 0.00431685 | 1.90E-11 | rs2495477 | G | A | 0.393802 | 0.000391412 | 45.0613651 |
| | 1 | 234852760 | 0.0310149 | 0.00413434 | 6.30E-14 | rs553427 | T | C | 0.517037 | 0.000488775 | 56.27569203 |
| | 2 | 44072576 | -0.0418288 | 0.00442685 | 3.40E-21 | rs4299376 | T | G | 0.676277 | 0.000775206 | 89.2798908 |

| | | | | | | | | | | | |
|---|---|---|---|---|---|---|---|---|---|---|---|
| | 2 | 21232195 | 0.0775471 | 0.00413507 | 1.80E-78 | rs693 | A | G | 0.520889 | 0.003046722 | 351.6882375 |
| | 2 | 27598097 | −0.0588935 | 0.0042513 | 1.20E-43 | rs4665972 | C | T | 0.604609 | 0.001664793 | 191.9038451 |
| | 4 | 3443931 | −0.0314943 | 0.00427972 | 1.90E-13 | rs13108218 | G | A | 0.615355 | 0.0047035 | 54.1533914 |
| | 5 | 122832289 | 0.0245047 | 0.0043978 | 2.50E-08 | rs7704651 | A | G | 0.331143 | 0.00269714 | 31.04704581 |
| | 5 | 74472939 | 0.0317896 | 0.00430715 | 1.60E-13 | rs7707394 | A | G | 0.357075 | 0.00473126 | 54.47316203 |
| | 5 | 52106278 | −0.0683986 | 0.0103891 | 4.60E-11 | rs1499279 | G | T | 0.041623 | 0.00376502 | 43.34420326 |
| | 5 | 74656539 | 0.0499804 | 0.00422035 | 2.30E-32 | rs12916 | C | T | 0.400341 | 0.001217212 | 140.2474486 |
| | 5 | 156443066 | 0.0375673 | 0.00431229 | 3.00E-18 | rs4704834 | G | A | 0.644028 | 0.00659038 | 75.89208767 |
| | 6 | 26098474 | −0.0561052 | 0.00779576 | 6.20E-13 | rs79220007 | C | T | 0.076083 | 0.00449869 | 51.79427533 |
| | 6 | 161111700 | −0.132249 | 0.0174416 | 3.40E-14 | rs186696265 | T | C | 0.014352 | 0.0049933 | 57.49162728 |
| | 6 | 160543148 | 0.0620741 | 0.0081296 | 2.20E-14 | rs12208357 | T | C | 0.07 | 0.00506355 | 58.30085989 |

| | | | | | | | | | | | |
|---|---|---|---|---|---|---|---|---|---|---|---|
| | 6 | 32586376 | 0.0412131 | 0.00451889 | 7.50E-20 | rs11755689 | G | A | 0.334143 | 0.00722247 | 83.17627725 |
| | 6 | 160575366 | 0.0355734 | 0.00549355 | 9.50E-11 | rs11753995 | A | G | 0.171039 | 0.00364233 | 41.93117579 |
| | 7 | 73037366 | -0.0374559 | 0.00514835 | 3.50E-13 | rs55747707 | A | G | 0.203642 | 0.00459724 | 52.92937897 |
| | 8 | 9184691 | 0.0549388 | 0.00687607 | 1.40E-15 | rs6601299 | C | T | 0.899281 | 0.00554408 | 63.83666821 |
| | 8 | 59398461 | -0.0247022 | 0.00438913 | 1.80E-08 | rs10504255 | A | G | 0.662958 | 0.00275161 | 31.67428581 |
| | 8 | 126506694 | -0.0693202 | 0.00429303 | 1.20E-58 | rs112875651 | A | G | 0.392379 | 0.02260485 | 260.7259267 |
| | 9 | 136149711 | 0.0581116 | 0.00534745 | 1.70E-27 | rs115478735 | T | A | 0.183304 | 0.01025131 | 118.0931355 |
| | 9 | 107661742 | -0.0284782 | 0.00469381 | 1.30E-09 | rs2740488 | C | A | 0.265313 | 0.00319763 | 36.81005887 |
| | 11 | 116648917 | -0.0826827 | 0.00609378 | 6.20E-42 | rs964184 | C | G | 0.86726 | 0.01597181 | 184.0976354 |
| | 11 | 61557803 | -0.0290753 | 0.00432308 | 1.70E-11 | rs102275 | C | T | 0.34979 | 0.00392902 | 45.23288794 |
| | 11 | 126244955 | 0.0534572 | 0.00673628 | 2.10E-15 | rs4307732 | A | G | 0.105933 | 0.00546924 | 62.97448226 |

| | | | | | | | | | | | |
|---|---|---|---|---|---|---|---|---|---|---|---|
| | 12 | 9172646 | 0.0253629 | 0.00421525 | 1.80E-09 | rs10842724 | C | T | 0.412903 | 0.00314449 | 36.2029197 |
| | 13 | 114547372 | 0.0238333 | 0.00430484 | 3.10E-08 | rs6602911 | T | C | 0.360028 | 0.00266276 | 30.65115066 |
| | 14 | 24871926 | 0.0299337 | 0.00417453 | 7.50E-13 | rs11621792 | T | C | 0.453813 | 0.00446585 | 51.41597587 |
| | 16 | 56988044 | -0.0398384 | 0.00438532 | 1.00E-19 | rs173539 | T | C | 0.331294 | 0.00716609 | 82.52654218 |
| | 16 | 72101525 | 0.0458758 | 0.00532357 | 6.80E-18 | rs34042070 | G | C | 0.186978 | 0.00644872 | 74.25980694 |
| | 17 | 67191270 | -0.0378675 | 0.00616893 | 8.30E-10 | rs72631343 | G | C | 0.129021 | 0.00327314 | 37.67957851 |
| | 17 | 45766771 | 0.0331451 | 0.00416537 | 1.80E-15 | rs56325564 | A | G | 0.482691 | 0.00549902 | 63.31752784 |
| | 17 | 7080316 | -0.0423013 | 0.00525746 | 8.60E-16 | rs55714927 | T | C | 0.190994 | 0.00562217 | 64.73630789 |
| | 17 | 67081278 | 0.102215 | 0.0139037 | 2.00E-13 | rs77542162 | G | A | 0.022666 | 0.00469415 | 54.04567452 |
| | 19 | 45358353 | 0.0494719 | 0.00418318 | 2.90E-32 | rs10410835 | C | T | 0.523103 | 0.01213862 | 139.8609673 |
| | 19 | 19379549 | -0.111477 | 0.00786601 | 1.40E-45 | rs58542926 | T | C | 0.074399 | 0.01742195 | 200.8417558 |

| | | | | | | | | | | | |
|---|---|---|---|---|---|---|---|---|---|---|---|
| | 19 | 45412955 | 0.161867 | 0.0126194 | 1.20E-37 | rs1081105 | C | A | 0.027621 | 0.001427616 | 164.5249149 |
| | 19 | 45215081 | -0.128561 | 0.0146862 | 2.10E-18 | rs11278137 | A | G | 0.021614 | 0.000665431 | 76.62880619 |
| | 19 | 45413233 | -0.455186 | 0.00761057 | 1.00E-200 | rs1065853 | T | G | 0.080581 | 0.030146854 | 3577.139484 |
| | 19 | 11197261 | -0.135066 | 0.00635102 | 2.30E-100 | rs12151108 | A | G | 0.119677 | 0.003914662 | 452.2697785 |
| | 19 | 11227480 | 0.0294324 | 0.00420834 | 2.70E-12 | rs2738447 | C | A | 0.59318 | 0.00424852 | 48.91275967 |
| | 19 | 45173951 | -0.121579 | 0.0095823 | 6.90E-37 | rs28807203 | C | A | 0.049207 | 0.001396893 | 160.9793091 |
| | 19 | 45254168 | 0.0515794 | 0.00619937 | 8.80E-17 | rs8103315 | A | C | 0.135788 | 0.000601158 | 69.22291912 |
| | 20 | 39678289 | 0.0278556 | 0.00414008 | 1.70E-11 | rs6016505 | T | C | 0.549538 | 0.000393214 | 45.26890826 |
| | 20 | 44551855 | 0.0699226 | 0.00518765 | 2.10E-41 | rs6073958 | C | T | 0.198731 | 0.001576164 | 181.6712711 |
| | 20 | 39186590 | -0.0274738 | 0.00446861 | 7.80E-10 | rs6102034 | G | C | 0.323224 | 0.000328354 | 37.79940912 |
| exposure | chr.exposure | pos.exposure | beta.exposure | se.exposure | pval.exposure | SNP | effect_allele.exposure | other_allele.exposure | eaf.exposure | R2 | F |

| | | | | | | | | | | | |
|---|---|---|---|---|---|---|---|---|---|---|---|
| Total lipid levels in small LDL \|\| id:ebi-a-GCST90092962 | 1 | 25622291 | -0.0256176 | 0.00451119 | 1.40E-08 | rs2986164 | A | G | 0.535826 | 0.00280134 | 32.24683497 |
| | 1 | 55520408 | 0.0313061 | 0.00432277 | 4.40E-13 | rs553741 | C | G | 0.645268 | 0.00455542 | 52.44766176 |
| | 1 | 109817590 | -0.126975 | 0.00496446 | 2.80E-144 | rs12740374 | T | G | 0.221064 | 0.05652275 | 654.161324 |
| | 1 | 230307182 | -0.0300771 | 0.00436259 | 5.40E-12 | rs4846922 | C | T | 0.654699 | 0.00412854 | 47.53084911 |
| | 1 | 55505647 | -0.305666 | 0.0157226 | 3.50E-84 | rs11591147 | T | G | 0.017457 | 0.03273513 | 377.9531668 |
| | 1 | 55526428 | 0.0922566 | 0.0115504 | 1.40E-15 | rs11206517 | G | T | 0.033012 | 0.00554055 | 63.79597715 |
| | 1 | 62999675 | 0.0307136 | 0.00431264 | 1.10E-12 | rs7534572 | G | C | 0.64747 | 0.00440531 | 50.71862683 |
| | 1 | 234852760 | 0.0321215 | 0.00412695 | 7.10E-15 | rs553427 | T | C | 0.517037 | 0.00526135 | 60.57949842 |
| | 2 | 21149771 | -0.0371066 | 0.00435734 | 1.70E-17 | rs140798831 | C | T | 0.656814 | 0.00629765 | 72.51901221 |
| | 2 | 21288321 | 0.109297 | 0.00535678 | 1.60E-92 | rs562338 | G | A | 0.819616 | 0.03604402 | 416.2951147 |
| | 2 | 27752463 | -0.0499132 | 0.00430415 | 4.30E-31 | rs11127048 | A | G | 0.614116 | 0.0116719 | 134.4771461 |

| | | | | | | | | | | | |
|---|---|---|---|---|---|---|---|---|---|---|---|
| | 2 | 44072576 | −0.045335 | 0.00441917 | 1.10E-24 | rs4299376 | T | G | 0.676277 | 0.00913653 | 105.2393786 |
| | 4 | 3443931 | −0.0270612 | 0.00427279 | 2.40E-10 | rs13108218 | G | A | 0.615355 | 0.00348427 | 40.11099703 |
| | 5 | 52106278 | −0.0668698 | 0.0103717 | 1.10E-10 | rs1499279 | G | T | 0.041623 | 0.00361074 | 41.56737583 |
| | 5 | 74472939 | 0.0360122 | 0.00429996 | 5.50E-17 | rs7707394 | A | G | 0.357075 | 0.00609114 | 70.13954237 |
| | 5 | 156443066 | 0.0432055 | 0.00430509 | 1.10E-23 | rs4704834 | G | A | 0.644028 | 0.00874432 | 100.7177588 |
| | 5 | 74656539 | 0.0560544 | 0.0042133 | 2.20E-40 | rs12916 | C | T | 0.400341 | 0.01535677 | 176.9974878 |
| | 6 | 16134247 | −0.0240462 | 0.00415548 | 7.20E-09 | rs6920309 | T | C | 0.456849 | 0.00290882 | 33.48443455 |
| | 6 | 160575366 | 0.035093 | 0.00548389 | 1.60E-10 | rs11753995 | A | G | 0.171039 | 0.00355715 | 40.95019513 |
| | 6 | 26123502 | −0.0604767 | 0.00793972 | 2.60E-14 | rs115740542 | C | T | 0.073601 | 0.00503894 | 58.01739819 |
| | 6 | 160543148 | 0.0640823 | 0.0081153 | 2.90E-15 | rs12208357 | T | C | 0.07 | 0.00541532 | 62.35330467 |
| | 6 | 32586376 | 0.0410279 | 0.00451094 | 9.40E-20 | rs11755689 | G | A | 0.334143 | 0.00718299 | 82.72121941 |

| | | | | | | | | | | | |
|---|---|---|---|---|---|---|---|---|---|---|---|
| | 6 | 161111700 | -0.144396 | 0.017411 | 1.10E-16 | rs186696265 | T | C | 0.014352 | 0.00597305 | 68.77891918 |
| | 7 | 73037366 | -0.029034 | 0.00514032 | 1.60E-08 | rs55747707 | A | G | 0.203642 | 0.00277144 | 31.90258581 |
| | 8 | 19824492 | -0.0261715 | 0.00456425 | 9.80E-09 | rs13702 | C | T | 0.287843 | 0.00285619 | 32.87841199 |
| | 8 | 59398461 | -0.0255669 | 0.00438204 | 5.40E-09 | rs10504255 | A | G | 0.662958 | 0.00295711 | 34.0404994 |
| | 8 | 126506694 | -0.0649649 | 0.0042861 | 6.80E-52 | rs112875651 | A | G | 0.392379 | 0.01992322 | 229.7340675 |
| | 8 | 9184691 | 0.0474781 | 0.00686497 | 4.60E-12 | rs6601299 | C | T | 0.899281 | 0.00415453 | 47.83015923 |
| | 9 | 136149711 | 0.0493035 | 0.00533905 | 2.60E-20 | rs115478735 | T | A | 0.183304 | 0.00740455 | 85.27465621 |
| | 9 | 107661742 | -0.0257859 | 0.00468643 | 3.70E-08 | rs2740488 | C | A | 0.265313 | 0.00263002 | 30.27420669 |
| | 11 | 61557803 | -0.0331746 | 0.00431584 | 1.50E-14 | rs102275 | C | T | 0.34979 | 0.00513157 | 59.08444643 |
| | 11 | 126244955 | 0.0519899 | 0.006725 | 1.10E-14 | rs4307732 | A | G | 0.105933 | 0.00519064 | 59.76485074 |
| | 11 | 116648917 | -0.0748199 | 0.00608357 | 9.20E-35 | rs964184 | C | G | 0.86726 | 0.01312621 | 151.2549791 |

| | | | | | | | | | | | |
|---|---|---|---|---|---|---|---|---|---|---|---|
| | 12 | 9172646 | 0.0252371 | 0.00420829 | 2.00E-09 | rs10842724 | C | T | 0.412903 | 0.00031241 | 35.96334086 |
| | 12 | 111904371 | 0.0247314 | 0.00413486 | 2.20E-09 | rs4766578 | A | T | 0.503234 | 0.000310766 | 35.77406045 |
| | 13 | 114551993 | 0.0264113 | 0.00439737 | 1.90E-09 | rs6602909 | C | T | 0.328942 | 0.000313365 | 36.07330971 |
| | 13 | 32947133 | -0.0270029 | 0.0047251 | 1.10E-08 | rs35596121 | CT | C | 0.287848 | 0.000283706 | 32.658126 |
| | 14 | 24871926 | 0.0284752 | 0.0041679 | 8.40E-12 | rs11621792 | T | C | 0.453813 | 0.00040543 | 46.67576426 |
| | 16 | 56988044 | -0.0242127 | 0.00437832 | 3.20E-08 | rs173539 | T | C | 0.331294 | 0.000265674 | 30.58185993 |
| | 16 | 72101525 | 0.0476163 | 0.00531507 | 3.30E-19 | rs34042070 | G | C | 0.186978 | 0.00069692 | 80.25752573 |
| | 17 | 7080316 | -0.0373731 | 0.00524896 | 1.10E-12 | rs55714927 | T | C | 0.190994 | 0.000440325 | 50.69488845 |
| | 17 | 45766771 | 0.0341996 | 0.00415864 | 2.00E-16 | rs56325564 | A | G | 0.482691 | 0.000587323 | 67.62882663 |
| | 17 | 67191270 | -0.0389993 | 0.00615895 | 2.40E-10 | rs72631343 | G | C | 0.129021 | 0.000348178 | 40.08227793 |
| | 17 | 67081278 | 0.117284 | 0.0138812 | 2.90E-17 | rs77542162 | G | A | 0.022666 | 0.000619936 | 71.38647999 |

| | | | | | | | | | | | |
|---|---|---|---|---|---|---|---|---|---|---|---|
| | 19 | 19379549 | -0.111197 | 0.00784572 | 1.30E-45 | rs58542926 | T | C | 0.074399 | 0.01742432 | 200.8690306 |
| | 19 | 45173951 | -0.138682 | 0.00955758 | 1.00E-47 | rs28807203 | C | A | 0.049207 | 0.01826178 | 210.5410386 |
| | 19 | 45412955 | 0.186329 | 0.0125868 | 1.40E-49 | rs1081105 | C | A | 0.027621 | 0.01900626 | 219.1405244 |
| | 19 | 45358353 | 0.0573978 | 0.00417239 | 4.70E-43 | rs10410835 | C | T | 0.523103 | 0.01641722 | 189.2400935 |
| | 19 | 45413233 | -0.525217 | 0.00759094 | 1.00E-200 | rs1065853 | T | G | 0.080581 | 0.03993729 4 | 4787.170376 |
| | 19 | 11197261 | -0.148601 | 0.00633463 | 1.10E-121 | rs12151108 | A | G | 0.119677 | 0.00475906 7 | 550.2922267 |
| | 19 | 11227480 | 0.0341762 | 0.00419748 | 3.90E-16 | rs2738447 | C | A | 0.59318 | 0.00575722 | 66.29224495 |
| | 19 | 45215081 | -0.135049 | 0.0146483 | 3.00E-20 | rs11278137 | A | G | 0.021614 | 0.00738041 | 84.99644197 |
| | 19 | 45254168 | 0.0594464 | 0.00618338 | 7.00E-22 | rs8103315 | A | C | 0.135788 | 0.00802497 | 92.42547631 |
| | 20 | 44545048 | -0.0327459 | 0.00476756 | 6.50E-12 | rs4810479 | T | C | 0.750217 | 0.00409766 | 47.17522499 |
| | 20 | 39179822 | 0.0937438 | 0.0121247 | 1.10E-14 | rs1883711 | C | G | 0.031207 | 0.00519171 | 59.77719843 |

| exposure | chr.exposure | pos.exposure | beta.exposure | se.exposure | pval.exposure | SNP | effect_allele.exposure | other_allele.exposure | eaf.exposure | R2 | F |
|---|---|---|---|---|---|---|---|---|---|---|---|
| Free cholesterol to total lipids ratio in chylomicrons and extremely large VLDL \|\| id:ebi-a-GCST90093045 | 1 | 27262545 | -0.0682279 | 0.0124534 | 4.30E-08 | rs182050989 | T | C | 0.028184 | 0.00268794 | 30.01513856 |
| | 2 | 27730940 | 0.0305836 | 0.0042046 4 | 3.50E-13 | rs1260326 | C | T | 0.603361 | 0.00473698 | 52.90683651 |
| | 5 | 55804552 | -0.0320896 | 0.0048971 2 | 5.60E-11 | rs40270 | C | A | 0.771897 | 0.00384475 | 42.93775507 |
| | 6 | 160711566 | -0.0388593 | 0.0047290 2 | 2.10E-16 | rs12179053 | T | C | 0.253029 | 0.00604468 | 67.52115525 |
| | 6 | 160710851 | 0.0792444 | 0.0056572 4 | 1.40E-44 | rs3127580 | T | C | 0.155659 | 0.017545 | 196.2095653 |
| | 6 | 161080457 | -0.0823436 | 0.014401 | 1.10E-08 | rs117857195 | T | G | 0.024355 | 0.00292776 | 32.69389165 |
| | 6 | 161339399 | -0.0726264 | 0.0121426 | 2.20E-09 | rs1247304 | G | A | 0.030815 | 0.00320342 | 35.77320708 |
| | 6 | 161010118 | 0.266534 | 0.0076670 9 | 1.00E-200 | rs10455872 | G | A | 0.077845 | 0.10709176 | 1208.471273 |
| | 6 | 32622063 | -0.0259386 | 0.0044867 3 | 7.40E-09 | rs17613367 | A | G | 0.698912 | 0.00299289 | 33.42145956 |
| | 6 | 160578069 | 0.133105 | 0.0179002 | 1.00E-13 | rs146534110 | T | G | 0.013369 | 0.00495046 | 55.29236831 |

| | | | | | | | | | | | |
|---|---|---|---|---|---|---|---|---|---|---|---|
| | 6 | 160922870 | 0.340251 | 0.0154875 | 5.70E-107 | rs117733303 | G | A | 0.017933 | 0.004304776 | 482.6456555 |
| | 6 | 161017363 | 0.150418 | 0.0114913 | 3.80E-39 | rs73596816 | A | G | 0.033259 | 0.001532439 | 171.3379292 |
| | 7 | 1117436 | -0.0264007 | 0.00430139 | 8.40E-10 | rs10274367 | C | T | 0.355358 | 0.0033733 | 37.67085363 |
| | 7 | 25983400 | 0.0261407 | 0.0042957 | 1.20E-09 | rs56001710 | T | A | 0.580369 | 0.00331597 | 37.03043024 |
| | 8 | 19881801 | 0.0516845 | 0.00687445 | 5.50E-14 | rs60223219 | G | A | 0.099172 | 0.00506073 | 56.52452779 |
| | 8 | 126500031 | 0.0324658 | 0.00421944 | 1.40E-14 | rs28601761 | G | C | 0.419999 | 0.0053003 | 59.20178564 |
| | 9 | 136149711 | 0.0462328 | 0.00532134 | 3.70E-18 | rs115478735 | T | A | 0.182911 | 0.00675698 | 75.4832508 |
| | 11 | 116648917 | 0.0441827 | 0.00604198 | 2.60E-13 | rs964184 | C | G | 0.866291 | 0.00478769 | 53.4734441 |
| | 11 | 61581368 | -0.0289204 | 0.00444312 | 7.60E-11 | rs174557 | G | A | 0.307577 | 0.00379364 | 42.3667085 |
| | 11 | 116916060 | -0.126067 | 0.0206764 | 1.10E-09 | rs144018203 | C | G | 0.010796 | 0.00332887 | 37.17450592 |
| | 15 | 58568394 | -0.0264191 | 0.00420259 | 3.20E-10 | rs77732800 | G | T | 0.483464 | 0.00353864 | 39.51793217 |

| | | | | | | | | | | | |
|---|---|---|---|---|---|---|---|---|---|---|---|
| | 15 | 58723426 | 0.12267 | 0.00498685 | 1.30E-133 | rs1077835 | G | A | 0.218143 | 0.00539094 | 605.0849341 |
| | 15 | 58683366 | -0.108126 | 0.00422597 | 2.20E-144 | rs1532085 | G | A | 0.615082 | 0.005829833 | 654.635683 |
| | 16 | 56993886 | -0.0487638 | 0.00439543 | 1.30E-28 | rs821840 | G | A | 0.324458 | 0.0110129 | 123.0791721 |
| | 17 | 64210580 | 0.135524 | 0.0120094 | 1.60E-29 | rs1801689 | C | A | 0.0302 | 0.01139417 | 127.3450364 |
| | 19 | 45430280 | -0.0469839 | 0.00442298 | 2.30E-26 | rs5112 | G | C | 0.535256 | 0.01009758 | 112.8393031 |
| | 19 | 19366632 | 0.0729289 | 0.00851491 | 1.10E-17 | rs72999033 | T | C | 0.063455 | 0.00656662 | 73.35528719 |
| | 19 | 18244690 | -0.0286749 | 0.00470238 | 1.10E-09 | rs885683 | A | G | 0.256772 | 0.00332975 | 37.18438478 |
| | 20 | 44545773 | 0.0477534 | 0.00526851 | 1.30E-19 | rs73307905 | C | A | 0.187284 | 0.00735362 | 82.15333513 |
| | 22 | 44324730 | 0.046922 | 0.00498841 | 5.10E-21 | rs738408 | T | C | 0.216638 | 0.00791904 | 88.47508072 |
| exposure | chr.exposure | pos.exposure | beta.exposure | se.exposure | pval.exposure | SNP | effect_allele.exposure | other_allele.exposure | eaf.exposure | R2 | F |
| Free choles | 1 | 55505647 | -0.216328 | 0.0152657 | 1.40E-45 | rs1591147 | T | G | 0.017401 | 0.01755961 | 200.809574 |

| | | | | | | | | | | | |
|---|---|---|---|---|---|---|---|---|---|---|---|
| terol to total lipids ratio in very large VLDL \|\| id: ebi-a-GCST90093021 | 1 | 55521109 | 0.0242126 | 0.00418467 | 7.20E-09 | rs693668 | A | G | 0.648668 | 0.00293177 | 33.47748431 |
| | 1 | 110232983 | -0.0256907 | 0.00447691 | 9.60E-09 | rs140584594 | G | A | 0.73012 | 0.00288374 | 32.92968219 |
| | 1 | 109821511 | 0.0419109 | 0.00484207 | 4.90E-18 | rs602633 | G | T | 0.78309 | 0.00655833 | 74.91766591 |
| | 1 | 230302838 | 0.0254225 | 0.00414849 | 8.90E-10 | rs59484402 | CT | C | 0.6115 | 0.00328851 | 37.55336067 |
| | 2 | 21267461 | 0.0485121 | 0.00422594 | 1.70E-30 | rs934197 | A | G | 0.336053 | 0.01153024 | 131.778903 |
| | 2 | 227106217 | -0.0360663 | 0.00417147 | 5.30E-18 | rs2972138 | G | A | 0.645844 | 0.00654375 | 74.75105455 |
| | 2 | 27730940 | 0.0778523 | 0.00408504 | 5.60E-81 | rs1260326 | C | T | 0.60381 | 0.03171442 | 363.1973404 |
| | 2 | 165514409 | 0.0289807 | 0.00420943 | 5.80E-12 | rs11394662 | AT | A | 0.384489 | 0.00415028 | 47.39838563 |
| | 2 | 121337280 | 0.0329637 | 0.00546958 | 1.70E-09 | rs9677933 | T | C | 0.158637 | 0.00318062 | 36.32087811 |
| | 2 | 44073881 | -0.0274781 | 0.00427539 | 1.30E-10 | rs6544713 | C | T | 0.67595 | 0.00361702 | 41.30611184 |
| | 3 | 186332182 | 0.0233098 | 0.00416038 | 2.10E-08 | rs2518134 | T | C | 0.632583 | 0.00274902 | 31.39087966 |

| | | | | | | | | | | | |
|---|---|---|---|---|---|---|---|---|---|---|---|
| | 4 | 3365381 | −0.0250865 | 0.00407818 | 7.70E-10 | rs881817 | G | A | 0.593646 | 0.00331352 | 37.83900941 |
| | 4 | 103895398 | 0.0227927 | 0.00399442 | 1.20E-08 | rs7659650 | A | T | 0.490612 | 0.00285132 | 32.55940688 |
| | 5 | 55868097 | −0.0321857 | 0.00451187 | 9.80E-13 | rs13179413 | T | C | 0.285561 | 0.00445559 | 50.88680144 |
| | 5 | 55804552 | −0.03554 | 0.00476272 | 8.50E-14 | rs40270 | C | A | 0.771812 | 0.00487527 | 55.68229645 |
| | 5 | 74472939 | 0.026638 | 0.00416621 | 1.60E-10 | rs7707394 | A | G | 0.357225 | 0.00357974 | 40.88022774 |
| | 5 | 74619639 | 0.0377805 | 0.00422219 | 3.60E-19 | rs79225634 | T | C | 0.340502 | 0.00700876 | 80.06675883 |
| | 6 | 32603825 | −0.0397338 | 0.00418044 | 2.00E-21 | rs3129774 | T | C | 0.440113 | 0.00790713 | 90.33761178 |
| | 6 | 43758873 | −0.028711 | 0.00402654 | 1.00E-12 | rs6905288 | A | G | 0.568457 | 0.00445169 | 50.84227721 |
| | 6 | 160922870 | 0.194224 | 0.0149001 | 7.70E-39 | rs117733303 | G | A | 0.018321 | 0.01486167 | 169.9103417 |
| | 6 | 161017363 | 0.0792551 | 0.0111478 | 1.20E-12 | rs73596816 | A | G | 0.033484 | 0.00442558 | 50.54386992 |
| | 6 | 32624128 | −0.0331093 | 0.00527125 | 3.40E-10 | rs184252375 | T | C | 0.311401 | 0.00345469 | 39.45164906 |

| | | | | | | | | | | | |
|---|---|---|---|---|---|---|---|---|---|---|---|
| | 6 | 160710851 | 0.0389959 | 0.00549578 | 1.30E-12 | rs3127580 | T | C | 0.156186 | 0.00440833 | 50.34676946 |
| | 6 | 161010118 | 0.15373 | 0.00741865 | 2.20E-95 | rs10455872 | G | A | 0.078784 | 0.00374739 | 429.3978655 |
| | 7 | 73033559 | 0.104963 | 0.00594018 | 7.10E-70 | rs34062580 | A | G | 0.130239 | 0.00272753 | 312.2235454 |
| | 7 | 1029585 | -0.0308468 | 0.0041184 | 6.90E-14 | rs9769088 | C | T | 0.390947 | 0.00491175 | 56.09905897 |
| | 7 | 25983400 | 0.0285592 | 0.00417595 | 8.00E-12 | rs56001710 | T | A | 0.580556 | 0.00409534 | 46.77070183 |
| | 7 | 28256240 | 0.0280485 | 0.00415952 | 1.50E-11 | rs498475 | A | G | 0.630615 | 0.00398149 | 45.47002983 |
| | 8 | 11105077 | -0.0340463 | 0.00403316 | 3.10E-17 | rs2572431 | T | C | 0.569443 | 0.00623827 | 71.25926394 |
| | 8 | 126507389 | 0.0336625 | 0.00435907 | 1.10E-14 | rs2954038 | A | C | 0.698612 | 0.00522113 | 59.63448017 |
| | 8 | 19820433 | 0.0804589 | 0.0066506 | 1.10E-33 | rs12679834 | C | T | 0.099996 | 0.01280431 | 146.3588259 |
| | 9 | 136149711 | 0.0570569 | 0.00517345 | 2.80E-28 | rs115478735 | T | A | 0.183191 | 0.01064338 | 121.6321051 |
| | 10 | 95342772 | -0.0380227 | 0.00632752 | 1.90E-09 | rs7091593 | T | A | 0.113962 | 0.00316204 | 36.10864938 |

| | | | | | | | | | | | |
|---|---|---|---|---|---|---|---|---|---|---|---|
| | 10 | 65104500 | 0.0317484 | 0.00407934 | 7.10E-15 | rs7896518 | G | A | 0.428268 | 0.00530297 | 60.56981823 |
| | 11 | 116916060 | -0.124055 | 0.0201952 | 8.10E-10 | rs144018203 | C | G | 0.0107 | 0.00330426 | 37.73328541 |
| | 11 | 5701074 | 0.0445647 | 0.00778554 | 1.00E-08 | rs11601507 | A | C | 0.069236 | 0.00286923 | 32.76399759 |
| | 11 | 61552680 | -0.0686296 | 0.00420079 | 5.40E-60 | rs174537 | T | G | 0.343434 | 0.00233256 | 266.9029561 |
| | 11 | 63988045 | -0.0510233 | 0.00852387 | 2.20E-09 | rs3802932 | A | G | 0.058725 | 0.00313771 | 35.83074547 |
| | 11 | 65473798 | -0.0277309 | 0.00440463 | 3.10E-10 | rs10750766 | A | C | 0.709801 | 0.00347092 | 39.637062 |
| | 11 | 116648917 | 0.0965108 | 0.00588676 | 2.10E-60 | rs964184 | C | G | 0.866906 | 0.00248897 | 268.7766658 |
| | 12 | 112013941 | -0.0262954 | 0.00475591 | 3.20E-08 | rs61941261 | C | T | 0.738662 | 0.00267708 | 30.56924408 |
| | 12 | 4384844 | 0.0886878 | 0.0155549 | 1.20E-08 | rs76895963 | G | T | 0.020551 | 0.00284679 | 32.50765996 |
| | 14 | 94844947 | 0.0984874 | 0.0143188 | 6.10E-12 | rs28929474 | T | C | 0.019985 | 0.00414242 | 47.30864294 |
| | 15 | 58683366 | -0.0841471 | 0.00410632 | 2.50E-93 | rs1532085 | G | A | 0.614243 | 0.00366491 | 419.9186053 |

| | | | | | | | | | | | |
|---|---|---|---|---|---|---|---|---|---|---|---|
| | 15 | 44363739 | -0.0771824 | 0.0128419 | 1.90E-09 | rs144022883 | A | G | 0.024842 | 0.0031632 | 36.12188301 |
| | 15 | 58602300 | 0.0269363 | 0.00401004 | 1.90E-11 | rs261265 | A | C | 0.454485 | 0.00395087 | 45.12018406 |
| | 15 | 58723479 | 0.106696 | 0.00484342 | 1.50E-107 | rs1077834 | C | T | 0.218263 | 0.00432879 | 485.2710757 |
| | 16 | 72144174 | -0.034967 | 0.00516538 | 1.30E-11 | rs9302635 | C | T | 0.182333 | 0.00401258 | 45.8252228 |
| | 16 | 56991363 | -0.0692196 | 0.00427601 | 6.10E-59 | rs183130 | T | C | 0.323372 | 0.00229087 | 262.0433468 |
| | 16 | 15148646 | 0.0261861 | 0.00438807 | 2.40E-09 | rs11075253 | A | C | 0.29562 | 0.0031185 | 35.61122731 |
| | 16 | 31283744 | 0.0351805 | 0.00643407 | 4.60E-08 | rs8056944 | G | C | 0.110188 | 0.00261821 | 29.89680945 |
| | 16 | 81534790 | 0.0266503 | 0.0043554 | 9.40E-10 | rs2925979 | C | T | 0.698804 | 0.00327862 | 37.44040192 |
| | 17 | 41840849 | -0.0874365 | 0.0119526 | 2.60E-13 | rs77697917 | T | C | 0.02874 | 0.00468536 | 53.51224231 |
| | 17 | 41163974 | -0.0536107 | 0.00911771 | 4.10E-09 | rs111351217 | A | G | 0.052287 | 0.00302751 | 34.57194374 |
| | 17 | 64210580 | 0.128884 | 0.0116329 | 1.60E-28 | rs1801689 | C | A | 0.030448 | 0.01074092 | 122.7479971 |

| | | | | | | | | | | | |
|---|---|---|---|---|---|---|---|---|---|---|---|
| | 17 | 67081278 | 0.140079 | 0.0134468 | 2.10E-25 | rs77542162 | G | A | 0.022652 | 0.00949689 | 108.5176964 |
| | 19 | 11192915 | -0.0937437 | 0.00613416 | 1.00E-52 | rs73015021 | G | A | 0.12061 | 0.002041608 | 233.5427413 |
| | 19 | 7197880 | 0.0258846 | 0.00442882 | 5.10E-09 | rs66921136 | C | T | 0.711844 | 0.00299132 | 34.15853595 |
| | 19 | 45173951 | -0.05845 | 0.00929643 | 3.20E-10 | rs28807203 | C | A | 0.04906 | 0.00346156 | 39.53020034 |
| | 19 | 45337918 | 0.0866785 | 0.0109639 | 2.70E-15 | rs147711004 | A | G | 0.03605 | 0.00547194 | 62.50072197 |
| | 19 | 11275842 | 0.0274852 | 0.00427345 | 1.30E-10 | rs3185010 | A | G | 0.327283 | 0.00362217 | 41.3649916 |
| | 19 | 45413233 | -0.186309 | 0.0073909 | 3.30E-140 | rs1065853 | T | G | 0.08013 | 0.005535392 | 635.4266254 |
| | 20 | 44559748 | -0.0978465 | 0.00514102 | 9.20E-81 | rs139953093 | T | G | 0.186266 | 0.003163022 | 362.229983 |
| | 20 | 17844684 | -0.024008 | 0.00422292 | 1.30E-08 | rs2618566 | T | G | 0.660275 | 0.00283041 | 32.32054296 |
| | 22 | 44379565 | 0.0279289 | 0.00493516 | 1.50E-08 | rs2294922 | C | G | 0.207878 | 0.00280459 | 32.02562242 |
| exposure | chr.exposure | pos.exposure | beta.exposure | se.exposure | pval.exposure | SNP | effect_allele.exposure | other_allele.exposure | eaf.exposure | R2 | F |

| | | | | | | | | | | | |
|---|---|---|---|---|---|---|---|---|---|---|---|
| Free cholesterol to total lipids ratio in large VLDL \|\| id:ebi-a-GCST90092873 | 1 | 182136401 | 0.0309269 | 0.00434526 | 1.10E-12 | rs61805075 | A | G | 0.329468 | 0.00440032 | 50.65632697 |
| | 1 | 55505647 | -0.184283 | 0.015583 | 2.90E-32 | rs11591147 | T | G | 0.017454 | 0.01213878 | 139.8494806 |
| | 1 | 109817192 | -0.0532758 | 0.00491639 | 2.30E-27 | rs7528419 | G | A | 0.222154 | 0.01019432 | 117.4247808 |
| | 1 | 230293530 | -0.0272658 | 0.00421362 | 9.70E-11 | rs4631704 | T | C | 0.608323 | 0.00363749 | 41.87146996 |
| | 2 | 44058201 | -0.0526926 | 0.00856348 | 7.60E-10 | rs77725792 | A | G | 0.06142 | 0.0032892 | 37.86091739 |
| | 2 | 21267461 | 0.0605181 | 0.00432325 | 1.60E-44 | rs934197 | A | G | 0.335698 | 0.01699985 | 195.9486703 |
| | 5 | 74619639 | 0.024621 | 0.00431618 | 1.20E-08 | rs79225634 | T | C | 0.340372 | 0.00282699 | 32.53903971 |
| | 6 | 160922870 | 0.107839 | 0.0151688 | 1.20E-12 | rs117733303 | G | A | 0.018497 | 0.00439028 | 50.54075265 |
| | 6 | 160985526 | 0.103028 | 0.00761838 | 1.10E-41 | rs118039278 | A | G | 0.078841 | 0.01586827 | 182.8848489 |
| | 6 | 160710851 | 0.0475178 | 0.00562224 | 2.90E-17 | rs3127580 | T | C | 0.156219 | 0.00620381 | 71.43093599 |
| | 6 | 32634681 | 0.040709 | 0.00568356 | 7.90E-13 | rs71542468 | T | G | 0.153799 | 0.00445636 | 51.30177858 |

| | | | | | | | | | | | |
|---|---|---|---|---|---|---|---|---|---|---|---|
| | 6 | 34689725 | −0.0240768 | 0.00431233 | 2.40E-08 | rs34265539 | ATT | A | 0.643939 | 0.00270826 | 31.17209611 |
| | 8 | 19824492 | −0.034316 | 0.00452077 | 3.20E-14 | rs13702 | C | T | 0.287835 | 0.00500478 | 57.61836698 |
| | 8 | 9183358 | −0.0446104 | 0.00711488 | 3.60E-10 | rs9987289 | G | A | 0.909165 | 0.00341525 | 39.3123972 |
| | 8 | 126506694 | −0.0428429 | 0.00424525 | 6.00E-24 | rs112875651 | A | G | 0.392369 | 0.00884303 | 101.8459188 |
| | 9 | 136149711 | 0.0771557 | 0.00528685 | 3.10E-48 | rs115478735 | T | A | 0.183296 | 0.01847452 | 212.977883 |
| | 9 | 15303583 | −0.0401284 | 0.00594927 | 1.50E-11 | rs1215112 | A | G | 0.863021 | 0.0039522 | 45.49553546 |
| | 9 | 107589255 | −0.0723144 | 0.0121605 | 2.70E-09 | rs2066718 | T | C | 0.029065 | 0.00307219 | 35.3621907 |
| | 9 | 139369062 | −0.0294084 | 0.00447233 | 4.80E-11 | rs3812595 | T | C | 0.263629 | 0.00375618 | 43.23819567 |
| | 10 | 115786233 | −0.0447513 | 0.00616515 | 3.90E-13 | rs72823013 | A | G | 0.126336 | 0.00457677 | 52.68857685 |
| | 11 | 117156893 | −0.132259 | 0.0207895 | 2.00E-10 | rs1047964 | T | G | 0.010074 | 0.00351596 | 40.47201546 |
| | 11 | 61603510 | 0.0296412 | 0.00427776 | 4.20E-12 | rs174576 | A | C | 0.350046 | 0.00417073 | 48.01215336 |

| | | | | | | | | | | | |
|---|---|---|---|---|---|---|---|---|---|---|---|
| | 11 | 47269159 | -0.0357304 | 0.00520642 | 6.80E-12 | rs4752973 | C | T | 0.189262 | 0.000409123 | 47.09662745 |
| | 11 | 126225876 | 0.0774553 | 0.00793111 | 1.60E-22 | rs112771035 | G | C | 0.071369 | 0.000828149 | 95.3732947 |
| | 12 | 125347764 | 0.0304572 | 0.00431469 | 1.70E-12 | rs10692614 | TCA | T | 0.638692 | 0.000432839 | 49.8279713 |
| | 12 | 57843711 | -0.0269424 | 0.00476552 | 1.60E-08 | rs2229357 | A | G | 0.240934 | 0.000277693 | 31.96277059 |
| | 12 | 125265201 | 0.0298785 | 0.00444031 | 1.70E-11 | rs921919 | A | G | 0.669519 | 0.000393327 | 45.27760539 |
| | 15 | 58614376 | 0.030853 | 0.00422211 | 2.70E-13 | rs34433629 | TA | T | 0.570204 | 0.00046384 | 53.3983899 |
| | 15 | 58738423 | -0.0546032 | 0.00449653 | 6.20E-34 | rs12914626 | T | C | 0.701923 | 0.00127985 | 147.4598037 |
| | 15 | 58679668 | 0.0904029 | 0.00428013 | 5.00E-99 | rs7350789 | A | G | 0.353947 | 0.003861931 | 446.111333 |
| | 16 | 57049137 | 0.139883 | 0.0120293 | 3.00E-31 | rs75911530 | A | G | 0.032888 | 0.001173744 | 135.2202129 |
| | 16 | 56702000 | 0.040847 | 0.00650963 | 3.50E-10 | rs904770 | C | T | 0.889382 | 0.000342053 | 39.3731511 |
| | 16 | 56740233 | -0.0316301 | 0.0050556 | 3.90E-10 | rs12923787 | T | C | 0.208879 | 0.000340049 | 39.14246494 |

| | | | | | | | | | | | |
|---|---|---|---|---|---|---|---|---|---|---|---|
| | 16 | 56993886 | -0.210368 | 0.0043597 | 1.00E-200 | rs821840 | G | A | 0.324518 | 0.019832638 | 2328.298169 |
| | 16 | 67985706 | -0.0410289 | 0.00625451 | 5.40E-11 | rs2292318 | T | C | 0.121141 | 0.00373822 | 43.03141391 |
| | 17 | 64210580 | 0.128006 | 0.0118785 | 4.50E-27 | rs1801689 | C | A | 0.030525 | 0.01008169 | 116.1261161 |
| | 17 | 67081278 | 0.12771 | 0.0137424 | 1.50E-20 | rs77542162 | G | A | 0.022664 | 0.00749951 | 86.36088628 |
| | 17 | 66889862 | 0.0283146 | 0.00493222 | 9.40E-09 | rs5821690 | CT | C | 0.25394 | 0.00286316 | 32.95554008 |
| | 17 | 45750596 | -0.0261133 | 0.00409766 | 1.90E-10 | rs3809868 | A | G | 0.518363 | 0.00352803 | 40.61104148 |
| | 18 | 47158234 | -0.0372316 | 0.00530609 | 2.30E-12 | rs9304381 | T | C | 0.818404 | 0.00427684 | 49.23417078 |
| | 18 | 47109955 | -0.133442 | 0.0178534 | 7.80E-14 | rs77960347 | G | A | 0.013235 | 0.00485251 | 55.86446782 |
| | 19 | 45356752 | 0.0375001 | 0.00479341 | 5.10E-15 | rs199956232 | CT | C | 0.266074 | 0.00531593 | 61.20235705 |
| | 19 | 11227480 | 0.0274402 | 0.00416911 | 4.60E-11 | rs2738447 | C | A | 0.593189 | 0.00376321 | 43.31918613 |
| | 19 | 45500925 | -0.0457335 | 0.00437528 | 1.40E-25 | rs2376867 | C | T | 0.324521 | 0.00948591 | 109.2570896 |

| | | | | | | | | | | | |
|---|---|---|---|---|---|---|---|---|---|---|---|
| | 19 | 11197261 | -0.0890661 | 0.00629154 | 1.70E-45 | rs12151108 | A | G | 0.119688 | 0.00173859 | 200.402622 |
| | 19 | 45411941 | 0.104559 | 0.0056591 | 3.20E-76 | rs429358 | C | T | 0.15483 | 0.002957847 | 341.366231 |
| | 20 | 39713869 | 0.0247818 | 0.00407853 | 1.20E-09 | rs2223745 | A | G | 0.47412 | 0.0032074 | 36.91907614 |
| | 20 | 44551855 | 0.107448 | 0.00512813 | 1.80E-97 | rs6073958 | C | T | 0.198741 | 0.0380066 | 439.0066223 |
| | 20 | 43008981 | 0.0733276 | 0.0124442 | 3.80E-09 | rs150224153 | T | C | 0.029235 | 0.00301651 | 34.72109624 |
| | 22 | 21917479 | 0.0306344 | 0.00521149 | 4.10E-09 | rs5998509 | T | C | 0.189548 | 0.00300192 | 34.5531298 |
| exposure | chr.exposure | pos.exposure | beta.exposure | se.exposure | pval.exposure | SNP | effect_allele.exposure | other_allele.exposure | eaf.exposure | R2 | F |
| Free cholesterol to total lipids ratio in medium | 1 | 33227593 | 0.038666 | 0.00702224 | 3.70E-08 | rs61798840 | A | T | 0.086911 | 0.00263382 | 30.3179344 |
| | 1 | 230301811 | 0.03054 | 0.00402679 | 3.30E-14 | rs11122450 | G | T | 0.611471 | 0.00499569 | 57.51916263 |
| | 1 | 55521109 | 0.0300474 | 0.00410804 | 2.60E-13 | rs693668 | A | G | 0.648549 | 0.0046466 | 53.49792308 |
| | 1 | 220970028 | 0.028956 | 0.00430002 | 1.70E-11 | rs2642438 | G | A | 0.70385 | 0.00393874 | 45.3449197 |

| | | | | | | | | | | | |
|---|---|---|---|---|---|---|---|---|---|---|---|
| VLDL \|\| id: ebi-a-GCST90092921 | 1 | 109817590 | -0.0729239 | 0.00472338 | 9.00E-54 | rs12740374 | T | G | 0.221064 | 0.002066941 | 238.3562002 |
| | 1 | 55492083 | -0.0295023 | 0.0044187 | 2.40E-11 | rs59784135 | G | A | 0.696422 | 0.0038721 | 44.57744454 |
| | 1 | 55505647 | -0.289129 | 0.014959 | 3.10E-83 | rs11591147 | T | G | 0.017457 | 0.03235663 | 373.5688335 |
| | 1 | 62999675 | -0.0331481 | 0.00410321 | 6.60E-16 | rs7534572 | G | C | 0.64747 | 0.00566782 | 65.26228028 |
| | 2 | 219699999 | -0.0675984 | 0.0101641 | 2.90E-11 | rs78058190 | A | G | 0.050108 | 0.00384203 | 44.23106813 |
| | 2 | 121347612 | 0.0272718 | 0.00456384 | 2.30E-09 | rs11688682 | C | G | 0.27151 | 0.00310188 | 35.707483 |
| | 2 | 227117778 | -0.0403768 | 0.0041012 | 7.20E-23 | rs2972156 | G | C | 0.645827 | 0.00841529 | 96.92467384 |
| | 2 | 21267461 | 0.0539686 | 0.00415144 | 1.20E-38 | rs934197 | A | G | 0.335688 | 0.01466358 | 168.9963114 |
| | 2 | 27730940 | 0.0650849 | 0.00401264 | 3.60E-59 | rs1260326 | C | T | 0.603982 | 0.02280872 | 263.0828512 |
| | 2 | 165514409 | 0.0320828 | 0.00413315 | 8.30E-15 | rs11394662 | AT | A | 0.384759 | 0.00523296 | 60.25244104 |
| | 2 | 44055510 | -0.0724725 | 0.00823247 | 1.30E-18 | rs114938914 | T | C | 0.061412 | 0.00672956 | 77.49588186 |

| | | | | | | | | | | | |
|---|---|---|---|---|---|---|---|---|---|---|---|
| | 3 | 12394840 | 0.0331728 | 0.00598957 | 3.10E-08 | rs1899951 | T | C | 0.122174 | 0.000266471 | 30.67364757 |
| | 4 | 103895398 | 0.0217497 | 0.00392454 | 3.00E-08 | rs7659650 | A | T | 0.490518 | 0.000266812 | 30.71294589 |
| | 4 | 3364530 | -0.0232812 | 0.0040004 | 5.90E-09 | rs60208975 | G | A | 0.592092 | 0.000294218 | 33.86852932 |
| | 5 | 74472939 | 0.0356694 | 0.0040944 | 3.00E-18 | rs7707394 | A | G | 0.357075 | 0.000659048 | 75.89331506 |
| | 5 | 74618186 | 0.0480586 | 0.00414665 | 4.60E-31 | rs13358429 | A | G | 0.340709 | 0.001165825 | 134.3197648 |
| | 5 | 55857025 | -0.0369785 | 0.00500812 | 1.50E-13 | rs11429307 | GT | G | 0.191517 | 0.000473517 | 54.518209 |
| | 6 | 32603825 | -0.0363057 | 0.00410647 | 9.50E-19 | rs3129774 | T | C | 0.44005 | 0.00067875 | 78.16363947 |
| | 6 | 161125454 | -0.0550341 | 0.00769012 | 8.30E-13 | rs12212146 | C | T | 0.074303 | 0.000444833 | 51.21414846 |
| | 6 | 43758873 | -0.0298957 | 0.00395552 | 4.10E-14 | rs6905288 | A | G | 0.568525 | 0.000496121 | 57.12191208 |
| | 6 | 29937977 | 0.0266486 | 0.00396743 | 1.90E-11 | rs2517671 | G | A | 0.419106 | 0.00039188 | 45.11518084 |
| | 6 | 160922870 | 0.163729 | 0.0145693 | 2.70E-29 | rs117733303 | G | A | 0.0185 | 0.001096201 | 126.2892252 |

| | | | | | | | | | | | |
|---|---|---|---|---|---|---|---|---|---|---|---|
| | 6 | 161010118 | 0.138417 | 0.00728015 | 1.30E-80 | rs10455872 | G | A | 0.078987 | 0.00313329 | 361.4853171 |
| | 6 | 160710851 | 0.0336199 | 0.00540048 | 4.80E-10 | rs3127580 | T | C | 0.156213 | 0.00336647 | 38.75435944 |
| | 7 | 28232457 | 0.0235145 | 0.00413709 | 1.30E-08 | rs849327 | G | A | 0.656011 | 0.00280642 | 32.30531736 |
| | 7 | 71386614 | 0.0249135 | 0.0044419 | 1.70E-08 | rs13229300 | T | C | 0.273994 | 0.00276118 | 31.78439414 |
| | 7 | 73020337 | 0.119033 | 0.00584723 | 4.00E-92 | rs3812316 | G | C | 0.129282 | 0.03588111 | 414.4067844 |
| | 7 | 1029585 | -0.0305228 | 0.00404568 | 4.50E-14 | rs9769088 | C | T | 0.390936 | 0.0049436 | 56.91911479 |
| | 7 | 25991826 | 0.0489604 | 0.00537583 | 8.40E-20 | rs4722551 | C | T | 0.1581 | 0.00720242 | 82.94521053 |
| | 7 | 72082604 | -0.0551575 | 0.00997348 | 3.20E-08 | rs28454618 | A | G | 0.046037 | 0.00265701 | 30.58497695 |
| | 8 | 11095346 | -0.0362558 | 0.00398038 | 8.30E-20 | rs34171564 | A | C | 0.572275 | 0.0072042 | 82.96565888 |
| | 8 | 19733244 | 0.0532227 | 0.00478931 | 1.10E-28 | rs1561748 | C | G | 0.267445 | 0.0107195 | 123.4924367 |
| | 8 | 19819724 | 0.13353 | 0.00651959 | 3.20E-93 | rs328 | G | C | 0.100307 | 0.03631864 | 419.4783784 |

| | | | | | | | | | | | |
|---|---|---|---|---|---|---|---|---|---|---|---|
| | 8 | 126484276 | 0.0306006 | 0.00413036 | 1.30E-13 | rs4336630 | G | T | 0.654138 | 0.000476727 | 54.88788848 |
| | 9 | 136149711 | 0.0681013 | 0.00508232 | 6.10E-41 | rs115478735 | T | A | 0.183304 | 0.001557766 | 179.5474513 |
| | 9 | 139321087 | −0.0239335 | 0.00416872 | 9.40E-09 | rs10747046 | G | A | 0.339246 | 0.000286336 | 32.96092783 |
| | 10 | 95342772 | −0.0394088 | 0.00621774 | 2.30E-10 | rs7091593 | T | A | 0.113925 | 0.000348949 | 40.17110318 |
| | 10 | 113949664 | −0.0262157 | 0.00441245 | 2.80E-09 | rs10787429 | C | T | 0.725575 | 0.000306636 | 35.29846135 |
| | 10 | 65124098 | 0.0407196 | 0.00393088 | 3.80E-25 | rs10822163 | G | C | 0.474416 | 0.00093157 | 107.3049821 |
| | 11 | 65473798 | −0.0332992 | 0.0043256 | 1.40E-14 | rs10750766 | A | C | 0.709783 | 0.000514688 | 59.26077666 |
| | 11 | 5701074 | 0.045984 | 0.00764895 | 1.80E-09 | rs11601507 | A | C | 0.069168 | 0.000313954 | 36.14117777 |
| | 11 | 116648917 | 0.14759 | 0.0057874 | 1.90E-143 | rs964184 | C | G | 0.86726 | 0.005619428 | 650.3382798 |
| | 11 | 61552680 | −0.0916644 | 0.00412699 | 2.70E-109 | rs174537 | T | G | 0.343118 | 0.004268443 | 493.318065 |
| | 11 | 117156893 | 0.124494 | 0.0199713 | 4.60E-10 | rs1047964 | T | G | 0.010073 | 0.000337544 | 38.85765817 |

| | | | | | | | | | | | |
|---|---|---|---|---|---|---|---|---|---|---|---|
| | 11 | 64004643 | -0.0661818 | 0.00938819 | 1.80E-12 | rs12294913 | G | C | 0.049058 | 0.000431637 | 49.69421806 |
| | 11 | 116916060 | -0.187322 | 0.0198763 | 4.30E-21 | rs144018203 | C | G | 0.010653 | 0.000771195 | 88.81758049 |
| | 12 | 112007756 | 0.0264821 | 0.00393472 | 1.70E-11 | rs653178 | T | C | 0.516036 | 0.000393458 | 45.29701974 |
| | 12 | 123350135 | -0.0744958 | 0.0132261 | 1.80E-08 | rs149654682 | A | G | 0.02394 | 0.000275596 | 31.72432753 |
| | 15 | 58602300 | 0.0237857 | 0.00394114 | 1.60E-09 | rs261265 | A | C | 0.454502 | 0.000316405 | 36.42340991 |
| | 15 | 58683366 | -0.0708071 | 0.00403474 | 6.00E-69 | rs1532085 | G | A | 0.613962 | 0.002669036 | 307.9746426 |
| | 15 | 58723479 | 0.0856182 | 0.00475658 | 2.00E-72 | rs1077834 | C | T | 0.218646 | 0.002807464 | 323.9925528 |
| | 15 | 43757184 | -0.119995 | 0.012903 | 1.40E-20 | rs190543502 | C | T | 0.024111 | 0.00075095 | 86.4843166 |
| | 16 | 81534790 | 0.0312484 | 0.0042807 | 2.90E-13 | rs2925979 | C | T | 0.698949 | 0.000462826 | 53.28665238 |
| | 16 | 15148646 | 0.032074 | 0.00431101 | 1.00E-13 | rs11075253 | A | C | 0.295789 | 0.000480764 | 55.35293347 |
| | 16 | 56991363 | -0.048047 | 0.00419969 | 2.60E-30 | rs183130 | T | C | 0.32403 | 0.001136049 | 130.8851985 |

| | | | | | | | | | | | |
|---|---|---|---|---|---|---|---|---|---|---|---|
| | 17 | 67100804 | −0.02635 | 0.00469749 | 2.00E-08 | rs200938405 | AC | A | 0.701626 | 0.00027334 | 31.46458174 |
| | 17 | 41926126 | −0.131093 | 0.011249 | 2.20E-31 | rs72836561 | T | C | 0.031482 | 0.001178722 | 135.8074589 |
| | 17 | 45750596 | −0.0281858 | 0.00393956 | 8.40E-13 | rs3809868 | A | G | 0.518366 | 0.000444595 | 51.1867724 |
| | 17 | 67081278 | 0.169684 | 0.0132115 | 9.30E-38 | rs77542162 | G | A | 0.022666 | 0.001431357 | 164.956708 |
| | 17 | 64210580 | 0.127505 | 0.0114199 | 6.00E-29 | rs1801689 | C | A | 0.030526 | 0.001082062 | 124.6586105 |
| | 19 | 11275842 | 0.030682 | 0.00419138 | 2.50E-13 | rs3185010 | A | G | 0.327174 | 0.000465419 | 53.58530538 |
| | 19 | 45173951 | −0.0995357 | 0.00910697 | 8.30E-28 | rs28807203 | C | A | 0.049207 | 0.001036936 | 119.4544658 |
| | 19 | 45413233 | −0.384366 | 0.00723305 | 1.00E-200 | rs1065853 | T | G | 0.080581 | 0.023950331 | 2823.835841 |
| | 19 | 11197598 | −0.125633 | 0.00603601 | 3.20E-96 | rs73015024 | T | G | 0.119667 | 0.00375032 | 433.2115351 |
| | 19 | 45357939 | 0.0415236 | 0.0039754 | 1.50E-25 | rs2927468 | G | A | 0.523287 | 0.00094713 | 109.0990033 |
| | 19 | 45412955 | 0.0824497 | 0.0119934 | 6.20E-12 | rs1081105 | C | A | 0.027621 | 0.000410495 | 47.25915751 |

| | | | | | | | | | | | |
|---|---|---|---|---|---|---|---|---|---|---|---|
| | 19 | 45254168 | 0.0472965 | 0.00589185 | 1.00E-15 | rs8103315 | A | C | 0.135788 | 0.000559634 | 64.43874836 |
| | 20 | 44590298 | −0.0781313 | 0.00505669 | 7.40E-54 | rs6073972 | G | C | 0.186129 | 0.002070189 | 238.7315914 |
| | 20 | 17844684 | −0.0312681 | 0.00414985 | 4.90E-14 | rs2618566 | T | G | 0.660294 | 0.000493079 | 56.77153524 |
| exposure | chr.exposure | pos.exposure | beta.exposure | se.exposure | pval.exposure | SNP | effect_allele.exposure | other_allele.exposure | eaf.exposure | R2 | F |
| Free cholesterol to total lipids ratio in small VLDL \|\| id:ebi-a-GCST90092973 | 1 | 55492083 | −0.0285897 | 0.00441299 | 9.30E-11 | rs59784135 | G | A | 0.696422 | 0.000364576 | 41.97065647 |
| | 1 | 230301811 | 0.0332726 | 0.00402159 | 1.30E-16 | rs11122450 | G | T | 0.611471 | 0.000594446 | 68.44951034 |
| | 1 | 39618556 | −0.0276903 | 0.00479587 | 7.80E-09 | rs67408364 | G | A | 0.213617 | 0.000289593 | 33.33596026 |
| | 1 | 55521313 | 0.0307924 | 0.00410394 | 6.20E-14 | rs472495 | T | G | 0.649121 | 0.000488951 | 56.29599848 |
| | 1 | 62999675 | −0.0457703 | 0.0040979 | 5.80E-29 | rs7534572 | G | C | 0.64747 | 0.001082846 | 124.7490531 |
| | 1 | 220970028 | 0.03154 | 0.00429446 | 2.10E-13 | rs2642438 | G | A | 0.70385 | 0.000468485 | 53.93848034 |
| | 1 | 27262545 | −0.0706754 | 0.0118695 | 2.60E-09 | rs182050989 | T | C | 0.02814 | 0.000307986 | 35.45391124 |

| | | | | | | | | | | | |
|---|---|---|---|---|---|---|---|---|---|---|---|
| | 1 | 55505647 | -0.271361 | 0.0149397 | 1.00E-73 | rs11591147 | T | G | 0.017457 | 0.002858646 | 329.916139 |
| | 1 | 109817590 | -0.0750547 | 0.00471727 | 5.30E-57 | rs12740374 | T | G | 0.221064 | 0.002194889 | 253.1434941 |
| | 2 | 227105921 | -0.043596 | 0.00409012 | 1.60E-26 | rs2943650 | T | C | 0.644754 | 0.000986246 | 113.6092363 |
| | 2 | 44072576 | -0.0347346 | 0.00419965 | 1.30E-16 | rs4299376 | T | G | 0.676277 | 0.000594064 | 68.40547468 |
| | 2 | 121337280 | 0.0297676 | 0.00536478 | 2.90E-08 | rs9677933 | T | C | 0.158677 | 0.000267461 | 30.78763015 |
| | 2 | 21237544 | 0.0568592 | 0.00483204 | 5.80E-32 | rs673548 | A | G | 0.206075 | 0.001201741 | 138.4627334 |
| | 2 | 21451827 | -0.028583 | 0.00422504 | 1.30E-11 | rs386397 | G | A | 0.688287 | 0.000397534 | 45.76636513 |
| | 2 | 165528624 | 0.0341959 | 0.00401728 | 1.70E-17 | rs1128249 | T | G | 0.392003 | 0.000629221 | 72.45632853 |
| | 2 | 27730940 | 0.0719562 | 0.00400734 | 4.30E-72 | rs1260326 | C | T | 0.603982 | 0.002793841 | 322.4159438 |
| | 2 | 219699999 | -0.0746792 | 0.0101507 | 1.90E-13 | rs78058190 | A | G | 0.050108 | 0.000470106 | 54.12523327 |
| | 4 | 3098321 | 0.0233341 | 0.00414217 | 1.80E-08 | rs7659144 | G | C | 0.344468 | 0.000275676 | 31.73355476 |

| | | | | | | | | | | | |
|---|---|---|---|---|---|---|---|---|---|---|---|
| | 5 | 55861894 | -0.0431486 | 0.00488679 | 1.10E-18 | rs9687846 | A | G | 0.201404 | 0.00676994 | 77.96120043 |
| | 5 | 74605220 | 0.0453834 | 0.00415831 | 9.90E-28 | rs10056811 | A | G | 0.336516 | 0.0103396 | 119.1112507 |
| | 5 | 78531337 | 0.0240696 | 0.00401312 | 2.00E-09 | rs1316753 | C | G | 0.394883 | 0.00312486 | 35.97210944 |
| | 5 | 74472939 | 0.0344934 | 0.00408995 | 3.30E-17 | rs7707394 | A | G | 0.357075 | 0.00617675 | 71.12601291 |
| | 6 | 160922870 | 0.15119 | 0.0145507 | 2.70E-25 | rs117733303 | G | A | 0.0185 | 0.00937268 | 107.9619731 |
| | 6 | 161010118 | 0.116991 | 0.00727086 | 3.00E-58 | rs10455872 | G | A | 0.078987 | 0.02244657 | 258.8962703 |
| | 6 | 26267755 | 0.0273727 | 0.00392536 | 3.10E-12 | rs7773004 | G | A | 0.497283 | 0.00422363 | 48.62602202 |
| | 6 | 32579626 | -0.033709 | 0.0039482 | 1.40E-17 | rs28752510 | A | G | 0.457441 | 0.00633011 | 72.89301303 |
| | 6 | 139831180 | 0.024235 | 0.00405749 | 2.30E-09 | rs679582 | A | G | 0.627691 | 0.00309905 | 35.67496734 |
| | 6 | 161125454 | -0.0620993 | 0.00768031 | 6.20E-16 | rs12212146 | C | T | 0.074303 | 0.00567756 | 65.37449728 |
| | 6 | 43764551 | -0.0284133 | 0.00394815 | 6.20E-13 | rs1358980 | T | C | 0.482622 | 0.00449835 | 51.79031083 |

| | | | | | | | | | | | |
|---|---|---|---|---|---|---|---|---|---|---|---|
| | 7 | 73025975 | 0.127759 | 0.0058387 | 3.90E-106 | rs13234131 | G | A | 0.129443 | 0.004143241 | 478.7878683 |
| | 7 | 130445428 | 0.0218307 | 0.00393067 | 2.80E-08 | rs35363532 | CT | C | 0.49038 | 0.000267965 | 30.84569836 |
| | 7 | 25992323 | 0.0529687 | 0.0053648 | 5.40E-23 | rs10642257 | AAGGCC | A | 0.160158 | 0.00846363 | 97.48190525 |
| | 7 | 73123473 | -0.0563707 | 0.00910569 | 6.00E-10 | rs72555385 | G | A | 0.049304 | 0.00332912 | 38.32424311 |
| | 7 | 128732786 | 0.0237905 | 0.00397276 | 2.10E-09 | rs35163364 | T | C | 0.432067 | 0.00311516 | 35.86038374 |
| | 8 | 126484276 | 0.0482528 | 0.00412196 | 1.20E-31 | rs4336630 | G | T | 0.654138 | 0.0118936 | 137.0345082 |
| | 8 | 8687054 | 0.0358827 | 0.003983 | 2.10E-19 | rs3789849 | C | G | 0.428164 | 0.00704752 | 81.16000552 |
| | 8 | 22608035 | -0.0800007 | 0.0144833 | 3.30E-08 | rs74340693 | G | A | 0.019296 | 0.00265051 | 30.5101943 |
| | 8 | 18244986 | -0.0282898 | 0.00510039 | 2.90E-08 | rs13282367 | G | T | 0.179298 | 0.00267257 | 30.7641865 |
| | 8 | 19733244 | 0.069669 | 0.00477957 | 4.00E-48 | rs1561748 | C | G | 0.267445 | 0.01842863 | 212.4682071 |
| | 8 | 126445055 | -0.0579883 | 0.00912751 | 2.10E-10 | rs72647336 | A | G | 0.056914 | 0.00350604 | 40.36161836 |

| | | | | | | | | | | | |
|---|---|---|---|---|---|---|---|---|---|---|---|
| | 8 | 19819724 | 0.179902 | 0.00650634 | 2.80E-168 | rs328 | G | C | 0.100307 | 0.006599567 | 764.5236537 |
| | 9 | 136149711 | 0.0528562 | 0.00507721 | 2.20E-25 | rs115478735 | T | A | 0.183304 | 0.000940861 | 108.3762384 |
| | 10 | 64934548 | 0.0429728 | 0.00392955 | 7.80E-28 | rs10822145 | T | C | 0.476857 | 0.00103811 | 119.5897942 |
| | 10 | 95342772 | -0.0403999 | 0.00621151 | 7.80E-11 | rs7091593 | T | A | 0.113925 | 0.000367451 | 42.30178125 |
| | 10 | 113934384 | -0.0296363 | 0.00439021 | 1.50E-11 | rs2803619 | C | G | 0.724355 | 0.00039582 | 45.56903458 |
| | 11 | 61552680 | -0.0832173 | 0.00412197 | 1.20E-90 | rs174537 | T | G | 0.343118 | 0.003529188 | 407.5773497 |
| | 11 | 65473798 | -0.0323705 | 0.00432034 | 6.80E-14 | rs10750766 | A | C | 0.709783 | 0.000487578 | 56.13780344 |
| | 11 | 116648917 | 0.177826 | 0.00578036 | 1.00E-200 | rs964184 | C | G | 0.86726 | 0.008156736 | 946.3966569 |
| | 11 | 116916060 | -0.234685 | 0.0198521 | 3.00E-32 | rs144018203 | C | G | 0.010653 | 0.001212895 | 139.7494826 |
| | 11 | 63957835 | -0.0634295 | 0.00834737 | 3.00E-14 | rs11231721 | A | G | 0.058839 | 0.000501485 | 57.73985647 |
| | 11 | 117156893 | 0.174579 | 0.019947 | 2.10E-18 | rs1047964 | T | G | 0.010073 | 0.00066517 | 76.59867901 |

| | | | | | | | | | | | |
|---|---|---|---|---|---|---|---|---|---|---|---|
| | 12 | 20473758 | 0.0219806 | 0.00395595 | 2.80E-08 | rs7134375 | A | C | 0.430211 | 0.00268197 | 30.8723686 |
| | 12 | 112007756 | 0.0266054 | 0.00393052 | 1.30E-11 | rs653178 | T | C | 0.516036 | 0.00397978 | 45.81756629 |
| | 12 | 123201726 | 0.0274084 | 0.0048167 | 1.30E-08 | rs28639492 | C | T | 0.210292 | 0.0028128 | 32.37879068 |
| | 15 | 58680954 | 0.0338902 | 0.00402707 | 3.90E-17 | rs2043085 | C | T | 0.612685 | 0.00615029 | 70.82104953 |
| | 15 | 58723426 | −0.0411538 | 0.00474706 | 4.30E-18 | rs1077835 | G | A | 0.219713 | 0.00652649 | 75.15586813 |
| | 15 | 43757184 | −0.157591 | 0.012894 | 2.40E-34 | rs190543502 | C | T | 0.024111 | 0.01296333 | 149.3756965 |
| | 16 | 15148646 | 0.0284862 | 0.00430732 | 3.80E-11 | rs11075253 | A | C | 0.295789 | 0.00379911 | 43.73682121 |
| | 16 | 81534790 | 0.0366812 | 0.00427703 | 9.80E-18 | rs2925979 | C | T | 0.698949 | 0.0063873 | 73.55207498 |
| | 17 | 45750596 | −0.0236266 | 0.00393607 | 1.90E-09 | rs3809868 | A | G | 0.518366 | 0.00312992 | 36.03041654 |
| | 17 | 64210580 | 0.0905008 | 0.0114098 | 2.20E-15 | rs1801689 | C | A | 0.030526 | 0.00546392 | 62.91312027 |
| | 17 | 67081278 | 0.132156 | 0.0131998 | 1.30E-23 | rs77542162 | G | A | 0.022666 | 0.00870269 | 100.2377988 |

| | | | | | | | | | | | |
|---|---|---|---|---|---|---|---|---|---|---|---|
| | 17 | 76392144 | 0.0232733 | 0.00396419 | 4.30E-09 | rs17561950 | A | G | 0.48446 | 0.000299412 | 34.46668099 |
| | 17 | 41926126 | -0.142483 | 0.0112391 | 7.90E-37 | rs72836561 | T | C | 0.031482 | 0.0013946 | 160.714656 |
| | 19 | 45215081 | -0.0868095 | 0.0139186 | 4.50E-10 | rs111278137 | A | G | 0.021614 | 0.00337901 | 38.89876866 |
| | 19 | 45357939 | 0.0491879 | 0.00396426 | 2.40E-35 | rs2927468 | G | A | 0.523287 | 0.00133599393 | 153.9517941 |
| | 19 | 45413233 | -0.489842 | 0.00721279 | 1.00E-200 | rs1065853 | T | G | 0.080581 | 0.038532968 | 4612.091511 |
| | 19 | 8565131 | 0.0584263 | 0.00702486 | 9.00E-17 | rs60318332 | G | A | 0.090006 | 0.00600721 | 69.17256403 |
| | 19 | 11197261 | -0.11405 | 0.00601907 | 4.60E-80 | rs12151108 | A | G | 0.119677 | 0.0031008 | 359.0246301 |
| | 19 | 45173951 | -0.122916 | 0.00908146 | 9.70E-42 | rs28807203 | C | A | 0.049207 | 0.00158930 5 | 183.188388 |
| | 19 | 45254168 | 0.0603799 | 0.00587535 | 9.00E-25 | rs8103315 | A | C | 0.135788 | 0.00916878 | 105.611435 |
| | 19 | 45412955 | 0.0892732 | 0.0119598 | 8.40E-14 | rs1081105 | C | A | 0.027621 | 0.00483924 | 55.71688432 |
| | 20 | 17844684 | -0.0283391 | 0.0041457 | 8.20E-12 | rs2618566 | T | G | 0.660294 | 0.00405875 | 46.72709873 |

| | | | | | | | | | | | |
|---|---|---|---|---|---|---|---|---|---|---|---|
| | 20 | 44576502 | -0.0592196 | 0.00505011 | 9.30E-32 | rs7679 | C | T | 0.186406 | 0.001193447 | 137.5060212 |
| | 20 | 12958687 | 0.0235469 | 0.00403438 | 5.30E-09 | rs438568 | G | A | 0.609873 | 0.000295922 | 34.06483791 |
| exposure | chr.exposure | pos.exposure | beta.exposure | se.exposure | pval.exposure | SNP | effect_allele.exposure | other_allele.exposure | eaf.exposure | R2 | F |
| Free cholesterol to total lipids ratio in very small VLDL \|\| id:ebi-a-GCST90093033 | 1 | 55492083 | -0.0280051 | 0.00461271 | 1.30E-09 | rs59784135 | G | A | 0.696422 | 0.00320196 | 36.85992142 |
| | 1 | 55505647 | -0.273288 | 0.0156158 | 1.40E-68 | rs11591147 | T | G | 0.017457 | 0.002654306 | 306.270453 |
| | 1 | 62999675 | -0.0530616 | 0.00428337 | 3.00E-35 | rs7534572 | G | C | 0.64747 | 0.001331691 | 153.4553217 |
| | 1 | 109817590 | -0.0861586 | 0.00493076 | 2.30E-68 | rs12740374 | T | G | 0.221064 | 0.002646131 | 305.3247365 |
| | 1 | 25818452 | 0.0237221 | 0.00412221 | 8.70E-09 | rs72660961 | T | C | 0.444274 | 0.000287683 | 33.11604713 |
| | 1 | 55521109 | 0.0340431 | 0.00428841 | 2.00E-15 | rs693668 | A | G | 0.648549 | 0.000547294 | 63.01704775 |
| | 1 | 220970028 | 0.0248794 | 0.00448882 | 3.00E-08 | rs2642438 | G | A | 0.70385 | 0.000266865 | 30.71905674 |
| | 2 | 44073881 | -0.0363455 | 0.00438562 | 1.20E-16 | rs6544713 | C | T | 0.676266 | 0.000596448 | 68.68023151 |

| | | | | | | | | | | | |
|---|---|---|---|---|---|---|---|---|---|---|---|
| | 2 | 227125416 | −0.029405 | 0.00426229 | 5.20E-12 | rs1399626 | A | T | 0.640654 | 0.00413399 | 47.59362995 |
| | 2 | 27730940 | 0.0619232 | 0.00419013 | 2.00E-49 | rs1260326 | C | T | 0.603982 | 0.01894179 | 218.395779 |
| | 2 | 21263900 | 0.0534299 | 0.00432943 | 5.40E-35 | rs1367117 | A | G | 0.335413 | 0.01321678 | 152.29399395 |
| | 3 | 186332182 | 0.0232707 | 0.0042648 | 4.90E-08 | rs2518134 | T | C | 0.632452 | 0.00258644 | 29.77241117 |
| | 5 | 131627840 | 0.0248276 | 0.0044486 | 3.10E-08 | rs270610 | A | G | 0.692896 | 0.0026609 | 30.62974587 |
| | 5 | 74472939 | 0.0317823 | 0.00427085 | 9.90E-14 | rs7707394 | A | G | 0.357075 | 0.00480979 | 55.37764219 |
| | 5 | 74656539 | 0.0460165 | 0.00418479 | 4.00E-28 | rs12916 | C | T | 0.400341 | 0.01049582 | 120.9128006 |
| | 6 | 25715657 | −0.0481596 | 0.00811581 | 3.00E-09 | rs116009877 | A | G | 0.07117 | 0.00305888 | 35.21230479 |
| | 6 | 29903991 | 0.0371322 | 0.00472013 | 3.60E-15 | rs9259970 | A | G | 0.282502 | 0.00537468 | 61.88507112 |
| | 6 | 161125454 | −0.0587796 | 0.00802619 | 2.40E-13 | rs12212146 | C | T | 0.074303 | 0.00465827 | 53.63235068 |
| | 6 | 32565652 | −0.0314236 | 0.00416481 | 4.50E-14 | rs28383169 | G | C | 0.428305 | 0.00494424 | 56.92642896 |

| | | | | | | | | | | | |
|---|---|---|---|---|---|---|---|---|---|---|---|
| | 6 | 43758873 | -0.0225587 | 0.00412839 | 4.60E-08 | rs6905288 | A | G | 0.568525 | 0.00259386 | 29.85789284 |
| | 6 | 161010118 | 0.0886495 | 0.00759831 | 1.90E-31 | rs10455872 | G | A | 0.078987 | 0.01181402 | 136.1165763 |
| | 7 | 73020337 | 0.107524 | 0.00610208 | 1.70E-69 | rs3812316 | G | C | 0.129282 | 0.02690776 | 310.4899037 |
| | 7 | 25991826 | 0.0421048 | 0.00561013 | 6.10E-14 | rs4722551 | C | T | 0.1581 | 0.00489212 | 56.3261179 |
| | 8 | 8687054 | 0.0292405 | 0.00416635 | 2.20E-12 | rs3789849 | C | G | 0.428164 | 0.00427824 | 49.25502459 |
| | 8 | 19733244 | 0.0361699 | 0.0049996 | 4.70E-13 | rs1561748 | C | G | 0.267445 | 0.00454589 | 52.33793093 |
| | 8 | 19907912 | 0.0846004 | 0.00671097 | 1.90E-36 | rs11991231 | C | T | 0.1037 | 0.0137901 | 158.915658 |
| | 8 | 126487691 | 0.040957 | 0.00447325 | 5.40E-20 | rs2980867 | G | T | 0.700748 | 0.00727924 | 83.83056388 |
| | 8 | 18277119 | 0.0249268 | 0.00411071 | 1.30E-09 | rs1466753 | T | C | 0.467416 | 0.00319413 | 36.76984775 |
| | 9 | 136149711 | 0.0515984 | 0.00530164 | 2.20E-22 | rs115478735 | T | A | 0.183304 | 0.00822408 | 94.72060289 |
| | 9 | 139321086 | -0.0244732 | 0.00434851 | 1.80E-08 | rs10747045 | G | T | 0.339211 | 0.00275153 | 31.67327889 |

| | | | | | | | | | | | |
|---|---|---|---|---|---|---|---|---|---|---|---|
| | 10 | 65161903 | 0.0304322 | 0.00410866 | 1.30E-13 | rs10761753 | A | G | 0.473682 | 0.000476488 | 54.86037138 |
| | 11 | 61588305 | -0.112889 | 0.00429338 | 2.30E-152 | rs174564 | G | A | 0.347045 | 0.005971672 | 691.3485345 |
| | 11 | 65405600 | 0.0329663 | 0.00506062 | 7.30E-11 | rs2306363 | T | G | 0.206718 | 0.000368609 | 42.43511762 |
| | 11 | 116714293 | -0.172544 | 0.0216448 | 1.60E-15 | rs141469619 | G | A | 0.010141 | 0.000551881 | 63.54553224 |
| | 11 | 116817978 | 0.049414 | 0.0088035 | 2.00E-08 | rs200671503 | TTA | T | 0.058045 | 0.000273693 | 31.50516344 |
| | 11 | 116648917 | 0.146102 | 0.0060386 | 2.50E-129 | rs964184 | C | G | 0.86726 | 0.005060913 | 585.3724099 |
| | 15 | 43757184 | -0.133543 | 0.0134628 | 3.40E-23 | rs190543502 | C | T | 0.024111 | 0.000854266 | 98.3929362 |
| | 15 | 58680954 | 0.0674712 | 0.00420471 | 6.00E-58 | rs2043085 | C | T | 0.612685 | 0.002232476 | 257.4881373 |
| | 15 | 58723426 | -0.0835269 | 0.00495647 | 1.00E-63 | rs1077835 | G | A | 0.219713 | 0.002461671 | 283.9881479 |
| | 15 | 101905966 | -0.0265436 | 0.00435329 | 1.10E-09 | rs8029570 | G | T | 0.333143 | 0.000322951 | 37.17721921 |
| | 16 | 56991363 | -0.0497532 | 0.00438081 | 6.80E-30 | rs183130 | T | C | 0.32403 | 0.001119539 | 128.9809746 |

| | | | | | | | | | | | |
|---|---|---|---|---|---|---|---|---|---|---|---|
| | 16 | 15104807 | 0.0392403 | 0.0051279 | 2.00E-14 | rs11645958 | T | C | 0.276072 | 0.000508578 | 58.55688901 |
| | 17 | 26694861 | 0.0266237 | 0.00409531 | 8.00E-11 | rs704 | A | G | 0.476026 | 0.00036711 | 42.26255126 |
| | 17 | 45750596 | -0.0266596 | 0.00411045 | 8.80E-11 | rs3809868 | A | G | 0.518366 | 0.000365395 | 42.06500751 |
| | 17 | 66425997 | 0.0273659 | 0.00492587 | 2.80E-08 | rs3785611 | T | C | 0.771343 | 0.00026812 | 30.86356089 |
| | 17 | 67081278 | 0.128975 | 0.0137846 | 8.20E-21 | rs77542162 | G | A | 0.022666 | 0.000760125 | 87.54174289 |
| | 19 | 11197598 | -0.122183 | 0.00630356 | 1.10E-83 | rs73015024 | T | G | 0.119667 | 0.00325407 | 375.7009031 |
| | 19 | 11228745 | 0.0311833 | 0.00418242 | 8.90E-14 | rs2569550 | C | T | 0.593326 | 0.000482805 | 55.58803378 |
| | 19 | 45173951 | -0.0912496 | 0.00951064 | 8.40E-22 | rs28807203 | C | A | 0.049207 | 0.000799259 | 92.05235534 |
| | 19 | 45413233 | -0.344014 | 0.00755366 | 1.00E-200 | rs1065853 | T | G | 0.080581 | 0.017704038 | 2074.100618 |
| | 19 | 45441608 | -0.0398019 | 0.00443231 | 2.70E-19 | rs8106814 | C | T | 0.504189 | 0.000700222 | 80.63801068 |
| | 20 | 17844684 | -0.0253474 | 0.00432858 | 4.70E-09 | rs2618566 | T | G | 0.660294 | 0.000297878 | 34.29007161 |

| | | | | | | | | | | | |
|---|---|---|---|---|---|---|---|---|---|---|---|
| | 20 | 44554015 | -0.0531644 | 0.00527604 | 7.00E-24 | rs6065906 | C | T | 0.185857 | 0.00881526 | 101.5355504 |
| | 20 | 12962718 | 0.0254171 | 0.00420653 | 1.50E-09 | rs364585 | G | A | 0.609491 | 0.00317146 | 36.5087082 |
| exposure | chr.exposure | pos.exposure | beta.exposure | se.exposure | pval.exposure | SNP | effect_allele.exposure | other_allele.exposure | eaf.exposure | R2 | F |
| Free cholesterol to total lipids ratio in IDL \|\| id:ebi-a-GCST90092836 | 1 | 63034222 | 0.0766004 | 0.00431132 | 1.30E-70 | rs10789114 | C | T | 0.349833 | 0.02735549 | 315.6705084 |
| | 1 | 55488831 | -0.026387 | 0.0046898 | 1.80E-08 | rs9436961 | A | T | 0.742768 | 0.00275007 | 31.65656889 |
| | 1 | 109817192 | -0.0522365 | 0.0049283 | 3.00E-26 | rs7528419 | G | A | 0.222163 | 0.00975265 | 112.3430782 |
| | 2 | 21136327 | -0.0242795 | 0.00418125 | 6.40E-09 | rs12710748 | C | T | 0.411398 | 0.00292909 | 33.71783335 |
| | 2 | 227036911 | 0.033749 | 0.00439863 | 1.70E-14 | rs62188784 | T | C | 0.318681 | 0.00511278 | 58.86801698 |
| | 2 | 165514409 | 0.0249161 | 0.00431636 | 7.80E-09 | rs11394662 | AT | A | 0.384759 | 0.00289462 | 33.32094152 |
| | 2 | 21293914 | -0.0250671 | 0.00411027 | 1.10E-09 | rs571468 | C | A | 0.46583 | 0.00323087 | 37.19289097 |
| | 2 | 27730940 | 0.054334 | 0.00419052 | 1.90E-38 | rs1260326 | C | T | 0.603982 | 0.01458701 | 168.1124873 |

| | | | | | | | | | | | |
|---|---|---|---|---|---|---|---|---|---|---|---|
| | 3 | 186332182 | 0.0379667 | 0.00426473 | 5.50E-19 | rs2518134 | T | C | 0.632452 | 0.000688202 | 79.25288331 |
| | 4 | 88181491 | −0.0245294 | 0.00421679 | 6.00E-09 | rs10020067 | A | C | 0.381441 | 0.000293951 | 33.83781848 |
| | 5 | 74605220 | 0.0257693 | 0.00434268 | 3.00E-09 | rs10056811 | A | G | 0.336516 | 0.000305879 | 35.21129827 |
| | 5 | 55861894 | −0.0316376 | 0.00510347 | 5.70E-10 | rs9687846 | A | G | 0.201404 | 0.000333829 | 38.42982279 |
| | 6 | 161125454 | −0.04768 | 0.00802815 | 2.90E-09 | rs12212146 | C | T | 0.074303 | 0.000306409 | 35.27231701 |
| | 6 | 25715657 | −0.0526389 | 0.0081178 | 8.90E-11 | rs116009877 | A | G | 0.07117 | 0.000365234 | 42.04645331 |
| | 6 | 32605508 | −0.0247547 | 0.00419063 | 3.50E-09 | rs2213287 | T | G | 0.397317 | 0.000303122 | 34.89387107 |
| | 6 | 43758873 | −0.0275956 | 0.0041294 | 2.30E-11 | rs6905288 | A | G | 0.568525 | 0.000387909 | 44.6578929 |
| | 6 | 161010118 | 0.0665856 | 0.00760017 | 1.90E-18 | rs10455872 | G | A | 0.078987 | 0.000666526 | 76.75496431 |
| | 6 | 139831180 | 0.0241704 | 0.00424126 | 1.20E-08 | rs679582 | A | G | 0.627691 | 0.000282129 | 32.47658341 |
| | 7 | 17978828 | 0.0227371 | 0.00412855 | 3.60E-08 | rs56287412 | C | T | 0.508001 | 0.000263483 | 30.329657 |

| | | | | | | | | | | | |
|---|---|---|---|---|---|---|---|---|---|---|---|
| | 7 | 25983400 | 0.0243107 | 0.0042811 | 1.40E-08 | rs56001710 | T | A | 0.580583 | 0.00280127 | 32.24605836 |
| | 7 | 73020337 | 0.0960658 | 0.00610168 | 7.50E-56 | rs3812316 | G | C | 0.129282 | 0.00214929 9 | 247.8741311 |
| | 8 | 19733244 | 0.0475289 | 0.00499871 | 1.90E-21 | rs1561748 | C | G | 0.267445 | 0.00784967 | 90.40492598 |
| | 8 | 126507389 | 0.0466259 | 0.00446778 | 1.70E-25 | rs2954038 | A | C | 0.698676 | 0.00945481 | 108.9088943 |
| | 8 | 19819724 | 0.130034 | 0.00680464 | 2.10E-81 | rs328 | G | C | 0.100307 | 0.03163153 | 365.1707548 |
| | 8 | 126445055 | -0.0556023 | 0.00954599 | 5.70E-09 | rs72647336 | A | G | 0.056914 | 0.00294719 | 33.9262623 |
| | 8 | 10649767 | -0.0245296 | 0.00423516 | 7.00E-09 | rs752446 | A | G | 0.375244 | 0.00291412 | 33.54545915 |
| | 8 | 18239697 | -0.0353641 | 0.00533537 | 3.40E-11 | rs13266992 | C | T | 0.180568 | 0.00381613 | 43.93277006 |
| | 9 | 136149711 | 0.0415936 | 0.00530145 | 4.30E-15 | rs115478735 | T | A | 0.183304 | 0.00534594 | 61.55397938 |
| | 9 | 139321087 | -0.0266722 | 0.00434846 | 8.60E-10 | rs10747046 | G | A | 0.339246 | 0.00326812 | 37.62176544 |
| | 10 | 64878118 | 0.0261637 | 0.00414091 | 2.60E-10 | rs6479877 | G | C | 0.443609 | 0.00346776 | 39.92079587 |

| | | | | | | | | | | | |
|---|---|---|---|---|---|---|---|---|---|---|---|
| | 11 | 61588305 | −0.1042 | 0.00429363 | 4.20E-130 | rs174564 | G | A | 0.347045 | 0.005091689 | 588.9503128 |
| | 11 | 116916060 | −0.225394 | 0.0207402 | 1.60E-27 | rs144018203 | C | G | 0.010653 | 0.001025194 | 118.1003743 |
| | 11 | 116648917 | 0.18513 | 0.00603895 | 1.00E-200 | rs964184 | C | G | 0.86726 | 0.008100115 | 939.7734837 |
| | 11 | 65473798 | −0.0326197 | 0.00451361 | 4.90E-13 | rs10750766 | A | C | 0.709783 | 0.000453636 | 52.22811064 |
| | 14 | 106135632 | −0.0283573 | 0.00473531 | 2.10E-09 | rs8015375 | G | A | 0.491435 | 0.000311523 | 35.86124613 |
| | 15 | 101892507 | 0.0231042 | 0.00422504 | 4.50E-08 | rs11247282 | C | T | 0.387749 | 0.000259777 | 29.90285626 |
| | 15 | 58723426 | −0.07834 | 0.00495659 | 2.90E-56 | rs1077835 | G | A | 0.219713 | 0.002165968 | 249.8006672 |
| | 15 | 102068658 | −0.0312655 | 0.00472213 | 3.60E-11 | rs1037117 | A | G | 0.254935 | 0.000380787 | 43.83765667 |
| | 15 | 58680954 | 0.0598749 | 0.00420482 | 5.20E-46 | rs2043085 | C | T | 0.612685 | 0.001758826 | 202.7623127 |
| | 15 | 43757184 | −0.160649 | 0.0134631 | 8.00E-33 | rs190543502 | C | T | 0.024111 | 0.001235724 | 142.3830731 |
| | 16 | 15129940 | 0.0242292 | 0.00417185 | 6.30E-09 | rs3198697 | T | C | 0.406727 | 0.000293012 | 33.72975855 |

| | | | | | | | | | | | |
|---|---|---|---|---|---|---|---|---|---|---|---|
| | 17 | 26694861 | 0.0237635 | 0.0040949 | 6.50E-09 | rs704 | A | G | 0.476026 | 0.00029255 | 33.67647785 |
| | 17 | 66425995 | 0.0271311 | 0.00492611 | 3.60E-08 | rs3785610 | G | T | 0.771374 | 0.00263515 | 30.33325707 |
| | 17 | 41840849 | -0.0803877 | 0.012259 | 5.50E-11 | rs77697917 | T | C | 0.028723 | 0.00373508 | 42.9993181 |
| | 18 | 47207205 | -0.0272854 | 0.00425187 | 1.40E-10 | rs55987145 | AC | A | 0.38656 | 0.00357716 | 41.18064522 |
| | 19 | 8429323 | 0.154011 | 0.0146982 | 1.10E-25 | rs116843064 | A | G | 0.019851 | 0.00953133 | 109.7912245 |
| | 19 | 19456917 | 0.0448465 | 0.00731933 | 8.90E-10 | rs58489806 | T | C | 0.086437 | 0.00326111 | 37.54109233 |
| | 19 | 45413233 | -0.180501 | 0.00755936 | 5.20E-126 | rs1065853 | T | G | 0.080581 | 0.0492987 | 570.1401431 |
| | 19 | 11206969 | -0.0414248 | 0.00619554 | 2.30E-11 | rs17242395 | A | G | 0.126447 | 0.00388317 | 44.70488299 |
| | 19 | 45442528 | -0.0317146 | 0.00436718 | 3.80E-13 | rs12977604 | G | C | 0.499959 | 0.00458047 | 52.73618923 |
| | 19 | 54800500 | 0.0290574 | 0.00492306 | 3.60E-09 | rs367070 | G | A | 0.225184 | 0.00302625 | 34.83659492 |
| | 20 | 44559748 | -0.0489288 | 0.00526955 | 1.60E-20 | rs139953093 | T | G | 0.186317 | 0.00748599 | 86.21335029 |

| exposure | chr.exposure | pos.exposure | beta.exposure | se.exposure | pval.exposure | SNP | effect_allele.exposure | other_allele.exposure | eaf.exposure | R2 | F |
|---|---|---|---|---|---|---|---|---|---|---|---|
| Free cholesterol to total lipids ratio in large LDL \|\| id:ebi-a-GCST90092861 | 1 | 39618556 | -0.0365775 | 0.00478981 | 2.20E-14 | rs67408364 | G | A | 0.213617 | 0.000506482 | 58.31548845 |
| | 1 | 55674000 | -0.127069 | 0.0184133 | 5.20E-12 | rs12750160 | T | C | 0.012266 | 0.000413646 | 47.62208641 |
| | 1 | 220970028 | 0.0351201 | 0.00428903 | 2.60E-16 | rs2642438 | G | A | 0.70385 | 0.000582281 | 67.04799143 |
| | 1 | 27262545 | -0.08494 | 0.0118544 | 7.80E-13 | rs182050989 | T | C | 0.02814 | 0.000445927 | 51.3402301 |
| | 1 | 109817590 | -0.0344208 | 0.0047113 | 2.80E-13 | rs12740374 | T | G | 0.221064 | 0.000463609 | 53.37683923 |
| | 1 | 230301811 | 0.0449235 | 0.0040165 | 4.80E-29 | rs11122450 | G | T | 0.611471 | 0.00108556 | 125.0961891 |
| | 1 | 63047987 | 0.0550569 | 0.00410315 | 4.70E-41 | rs11207994 | T | C | 0.350943 | 0.00156277 | 180.0450451 |
| | 2 | 27730940 | 0.0939994 | 0.00399989 | 4.00E-122 | rs1260326 | C | T | 0.603982 | 0.00477635 | 552.2637267 |
| | 2 | 21226560 | -0.106819 | 0.00482423 | 1.20E-108 | rs2678379 | G | A | 0.793822 | 0.00424216 | 490.2678991 |
| | 2 | 21372547 | -0.0502604 | 0.00814755 | 6.90E-10 | rs11689541 | C | T | 0.062039 | 0.000330557 | 38.0531195 |

| | | | | | | | | | | | |
|---|---|---|---|---|---|---|---|---|---|---|---|
| | 2 | 165528624 | 0.0435363 | 0.0040098 | 1.80E-27 | rs1128249 | T | G | 0.392003 | 0.001023306 | 117.882697 |
| | 2 | 219699999 | -0.0870386 | 0.0101318 | 8.60E-18 | rs78058190 | A | G | 0.050108 | 0.000640862 | 73.79773446 |
| | 2 | 227105921 | -0.0494689 | 0.00408251 | 8.60E-34 | rs2943650 | T | C | 0.644754 | 0.001274233 | 146.8258233 |
| | 3 | 135956305 | -0.0351378 | 0.00463715 | 3.50E-14 | rs684773 | C | A | 0.767075 | 0.000498681 | 57.41684128 |
| | 3 | 52359678 | 0.030787 | 0.00508456 | 1.40E-09 | rs6796333 | C | T | 0.180898 | 0.00031848 | 36.66236211 |
| | 3 | 150097635 | -0.0461047 | 0.00777365 | 3.00E-09 | rs9844972 | C | G | 0.070763 | 0.000305563 | 35.17490126 |
| | 3 | 172253678 | -0.033885 | 0.0056241 | 1.70E-09 | rs6764884 | T | G | 0.143359 | 0.000315329 | 36.29956027 |
| | 4 | 3443931 | 0.0273666 | 0.00405591 | 1.50E-11 | rs13108218 | G | A | 0.615355 | 0.000395445 | 45.5257933 |
| | 4 | 87985166 | -0.0269916 | 0.00401584 | 1.80E-11 | rs3775228 | T | C | 0.398282 | 0.000392398 | 45.17486969 |
| | 4 | 77414144 | -0.0233471 | 0.0039354 | 3.00E-09 | rs10008637 | C | T | 0.461943 | 0.000305737 | 35.19496817 |
| | 4 | 103188709 | -0.0520563 | 0.00746961 | 3.20E-12 | rs13107325 | T | C | 0.074282 | 0.000421852 | 48.56721396 |

| | | | | | | | | | | | |
|---|---|---|---|---|---|---|---|---|---|---|---|
| | 5 | 156443066 | -0.0227333 | 0.00408998 | 2.70E-08 | rs4704834 | G | A | 0.644028 | 0.000268385 | 30.89406458 |
| | 5 | 78433363 | 0.0298741 | 0.00412747 | 4.60E-13 | rs200450501 | TAC | T | 0.347633 | 0.00455006 | 52.38588226 |
| | 5 | 55857025 | -0.0579755 | 0.00499676 | 4.00E-31 | rs11429307 | GT | G | 0.191517 | 0.01168414 | 134.6184164 |
| | 5 | 74472939 | 0.0246455 | 0.00408511 | 1.60E-09 | rs7707394 | A | G | 0.357075 | 0.00316172 | 36.39654911 |
| | 5 | 75015242 | 0.0304151 | 0.00408488 | 9.60E-14 | rs2112347 | G | T | 0.361738 | 0.00481508 | 55.43861142 |
| | 6 | 32583057 | -0.0397462 | 0.00399327 | 2.40E-23 | rs3129755 | G | A | 0.565325 | 0.00860108 | 99.06638795 |
| | 6 | 43758873 | -0.034703 | 0.00394409 | 1.40E-18 | rs6905288 | A | G | 0.568525 | 0.00672266 | 77.41638 |
| | 6 | 160922870 | 0.174215 | 0.0145272 | 3.90E-33 | rs117733303 | G | A | 0.0185 | 0.01248123 | 143.8135185 |
| | 6 | 26340872 | -0.0246474 | 0.00402128 | 8.80E-10 | rs55964643 | A | G | 0.503231 | 0.00326336 | 37.56695988 |
| | 6 | 161125454 | -0.0610643 | 0.0076679 | 1.70E-15 | rs12212146 | C | T | 0.074303 | 0.00550776 | 63.41826224 |
| | 6 | 31303922 | -0.0293601 | 0.00406249 | 4.90E-13 | rs28752924 | C | T | 0.445537 | 0.00453655 | 52.23034606 |

| | | | | | | | | | | | |
|---|---|---|---|---|---|---|---|---|---|---|---|
| | 6 | 32529147 | -0.0616489 | 0.00958202 | 1.20E-10 | rs71536559 | T | G | 0.052986 | 0.00359561 | 41.39319698 |
| | 6 | 127454893 | -0.0618334 | 0.0091573 | 1.50E-11 | rs72959041 | A | G | 0.049614 | 0.00396034 | 45.5935935 |
| | 6 | 139834012 | 0.0335016 | 0.00405103 | 1.30E-16 | rs632057 | G | T | 0.627778 | 0.00593929 | 68.39000438 |
| | 6 | 161010118 | 0.110249 | 0.00725912 | 4.30E-52 | rs10455872 | G | A | 0.078987 | 0.02000342 | 230.6608132 |
| | 7 | 73020337 | 0.143174 | 0.00583398 | 5.30E-133 | rs3812316 | G | C | 0.129282 | 0.00520623 9 | 602.2695376 |
| | 7 | 73123473 | -0.074315 | 0.00909459 | 3.00E-16 | rs72555385 | G | A | 0.049304 | 0.00579866 | 66.76966026 |
| | 7 | 128732786 | 0.0238849 | 0.00396792 | 1.70E-09 | rs35163364 | T | C | 0.432067 | 0.00314758 | 36.23376741 |
| | 7 | 17978828 | 0.0230627 | 0.00394742 | 5.10E-09 | rs56287412 | C | T | 0.508001 | 0.00296522 | 34.13391302 |
| | 7 | 25992323 | 0.0460183 | 0.00535826 | 8.80E-18 | rs10642257 | AAGGCC | A | 0.160158 | 0.00640513 | 73.75747285 |
| | 7 | 130447130 | -0.028344 | 0.00430656 | 4.70E-11 | rs59326756 | G | A | 0.293748 | 0.00376262 | 43.31653674 |
| | 8 | 19733244 | 0.0752029 | 0.00477137 | 5.70E-56 | rs1561748 | C | G | 0.267445 | 0.02153967 | 248.4136396 |

| | | | | | | | | | | | |
|---|---|---|---|---|---|---|---|---|---|---|---|
| | 8 | 19980272 | 0.0487574 | 0.00769855 | 2.40E-10 | rs71205961 | CTTTTTT | C | 0.911027 | 0.00348421 | 40.11027692 |
| | 8 | 126500031 | 0.0625079 | 0.00401015 | 8.90E-55 | rs28601761 | G | C | 0.420159 | 0.002106809 | 242.9634995 |
| | 8 | 126629878 | −0.0314171 | 0.00481448 | 6.80E-11 | rs7009450 | T | G | 0.210014 | 0.00369884 | 42.58198153 |
| | 8 | 11094566 | −0.032298 | 0.00396929 | 4.10E-16 | rs7007394 | A | G | 0.573492 | 0.00575001 | 66.20915615 |
| | 8 | 18248661 | −0.0287127 | 0.00506044 | 1.40E-08 | rs4646246 | G | A | 0.182596 | 0.00279668 | 32.19318603 |
| | 8 | 19819724 | 0.195349 | 0.00649517 | 1.00E-198 | rs328 | G | C | 0.100307 | 0.00779890 5 | 904.552529 |
| | 9 | 136149711 | 0.0307315 | 0.0050714 | 1.40E-09 | rs115478735 | T | A | 0.183304 | 0.00318982 | 36.72013228 |
| | 9 | 139321087 | −0.0231552 | 0.00415976 | 2.60E-08 | rs10747046 | G | A | 0.339246 | 0.00269176 | 30.98510531 |
| | 9 | 92119579 | −0.0259672 | 0.00450009 | 7.90E-09 | rs13287131 | C | T | 0.258077 | 0.00289251 | 33.29663145 |
| | 10 | 94839642 | 0.026122 | 0.00393741 | 3.30E-11 | rs2068888 | A | G | 0.450101 | 0.00382312 | 44.01330919 |
| | 10 | 95309022 | 0.0384442 | 0.00594171 | 9.80E-11 | rs10786114 | T | C | 0.875672 | 0.00363641 | 41.86308273 |

| | | | | | | | | | | | |
|---|---|---|---|---|---|---|---|---|---|---|---|
| | 10 | 113934384 | -0.039834 | 0.00438395 | 1.00E-19 | rs2803619 | C | G | 0.724355 | 0.00716899 | 82.55989005 |
| | 10 | 65161903 | 0.0539741 | 0.00392837 | 5.90E-43 | rs10761753 | A | G | 0.473682 | 0.01637672 | 188.7724009 |
| | 11 | 27631005 | -0.0258945 | 0.00468969 | 3.40E-08 | rs141078087 | AT | A | 0.227747 | 0.00264853 | 30.48731936 |
| | 11 | 47275064 | 0.0244479 | 0.00441812 | 3.10E-08 | rs10838681 | A | G | 0.266784 | 0.00266002 | 30.6196737 |
| | 11 | 63882495 | -0.0633625 | 0.00832471 | 2.70E-14 | rs35927325 | T | C | 0.058816 | 0.00503153 | 57.93204164 |
| | 11 | 65405600 | 0.0302135 | 0.00483767 | 4.20E-10 | rs2306363 | T | G | 0.206718 | 0.00338825 | 39.00515863 |
| | 11 | 116389017 | -0.0235986 | 0.00427529 | 3.40E-08 | rs883863 | A | G | 0.308012 | 0.00264679 | 30.46728621 |
| | 11 | 116714293 | -0.290076 | 0.0206912 | 1.20E-44 | rs141469619 | G | A | 0.010141 | 0.01704919 | 196.5371669 |
| | 11 | 116648917 | 0.204 | 0.00577256 | 1.00E-200 | rs964184 | C | G | 0.86726 | 0.01073565 | 1248.866097 |
| | 11 | 61579463 | -0.0835689 | 0.00411941 | 1.70E-91 | rs174554 | G | A | 0.341712 | 0.03563376 | 411.5397466 |
| | 11 | 117156893 | 0.225716 | 0.0199201 | 9.20E-30 | rs1047964 | T | G | 0.010073 | 0.01114423 | 128.390862 |

| | | | | | | | | | | | |
|---|---|---|---|---|---|---|---|---|---|---|---|
| | 12 | 124506631 | 0.0267773 | 0.00401561 | 2.60E-11 | rs10773049 | C | T | 0.393095 | 0.00386238 | 44.46547814 |
| | 12 | 57692470 | 0.0290881 | 0.00458361 | 2.20E-10 | rs12313762 | T | C | 0.238672 | 0.00349829 | 40.2724309 |
| | 15 | 58680184 | -0.0469903 | 0.00410699 | 2.60E-30 | rs7177289 | T | C | 0.354013 | 0.01136235 | 130.9066162 |
| | 15 | 43726625 | -0.203739 | 0.0124022 | 1.20E-60 | rs150844304 | C | A | 0.025671 | 0.02339517 | 269.8629923 |
| | 15 | 58738423 | 0.0238023 | 0.00431512 | 3.50E-08 | rs12914626 | T | C | 0.701916 | 0.0026432 | 30.42597797 |
| | 16 | 81534790 | 0.0390907 | 0.0042728 | 5.80E-20 | rs2925979 | C | T | 0.698949 | 0.00726773 | 83.69783366 |
| | 16 | 15172118 | 0.0286741 | 0.00430756 | 2.80E-11 | rs11644601 | C | T | 0.296695 | 0.00384895 | 44.3107777 |
| | 16 | 57005479 | 0.0216107 | 0.00396222 | 4.90E-08 | rs1532624 | A | C | 0.439995 | 0.00258429 | 29.74766963 |
| | 16 | 69733460 | -0.0225979 | 0.00399802 | 1.60E-08 | rs7359336 | A | G | 0.587596 | 0.00277535 | 31.94763343 |
| | 17 | 76391454 | 0.0236378 | 0.0040166 | 4.00E-09 | rs4969179 | G | T | 0.603899 | 0.00300856 | 34.63294251 |
| | 17 | 41926126 | -0.12824 | 0.0112271 | 3.20E-30 | rs72836561 | T | C | 0.031482 | 0.01132433 | 130.4680854 |

| | | | | | | | | | | | |
|---|---|---|---|---|---|---|---|---|---|---|---|
| | 17 | 64210580 | 0.0711442 | 0.0113976 | 4.30E-10 | rs1801689 | C | A | 0.030526 | 0.00338453 | 38.96230396 |
| | 18 | 19898816 | 0.0215869 | 0.00393375 | 4.10E-08 | rs11662649 | C | T | 0.529393 | 0.00261605 | 30.11337968 |
| | 18 | 31246594 | -0.0216674 | 0.00394903 | 4.10E-08 | rs4799707 | A | G | 0.553054 | 0.00261524 | 30.10406813 |
| | 19 | 45337918 | 0.100005 | 0.010727 | 1.10E-20 | rs147711004 | A | G | 0.036038 | 0.0075466 | 86.91191506 |
| | 19 | 45173951 | -0.0976375 | 0.00908197 | 5.90E-27 | rs28807203 | C | A | 0.049207 | 0.01003298 | 115.5754593 |
| | 19 | 11209757 | -0.051336 | 0.00597639 | 8.70E-18 | rs141820146 | CCAAA | C | 0.124925 | 0.00640738 | 73.78339663 |
| | 19 | 45413233 | -0.384964 | 0.00721319 | 1.00E-200 | rs1065853 | T | G | 0.080581 | 0.02415238 9 | 2848.248944 |
| | 19 | 8429323 | 0.256634 | 0.0140251 | 8.60E-75 | rs116843064 | A | G | 0.019851 | 0.02900997 | 334.818083 |
| | 19 | 8442202 | 0.0525859 | 0.00848798 | 5.80E-10 | rs62117489 | A | C | 0.056468 | 0.00333409 | 38.38153854 |
| | 20 | 44576502 | -0.0750549 | 0.00504478 | 4.60E-50 | rs7679 | C | T | 0.186406 | 0.01919693 | 221.3431521 |
| | 20 | 62711459 | 0.0215681 | 0.00392686 | 4.00E-08 | rs8126001 | T | C | 0.488461 | 0.00262067 | 30.16653251 |

| | | | | | | | | | | | |
|---|---|---|---|---|---|---|---|---|---|---|---|
| | 20 | 12959398 | 0.0250076 | 0.00402647 | 5.30E-10 | rs4814176 | C | T | 0.608192 | 0.00335075 | 38.57336665 |
| | 22 | 38600542 | -0.0220506 | 0.00398905 | 3.20E-08 | rs2267373 | T | C | 0.580695 | 0.00265448 | 30.55584616 |
| exposure | chr.exposure | pos.exposure | beta.exposure | se.exposure | pval.exposure | SNP | effect_allele.exposure | other_allele.exposure | eaf.exposure | R2 | F |
| Free cholesterol to total lipids ratio in medium LDL \|\| id:ebi-a-GCST90092909 | 1 | 39618556 | -0.0302138 | 0.00481101 | 3.40E-10 | rs67408364 | G | A | 0.213617 | 0.00342596 | 39.43943033 |
| | 1 | 27284913 | -0.0903551 | 0.0130595 | 4.60E-12 | rs79598313 | T | C | 0.023201 | 0.00415781 | 47.86793523 |
| | 1 | 63060037 | 0.0754242 | 0.00411477 | 4.80E-75 | rs79439217 | TAAGAA | T | 0.352737 | 0.02911098 | 335.9872117 |
| | 1 | 109817590 | -0.0314884 | 0.00473215 | 2.80E-11 | rs12740374 | T | G | 0.221064 | 0.003846 | 44.2768417 |
| | 1 | 219618035 | -0.0301897 | 0.00533319 | 1.50E-08 | rs12747505 | G | A | 0.161893 | 0.00278365 | 32.04320402 |
| | 1 | 230302838 | 0.03714 | 0.00408055 | 8.90E-20 | rs59484402 | CT | C | 0.611688 | 0.00719327 | 82.83976223 |
| | 2 | 227104111 | -0.0457661 | 0.00411413 | 9.60E-29 | rs2972140 | C | T | 0.650637 | 0.01074132 | 123.744026 |
| | 2 | 165528876 | 0.0489245 | 0.00402599 | 5.60E-34 | rs13389219 | T | C | 0.392532 | 0.01281572 | 147.6725793 |

| | | | | | | | | | | | |
|---|---|---|---|---|---|---|---|---|---|---|---|
| | 2 | 219699999 | -0.0805543 | 0.0101757 | 2.40E-15 | rs78058190 | A | G | 0.050108 | 0.00544258 | 62.66734843 |
| | 2 | 21226560 | -0.102309 | 0.00484513 | 5.70E-99 | rs2678379 | G | A | 0.793822 | 0.003859491 | 445.8710384 |
| | 2 | 27730940 | 0.0913448 | 0.00401722 | 1.90E-114 | rs1260326 | C | T | 0.603982 | 0.004472622 | 517.0218276 |
| | 3 | 52359678 | 0.0280909 | 0.00510619 | 3.80E-08 | rs6796333 | C | T | 0.180898 | 0.00262915 | 30.2642432 |
| | 3 | 135956305 | -0.0412333 | 0.00465688 | 8.40E-19 | rs684773 | C | A | 0.767075 | 0.00680764 | 78.39569712 |
| | 4 | 100508556 | 0.0433556 | 0.00709668 | 1.00E-09 | rs2035816 | G | A | 0.084301 | 0.00324214 | 37.32264454 |
| | 4 | 3443931 | 0.0269589 | 0.00407311 | 3.60E-11 | rs13108218 | G | A | 0.615355 | 0.00380522 | 43.80710059 |
| | 4 | 87976359 | -0.0333384 | 0.00403766 | 1.50E-16 | rs1471251 | T | A | 0.397121 | 0.0059206 | 68.17457945 |
| | 5 | 57610069 | 0.0229089 | 0.00404322 | 1.50E-08 | rs37538 | C | G | 0.602788 | 0.00278885 | 32.10304146 |
| | 5 | 78531337 | 0.0243655 | 0.00402433 | 1.40E-09 | rs1316753 | C | G | 0.394883 | 0.00318433 | 36.65691696 |
| | 5 | 156443066 | -0.025185 | 0.00410627 | 8.60E-10 | rs4704834 | G | A | 0.644028 | 0.00326768 | 37.61675759 |

| | | | | | | | | | | | |
|---|---|---|---|---|---|---|---|---|---|---|---|
| | 5 | 55857025 | −0.0537422 | 0.00501666 | 8.90E-27 | rs11429307 | GT | G | 0.191517 | 0.000996234 | 114.7609138 |
| | 6 | 31303922 | −0.0268159 | 0.00407698 | 4.80E-11 | rs28752924 | C | T | 0.445537 | 0.00375783 | 43.26134775 |
| | 6 | 32529147 | −0.0696528 | 0.0096162 | 4.40E-13 | rs71536559 | T | G | 0.052986 | 0.00455685 | 52.46414948 |
| | 6 | 139834012 | 0.0335237 | 0.00406549 | 1.60E-16 | rs632057 | G | T | 0.627778 | 0.00590492 | 67.99399346 |
| | 6 | 161125454 | −0.0685887 | 0.00769525 | 5.00E-19 | rs12212146 | C | T | 0.074303 | 0.00689847 | 79.44235834 |
| | 6 | 25582712 | −0.0388513 | 0.00667244 | 5.80E-09 | rs79681349 | G | C | 0.096452 | 0.00294514 | 33.9026939 |
| | 6 | 43757896 | −0.0285603 | 0.00393469 | 3.90E-13 | rs998584 | A | C | 0.482313 | 0.00457613 | 52.68620766 |
| | 6 | 32583057 | −0.037327 | 0.00400751 | 1.20E-20 | rs3129755 | G | A | 0.565325 | 0.0075329 | 86.75397769 |
| | 6 | 127454893 | −0.0632303 | 0.00918997 | 6.00E-12 | rs72959041 | A | G | 0.049614 | 0.00411184 | 47.33852804 |
| | 6 | 160922870 | 0.261111 | 0.014579 | 9.80E-72 | rs117733303 | G | A | 0.0185 | 0.002779576 | 320.7652526 |
| | 6 | 161010118 | 0.168768 | 0.00728501 | 9.91E-119 | rs10455872 | G | A | 0.078987 | 0.004641858 | 536.6762239 |

| | | | | | | | | | | | |
|---|---|---|---|---|---|---|---|---|---|---|---|
| | 7 | 17978828 | 0.0242365 | 0.00396382 | 9.70E-10 | rs56287412 | C | T | 0.508001 | 0.00324761 | 37.38560473 |
| | 7 | 25992323 | 0.0473398 | 0.00538053 | 1.40E-18 | rs10642257 | AAGGCC | A | 0.160158 | 0.00672207 | 77.40966278 |
| | 7 | 73123473 | -0.0642671 | 0.00913239 | 2.00E-12 | rs72555385 | G | A | 0.049304 | 0.00430144 | 49.52231737 |
| | 7 | 73020337 | 0.124664 | 0.00585823 | 1.70E-100 | rs3812316 | G | C | 0.129282 | 0.03919551 | 452.8368187 |
| | 7 | 128732786 | 0.0223148 | 0.00398441 | 2.10E-08 | rs35163364 | T | C | 0.432067 | 0.00272478 | 31.36536942 |
| | 7 | 130432481 | -0.0278279 | 0.00432559 | 1.20E-10 | rs6971365 | C | T | 0.292826 | 0.00359506 | 41.38687309 |
| | 8 | 126500031 | 0.0736808 | 0.00402649 | 8.40E-75 | rs28601761 | G | C | 0.420159 | 0.02901257 | 334.8481223 |
| | 8 | 8687054 | 0.0338985 | 0.00399236 | 2.10E-17 | rs3789849 | C | G | 0.428164 | 0.00626069 | 72.09315359 |
| | 8 | 19819724 | 0.205728 | 0.00652163 | 1.00E-200 | rs328 | G | C | 0.100307 | 0.08572902 | 995.1005103 |
| | 8 | 126631799 | -0.0373457 | 0.0048264 | 1.00E-14 | rs3860847 | A | G | 0.212508 | 0.00519998 | 59.87245084 |
| | 8 | 18239697 | -0.0321388 | 0.00511347 | 3.30E-10 | rs13266992 | C | T | 0.180568 | 0.0034314 | 39.50211434 |

| | | | | | | | | | | | |
|---|---|---|---|---|---|---|---|---|---|---|---|
| | 8 | 19733244 | 0.0750764 | 0.0047908 | 2.40E-55 | rs1561748 | C | G | 0.267445 | 0.002129402 | 245.5744894 |
| | 8 | 19980272 | 0.0428244 | 0.00772991 | 3.00E-08 | rs71205961 | CTTTTTT | C | 0.911027 | 0.000266631 | 30.69206797 |
| | 9 | 92214291 | -0.022674 | 0.00397027 | 1.10E-08 | rs3215882 | AG | A | 0.511059 | 0.000283326 | 32.61434438 |
| | 10 | 65165184 | 0.0393254 | 0.00393865 | 1.80E-23 | rs10822168 | A | G | 0.474364 | 0.000865502 | 99.68824956 |
| | 10 | 94839642 | 0.0247737 | 0.00395432 | 3.70E-10 | rs2068888 | A | G | 0.450101 | 0.000340944 | 39.24917912 |
| | 10 | 95309022 | 0.0372275 | 0.00596723 | 4.40E-10 | rs10786114 | T | C | 0.875672 | 0.000338087 | 38.92016223 |
| | 11 | 63957835 | -0.0580854 | 0.00837178 | 4.00E-12 | rs11231721 | A | G | 0.058839 | 0.000418128 | 48.13830642 |
| | 11 | 27631005 | -0.0277889 | 0.00470976 | 3.60E-09 | rs141078087 | AT | A | 0.227747 | 0.000302417 | 34.81269221 |
| | 11 | 117156893 | 0.240707 | 0.0200053 | 2.40E-33 | rs1047964 | T | G | 0.010073 | 0.001256417 | 144.7703938 |
| | 11 | 116714293 | -0.305695 | 0.0207797 | 5.50E-49 | rs141469619 | G | A | 0.010141 | 0.001877045 | 216.4166034 |
| | 11 | 61579463 | -0.0718788 | 0.00413704 | 1.30E-67 | rs174554 | G | A | 0.341712 | 0.002616237 | 301.8663117 |

| | | | | | | | | | | | |
|---|---|---|---|---|---|---|---|---|---|---|---|
| | 11 | 116389017 | −0.0259156 | 0.00429359 | 1.60E-09 | rs883863 | A | G | 0.308012 | 0.00316473 | 36.43123107 |
| | 11 | 116648917 | 0.214009 | 0.00579726 | 1.00E-200 | rs964184 | C | G | 0.86726 | 0.01170303 | 1362.733621 |
| | 11 | 65473798 | −0.0296634 | 0.00433297 | 7.60E-12 | rs10750766 | A | C | 0.709783 | 0.00407086 | 46.86654562 |
| | 12 | 124409502 | 0.02801 | 0.00422065 | 3.20E-11 | rs7133378 | A | G | 0.319789 | 0.00382555 | 44.04128592 |
| | 12 | 57692470 | 0.0293525 | 0.00460327 | 1.80E-10 | rs12313762 | T | C | 0.238672 | 0.0035318 | 40.65834984 |
| | 13 | 95231825 | −0.027191 | 0.00411737 | 4.00E-11 | rs2275647 | A | G | 0.351884 | 0.00378825 | 43.61170045 |
| | 13 | 114623821 | 0.0371442 | 0.00636686 | 5.40E-09 | rs41284486 | A | G | 0.111972 | 0.00295662 | 34.03485227 |
| | 13 | 114544024 | −0.0243037 | 0.00430044 | 1.60E-08 | rs7140110 | C | T | 0.299365 | 0.00277454 | 31.93827517 |
| | 14 | 64235556 | −0.0295206 | 0.0053774 | 4.00E-08 | rs7157785 | T | G | 0.159921 | 0.00261809 | 30.13686877 |
| | 15 | 58737341 | 0.0532876 | 0.00564463 | 3.70E-21 | rs473224 | G | T | 0.853927 | 0.00773816 | 89.11972023 |
| | 15 | 99183876 | −0.0229454 | 0.00410541 | 2.30E-08 | rs7402939 | C | T | 0.623485 | 0.00271364 | 31.2370956 |

| | | | | | | | | | | | |
|---|---|---|---|---|---|---|---|---|---|---|---|
| | 15 | 58577163 | 0.0291842 | 0.004859 | 1.90E-09 | rs10162642 | A | G | 0.210107 | 0.00313371 | 36.07399494 |
| | 15 | 43726625 | -0.177029 | 0.0124524 | 7.20E-46 | rs150844304 | C | A | 0.025671 | 0.01753127 | 202.1041163 |
| | 15 | 58678720 | 0.0600579 | 0.00414892 | 1.70E-47 | rs261290 | C | T | 0.654645 | 0.01817493 | 209.5379138 |
| | 16 | 56993886 | 0.0579917 | 0.00420869 | 3.40E-43 | rs821840 | G | A | 0.324526 | 0.01647079 | 189.8585923 |
| | 16 | 72219756 | 0.0300009 | 0.00526938 | 1.20E-08 | rs34417180 | A | C | 0.166852 | 0.00281592 | 32.41469814 |
| | 16 | 81534790 | 0.0327563 | 0.00428995 | 2.20E-14 | rs2925979 | C | T | 0.698949 | 0.00506358 | 58.30122384 |
| | 16 | 15172118 | 0.0286356 | 0.00432485 | 3.60E-11 | rs11644601 | C | T | 0.296695 | 0.00380801 | 43.83923109 |
| | 17 | 41926126 | -0.126937 | 0.011274 | 2.10E-29 | rs72836561 | T | C | 0.031482 | 0.0110036 | 126.7689379 |
| | 17 | 64210580 | 0.0903281 | 0.0114452 | 3.00E-15 | rs1801689 | C | A | 0.030526 | 0.00540949 | 62.28614183 |
| | 17 | 76395421 | 0.0222124 | 0.00402975 | 3.50E-08 | rs2292643 | A | G | 0.603709 | 0.00263945 | 30.38275949 |
| | 17 | 73516008 | 0.0253136 | 0.00454091 | 2.50E-08 | rs59476633 | A | G | 0.251287 | 0.00269959 | 31.0752399 |

| | | | | | | | | | | | |
|---|---|---|---|---|---|---|---|---|---|---|---|
| | 19 | 45413233 | -0.373861 | 0.00724448 | 1.00E-200 | rs1065853 | T | G | 0.080581 | 0.022618439 | 2663.166648 |
| | 19 | 45470298 | -0.034132 | 0.00394313 | 4.90E-18 | rs11878597 | C | T | 0.45019 | 0.000650656 | 74.92620486 |
| | 19 | 19393890 | 0.0651439 | 0.00756893 | 7.50E-18 | rs200210321 | AG | A | 0.073166 | 0.000643268 | 74.07488486 |
| | 19 | 8429323 | 0.260644 | 0.0140859 | 1.90E-76 | rs116843064 | A | G | 0.019851 | 0.002966393 | 342.3881414 |
| | 19 | 45041317 | -0.094855 | 0.0116454 | 3.80E-16 | rs144439590 | T | C | 0.03079 | 0.000576174 | 66.34437212 |
| | 19 | 8442202 | 0.0529337 | 0.0085248 | 5.30E-10 | rs62117489 | A | C | 0.056468 | 0.000334922 | 38.55569696 |
| | 19 | 45401783 | -0.0473734 | 0.00424709 | 6.80E-29 | rs56290633 | C | T | 0.539526 | 0.001079965 | 124.4166875 |
| | 20 | 44576502 | -0.050476 | 0.00506586 | 2.20E-23 | rs7679 | C | T | 0.186406 | 0.000861949 | 99.27866962 |
| exposure | chr.exposure | pos.exposure | beta.exposure | se.exposure | pval.exposure | SNP | effect_allele.exposure | other_allele.exposure | eaf.exposure | R2 | F |
| Free cholesterol to tot | 1 | 230302838 | 0.0307092 | 0.00412528 | 9.80E-14 | rs59484402 | CT | C | 0.611688 | 0.000481298 | 55.41439493 |
| | 1 | 27262545 | -0.0724692 | 0.0120374 | 1.70E-09 | rs182050989 | T | C | 0.02814 | 0.000314845 | 36.2438231 |

| | | | | | | | | | | | |
|---|---|---|---|---|---|---|---|---|---|---|---|
| al lipids ratio in small LDL \|\| id: ebi-a-GCST90092961 | 1 | 62963914 | -0.0856081 | 0.00415388 | 2.30E-94 | rs1168034 | C | G | 0.644669 | 0.003677177 | 424.7313096 |
| | 1 | 109817590 | -0.0477834 | 0.00478403 | 1.70E-23 | rs12740374 | T | G | 0.221064 | 0.00866129 | 99.76053294 |
| | 2 | 21226560 | -0.0860161 | 0.00490042 | 5.70E-69 | rs2678379 | G | A | 0.793822 | 0.0267008 | 308.0954894 |
| | 2 | 165528624 | 0.0414424 | 0.00407313 | 2.60E-24 | rs1128249 | T | G | 0.392003 | 0.00898743 | 103.5203438 |
| | 2 | 227077377 | -0.0391468 | 0.00426137 | 4.10E-20 | rs2943635 | T | C | 0.679832 | 0.00732771 | 84.38912779 |
| | 2 | 20369562 | 0.0217505 | 0.0039818 | 4.70E-08 | rs870526 | T | C | 0.521406 | 0.00259215 | 29.83816133 |
| | 2 | 27730940 | 0.0837481 | 0.00406306 | 2.10E-94 | rs1260326 | C | T | 0.603982 | 0.00378203 | 424.8502781 |
| | 3 | 135956305 | -0.0398109 | 0.00470782 | 2.80E-17 | rs684773 | C | A | 0.767075 | 0.00620994 | 71.50834795 |
| | 4 | 88018991 | -0.0333297 | 0.00406114 | 2.30E-16 | rs2035403 | G | A | 0.398315 | 0.00584932 | 67.35337136 |
| | 4 | 3098321 | 0.0252984 | 0.0042 | 1.70E-09 | rs7659144 | G | C | 0.344468 | 0.00315169 | 36.28106122 |
| | 4 | 100508556 | 0.0411107 | 0.007175 | 1.00E-08 | rs2035816 | G | A | 0.084301 | 0.00285139 | 32.82314623 |

| | | | | | | | | | | | |
|---|---|---|---|---|---|---|---|---|---|---|---|
| | 5 | 156443066 | －0.0262823 | 0.00415133 | 2.40E-10 | rs4704834 | G | A | 0.644028 | 0.00348172 | 40.08156524 |
| | 5 | 55857025 | －0.044673 | 0.00507171 | 1.30E-18 | rs11429307 | GT | G | 0.191517 | 0.00673723 | 77.58430316 |
| | 6 | 31303922 | －0.0252414 | 0.00412186 | 9.10E-10 | rs28752924 | C | T | 0.445537 | 0.00325755 | 37.50013724 |
| | 6 | 31850308 | －0.0639319 | 0.00960789 | 2.90E-11 | rs74434374 | A | C | 0.044813 | 0.00384596 | 44.27633155 |
| | 6 | 32587213 | －0.0342966 | 0.00409886 | 5.90E-17 | rs28383314 | C | T | 0.622964 | 0.00608001 | 70.01134471 |
| | 6 | 139834012 | 0.029756 | 0.00411024 | 4.50E-13 | rs632057 | G | T | 0.627778 | 0.00455208 | 52.40915887 |
| | 6 | 161125454 | －0.0695974 | 0.00777996 | 3.70E-19 | rs12212146 | C | T | 0.074303 | 0.006949 | 80.02464486 |
| | 6 | 160922870 | 0.279322 | 0.0147395 | 4.40E-80 | rs117733303 | G | A | 0.0185 | 0.003110887 | 359.1180511 |
| | 6 | 161010118 | 0.188662 | 0.00736521 | 1.00E-144 | rs10455872 | G | A | 0.078987 | 0.00569204 | 656.1317479 |
| | 6 | 43758873 | －0.0225879 | 0.00400174 | 1.70E-08 | rs6905288 | A | G | 0.568525 | 0.00276775 | 31.8600482 |
| | 6 | 160710851 | 0.0378014 | 0.00546358 | 4.60E-12 | rs3127580 | T | C | 0.156213 | 0.00415789 | 47.86891769 |

| | | | | | | | | | | | |
|---|---|---|---|---|---|---|---|---|---|---|---|
| | 6 | 26340872 | −0.0258975 | 0.00408005 | 2.20E-10 | rs55964643 | A | G | 0.503231 | 0.00349966 | 40.28813515 |
| | 6 | 127436064 | −0.0222294 | 0.00398807 | 2.50E-08 | rs1936800 | T | C | 0.52623 | 0.00269902 | 31.06865042 |
| | 7 | 73123473 | −0.0578094 | 0.00923354 | 3.80E-10 | rs72555385 | G | A | 0.049304 | 0.00340491 | 39.19700699 |
| | 7 | 17978828 | 0.0220135 | 0.00400772 | 4.00E-08 | rs56287412 | C | T | 0.508001 | 0.00262097 | 30.17004129 |
| | 7 | 25991826 | 0.0510971 | 0.00544559 | 6.40E-21 | rs4722551 | C | T | 0.1581 | 0.00764475 | 88.04304966 |
| | 7 | 73020337 | 0.113071 | 0.00592311 | 3.10E-81 | rs3812316 | G | C | 0.129282 | 0.03156621 | 364.4142257 |
| | 7 | 128732786 | 0.0224704 | 0.00402854 | 2.40E-08 | rs35163364 | T | C | 0.432067 | 0.00270272 | 31.1113387 |
| | 8 | 18248661 | −0.0345109 | 0.00513819 | 1.90E-11 | rs4646246 | G | A | 0.182596 | 0.00391845 | 45.11123364 |
| | 8 | 126500031 | 0.0683624 | 0.00407177 | 2.90E-63 | rs28601761 | G | C | 0.420159 | 0.02443421 | 281.8776084 |
| | 8 | 10649767 | −0.0353009 | 0.00410466 | 8.00E-18 | rs752446 | A | G | 0.375244 | 0.00642289 | 73.96216749 |
| | 8 | 19819724 | 0.19417 | 0.00659497 | 1.60E-190 | rs328 | G | C | 0.100307 | 0.0747605 | 866.8242749 |

| | | | | | | | | | | | |
|---|---|---|---|---|---|---|---|---|---|---|---|
| | 8 | 19733244 | 0.0702187 | 0.00484468 | 1.30E-47 | rs1561748 | C | G | 0.267445 | 0.00182216 | 210.0718351 |
| | 8 | 126629700 | -0.0350443 | 0.00487872 | 6.80E-13 | rs7005453 | A | G | 0.211239 | 0.00448147 | 51.59592988 |
| | 9 | 92214291 | -0.022241 | 0.00401334 | 3.00E-08 | rs3215882 | AG | A | 0.511059 | 0.00266792 | 30.71066109 |
| | 9 | 136149711 | 0.0285952 | 0.00514792 | 2.80E-08 | rs115478735 | T | A | 0.183304 | 0.0026804 | 30.85426113 |
| | 10 | 65176105 | 0.0353601 | 0.00399418 | 8.50E-19 | rs10640079 | GT | G | 0.474723 | 0.00680563 | 78.3725822 |
| | 10 | 104091096 | 0.0452073 | 0.00795991 | 1.40E-08 | rs17780834 | T | A | 0.067232 | 0.00280202 | 32.25472006 |
| | 11 | 64021821 | -0.042658 | 0.00703304 | 1.30E-09 | rs2302263 | T | C | 0.08812 | 0.00319572 | 36.78809144 |
| | 11 | 116648917 | 0.216949 | 0.00586065 | 1.00E-200 | rs964184 | C | G | 0.86726 | 0.01176726 | 1370.301666 |
| | 11 | 116714293 | -0.30244 | 0.0210069 | 5.40E-47 | rs141469619 | G | A | 0.010141 | 0.017979 | 207.2750244 |
| | 11 | 61606642 | -0.0796679 | 0.00415574 | 6.50E-82 | rs174580 | G | A | 0.351972 | 0.03183302 | 367.504325 |
| | 11 | 65473798 | -0.0295236 | 0.00438035 | 1.60E-11 | rs10750766 | A | C | 0.709783 | 0.00394586 | 45.42693669 |

| | | | | | | | | | | | |
|---|---|---|---|---|---|---|---|---|---|---|---|
| | 11 | 117156893 | 0.207922 | 0.0202241 | 8.60E-25 | rs1047964 | T | G | 0.010073 | 0.00917606 | 105.6951191 |
| | 11 | 116389017 | -0.0262254 | 0.00434053 | 1.50E-09 | rs883863 | A | G | 0.308012 | 0.00317113 | 36.50490056 |
| | 12 | 109885374 | -0.0279725 | 0.0046179 | 1.40E-09 | rs888190 | A | G | 0.249286 | 0.00318733 | 36.69154246 |
| | 13 | 95236825 | -0.0281817 | 0.00415345 | 1.20E-11 | rs9556403 | G | A | 0.352423 | 0.00399885 | 46.03719491 |
| | 13 | 114623821 | 0.0352759 | 0.00643592 | 4.20E-08 | rs41284486 | A | G | 0.111972 | 0.00260984 | 30.04189915 |
| | 14 | 64235556 | -0.0304656 | 0.00543611 | 2.10E-08 | rs7157785 | T | G | 0.159921 | 0.00272846 | 31.40765064 |
| | 15 | 102065008 | -0.0297156 | 0.00481684 | 6.90E-10 | rs2084152 | C | T | 0.222111 | 0.00330593 | 38.05721628 |
| | 15 | 58678720 | 0.100276 | 0.0041915 | 1.70E-126 | rs261290 | C | T | 0.654645 | 0.04948723 | 572.3313227 |
| | 15 | 58737341 | 0.102152 | 0.00570255 | 9.30E-72 | rs473224 | G | T | 0.853927 | 0.02780602 | 320.8839511 |
| | 15 | 43726625 | -0.164793 | 0.0125802 | 3.30E-39 | rs150844304 | C | A | 0.025671 | 0.0148884 | 171.5911485 |
| | 15 | 58577163 | 0.0430892 | 0.00490886 | 1.70E-18 | rs10162642 | A | G | 0.210107 | 0.00669079 | 77.04918013 |

| | | | | | | | | | | | |
|---|---|---|---|---|---|---|---|---|---|---|---|
| | 16 | 15172118 | 0.0282431 | 0.00437254 | 1.10E-10 | rs11644601 | C | T | 0.296695 | 0.00362403 | 41.72050894 |
| | 16 | 72219756 | 0.0300503 | 0.00532748 | 1.70E-08 | rs34417180 | A | C | 0.166852 | 0.00276393 | 31.81606172 |
| | 16 | 56985555 | 0.0327669 | 0.00482169 | 1.10E-11 | rs12448528 | G | A | 0.780626 | 0.00401135 | 46.1811341 |
| | 16 | 81534790 | 0.0269101 | 0.00433725 | 5.50E-10 | rs2925979 | C | T | 0.698949 | 0.00334387 | 38.49409947 |
| | 17 | 41926126 | -0.118812 | 0.011397 | 1.90E-25 | rs72836561 | T | C | 0.031482 | 0.00943457 | 108.6755806 |
| | 17 | 64210580 | 0.0885428 | 0.0115701 | 2.00E-14 | rs1801689 | C | A | 0.030526 | 0.00508632 | 58.56319071 |
| | 17 | 76395421 | 0.0224885 | 0.00407371 | 3.40E-08 | rs2292643 | A | G | 0.603709 | 0.00264739 | 30.47426301 |
| | 18 | 47109955 | -0.109384 | 0.0173966 | 3.20E-10 | rs77960347 | G | A | 0.013234 | 0.00343417 | 39.53404919 |
| | 19 | 19407718 | 0.0550856 | 0.0074945 | 2.00E-13 | rs10401969 | C | T | 0.076001 | 0.00469223 | 54.02357161 |
| | 19 | 8442202 | 0.0537107 | 0.008607 | 4.40E-10 | rs62117489 | A | C | 0.056468 | 0.0033827 | 38.9413023 |
| | 19 | 45039971 | -0.0819983 | 0.013007 | 2.90E-10 | rs74747585 | C | T | 0.025173 | 0.00345222 | 39.74183072 |

| | | | | | | | | | | | |
|---|---|---|---|---|---|---|---|---|---|---|---|
| | 19 | 45173951 | −0.119148 | 0.00920932 | 2.80E-38 | rs28807203 | C | A | 0.049207 | 0.001452378 | 167.3828037 |
| | 19 | 45337918 | 0.126939 | 0.0108774 | 1.80E-31 | rs147711004 | A | G | 0.036038 | 0.001182004 | 136.1859796 |
| | 19 | 45413233 | −0.486644 | 0.00731434 | 1.00E-200 | rs1065853 | T | G | 0.080581 | 0.037040191 | 4426.545272 |
| | 19 | 44805948 | −0.0940235 | 0.0172352 | 4.90E-08 | rs74796695 | G | T | 0.01343 | 0.00258536 | 29.75997931 |
| | 19 | 45215081 | −0.0901517 | 0.0141146 | 1.70E-10 | rs111278137 | A | G | 0.021614 | 0.00354364 | 40.79464364 |
| | 19 | 8429323 | 0.238796 | 0.0142218 | 2.80E-63 | rs116843064 | A | G | 0.019851 | 0.02443852 | 281.9274831 |
| | 21 | 30237130 | −0.0351454 | 0.00631375 | 2.60E-08 | rs2705683 | T | A | 0.885849 | 0.00269177 | 30.98522616 |
| exposure | chr.exposure | pos.exposure | beta.exposure | se.exposure | pval.exposure | SNP | effect_allele.exposure | other_allele.exposure | eaf.exposure | R2 | F |
| Free cholesterol levels in very lar | 1 | 25818452 | 0.0212949 | 0.0038699 | 3.70E-08 | rs72660961 | T | C | 0.444274 | 0.00263045 | 30.27918467 |
| | 1 | 62999675 | −0.032152 | 0.00402119 | 1.30E-15 | rs7534572 | G | C | 0.64747 | 0.00555211 | 63.92919724 |
| | 1 | 55505647 | −0.178287 | 0.01466 | 5.00E-34 | rs11591147 | T | G | 0.017457 | 0.01283531 | 147.898533 |

| | | | | | | | | | | | |
|---|---|---|---|---|---|---|---|---|---|---|---|
| ge HDL \|\| id: ebi-a-GCST90093008 | 1 | 161193683 | -0.0474699 | 0.00397457 | 7.00E-33 | rs5082 | A | G | 0.620747 | 0.001237972 | 142.6424487 |
| | 1 | 220970028 | 0.0274731 | 0.00421406 | 7.10E-11 | rs2642438 | G | A | 0.70385 | 0.00369187 | 42.5017054 |
| | 1 | 178529898 | -0.0224117 | 0.00385853 | 8.20E-09 | rs12753251 | A | G | 0.533401 | 0.00288642 | 33.2265039 |
| | 1 | 109817590 | -0.0572091 | 0.00462896 | 4.40E-35 | rs12740374 | T | G | 0.221064 | 0.01325501 | 152.7410697 |
| | 1 | 230302838 | 0.0225986 | 0.00399157 | 1.50E-08 | rs59484402 | CT | C | 0.611688 | 0.0027845 | 32.05295124 |
| | 2 | 44055510 | -0.0586474 | 0.00806623 | 3.60E-13 | rs114938914 | T | C | 0.061412 | 0.00459145 | 52.86263127 |
| | 2 | 111910083 | -0.0453551 | 0.00792804 | 1.10E-08 | rs3789067 | T | C | 0.062911 | 0.00284308 | 32.72751606 |
| | 2 | 21271707 | -0.0603671 | 0.00565154 | 1.20E-26 | rs34722314 | A | T | 0.136126 | 0.00990442 | 114.0931228 |
| | 2 | 227112754 | -0.0332643 | 0.00401163 | 1.10E-16 | rs2943654 | T | C | 0.64513 | 0.00597102 | 68.75550703 |
| | 2 | 27730940 | 0.0725692 | 0.00393161 | 4.50E-76 | rs1260326 | C | T | 0.603982 | 0.02951703 | 340.6875485 |
| | 2 | 165528876 | 0.0317312 | 0.0039402 | 8.10E-16 | rs13389219 | T | C | 0.392532 | 0.00563228 | 64.85282695 |

| | | | | | | | | | | | |
|---|---|---|---|---|---|---|---|---|---|---|---|
| | 3 | 12450838 | -0.149919 | 0.0230473 | 7.80E-11 | rs147667955 | T | C | 0.007221 | 0.00367541 | 42.31220868 |
| | 3 | 98416900 | -0.0229274 | 0.00387287 | 3.20E-09 | rs10935473 | T | G | 0.442432 | 0.00304442 | 35.04582028 |
| | 4 | 103188709 | -0.0425959 | 0.00733875 | 6.50E-09 | rs13107325 | T | C | 0.074282 | 0.00292656 | 33.6886698 |
| | 4 | 3443931 | 0.0305107 | 0.00398485 | 1.90E-14 | rs13108218 | G | A | 0.615355 | 0.00509157 | 58.62364794 |
| | 4 | 100052733 | -0.0256416 | 0.00419863 | 1.00E-09 | rs1126670 | A | C | 0.698741 | 0.00323986 | 37.29644907 |
| | 5 | 74472939 | 0.0256839 | 0.00401285 | 1.50E-10 | rs7707394 | A | G | 0.357075 | 0.00355839 | 40.96458323 |
| | 5 | 74666492 | 0.0348307 | 0.00396267 | 1.50E-18 | rs7733436 | T | C | 0.381464 | 0.00670888 | 77.25756993 |
| | 5 | 55857025 | -0.0431951 | 0.00490838 | 1.40E-18 | rs11429307 | GT | G | 0.191517 | 0.00672501 | 77.44351638 |
| | 6 | 139839768 | 0.0214649 | 0.00386229 | 2.70E-08 | rs151288714 | AAAAC | A | 0.51752 | 0.00268314 | 30.88591217 |
| | 6 | 98583488 | 0.0225346 | 0.00394756 | 1.10E-08 | rs72934503 | G | A | 0.451389 | 0.00283082 | 32.58627213 |
| | 6 | 127454893 | -0.0519779 | 0.00900599 | 7.90E-09 | rs72959041 | A | G | 0.049614 | 0.00289362 | 33.30941382 |

| | | | | | | | | | | | |
|---|---|---|---|---|---|---|---|---|---|---|---|
| | 6 | 161005389 | -0.0542019 | 0.00626534 | 5.10E-18 | rs191555775 | T | A | 0.106389 | 0.00649905 | 74.83972554 |
| | 6 | 25334241 | -0.0289023 | 0.0051516 | 2.00E-08 | rs12196438 | T | C | 0.168984 | 0.00273435 | 31.47552571 |
| | 6 | 31324586 | -0.0298158 | 0.00392229 | 2.90E-14 | rs1050556 | T | C | 0.503484 | 0.00501866 | 57.78378476 |
| | 6 | 43758873 | -0.0283226 | 0.00387892 | 2.80E-13 | rs6905288 | A | G | 0.568525 | 0.00463059 | 53.3134816 |
| | 7 | 72879234 | -0.0467783 | 0.00699909 | 2.30E-11 | rs111432585 | GT | G | 0.907696 | 0.00387998 | 44.66817057 |
| | 7 | 1029585 | -0.0341069 | 0.00396643 | 8.00E-18 | rs9769088 | C | T | 0.390936 | 0.00642094 | 73.93964476 |
| | 7 | 73033559 | 0.102042 | 0.00572137 | 3.80E-71 | rs34062580 | A | G | 0.130304 | 0.0275646 | 318.0902136 |
| | 7 | 94953895 | -0.0280866 | 0.00389067 | 5.20E-13 | rs705379 | A | G | 0.477476 | 0.00452632 | 52.11251082 |
| | 7 | 28232457 | 0.0244805 | 0.00405605 | 1.60E-09 | rs849327 | G | A | 0.656011 | 0.00316438 | 36.42725282 |
| | 8 | 19733244 | 0.0361612 | 0.00469746 | 1.40E-14 | rs1561748 | C | G | 0.267445 | 0.0051467 | 59.25867533 |
| | 8 | 126506694 | 0.0303318 | 0.00399773 | 3.30E-14 | rs112875651 | A | G | 0.392379 | 0.00499971 | 57.56544964 |

| | | | | | | | | | | | |
|---|---|---|---|---|---|---|---|---|---|---|---|
| | 8 | 11756600 | 0.036356 | 0.00396965 | 5.30E-20 | rs4841605 | A | G | 0.606733 | 0.000728323 | 83.87648216 |
| | 8 | 19820916 | 0.0920273 | 0.00639907 | 6.80E-47 | rs117199990 | T | C | 0.100233 | 0.001793959 | 206.8197892 |
| | 9 | 15298666 | 0.0412572 | 0.00556699 | 1.30E-13 | rs675849 | C | T | 0.860645 | 0.000477028 | 54.92259779 |
| | 9 | 107661742 | -0.0266889 | 0.00437297 | 1.00E-09 | rs2740488 | C | A | 0.265313 | 0.000323564 | 37.24781532 |
| | 10 | 94839642 | 0.0248782 | 0.00386859 | 1.30E-10 | rs2068888 | A | G | 0.450101 | 0.000359227 | 41.3547074 |
| | 10 | 113949664 | -0.0332211 | 0.00432481 | 1.60E-14 | rs10787429 | C | T | 0.725575 | 0.000512465 | 59.00467251 |
| | 10 | 65301725 | 0.0318091 | 0.00385347 | 1.50E-16 | rs7897379 | C | T | 0.479379 | 0.000591745 | 68.1382932 |
| | 11 | 5701074 | 0.0440674 | 0.00749637 | 4.10E-09 | rs11601507 | A | C | 0.069168 | 0.000300189 | 34.55614437 |
| | 11 | 10324061 | -0.0237266 | 0.00387895 | 9.50E-10 | rs7924631 | G | A | 0.552641 | 0.000325008 | 37.41408311 |
| | 11 | 64004643 | -0.0522742 | 0.00920092 | 1.30E-08 | rs12294913 | G | C | 0.049058 | 0.000280403 | 32.27785999 |
| | 11 | 116648917 | 0.111217 | 0.00567195 | 1.30E-85 | rs964184 | C | G | 0.86726 | 0.003329827 | 384.4766922 |

| | | | | | | | | | | | |
|---|---|---|---|---|---|---|---|---|---|---|---|
| | 11 | 47324666 | 0.0263507 | 0.00411167 | 1.50E-10 | rs4752824 | A | T | 0.321813 | 0.00356768 | 41.07147153 |
| | 11 | 61586328 | -0.103795 | 0.00404979 | 7.10E-145 | rs35473591 | CT | C | 0.34192 | 0.00567558 | 656.8713369 |
| | 11 | 117156893 | 0.208729 | 0.0195729 | 1.50E-26 | rs1047964 | T | G | 0.010073 | 0.00987231 | 113.7228363 |
| | 12 | 20582651 | 0.0277264 | 0.00484846 | 1.10E-08 | rs12369443 | G | A | 0.199473 | 0.00284085 | 32.70180971 |
| | 12 | 125347764 | -0.0315009 | 0.00406474 | 9.20E-15 | rs10692614 | TCA | T | 0.638692 | 0.00521611 | 60.05827219 |
| | 12 | 57848708 | 0.0244642 | 0.00425512 | 9.00E-09 | rs3809113 | G | C | 0.710445 | 0.00287148 | 33.05452076 |
| | 12 | 125265201 | -0.0319942 | 0.00418288 | 2.00E-14 | rs921919 | A | G | 0.66951 | 0.00508117 | 58.50381025 |
| | 12 | 123209295 | 0.0278995 | 0.00474585 | 4.10E-09 | rs3922628 | A | T | 0.210496 | 0.00300211 | 34.55869048 |
| | 15 | 43757184 | -0.0793907 | 0.0126297 | 3.30E-10 | rs190543502 | C | T | 0.024111 | 0.00343239 | 39.51351323 |
| | 15 | 58833993 | 0.0744357 | 0.0114988 | 9.60E-11 | rs6078 | A | G | 0.028853 | 0.00363992 | 41.90346782 |
| | 15 | 58568394 | -0.0340491 | 0.00392976 | 4.50E-18 | rs77732800 | G | T | 0.483171 | 0.00651911 | 75.07090834 |

| | | | | | | | | | | | |
|---|---|---|---|---|---|---|---|---|---|---|---|
| | 15 | 58680954 | -0.134605 | 0.00394454 | 1.00E-200 | rs2043085 | C | T | 0.612685 | 0.010017281 | 1164.453387 |
| | 15 | 58723939 | 0.168185 | 0.00465093 | 1.00E-200 | rs2070895 | A | G | 0.218679 | 0.011235192 | 1307.637496 |
| | 15 | 59058720 | -0.0544269 | 0.00868616 | 3.70E-10 | rs77516617 | A | G | 0.052809 | 0.00341048 | 39.26123895 |
| | 16 | 56852822 | 0.0247394 | 0.00389366 | 2.10E-10 | rs1561139 | T | G | 0.42589 | 0.00350673 | 40.36962603 |
| | 16 | 81534790 | 0.0301558 | 0.00419042 | 6.20E-13 | rs2925979 | C | T | 0.698949 | 0.00449805 | 51.78679728 |
| | 16 | 15148646 | 0.033009 | 0.00422009 | 5.20E-15 | rs11075253 | A | C | 0.295789 | 0.00531353 | 61.1806042 |
| | 16 | 56993886 | 0.133779 | 0.00411104 | 1.00E-200 | rs821840 | G | A | 0.324526 | 0.00917739 | 1058.924357 |
| | 17 | 41926126 | -0.0763793 | 0.0110279 | 4.30E-12 | rs72836561 | T | C | 0.031482 | 0.00416656 | 47.96872348 |
| | 17 | 7080316 | 0.030563 | 0.00489749 | 4.40E-10 | rs55714927 | T | C | 0.190994 | 0.00338291 | 38.94370793 |
| | 17 | 66457658 | 0.0353105 | 0.0046364 | 2.60E-14 | rs2909211 | T | C | 0.778897 | 0.00503755 | 58.0013888 |
| | 19 | 45254168 | 0.0374664 | 0.0057806 | 9.10E-11 | rs8103315 | A | C | 0.135788 | 0.00364899 | 42.00785886 |

| | | | | | | | | | | | |
|---|---|---|---|---|---|---|---|---|---|---|---|
| | 19 | 45357939 | 0.0309599 | 0.00390033 | 2.10E-15 | rs2927468 | G | A | 0.523287 | 0.00547207 | 63.00700612 |
| | 19 | 45397952 | 0.0697721 | 0.0121211 | 8.60E-09 | rs116881820 | C | T | 0.026042 | 0.00287837 | 33.13385544 |
| | 19 | 8570490 | 0.0351815 | 0.00599355 | 4.40E-09 | rs201801505 | CA | C | 0.123685 | 0.00299311 | 34.45505053 |
| | 19 | 11192915 | -0.0765873 | 0.00590066 | 1.60E-38 | rs73015021 | G | A | 0.120658 | 0.01461738 | 168.46310002 |
| | 19 | 45413233 | -0.304976 | 0.00709647 | 1.00E-200 | rs1065853 | T | G | 0.080581 | 0.01579518 3 | 1846.88153 |
| | 19 | 45173951 | -0.0724142 | 0.00893501 | 5.30E-16 | rs28807203 | C | A | 0.049207 | 0.0057043 | 65.68252516 |
| | 20 | 17844684 | -0.0247113 | 0.00406336 | 1.20E-09 | rs2618566 | T | G | 0.660294 | 0.00321273 | 36.98392814 |
| | 20 | 44105636 | -0.0477426 | 0.00602382 | 2.30E-15 | rs6065819 | A | C | 0.116575 | 0.00545536 | 62.8146021 |
| | 20 | 44776543 | -0.0752732 | 0.0106806 | 1.80E-12 | rs2024570 | C | T | 0.962637 | 0.00431414 | 49.66859892 |
| | 20 | 44055710 | 0.0295493 | 0.00387862 | 2.60E-14 | rs3092498 | C | T | 0.517382 | 0.00504096 | 58.04066527 |
| | 20 | 44563007 | -0.22351 | 0.00494722 | 1.00E-200 | rs1057208 | T | C | 0.186277 | 0.01727247 | 2041.098306 |

| exposure | chr.exposure | pos.exposure | beta.exposure | se.exposure | pval.exposure | SNP | effect_allele.exposure | other_allele.exposure | eaf.exposure | R2 | F |
|---|---|---|---|---|---|---|---|---|---|---|---|
| Free cholesterol levels in large HDL \|\| id:ebi-a-GCST90092848 | 1 | 40083641 | -0.0273301 | 0.00439418 | 5.00E-10 | rs61781392 | A | C | 0.216719 | 0.00336026 | 38.68291627 |
| | 1 | 161193683 | -0.0256812 | 0.00373543 | 6.20E-12 | rs5082 | A | G | 0.620747 | 0.00410548 | 47.26524068 |
| | 1 | 178526271 | -0.022855 | 0.00362744 | 3.00E-10 | rs6681348 | C | A | 0.532241 | 0.0034483 | 39.69670211 |
| | 1 | 26987646 | -0.0756645 | 0.0122192 | 5.90E-10 | rs193084249 | G | A | 0.023362 | 0.00333079 | 38.34345463 |
| | 1 | 182119332 | -0.0326159 | 0.00387095 | 3.60E-17 | rs6694102 | A | G | 0.320921 | 0.00616522 | 70.99308728 |
| | 1 | 220970028 | 0.0331066 | 0.00396052 | 6.30E-17 | rs2642438 | G | A | 0.70385 | 0.00606811 | 69.87425559 |
| | 1 | 110470764 | -0.0288007 | 0.00505515 | 1.20E-08 | rs333947 | A | G | 0.150128 | 0.00281973 | 32.45864945 |
| | 1 | 230303848 | 0.042444 | 0.0037089 | 2.50E-30 | rs1546954 | G | T | 0.612656 | 0.01136687 | 130.958839 |
| | 2 | 27730940 | 0.0320993 | 0.00369578 | 3.80E-18 | rs1260326 | C | T | 0.603982 | 0.00655069 | 75.43473238 |
| | 2 | 227095876 | -0.0393663 | 0.00377263 | 1.70E-25 | rs1024137 | T | G | 0.646501 | 0.00945241 | 108.8812806 |

| | | | | | | | | | | | |
|---|---|---|---|---|---|---|---|---|---|---|---|
| | 2 | 21231524 | 0.057836 | 0.00445874 | 1.80E-38 | rs676210 | A | G | 0.205766 | 0.001459924 | 168.2537118 |
| | 2 | 165528876 | 0.0353216 | 0.00370385 | 1.50E-21 | rs13389219 | T | C | 0.392532 | 0.0078963 | 90.94239937 |
| | 2 | 242237902 | 0.0238659 | 0.00377027 | 2.50E-10 | rs59104589 | T | C | 0.358836 | 0.00348058 | 40.06850887 |
| | 2 | 20363666 | -0.0202477 | 0.00363378 | 2.50E-08 | rs35135293 | T | C | 0.516755 | 0.00269718 | 31.0474965 |
| | 2 | 219699999 | -0.0567464 | 0.0093615 | 1.30E-09 | rs78058190 | A | G | 0.050108 | 0.00319183 | 36.74330342 |
| | 3 | 185804583 | -0.0309233 | 0.00528738 | 5.00E-09 | rs57912727 | C | A | 0.136685 | 0.00297135 | 34.20448627 |
| | 3 | 12206489 | -0.0897512 | 0.0149363 | 1.90E-09 | rs11922042 | C | T | 0.017207 | 0.00313654 | 36.1066274 |
| | 4 | 970112 | 0.0219546 | 0.00366053 | 2.00E-09 | rs12510382 | G | A | 0.459916 | 0.00312478 | 35.97125327 |
| | 4 | 89704367 | -0.0213503 | 0.00371714 | 9.30E-09 | rs13118477 | A | G | 0.390372 | 0.00286588 | 32.99002003 |
| | 4 | 100045141 | -0.0246625 | 0.00393917 | 3.80E-10 | rs1139490 | T | C | 0.696979 | 0.00340494 | 39.19739464 |
| | 4 | 103188709 | -0.0665832 | 0.00689695 | 4.70E-22 | rs13107325 | T | C | 0.074282 | 0.00809201 | 93.19822143 |

| | | | | | | | | | | | |
|---|---|---|---|---|---|---|---|---|---|---|---|
| | 4 | 69482383 | 0.0285236 | 0.0049969 | 1.10E-08 | rs368166328 | CT | C | 0.240065 | 0.000283059 | 32.58365591 |
| | 5 | 55857025 | -0.04437 | 0.00461226 | 6.60E-22 | rs11429307 | GT | G | 0.191517 | 0.00803516 | 92.54303696 |
| | 5 | 75003678 | 0.0271232 | 0.00370274 | 2.40E-13 | rs2307111 | C | T | 0.396595 | 0.00466043 | 53.65718314 |
| | 6 | 34335088 | -0.0598587 | 0.00886533 | 1.50E-11 | rs147627829 | A | G | 0.04407 | 0.00395991 | 45.58869927 |
| | 6 | 43757896 | -0.0345461 | 0.00362424 | 1.50E-21 | rs998584 | A | C | 0.482313 | 0.00788886 | 90.85662609 |
| | 6 | 161092438 | -0.0544669 | 0.00600638 | 1.20E-19 | rs11751347 | T | C | 0.102257 | 0.00714039 | 82.23035377 |
| | 6 | 31287454 | -0.0277589 | 0.00394288 | 1.90E-12 | rs9265113 | T | C | 0.407634 | 0.0043051 | 49.56439823 |
| | 6 | 127519234 | -0.0276723 | 0.00363909 | 2.90E-14 | rs10457487 | A | C | 0.484321 | 0.00502203 | 57.8225678 |
| | 6 | 98583488 | 0.0248027 | 0.00371038 | 2.30E-11 | rs72934503 | G | A | 0.451389 | 0.00388137 | 44.68415939 |
| | 7 | 28224053 | 0.0222535 | 0.00380511 | 5.00E-09 | rs849336 | G | A | 0.655418 | 0.00297116 | 34.2022496 |
| | 7 | 1066297 | -0.03456 | 0.00367795 | 5.60E-21 | rs3735687 | G | A | 0.422805 | 0.00766647 | 88.29340757 |

| | | | | | | | | | | | |
|---|---|---|---|---|---|---|---|---|---|---|---|
| | 7 | 73037956 | 0.0677603 | 0.00530173 | 2.10E-37 | rs34060476 | G | A | 0.13482 | 0.001417399 | 163.345765 |
| | 7 | 94953895 | -0.0261218 | 0.00365589 | 9.00E-13 | rs705379 | A | G | 0.477476 | 0.000443425 | 51.05195906 |
| | 7 | 73123473 | -0.0532898 | 0.00839741 | 2.20E-10 | rs72555385 | G | A | 0.049304 | 0.000349815 | 40.27076847 |
| | 7 | 130437476 | -0.0295052 | 0.00402147 | 2.20E-13 | rs7810507 | A | G | 0.281085 | 0.000467538 | 53.82944617 |
| | 8 | 19733244 | 0.0503444 | 0.00441158 | 3.60E-30 | rs1561748 | C | G | 0.267445 | 0.001130356 | 130.2286303 |
| | 8 | 19820933 | 0.132926 | 0.00600747 | 1.70E-108 | rs145391587 | C | A | 0.100287 | 0.004236289 | 489.5861273 |
| | 8 | 144305667 | 0.0307642 | 0.00548127 | 2.00E-08 | rs79744701 | A | G | 0.12409 | 0.000273655 | 31.50078045 |
| | 8 | 9173358 | 0.033733 | 0.00364723 | 2.30E-20 | rs7012814 | A | G | 0.474265 | 0.000742769 | 85.54134648 |
| | 8 | 126506694 | 0.0290893 | 0.00375443 | 9.30E-15 | rs112875651 | A | G | 0.392379 | 0.000521368 | 60.03036315 |
| | 8 | 116610180 | 0.0259213 | 0.00366203 | 1.50E-12 | rs2737205 | C | T | 0.561358 | 0.000435184 | 50.10282497 |
| | 9 | 15303583 | 0.0483342 | 0.00526964 | 4.60E-20 | rs1215112 | A | G | 0.86303 | 0.000730504 | 84.12781859 |

| | | | | | | | | | | | |
|---|---|---|---|---|---|---|---|---|---|---|---|
| | 9 | 107647019 | 0.0379374 | 0.00582593 | 7.40E-11 | rs11789603 | T | C | 0.108822 | 0.00036833 | 42.40304603 |
| | 9 | 107661742 | -0.0347326 | 0.00411027 | 2.90E-17 | rs2740488 | C | A | 0.265313 | 0.00620093 | 71.40462447 |
| | 10 | 65191645 | 0.021735 | 0.00361793 | 1.90E-09 | rs7924036 | T | G | 0.504241 | 0.00313513 | 36.09037725 |
| | 10 | 94839724 | 0.0252891 | 0.00363689 | 3.60E-12 | rs4418728 | T | G | 0.450703 | 0.00419968 | 48.35026607 |
| | 10 | 113934384 | -0.0390825 | 0.00404757 | 4.60E-22 | rs2803619 | C | G | 0.724355 | 0.008095 | 93.23272734 |
| | 10 | 45998984 | 0.0310674 | 0.00434632 | 8.80E-13 | rs148063610 | CAAATAAAT | C | 0.236628 | 0.00443778 | 51.09270239 |
| | 10 | 113983758 | -0.0325037 | 0.00518338 | 3.60E-10 | rs60847460 | T | C | 0.14333 | 0.00341573 | 39.32167025 |
| | 11 | 47362465 | 0.0377179 | 0.00464383 | 4.60E-16 | rs10769254 | C | G | 0.18914 | 0.0057291 | 65.96823069 |
| | 11 | 118957388 | -0.02491 | 0.00381282 | 6.40E-11 | rs141949189 | T | A | 0.431973 | 0.00370754 | 42.68224961 |
| | 11 | 64004643 | -0.0517449 | 0.00864653 | 2.20E-09 | rs12294913 | G | C | 0.049058 | 0.00311106 | 35.81325962 |
| | 11 | 47322007 | -0.0412926 | 0.00442919 | 1.10E-20 | rs200644264 | CG | C | 0.788465 | 0.00754676 | 86.91371015 |

| | | | | | | | | | | | |
|---|---|---|---|---|---|---|---|---|---|---|---|
| | 11 | 116916060 | -0.128365 | 0.0183061 | 2.30E-12 | rs144018203 | C | G | 0.010653 | 0.00427079 | 49.16930184 |
| | 11 | 117156893 | 0.21598 | 0.0183936 | 7.70E-32 | rs1047964 | T | G | 0.010073 | 0.01196647 | 137.8751667 |
| | 11 | 61586328 | -0.0852872 | 0.00380578 | 3.20E-111 | rs35473591 | CT | C | 0.34192 | 0.0043449221 | 502.1954882 |
| | 11 | 75531695 | -0.0407718 | 0.00590072 | 4.90E-12 | rs10899133 | T | C | 0.105503 | 0.00414689 | 47.74217591 |
| | 11 | 116648917 | 0.0957812 | 0.0053302 | 3.40E-72 | rs964184 | C | G | 0.86726 | 0.00279801 | 322.8984722 |
| | 12 | 20470199 | 0.0278135 | 0.00456822 | 1.10E-09 | rs11045171 | G | A | 0.197429 | 0.0032201 | 37.06890155 |
| | 12 | 57848708 | 0.0261742 | 0.00399721 | 5.80E-11 | rs3809113 | G | C | 0.710445 | 0.00372446 | 42.87709516 |
| | 12 | 109871179 | 0.0249983 | 0.00361433 | 4.60E-12 | rs7308864 | G | A | 0.523292 | 0.00415506 | 47.83632399 |
| | 12 | 124620185 | 0.0266491 | 0.0037825 | 1.80E-12 | rs10846606 | G | C | 0.366512 | 0.00431134 | 49.636331 |
| | 12 | 123209295 | 0.0332537 | 0.00445819 | 8.70E-14 | rs3922628 | A | T | 0.210496 | 0.00483221 | 55.63591751 |
| | 12 | 125265201 | -0.0426255 | 0.00392934 | 2.00E-27 | rs921919 | A | G | 0.66951 | 0.01021524 | 117.6771675 |

| | | | | | | | | | | | |
|---|---|---|---|---|---|---|---|---|---|---|---|
| | 12 | 112083162 | −0.0200743 | 0.0036757 | 4.70E-08 | rs11065991 | T | C | 0.415862 | 0.00259108 | 29.82587244 |
| | 12 | 125349480 | 0.0380004 | 0.00378222 | 9.50E-24 | rs36226283 | A | G | 0.358958 | 0.00876384 | 100.9427693 |
| | 15 | 42091212 | −0.0212858 | 0.00384966 | 3.20E-08 | rs2925339 | G | A | 0.673024 | 0.0026559 | 30.57226798 |
| | 15 | 43757184 | −0.0705361 | 0.0118589 | 2.70E-09 | rs190543502 | C | T | 0.024111 | 0.00307322 | 35.37744998 |
| | 15 | 58737661 | −0.159244 | 0.00515474 | 1.00E-200 | rs488490 | A | C | 0.852654 | 0.008224671 | 954.3442885 |
| | 15 | 58580781 | −0.0694391 | 0.00438646 | 1.90E-56 | rs1560390 | C | T | 0.219673 | 0.0217284 | 250.5949254 |
| | 15 | 58678512 | 0.165252 | 0.00395984 | 1.00E-200 | rs10468017 | T | C | 0.295937 | 0.0149076 | 1741.528654 |
| | 15 | 59058720 | −0.0525563 | 0.00815605 | 1.20E-10 | rs77516617 | A | G | 0.052809 | 0.00360683 | 41.52238198 |
| | 16 | 56752882 | −0.0817807 | 0.0124615 | 5.30E-11 | rs75714888 | A | C | 0.021624 | 0.00374103 | 43.06787619 |
| | 16 | 57151796 | −0.0236635 | 0.00404359 | 4.90E-09 | rs8058512 | T | C | 0.724045 | 0.002975 | 34.24649938 |
| | 16 | 15130594 | −0.0313282 | 0.00392276 | 1.40E-15 | rs6498540 | G | A | 0.302064 | 0.0055391 | 63.77931659 |

| | | | | | | | | | | | |
|---|---|---|---|---|---|---|---|---|---|---|---|
| | 16 | 56993324 | 0.204574 | 0.00384784 | 1.00E-200 | rs3764261 | A | C | 0.324412 | 0.02397294 1 | 2826.567208 |
| | 16 | 57049137 | −0.136175 | 0.0106236 | 1.30E-37 | rs75911530 | A | G | 0.032885 | 0.001425687 | 164.3023593 |
| | 16 | 67976320 | −0.0828058 | 0.00998623 | 1.10E-16 | rs4986970 | T | A | 0.033919 | 0.00597106 | 68.75603726 |
| | 16 | 81534790 | 0.0357845 | 0.00392435 | 7.60E-20 | rs2925979 | C | T | 0.698949 | 0.00721994 | 83.14705887 |
| | 17 | 37808759 | −0.038076 | 0.00669617 | 1.30E-08 | rs34695955 | G | T | 0.079539 | 0.0028088 | 32.33271298 |
| | 17 | 41926126 | −0.136505 | 0.010365 | 1.30E-39 | rs72836561 | T | C | 0.031482 | 0.01504863 | 173.4406762 |
| | 18 | 47109955 | 0.195543 | 0.0158169 | 4.10E-35 | rs77960347 | G | A | 0.013234 | 0.0132635 | 152.8390263 |
| | 18 | 47160953 | 0.054657 | 0.00475516 | 1.40E-30 | rs7241918 | T | G | 0.8229 | 0.01146715 | 132.1154975 |
| | 18 | 57861961 | −0.0234634 | 0.00411995 | 1.20E-08 | rs1457489 | A | G | 0.26462 | 0.00281753 | 32.43324856 |
| | 18 | 47126172 | 0.066512 | 0.010128 | 5.10E-11 | rs9955201 | A | G | 0.033777 | 0.00374613 | 43.12658616 |
| | 19 | 45413233 | −0.103029 | 0.00667843 | 1.10E-53 | rs1065853 | T | G | 0.080581 | 0.0206379 | 237.9921679 |

| | | | | | | | | | | | |
|---|---|---|---|---|---|---|---|---|---|---|---|
| | 19 | 11347493 | -0.0487456 | 0.00679416 | 7.30E-13 | rs737337 | C | T | 0.076789 | 0.000447093 | 51.4744545 |
| | 19 | 8429323 | 0.187437 | 0.0129853 | 3.10E-47 | rs116843064 | A | G | 0.019851 | 0.001807231 | 208.3526969 |
| | 19 | 45444524 | 0.0351453 | 0.00420555 | 6.40E-17 | rs72235114 | CT | C | 0.294931 | 0.00606482 | 69.8363223 |
| | 20 | 44055710 | 0.0232257 | 0.00364647 | 1.90E-10 | rs3092498 | C | T | 0.517382 | 0.00352397 | 40.56811108 |
| | 20 | 44776543 | -0.0564791 | 0.0100413 | 1.90E-08 | rs2024570 | C | T | 0.962637 | 0.00274833 | 31.63647609 |
| | 20 | 43008981 | -0.0668026 | 0.0110164 | 1.30E-09 | rs150224153 | T | C | 0.029237 | 0.00319419 | 36.77052164 |
| | 20 | 44113286 | -0.0571827 | 0.00862964 | 3.40E-11 | rs76070947 | G | C | 0.046949 | 0.00381391 | 43.90726748 |
| | 20 | 44563007 | -0.175697 | 0.00465111 | 1.00E-200 | rs1057208 | T | C | 0.186277 | 0.0122477444 | 1426.947263 |

| exposure | chr.exposure | pos.exposure | beta.exposure | se.exposure | pval.exposure | SNP | effect_allele.exposure | other_allele.exposure | eaf.exposure | R2 | F |
|---|---|---|---|---|---|---|---|---|---|---|---|
| Free cholesterol levels | 1 | 40035928 | -0.0377852 | 0.00473016 | 1.40E-15 | rs3768321 | T | G | 0.196524 | 0.00554171 | 63.80933823 |
| | 1 | 182154990 | -0.026676 | 0.00397384 | 1.90E-11 | rs61805076 | C | T | 0.33405 | 0.0039142 | 45.06227524 |

| | | | | | | | | | | | |
|---|---|---|---|---|---|---|---|---|---|---|---|
| in medium HDL \|\| id: ebi-a-GCST90092896 | 1 | 26987646 | -0.0968817 | 0.0126809 | 2.20E-14 | rs193084249 | G | A | 0.023362 | 0.00506939 | 58.36813254 |
| | 1 | 109822166 | -0.0302259 | 0.00446357 | 1.30E-11 | rs599839 | A | G | 0.77208 | 0.00398303 | 45.85495111 |
| | 1 | 110232983 | 0.0334958 | 0.00420195 | 1.60E-15 | rs140584594 | G | A | 0.730101 | 0.00551864 | 63.54353936 |
| | 1 | 150940625 | 0.0341585 | 0.00452864 | 4.60E-14 | rs267738 | G | T | 0.219652 | 0.00494129 | 56.89242406 |
| | 1 | 220970028 | 0.036829 | 0.00411018 | 3.20E-19 | rs2642438 | G | A | 0.70385 | 0.00697185 | 80.28799687 |
| | 1 | 230304914 | 0.049556 | 0.00384932 | 6.30E-38 | rs10864728 | G | A | 0.61323 | 0.0143811 | 165.7360212 |
| | 1 | 62906489 | -0.0399158 | 0.0039579 | 6.40E-24 | rs638714 | T | G | 0.345938 | 0.00883017 | 101.7073863 |
| | 2 | 165508389 | 0.0228023 | 0.00382123 | 2.40E-09 | rs10184004 | T | C | 0.406103 | 0.00309321 | 35.60765762 |
| | 2 | 227092802 | -0.0305911 | 0.00391679 | 5.70E-15 | rs2176040 | G | A | 0.646742 | 0.00529776 | 60.99890867 |
| | 2 | 20363666 | -0.0271563 | 0.00377161 | 6.00E-13 | rs35135293 | T | C | 0.516755 | 0.00450282 | 51.84180221 |
| | 2 | 21231524 | 0.0644724 | 0.00462785 | 4.10E-44 | rs676210 | A | G | 0.205766 | 0.01683643 | 194.0803554 |

| | | | | | | | | | | | |
|---|---|---|---|---|---|---|---|---|---|---|---|
| | 2 | 27730940 | -0.0328273 | 0.00383596 | 1.20E-17 | rs1260326 | C | T | 0.603982 | 0.00635973 | 73.23431788 |
| | 2 | 211540507 | -0.0231807 | 0.00404209 | 9.80E-09 | rs1047891 | A | C | 0.314984 | 0.002857 | 32.88770642 |
| | 4 | 103188709 | -0.0703087 | 0.00715515 | 8.70E-23 | rs13107325 | T | C | 0.074282 | 0.00838318 | 96.55462976 |
| | 4 | 69373407 | 0.0253536 | 0.00448561 | 1.60E-08 | rs9884390 | C | T | 0.234037 | 0.00277529 | 31.94689823 |
| | 5 | 55861894 | -0.0258316 | 0.00467388 | 3.30E-08 | rs9687846 | A | G | 0.201404 | 0.00265354 | 30.54498743 |
| | 6 | 161092438 | -0.0366523 | 0.00623137 | 4.10E-09 | rs11751347 | T | C | 0.102257 | 0.00300537 | 34.59615541 |
| | 6 | 34335088 | -0.0531585 | 0.00919741 | 7.50E-09 | rs147627829 | A | G | 0.04407 | 0.00290189 | 33.40463989 |
| | 6 | 43764551 | -0.0240497 | 0.0037802 | 2.00E-10 | rs1358980 | T | C | 0.482622 | 0.00351584 | 40.47456812 |
| | 6 | 127468274 | -0.0223858 | 0.00377258 | 3.00E-09 | rs7741021 | C | A | 0.477254 | 0.00305864 | 35.20957436 |
| | 6 | 31319923 | -0.0355387 | 0.00470225 | 5.20E-14 | rs36057735 | G | C | 0.19903 | 0.00491875 | 56.63286259 |
| | 7 | 1066297 | -0.0241448 | 0.00381541 | 2.50E-10 | rs3735687 | G | A | 0.422805 | 0.00347861 | 40.04582542 |

| | | | | | | | | | | | |
|---|---|---|---|---|---|---|---|---|---|---|---|
| | 7 | 36192606 | -0.0212262 | 0.00380522 | 2.40E-08 | rs12533197 | G | T | 0.439178 | 0.000270309 | 31.11554519 |
| | 7 | 130437476 | -0.0230121 | 0.00417178 | 3.50E-08 | rs7810507 | A | G | 0.281085 | 0.000264331 | 30.42721212 |
| | 8 | 19766233 | -0.128648 | 0.0141558 | 1.00E-19 | rs139915535 | G | A | 0.018003 | 0.000717164 | 82.59042369 |
| | 8 | 144302570 | 0.0303942 | 0.00481665 | 2.80E-10 | rs7817574 | C | T | 0.185333 | 0.000345886 | 39.818391 |
| | 8 | 9184231 | 0.0986528 | 0.00653607 | 1.80E-51 | rs4240624 | A | G | 0.909234 | 0.00197569 2 | 227.812681 |
| | 8 | 19824667 | 0.0876187 | 0.00415169 | 7.30E-99 | rs15285 | T | C | 0.286526 | 0.00385530 6 | 445.3856568 |
| | 8 | 116624879 | 0.03372 | 0.00380719 | 8.20E-19 | rs2245221 | A | G | 0.558968 | 0.000681181 | 78.44377442 |
| | 8 | 19750044 | -0.06699 | 0.00923033 | 3.90E-13 | rs59347135 | G | C | 0.045803 | 0.000457488 | 52.67177473 |
| | 9 | 107661742 | -0.0429908 | 0.00426288 | 6.40E-24 | rs2740488 | C | A | 0.265313 | 0.000882986 | 101.703826 |
| | 9 | 107647019 | 0.0555527 | 0.00604225 | 3.80E-20 | rs11789603 | T | C | 0.108822 | 0.000733984 | 84.52893842 |
| | 9 | 15304782 | 0.0446113 | 0.00540561 | 1.50E-16 | rs686030 | A | C | 0.859548 | 0.000591474 | 68.10716325 |

| | | | | | | | | | | | |
|---|---|---|---|---|---|---|---|---|---|---|---|
| | 9 | 107586753 | 0.0322456 | 0.00558023 | 7.50E-09 | rs2066714 | C | T | 0.12893 | 0.00290007 | 33.39098095 |
| | 10 | 45998984 | 0.0344939 | 0.00450825 | 2.00E-14 | rs148063610 | CAAATAAAT | C | 0.236628 | 0.00508441 | 58.54112574 |
| | 10 | 113921825 | -0.0409963 | 0.00418036 | 1.10E-22 | rs2792735 | A | G | 0.72034 | 0.0083501 | 96.17327611 |
| | 10 | 113983758 | -0.0375285 | 0.00537649 | 2.90E-12 | rs60847460 | T | C | 0.14333 | 0.00423188 | 48.72111065 |
| | 11 | 75451281 | -0.0633171 | 0.00516029 | 1.30E-34 | rs559355 | T | A | 0.157979 | 0.01306528 | 150.5519085 |
| | 11 | 61605499 | -0.052057 | 0.00392453 | 3.70E-40 | rs174578 | A | T | 0.350977 | 0.01526554 | 175.9443879 |
| | 11 | 116727936 | 0.0673791 | 0.00479735 | 8.30E-45 | rs625145 | T | A | 0.188128 | 0.01711183 | 197.2604661 |
| | 11 | 116916060 | -0.123166 | 0.0189956 | 8.90E-11 | rs144018203 | C | G | 0.010653 | 0.00365182 | 42.04052156 |
| | 11 | 47475189 | -0.036707 | 0.00391809 | 7.40E-21 | rs35184771 | T | G | 0.352771 | 0.00762097 | 87.76905465 |
| | 11 | 126225876 | -0.0429592 | 0.00728099 | 3.60E-09 | rs112771035 | G | C | 0.07137 | 0.00302408 | 34.81159751 |
| | 12 | 103556109 | -0.0216409 | 0.00377671 | 1.00E-08 | rs7956099 | C | T | 0.474982 | 0.00285228 | 32.83339658 |

| | | | | | | | | | | | |
|---|---|---|---|---|---|---|---|---|---|---|---|
| | 12 | 125324798 | -0.0566485 | 0.0057719 | 9.70E-23 | rs61941676 | A | C | 0.127218 | 0.000836312 | 96.3233737 |
| | 12 | 109873227 | 0.0298977 | 0.00374955 | 1.50E-15 | rs6606717 | C | A | 0.523165 | 0.000552166 | 63.57841743 |
| | 12 | 125259888 | -0.03916 | 0.00408403 | 8.90E-22 | rs838876 | G | A | 0.681442 | 0.000798276 | 91.93904143 |
| | 12 | 123796238 | 0.0378927 | 0.00691508 | 4.30E-08 | rs4759375 | T | C | 0.084883 | 0.000260853 | 30.02680598 |
| | 14 | 24759319 | -0.0214452 | 0.00377536 | 1.30E-08 | rs941507 | C | G | 0.46081 | 0.000280294 | 32.26531533 |
| | 15 | 58569330 | 0.0363983 | 0.00556113 | 5.90E-11 | rs34663616 | A | C | 0.137646 | 0.000372106 | 42.83793798 |
| | 15 | 58680954 | -0.111594 | 0.00384854 | 7.29E-185 | rs2043085 | C | T | 0.612685 | 0.007253053 | 840.7795354 |
| | 15 | 58723939 | 0.122025 | 0.00453775 | 2.80E-159 | rs2070895 | A | G | 0.218679 | 0.006244371 | 723.1176369 |
| | 16 | 15118254 | -0.0241595 | 0.00435869 | 3.00E-08 | rs4985126 | C | T | 0.562849 | 0.000266895 | 30.72248721 |
| | 16 | 56993324 | 0.18181 | 0.00399877 | 1.00E-200 | rs3764261 | A | C | 0.324412 | 0.017645881 | 2067.164964 |
| | 16 | 67976320 | -0.0758808 | 0.0103779 | 2.60E-13 | rs4986970 | T | A | 0.033919 | 0.00046434 | 53.46102622 |

| | | | | | | | | | | | |
|---|---|---|---|---|---|---|---|---|---|---|---|
| | 16 | 57049137 | －0.144979 | 0.0110403 | 2.20E-39 | rs75911530 | A | G | 0.032885 | 0.001496204 | 172.4411469 |
| | 16 | 57151796 | －0.0252074 | 0.0042022 | 2.00E-09 | rs8058512 | T | C | 0.724045 | 0.000312579 | 35.9828138 |
| | 16 | 81534790 | 0.0303773 | 0.00407828 | 9.40E-14 | rs2925979 | C | T | 0.698949 | 0.000481867 | 55.48003338 |
| | 17 | 26713970 | －0.0266925 | 0.00375281 | 1.10E-12 | rs967645 | T | C | 0.516505 | 0.000439407 | 50.58919873 |
| | 17 | 41926126 | －0.135899 | 0.0107492 | 1.20E-36 | rs72836561 | T | C | 0.031482 | 0.00138698 | 159.8352952 |
| | 17 | 66823805 | －0.0524937 | 0.00807267 | 7.90E-11 | rs112001035 | A | G | 0.05973 | 0.000367293 | 42.28364552 |
| | 17 | 76391653 | 0.0216023 | 0.00376575 | 9.70E-09 | rs4969141 | T | C | 0.489528 | 0.000285868 | 32.90708923 |
| | 18 | 47109955 | 0.303811 | 0.0163922 | 1.10E-76 | rs77960347 | G | A | 0.013234 | 0.002975985 | 343.4985849 |
| | 18 | 47120499 | 0.0881943 | 0.00939765 | 6.30E-21 | rs12608026 | G | T | 0.042399 | 0.000764721 | 88.07141109 |
| | 18 | 47160953 | 0.0905511 | 0.00492811 | 2.10E-75 | rs7241918 | T | G | 0.8229 | 0.002925143 | 337.6129735 |
| | 19 | 54814524 | 0.0233901 | 0.0040076 | 5.30E-09 | rs1761457 | G | A | 0.326718 | 0.00029591 | 34.06339053 |

| | | | | | | | | | | | |
|---|---|---|---|---|---|---|---|---|---|---|---|
| | 19 | 11347493 | -0.0693601 | 0.00704576 | 7.30E-23 | rs737337 | C | T | 0.076789 | 0.00841377 | 96.90723052 |
| | 19 | 8429323 | 0.168157 | 0.0134662 | 8.80E-36 | rs116843064 | A | G | 0.019851 | 0.01353145 | 155.9309171 |
| | 19 | 11243260 | -0.0234356 | 0.00376671 | 4.90E-10 | rs2978615 | T | C | 0.490198 | 0.00336259 | 38.70973836 |
| | 19 | 45421254 | -0.0483057 | 0.00516023 | 7.90E-21 | rs12721046 | A | G | 0.157696 | 0.00760888 | 87.62965332 |
| | 19 | 45330999 | -0.0259258 | 0.00467201 | 2.90E-08 | rs6509173 | G | A | 0.793731 | 0.00267506 | 30.79280543 |
| | 19 | 45448465 | 0.0476713 | 0.00392528 | 6.10E-34 | rs5167 | G | T | 0.352266 | 0.01279997 | 147.4908777 |
| | 20 | 17597531 | -0.0293294 | 0.00502564 | 5.30E-09 | rs2236252 | T | C | 0.167033 | 0.00295861 | 34.05775823 |
| | 20 | 46476143 | -0.0260408 | 0.00458823 | 1.40E-08 | rs3859588 | A | T | 0.217254 | 0.00279827 | 32.2114842 |
| | 20 | 43042364 | -0.118948 | 0.0109634 | 2.00E-27 | rs1800961 | T | C | 0.0302 | 0.01021815 | 117.7107756 |
| | 20 | 46340596 | 0.0370022 | 0.00464626 | 1.70E-15 | rs4239651 | C | T | 0.794242 | 0.00550809 | 63.42208672 |
| | 22 | 21982892 | -0.0408748 | 0.00486199 | 4.20E-17 | rs2298428 | T | C | 0.182552 | 0.00613775 | 70.67659385 |

| exposure | chr.exposure | pos.exposure | beta.exposure | se.exposure | pval.exposure | SNP | effect_allele.exposure | other_allele.exposure | eaf.exposure | R2 | F |
|---|---|---|---|---|---|---|---|---|---|---|---|
| Free cholesterol levels in small HDL \|\| id:ebi-a-GCST90092948 | 1 | 220970028 | 0.026725 4 | 0.0044306 2 | 1.60E-09 | rs2642438 | G | A | 0.70385 | 0.000316064 | 36.38411982 |
| | 1 | 161194641 | 0.029280 9 | 0.0041753 8 | 2.30E-12 | rs4656292 | G | A | 0.619567 | 0.000427153 | 49.17782101 |
| | 1 | 62963737 | 0.063085 9 | 0.0042254 1 | 2.10E-50 | rs1002687 | A | G | 0.644763 | 0.0019332 1 | 222.9047623 |
| | 1 | 110229787 | 0.032116 9 | 0.0048071 | 2.40E-11 | rs36209093 | T | C | 0.674022 | 0.000387726 | 44.63683345 |
| | 1 | 23792009 | 0.037664 9 | 0.0066458 9 | 1.40E-08 | rs612177 | T | G | 0.893313 | 0.000279022 | 32.11883559 |
| | 1 | 66073835 | 0.025957 7 | 0.0043825 1 | 3.20E-09 | rs11417022 | GT | G | 0.390629 | 0.000304752 | 35.08157142 |
| | 1 | 150940625 | 0.033817 1 | 0.0048817 | 4.30E-12 | rs267738 | G | T | 0.219652 | 0.000416814 | 47.98693396 |
| | 2 | 20367135 | -0.026894 4 | 0.0040585 9 | 3.40E-11 | rs11895352 | T | C | 0.475363 | 0.000381417 | 43.91023993 |
| | 2 | 21319016 | 0.05387 | 0.0058209 | 2.20E-20 | rs672889 | G | T | 0.860114 | 0.000743675 | 85.6458111 |
| | 2 | 27730940 | -0.097683 4 | 0.0041328 4 | 1.60E-123 | rs1260326 | C | T | 0.603982 | 0.004830964 | 558.6461405 |

| | | | | | | | | | | | |
|---|---|---|---|---|---|---|---|---|---|---|---|
| | 3 | 119560606 | -0.0231387 | 0.00415568 | 2.60E-08 | rs6805251 | C | T | 0.615535 | 0.0026932 | 31.00174693 |
| | 4 | 39414993 | 0.0254216 | 0.00416565 | 1.00E-09 | rs11940694 | G | A | 0.605104 | 0.00323513 | 37.24197115 |
| | 4 | 3443931 | -0.0334438 | 0.0041863 | 1.40E-15 | rs13108218 | G | A | 0.615355 | 0.00554271 | 63.82090998 |
| | 6 | 31323953 | 0.0383404 | 0.00422753 | 1.20E-19 | rs1131285 | G | C | 0.648353 | 0.00714204 | 82.24932358 |
| | 6 | 34889423 | -0.0687856 | 0.0111785 | 7.60E-10 | rs79834165 | C | T | 0.033984 | 0.0032891 | 37.86347326 |
| | 7 | 73012785 | -0.0665509 | 0.00600844 | 1.60E-28 | rs79624003 | G | A | 0.129851 | 0.01064913 | 122.6808739 |
| | 8 | 9184231 | 0.0982997 | 0.00704622 | 3.10E-44 | rs4240624 | A | G | 0.909234 | 0.01688304 | 194.6186428 |
| | 8 | 11689763 | -0.0259602 | 0.00429267 | 1.50E-09 | rs34120986 | A | G | 0.371642 | 0.00317699 | 36.57240444 |
| | 8 | 116624987 | 0.0231985 | 0.00410632 | 1.60E-08 | rs2737214 | G | C | 0.558518 | 0.00277259 | 31.91586952 |
| | 8 | 126506694 | -0.0598217 | 0.00419928 | 4.80E-46 | rs112875651 | A | G | 0.392379 | 0.01760335 | 202.9365563 |
| | 8 | 19855600 | 0.0287452 | 0.00458644 | 3.70E-10 | rs17411168 | C | T | 0.265208 | 0.00341211 | 39.28001897 |

| | | | | | | | | | | | |
|---|---|---|---|---|---|---|---|---|---|---|---|
| | 9 | 15300968 | 0.0320826 | 0.00488906 | 5.30E-11 | rs4008004 | A | C | 0.221856 | 0.00037404 | 43.06067481 |
| | 9 | 107661742 | -0.0467307 | 0.00459215 | 2.50E-24 | rs2740488 | C | A | 0.265313 | 0.00089903 | 103.5535267 |
| | 9 | 136155000 | 0.0425522 | 0.00524267 | 4.80E-16 | rs635634 | T | C | 0.183543 | 0.000572115 | 65.87664084 |
| | 9 | 107647019 | 0.0575105 | 0.00650895 | 1.00E-18 | rs11789603 | T | C | 0.108822 | 0.000677908 | 78.06652867 |
| | 10 | 113921825 | -0.0252039 | 0.00450458 | 2.20E-08 | rs2792735 | A | G | 0.72034 | 0.000271958 | 31.30540593 |
| | 10 | 5261832 | -0.0407791 | 0.00626679 | 7.70E-11 | rs117488242 | G | A | 0.131657 | 0.000367805 | 42.34259222 |
| | 11 | 75474195 | -0.0648916 | 0.00696074 | 1.10E-20 | rs72997616 | A | C | 0.094104 | 0.000754624 | 86.90776734 |
| | 11 | 116705516 | -0.0904734 | 0.00447068 | 4.60E-91 | rs525028 | A | G | 0.708551 | 0.003546047 | 409.5313045 |
| | 11 | 47391745 | -0.0282135 | 0.00431657 | 6.30E-11 | rs67472071 | A | G | 0.324584 | 0.00037108 | 42.7197603 |
| | 12 | 4384844 | -0.0958104 | 0.0157379 | 1.10E-09 | rs76895963 | G | T | 0.020556 | 0.000321947 | 37.0616027 |
| | 12 | 110013639 | 0.0225944 | 0.00404513 | 2.30E-08 | rs6606734 | G | T | 0.523174 | 0.000271026 | 31.1981694 |

| | | | | | | | | | | | |
|---|---|---|---|---|---|---|---|---|---|---|---|
| | 14 | 24871530 | 0.0287969 | 0.00410511 | 2.30E-12 | rs11620783 | T | C | 0.431558 | 0.00427414 | 49.20784331 |
| | 15 | 58580781 | -0.033055 | 0.00491461 | 1.70E-11 | rs1560390 | C | T | 0.219673 | 0.00392933 | 45.23646455 |
| | 15 | 58737341 | -0.0690999 | 0.00579921 | 9.80E-33 | rs473224 | G | T | 0.853927 | 0.0123218 | 141.9742617 |
| | 15 | 58678720 | -0.0799617 | 0.00426254 | 1.60E-78 | rs261290 | C | T | 0.654645 | 0.03048553 | 351.9002334 |
| | 16 | 67976320 | -0.064083 | 0.0112121 | 1.10E-08 | rs4986970 | T | A | 0.033919 | 0.0028378 | 32.66661971 |
| | 16 | 57049137 | -0.0823934 | 0.0119276 | 4.90E-12 | rs75911530 | A | G | 0.032885 | 0.00414468 | 47.71678436 |
| | 16 | 56993324 | 0.076061 | 0.00432017 | 2.20E-69 | rs3764261 | A | C | 0.324412 | 0.02686252 | 309.966558 |
| | 17 | 7080316 | -0.0391595 | 0.00514349 | 2.70E-14 | rs55714927 | T | C | 0.190994 | 0.00503422 | 57.9630098 |
| | 17 | 41926126 | -0.0657839 | 0.0115818 | 1.30E-08 | rs72836561 | T | C | 0.031482 | 0.00280258 | 32.2611292 |
| | 18 | 47109955 | 0.243656 | 0.0176674 | 2.90E-43 | rs77960347 | G | A | 0.013234 | 0.01650002 | 190.1960045 |
| | 18 | 47158234 | 0.0721067 | 0.00525047 | 6.40E-43 | rs9304381 | T | C | 0.818404 | 0.016362 | 188.6024501 |

| | | | | | | | | | | | |
|---|---|---|---|---|---|---|---|---|---|---|---|
| | 19 | 45413233 | -0.0712435 | 0.00746273 | 1.30E-21 | rs1065853 | T | G | 0.080581 | 0.00791305 | 91.13548056 |
| | 19 | 19379549 | -0.0807326 | 0.00771321 | 1.20E-25 | rs58542926 | T | C | 0.074399 | 0.00951058 | 109.5519266 |
| | 19 | 45440529 | 0.0257907 | 0.00461355 | 2.30E-08 | rs73045691 | A | G | 0.304293 | 0.00271475 | 31.2498724 |
| | 19 | 11330942 | -0.0995971 | 0.0111918 | 5.60E-19 | rs17699030 | G | A | 0.033711 | 0.00687682 | 79.19284721 |
| | 19 | 49248052 | -0.0242633 | 0.00406783 | 2.50E-09 | rs838146 | T | C | 0.500208 | 0.00309053 | 35.57677676 |
| | 20 | 39678289 | 0.0229373 | 0.0040501 | 1.50E-08 | rs6016505 | T | C | 0.549538 | 0.00278628 | 32.07344052 |
| | 20 | 44559748 | 0.109722 | 0.0519862 | 7.00E-99 | rs139953093 | T | G | 0.186317 | 0.03855903 | 445.4548957 |
| | 22 | 21916272 | -0.0368558 | 0.00528804 | 3.20E-12 | rs5754102 | A | C | 0.183262 | 0.00421921 | 48.57520859 |
| exposure | chr.exposure | pos.exposure | beta.exposure | se.exposure | pval.exposure | SNP | effect_allele.exposure | other_allele.exposure | eaf.exposure | R2 | F |
| Free cholesterol levels | 1 | 26987646 | -0.0814865 | 0.0123501 | 4.20E-11 | rs193084249 | G | A | 0.023362 | 0.00378145 | 43.53342463 |
| | 1 | 110470764 | -0.0293502 | 0.00510931 | 9.20E-09 | rs333947 | A | G | 0.150128 | 0.00286659 | 32.99818667 |

| | | | | | | | | | | | |
|---|---|---|---|---|---|---|---|---|---|---|---|
| in HDL \|\| id: ebi-a-GCST90092824 | 1 | 178512331 | -0.0206066 | 0.00375968 | 4.20E-08 | rs12405456 | G | T | 0.386143 | 0.0026097 | 30.04023807 |
| | 1 | 220970028 | 0.037005 | 0.00400295 | 2.40E-20 | rs2642438 | G | A | 0.70385 | 0.00742046 | 85.45804208 |
| | 1 | 230304352 | 0.0444331 | 0.00374937 | 2.10E-32 | rs4846921 | A | G | 0.613232 | 0.01218876 | 140.4394372 |
| | 1 | 62900812 | -0.0271579 | 0.00376131 | 5.20E-13 | rs624700 | C | G | 0.416637 | 0.00452804 | 52.13226147 |
| | 1 | 182171957 | -0.0279964 | 0.00387145 | 4.80E-13 | rs1779823 | T | G | 0.333943 | 0.00454205 | 52.29371826 |
| | 1 | 40035928 | -0.0346647 | 0.00460675 | 5.30E-14 | rs3768321 | T | G | 0.196524 | 0.00491773 | 56.62106781 |
| | 2 | 227092802 | -0.0350052 | 0.00381451 | 4.40E-20 | rs2176040 | G | A | 0.646742 | 0.00731244 | 84.21317928 |
| | 2 | 20371380 | -0.0278482 | 0.00369222 | 4.60E-14 | rs907866 | A | G | 0.443212 | 0.0049408 | 56.88680859 |
| | 2 | 165508389 | 0.0273196 | 0.00372144 | 2.10E-13 | rs10184004 | T | C | 0.406103 | 0.00468075 | 53.8913524 |
| | 2 | 21231524 | 0.0588258 | 0.004507 | 6.20E-39 | rs676210 | A | G | 0.205766 | 0.01478124 | 170.3542686 |
| | 2 | 211377711 | 0.0230631 | 0.00421168 | 4.40E-08 | rs189333486 | T | G | 0.259938 | 0.00260498 | 29.98589968 |

| | | | | | | | | | | | |
|---|---|---|---|---|---|---|---|---|---|---|---|
| | 4 | 69373407 | 0.0283795 | 0.00436895 | 8.30E-11 | rs9884390 | C | T | 0.234037 | 0.000366512 | 42.19371671 |
| | 4 | 103188709 | -0.0715063 | 0.00696906 | 1.10E-24 | rs13107325 | T | C | 0.074282 | 0.00913978 | 105.2767958 |
| | 5 | 75003678 | 0.0267853 | 0.00374151 | 8.10E-13 | rs2307111 | C | T | 0.396595 | 0.00445142 | 51.24973942 |
| | 5 | 55860866 | -0.0318337 | 0.00455415 | 2.70E-12 | rs3936510 | T | G | 0.201952 | 0.00424393 | 48.85983871 |
| | 6 | 31298366 | -0.0284805 | 0.00388355 | 2.20E-13 | rs9265559 | G | C | 0.350847 | 0.00467119 | 53.78112335 |
| | 6 | 161092438 | -0.0460396 | 0.00606941 | 3.30E-14 | rs11751347 | T | C | 0.102257 | 0.00499742 | 57.53903634 |
| | 6 | 127519234 | -0.0235871 | 0.00367728 | 1.40E-10 | rs10457487 | A | C | 0.484321 | 0.00357382 | 41.14226048 |
| | 6 | 34689725 | 0.0258782 | 0.00385799 | 2.00E-11 | rs34265539 | ATT | A | 0.64394 | 0.00390813 | 44.99231993 |
| | 6 | 43764551 | -0.0276815 | 0.00368195 | 5.60E-14 | rs1358980 | T | C | 0.482622 | 0.00490911 | 56.52179154 |
| | 7 | 130437476 | -0.024275 | 0.00406333 | 2.30E-09 | rs7810507 | A | G | 0.281085 | 0.00310036 | 35.6900158 |
| | 7 | 73039406 | 0.0258426 | 0.00455232 | 1.40E-08 | rs35493868 | G | C | 0.203675 | 0.00279948 | 32.22547281 |

| | | | | | | | | | | | |
|---|---|---|---|---|---|---|---|---|---|---|---|
| | 7 | 1066297 | -0.0306837 | 0.00371623 | 1.50E-16 | rs3735687 | G | A | 0.422805 | 0.00592033 | 68.17148247 |
| | 8 | 9184231 | 0.0836687 | 0.00636614 | 1.90E-39 | rs4240624 | A | G | 0.909234 | 0.01498701 | 172.7293444 |
| | 8 | 116624879 | 0.0319446 | 0.00370821 | 7.00E-18 | rs2245221 | A | G | 0.558968 | 0.00644435 | 74.20936573 |
| | 8 | 144305667 | 0.0335832 | 0.005539 | 1.30E-09 | rs79744701 | A | G | 0.12409 | 0.00319327 | 36.75986212 |
| | 8 | 19750044 | -0.0627706 | 0.00899036 | 2.90E-12 | rs59347135 | G | C | 0.045803 | 0.00423416 | 48.74733155 |
| | 8 | 19766233 | -0.121014 | 0.0137878 | 1.70E-18 | rs139915535 | G | A | 0.018003 | 0.00668934 | 77.03245429 |
| | 8 | 19824667 | 0.0843391 | 0.00404376 | 1.30E-96 | rs15285 | T | C | 0.286526 | 0.03765662 | 434.9903604 |
| | 9 | 15304782 | 0.0535283 | 0.0052655 | 2.80E-24 | rs686030 | A | C | 0.859548 | 0.00897202 | 103.3427752 |
| | 9 | 107661742 | -0.0472246 | 0.00415239 | 5.70E-30 | rs2740488 | C | A | 0.265313 | 0.01122652 | 129.3399699 |
| | 9 | 107586753 | 0.0362479 | 0.00543559 | 2.60E-11 | rs2066714 | C | T | 0.12893 | 0.00386275 | 44.46975866 |
| | 9 | 107647019 | 0.055917 | 0.00588563 | 2.10E-21 | rs11789603 | T | C | 0.108822 | 0.00783707 | 90.25977884 |

| | | | | | | | | | | | |
|---|---|---|---|---|---|---|---|---|---|---|---|
| | 10 | 45998984 | 0.0348741 | 0.00439149 | 2.00E-15 | rs148063610 | CAAATAAAT | C | 0.236628 | 0.00547693 | 63.06300671 |
| | 10 | 113921825 | −0.0420859 | 0.00407209 | 4.90E-25 | rs2792735 | A | G | 0.72034 | 0.00927317 | 106.8146825 |
| | 10 | 113983758 | −0.0360127 | 0.00523725 | 6.10E-12 | rs60847460 | T | C | 0.14333 | 0.00410695 | 47.28214907 |
| | 11 | 61592362 | −0.0741511 | 0.00382738 | 1.30E-83 | rs174566 | G | A | 0.348122 | 0.03250951 | 375.3396823 |
| | 11 | 117221862 | −0.0481487 | 0.0085266 | 1.60E-08 | rs141368429 | T | C | 0.056176 | 0.00277006 | 31.88671678 |
| | 11 | 48944029 | 0.0321914 | 0.0052627 | 9.50E-10 | rs12295878 | T | C | 0.140343 | 0.00325023 | 37.41579297 |
| | 11 | 116640237 | 0.0635881 | 0.00727914 | 2.40E-18 | rs6589565 | G | A | 0.932176 | 0.00662668 | 76.3104483 |
| | 11 | 126229617 | −0.0418713 | 0.0069996 | 2.20E-09 | rs73632745 | T | C | 0.073721 | 0.00311165 | 35.82001311 |
| | 11 | 47475189 | −0.0373258 | 0.00381554 | 1.30E-22 | rs35184771 | T | G | 0.352771 | 0.00830879 | 95.69707862 |
| | 11 | 75451281 | −0.0510297 | 0.00502522 | 3.20E-24 | rs559355 | T | A | 0.157979 | 0.0089524 | 103.116538 |
| | 11 | 116727936 | 0.0560751 | 0.00467178 | 3.40E-33 | rs625145 | T | A | 0.188128 | 0.0125033 | 144.0680904 |

| | | | | | | | | | | | |
|---|---|---|---|---|---|---|---|---|---|---|---|
| | 12 | 109871179 | 0.0293081 | 0.00365238 | 1.00E-15 | rs7308864 | G | A | 0.523292 | 0.000559208 | 64.38962115 |
| | 12 | 112083162 | -0.0217289 | 0.00371439 | 4.90E-09 | rs11065991 | T | C | 0.415862 | 0.000297279 | 34.22101857 |
| | 12 | 125265201 | -0.0418948 | 0.0039707 | 5.00E-26 | rs921919 | A | G | 0.66951 | 0.000966404 | 111.3213703 |
| | 12 | 125326153 | -0.0417392 | 0.0045235 | 2.80E-20 | rs7136506 | C | T | 0.215774 | 0.000739283 | 85.13958026 |
| | 12 | 20470199 | 0.0260772 | 0.0046163 | 1.60E-08 | rs11045171 | G | A | 0.197429 | 0.000277208 | 31.90996434 |
| | 12 | 123348440 | 0.0491463 | 0.00790215 | 5.00E-10 | rs142265900 | AGAT | A | 0.057566 | 0.000335999 | 38.67974199 |
| | 12 | 124496316 | 0.0264466 | 0.00395748 | 2.30E-11 | rs1054852 | G | A | 0.377477 | 0.000387906 | 44.65752892 |
| | 12 | 125434716 | 0.0242575 | 0.00427279 | 1.40E-08 | rs4078216 | A | G | 0.24095 | 0.000279988 | 32.23008385 |
| | 15 | 58678720 | -0.148741 | 0.00384624 | 1.00E-200 | rs261290 | C | T | 0.654645 | 0.012828441 | 1495.481642 |
| | 15 | 58737341 | -0.148425 | 0.00523283 | 5.60E-177 | rs473224 | G | T | 0.853927 | 0.006942373 | 804.5134621 |
| | 15 | 58577163 | -0.0648721 | 0.00450452 | 5.10E-47 | rs10162642 | A | G | 0.210107 | 0.001798993 | 207.4012297 |

| | | | | | | | | | | | |
|---|---|---|---|---|---|---|---|---|---|---|---|
| | 16 | 15129459 | −0.0266089 | 0.00385552 | 5.10E-12 | rs4985155 | G | A | 0.334889 | 0.0004137115 | 47.6299773 |
| | 16 | 57049137 | −0.142841 | 0.0107424 | 2.40E-40 | rs75911530 | A | G | 0.032885 | 0.001534013 | 176.8054212 |
| | 16 | 56752882 | −0.077622 | 0.012601 | 7.30E-10 | rs75714888 | A | C | 0.021624 | 0.00329617 | 37.94478412 |
| | 16 | 81534790 | 0.0326831 | 0.00396826 | 1.80E-16 | rs2925979 | C | T | 0.698949 | 0.00589092 | 67.83263677 |
| | 16 | 67976320 | −0.08649 | 0.010098 | 1.10E-17 | rs4986970 | T | A | 0.033919 | 0.00637055 | 73.3590188 |
| | 16 | 56993324 | 0.196289 | 0.00389089 | 1.00E-200 | rs3764261 | A | C | 0.324412 | 0.02163693 | 2544.992293 |
| | 16 | 57151796 | −0.0256572 | 0.00408883 | 3.50E-10 | rs8058512 | T | C | 0.724045 | 0.0034203 | 39.37430152 |
| | 17 | 41926126 | −0.138482 | 0.0104711 | 6.30E-40 | rs72836561 | T | C | 0.031482 | 0.01517522 | 174.9018859 |
| | 17 | 26695832 | −0.0204958 | 0.00372243 | 3.70E-08 | rs2071379 | G | A | 0.596242 | 0.00263363 | 30.31580621 |
| | 17 | 66823805 | −0.0468842 | 0.00786385 | 2.60E-09 | rs112001035 | A | G | 0.05973 | 0.00308219 | 35.48075642 |
| | 18 | 47120499 | 0.0764042 | 0.00915669 | 7.20E-17 | rs12608026 | G | T | 0.042399 | 0.00604627 | 69.62254509 |

| | | | | | | | | | | | |
|---|---|---|---|---|---|---|---|---|---|---|---|
| | 18 | 47160953 | 0.077373 | 0.00480175 | 2.10E-58 | rs7241918 | T | G | 0.8229 | 0.002251094 | 259.6403791 |
| | 18 | 57867526 | −0.0226316 | 0.00414924 | 4.90E-08 | rs688671 | G | A | 0.267133 | 0.000258448 | 29.7499238 |
| | 18 | 47109955 | 0.264028 | 0.0159719 | 2.20E-61 | rs77960347 | G | A | 0.013234 | 0.002368914 | 273.2620106 |
| | 19 | 45413576 | −0.0375439 | 0.00452384 | 1.00E-16 | rs75627662 | T | C | 0.20606 | 0.000598132 | 68.87422874 |
| | 19 | 54813266 | 0.0215071 | 0.00391349 | 3.90E-08 | rs431001 | A | G | 0.325865 | 0.00026237 | 30.20144374 |
| | 19 | 8429323 | 0.176342 | 0.0131196 | 3.50E-41 | rs116843064 | A | G | 0.019851 | 0.001567407 | 180.6603254 |
| | 19 | 45448465 | 0.0413167 | 0.00382423 | 3.30E-27 | rs5167 | G | T | 0.352266 | 0.001013248 | 116.7228002 |
| | 19 | 11347493 | −0.0605343 | 0.00686439 | 1.20E-18 | rs737337 | C | T | 0.076789 | 0.000675303 | 77.76632964 |
| | 20 | 43008981 | −0.0809626 | 0.0111353 | 3.60E-13 | rs150224153 | T | C | 0.029237 | 0.000459154 | 52.86369326 |
| | 20 | 44551855 | −0.0918399 | 0.00458895 | 4.20E-89 | rs6073958 | C | T | 0.198731 | 0.003468326 | 400.5240557 |
| | 20 | 46340974 | 0.0355302 | 0.00452416 | 4.00E-15 | rs6018652 | A | G | 0.792529 | 0.000535648 | 61.67538162 |

| | | | | | | | | | | | |
|---|---|---|---|---|---|---|---|---|---|---|---|
| | 22 | 21917479 | −0.0348674 | 0.00466685 | 7.90E-14 | rs5998509 | T | C | 0.189556 | 0.00484812 | 55.81923703 |
| exposure | chr.exposure | pos.exposure | beta.exposure | se.exposure | pval.exposure | SNP | effect_allele.exposure | other_allele.exposure | eaf.exposure | R2 | F |
| Total lipid levels in very large HDL \|\| id:ebi-a-GCST90093010 | 1 | 109817590 | −0.0341041 | 0.00446359 | 2.20E-14 | rs12740374 | T | G | 0.221064 | 0.00507001 | 58.37636473 |
| | 1 | 161193683 | −0.0423312 | 0.00383258 | 2.30E-28 | rs5082 | A | G | 0.620747 | 0.01058944 | 121.9919447 |
| | 1 | 178526271 | −0.022289 | 0.00372178 | 2.10E-09 | rs6681348 | C | A | 0.532241 | 0.00311557 | 35.8651148 |
| | 1 | 182119332 | −0.0299395 | 0.00397163 | 4.80E-14 | rs6694102 | A | G | 0.320921 | 0.00493548 | 56.82559264 |
| | 1 | 230297659 | 0.0295629 | 0.00380788 | 8.30E-15 | rs2281719 | T | C | 0.612353 | 0.00523471 | 60.27261212 |
| | 1 | 220970028 | 0.0267361 | 0.00406352 | 4.70E-11 | rs2642438 | G | A | 0.70385 | 0.00376028 | 43.28961895 |
| | 1 | 55505647 | −0.107163 | 0.0141363 | 3.40E-14 | rs11591147 | T | G | 0.017457 | 0.00499107 | 57.46596078 |
| | 2 | 44055510 | −0.0436827 | 0.00777823 | 2.00E-08 | rs114938914 | T | C | 0.061412 | 0.00273988 | 31.53913938 |
| | 2 | 27730940 | 0.0531173 | 0.00379123 | 1.30E-44 | rs1260326 | C | T | 0.603982 | 0.01702802 | 196.2927098 |

| | | | | | | | | | | | |
|---|---|---|---|---|---|---|---|---|---|---|---|
| | 2 | 165528876 | 0.0339588 | 0.00379952 | 4.00E-19 | rs13389219 | T | C | 0.392532 | 0.00693647 | 79.8802937 |
| | 2 | 242237902 | 0.0244836 | 0.00386765 | 2.40E-10 | rs59104589 | T | C | 0.358836 | 0.00348095 | 40.07270613 |
| | 2 | 227112754 | -0.0359695 | 0.0038684 | 1.40E-20 | rs2943654 | T | C | 0.64513 | 0.00750711 | 86.45667017 |
| | 2 | 21271707 | -0.0505768 | 0.00544975 | 1.70E-20 | rs34722314 | A | T | 0.136126 | 0.00747855 | 86.12752916 |
| | 3 | 12206489 | -0.105194 | 0.0153239 | 6.70E-12 | rs11922042 | C | T | 0.017207 | 0.00409315 | 47.12330833 |
| | 3 | 98416900 | -0.0240551 | 0.00373389 | 1.20E-10 | rs10935473 | T | G | 0.442432 | 0.00360518 | 41.50340675 |
| | 4 | 103188709 | -0.052796 | 0.00707617 | 8.60E-14 | rs13107325 | T | C | 0.074282 | 0.00483491 | 55.66702048 |
| | 4 | 3443931 | 0.0256029 | 0.00384228 | 2.70E-11 | rs13108218 | G | A | 0.615355 | 0.00385678 | 44.40100005 |
| | 4 | 100066125 | 0.0275358 | 0.00419088 | 5.00E-11 | rs4699716 | A | G | 0.271935 | 0.00374986 | 43.16953775 |
| | 4 | 89704367 | -0.0217413 | 0.00381373 | 1.20E-08 | rs13118477 | A | G | 0.390372 | 0.0028232 | 32.49852001 |
| | 5 | 55857025 | -0.0434084 | 0.00473221 | 4.60E-20 | rs11429307 | GT | G | 0.191517 | 0.00730625 | 84.14183736 |

| | | | | | | | | | | | |
|---|---|---|---|---|---|---|---|---|---|---|---|
| | 5 | 74967708 | -0.031669 | 0.00390003 | 4.70E-16 | rs34360 | A | C | 0.652339 | 0.000572633 | 65.93640382 |
| | 6 | 31320309 | -0.0257964 | 0.00412638 | 4.10E-10 | rs2507993 | T | C | 0.48033 | 0.000339488 | 39.08158941 |
| | 6 | 161005389 | -0.0516925 | 0.00604191 | 1.20E-17 | rs191555775 | T | A | 0.106389 | 0.000635657 | 73.19796743 |
| | 6 | 98583488 | 0.0256906 | 0.00380679 | 1.50E-11 | rs72934503 | G | A | 0.451389 | 0.000395596 | 45.5431544 |
| | 6 | 127521127 | -0.0243786 | 0.00375851 | 8.80E-11 | rs4318902 | G | A | 0.556738 | 0.000365443 | 42.07058542 |
| | 6 | 32564898 | -0.0256245 | 0.00461479 | 2.80E-08 | rs28366346 | G | A | 0.272121 | 0.000267845 | 30.83183453 |
| | 6 | 34354073 | -0.0518496 | 0.00894274 | 6.70E-09 | rs115447786 | T | C | 0.04509 | 0.000292022 | 33.61569266 |
| | 6 | 43757896 | -0.0323856 | 0.00371842 | 3.10E-18 | rs998584 | A | C | 0.482313 | 0.000658709 | 75.85417039 |
| | 6 | 139839768 | 0.0204226 | 0.00372456 | 4.20E-08 | rs151288714 | AAAAC | A | 0.51752 | 0.000261187 | 30.0652324 |
| | 7 | 72879234 | -0.0395605 | 0.00674888 | 4.60E-09 | rs111432585 | GT | G | 0.907696 | 0.000298485 | 34.35995481 |
| | 7 | 1029585 | -0.0365172 | 0.00382464 | 1.30E-21 | rs9769088 | C | T | 0.390936 | 0.000791521 | 91.16040941 |

| | | | | | | | | | | | |
|---|---|---|---|---|---|---|---|---|---|---|---|
| | 7 | 73033559 | 0.080317 | 0.00551684 | 5.20E-48 | rs34062580 | A | G | 0.130304 | 0.001838348 | 211.9466867 |
| | 7 | 130455588 | -0.0245021 | 0.00398878 | 8.10E-10 | rs1838931 | T | C | 0.320578 | 0.000327776 | 37.73278849 |
| | 7 | 94953895 | -0.0283984 | 0.00375158 | 3.70E-14 | rs705379 | A | G | 0.477476 | 0.000497663 | 57.29962407 |
| | 7 | 28224053 | 0.0254888 | 0.0039047 | 6.70E-11 | rs849336 | G | A | 0.655418 | 0.000370131 | 42.61042704 |
| | 7 | 98005617 | -0.0239661 | 0.00434216 | 3.40E-08 | rs6465679 | T | A | 0.756109 | 0.000264643 | 30.46320142 |
| | 8 | 126506694 | 0.0296936 | 0.00385448 | 1.30E-14 | rs112875651 | A | G | 0.392379 | 0.000515422 | 59.34533244 |
| | 8 | 19820933 | 0.0968537 | 0.00616756 | 1.40E-55 | rs145391587 | C | A | 0.100287 | 0.0021383 | 246.6028654 |
| | 8 | 11756600 | 0.0288392 | 0.0038274 | 4.90E-14 | rs4841605 | A | G | 0.606733 | 0.000493102 | 56.77421853 |
| | 8 | 19733244 | 0.0373685 | 0.00452914 | 1.60E-16 | rs1561748 | C | G | 0.267445 | 0.000591174 | 68.07259274 |
| | 9 | 107661742 | -0.0311528 | 0.00421673 | 1.50E-13 | rs2740488 | C | A | 0.265313 | 0.000474056 | 54.58018945 |
| | 9 | 15303583 | 0.0444415 | 0.00540613 | 2.00E-16 | rs1215112 | A | G | 0.86303 | 0.00058687 | 67.57668765 |

| | | | | | | | | | | | |
|---|---|---|---|---|---|---|---|---|---|---|---|
| | 9 | 136155000 | -0.027738 | 0.00481407 | 8.30E-09 | rs635634 | T | C | 0.183543 | 0.00288398 | 33.19845999 |
| | 10 | 94839724 | 0.0257744 | 0.00373185 | 5.00E-12 | rs4418728 | T | G | 0.450703 | 0.00414325 | 47.7003123 |
| | 10 | 113934384 | -0.0322601 | 0.00415326 | 8.00E-15 | rs2803619 | C | G | 0.724355 | 0.00523984 | 60.33170962 |
| | 10 | 45938301 | 0.0246977 | 0.00399984 | 6.60E-10 | rs2242334 | T | G | 0.683532 | 0.00331189 | 38.12591155 |
| | 10 | 65191645 | 0.0233624 | 0.0037124 | 3.10E-10 | rs7924036 | T | G | 0.504241 | 0.00344008 | 39.60206527 |
| | 11 | 61586328 | -0.0939693 | 0.00390511 | 6.10E-128 | rs35473591 | CT | C | 0.34192 | 0.05006317 | 579.0257695 |
| | 11 | 117156893 | 0.204021 | 0.0188736 | 3.10E-27 | rs1047964 | T | G | 0.010073 | 0.0101436 | 116.8510726 |
| | 11 | 10370634 | -0.0210582 | 0.00374233 | 1.80E-08 | rs1580005 | C | T | 0.477839 | 0.00275063 | 31.66290678 |
| | 11 | 47316568 | 0.0344639 | 0.00435475 | 2.50E-15 | rs113012334 | GT | G | 0.289459 | 0.0054395 | 62.63179227 |
| | 11 | 64004643 | -0.0534309 | 0.0088722 | 1.70E-09 | rs12294913 | G | C | 0.049058 | 0.00315049 | 36.26726328 |
| | 11 | 116648917 | 0.0890027 | 0.00546931 | 1.50E-59 | rs964184 | C | G | 0.86726 | 0.02295809 | 264.8096065 |

| | | | | | | | | | | | |
|---|---|---|---|---|---|---|---|---|---|---|---|
| | 11 | 118957388 | −0.0228366 | 0.00391234 | 5.30E-09 | rs141949189 | T | A | 0.431973 | 0.000295974 | 34.07078677 |
| | 12 | 20582651 | 0.0273422 | 0.00467393 | 4.90E-09 | rs12369443 | G | A | 0.199473 | 0.00029728 | 34.22117807 |
| | 12 | 123209295 | 0.0307209 | 0.00457502 | 1.90E-11 | rs3922628 | A | T | 0.210496 | 0.00039165 | 645.08938666 |
| | 12 | 124620185 | 0.0248727 | 0.00388162 | 1.50E-10 | rs10846606 | G | C | 0.366512 | 0.00035666 | 341.05937005 |
| | 12 | 57848708 | 0.0265328 | 0.00410195 | 9.90E-11 | rs3809113 | G | C | 0.710445 | 0.00036343 | 41.83867233 |
| | 12 | 125265201 | −0.0384262 | 0.0040323 | 1.60E-21 | rs921919 | A | G | 0.66951 | 0.00078849 | 690.81166919 |
| | 12 | 125349480 | 0.0369716 | 0.00388133 | 1.60E-21 | rs36226283 | A | G | 0.358958 | 0.00078781 | 890.73352961 |
| | 15 | 41053250 | 0.0205872 | 0.00375347 | 4.10E-08 | rs6492964 | A | G | 0.560902 | 0.00026134 | 130.08299846 |
| | 15 | 58678720 | −0.166925 | 0.00390274 | 1.00E-200 | rs261290 | C | T | 0.654645 | 0.01564756 | 41829.346539 |
| | 15 | 58833993 | 0.0814449 | 0.011076 | 1.90E-13 | rs6078 | A | G | 0.028853 | 0.00046962 | 454.06974819 |
| | 15 | 58568394 | −0.038356 | 0.00378525 | 3.90E-24 | rs77732800 | G | T | 0.483171 | 0.00089142 | 1102.67623 52 |

| | | | | | | | | | | | |
|---|---|---|---|---|---|---|---|---|---|---|---|
| | 15 | 58725839 | 0.133717 | 0.00408605 | 1.00E-200 | rs633695 | G | A | 0.292354 | 0.009220101 | 1070.923251 |
| | 16 | 15130594 | -0.0317858 | 0.0040322 | 3.20E-15 | rs6498540 | G | A | 0.302064 | 0.000539685 | 62.14048158 |
| | 16 | 56852822 | 0.0343984 | 0.00374816 | 4.40E-20 | rs1561139 | T | G | 0.42589 | 0.000731333 | 84.22338535 |
| | 16 | 67506247 | 0.0700739 | 0.00997095 | 2.10E-12 | rs8044652 | C | T | 0.962945 | 0.000428989 | 49.38919568 |
| | 16 | 57065556 | -0.0282578 | 0.00405187 | 3.10E-12 | rs289754 | T | C | 0.30631 | 0.000422449 | 48.63603254 |
| | 16 | 56993324 | 0.176023 | 0.00395519 | 1.00E-200 | rs3764261 | A | C | 0.324412 | 0.016919435 | 1980.599145 |
| | 16 | 81534790 | 0.0316963 | 0.00403383 | 3.90E-15 | rs2925979 | C | T | 0.698949 | 0.000536218 | 61.74110626 |
| | 17 | 66457658 | 0.0287309 | 0.00447081 | 1.30E-10 | rs2909211 | T | C | 0.778897 | 0.000358726 | 41.29699833 |
| | 17 | 7080316 | 0.0316852 | 0.00472257 | 2.00E-11 | rs55714927 | T | C | 0.190994 | 0.000391002 | 45.01409779 |
| | 17 | 41926126 | -0.0979738 | 0.0010634 | 3.20E-20 | rs72836561 | T | C | 0.031482 | 0.000737053 | 84.88267278 |
| | 18 | 47109955 | 0.125395 | 0.0162309 | 1.10E-14 | rs77960347 | G | A | 0.013234 | 0.000518373 | 59.68534216 |

| | | | | | | | | | | | |
|---|---|---|---|---|---|---|---|---|---|---|---|
| | 18 | 47160953 | 0.0300309 | 0.00487963 | 7.50E-10 | rs7241918 | T | G | 0.8229 | 0.000329013 | 37.87523731 |
| | 19 | 45413233 | -0.172329 | 0.00685056 | 1.20E-139 | rs1065853 | T | G | 0.080581 | 0.005468594 | 632.7862582 |
| | 19 | 8429323 | 0.150339 | 0.01332 | 1.50E-29 | rs116843064 | A | G | 0.019851 | 0.001105725 | 127.3876474 |
| | 19 | 11207777 | -0.0376504 | 0.00560787 | 1.90E-11 | rs117423069 | T | C | 0.126605 | 0.000391531 | 45.07500324 |
| | 19 | 45440529 | 0.0240118 | 0.00423509 | 1.40E-08 | rs73045691 | A | G | 0.304293 | 0.000279251 | 32.14523456 |
| | 20 | 44563007 | -0.21718 | 0.0047702 | 1.00E-200 | rs1057208 | T | C | 0.186277 | 0.017693194 | 2072.807321 |
| | 20 | 44055710 | 0.0275427 | 0.00373983 | 1.80E-13 | rs3092498 | C | T | 0.517382 | 0.000471083 | 54.23776107 |
| | 20 | 44113286 | -0.0718587 | 0.0088506 | 4.70E-16 | rs76070947 | G | C | 0.046949 | 0.000572476 | 65.91826041 |
| | 20 | 17844684 | -0.0213801 | 0.00391796 | 4.80E-08 | rs2618566 | T | G | 0.660294 | 0.00025869 | 29.77775286 |
| | 20 | 44776543 | -0.0702371 | 0.0102984 | 9.10E-12 | rs2024570 | C | T | 0.962637 | 0.000404027 | 46.51425651 |
| exposure | chr.exposure | pos.exposure | beta.exposure | se.exposure | pval.exposure | SNP | effect_allele.exposure | other_allele.exposure | eaf.exposure | R2 | F |

| | | | | | | | | | | | |
|---|---|---|---|---|---|---|---|---|---|---|---|
| Total lipid levels in large HDL \|\| id:ebi-a-GCST90092850 | 1 | 27180088 | -0.0847704 | 0.0121644 | 3.20E-12 | rs75460349 | C | A | 0.023255 | 0.0042181 | 48.56233183 |
| | 1 | 161193683 | -0.0216878 | 0.00373823 | 6.60E-09 | rs5082 | A | G | 0.620747 | 0.00292391 | 33.65824092 |
| | 1 | 220970028 | 0.0323664 | 0.00396349 | 3.20E-16 | rs2642438 | G | A | 0.70385 | 0.00579128 | 66.68462494 |
| | 1 | 40083641 | -0.0303059 | 0.00439748 | 5.50E-12 | rs61781392 | A | C | 0.216719 | 0.00412534 | 47.49403612 |
| | 1 | 178630827 | 0.0229599 | 0.00381018 | 1.70E-09 | rs58388121 | TAA | T | 0.371192 | 0.00315431 | 36.31127099 |
| | 1 | 230303848 | 0.0450802 | 0.00371169 | 6.10E-34 | rs1546954 | G | T | 0.612656 | 0.01280161 | 147.50975 |
| | 1 | 110470764 | -0.0291523 | 0.00505894 | 8.30E-09 | rs333947 | A | G | 0.150128 | 0.00288466 | 33.20618967 |
| | 1 | 182119332 | -0.0329644 | 0.00387386 | 1.70E-17 | rs6694102 | A | G | 0.320921 | 0.00628814 | 72.40940191 |
| | 2 | 165528876 | 0.0355411 | 0.0037068 | 9.00E-22 | rs13389219 | T | C | 0.392532 | 0.00798195 | 91.92970658 |
| | 2 | 227095876 | -0.0392772 | 0.00377564 | 2.40E-25 | rs1024137 | T | G | 0.646501 | 0.00939473 | 108.216214 |
| | 2 | 242237902 | 0.0244843 | 0.00377328 | 8.60E-11 | rs59104589 | T | C | 0.358836 | 0.00365739 | 42.10462167 |

| | | | | | | | | | | | |
|---|---|---|---|---|---|---|---|---|---|---|---|
| | 2 | 21231524 | 0.0594639 | 0.00446229 | 1.60E-40 | rs676210 | A | G | 0.205766 | 0.00154068 6 | 177.5757464 |
| | 2 | 219699999 | −0.0564707 | 0.00936896 | 1.70E-09 | rs78058190 | A | G | 0.050108 | 0.000315587 | 36.32921596 |
| | 2 | 20363666 | −0.0214195 | 0.00363668 | 3.90E-09 | rs35135293 | T | C | 0.516755 | 0.000301349 | 34.68973112 |
| | 2 | 27730940 | 0.0256137 | 0.00369872 | 4.40E-12 | rs1260326 | C | T | 0.603982 | 0.000416537 | 47.95503123 |
| | 3 | 185804583 | −0.0301032 | 0.00529138 | 1.30E-08 | rs57912727 | C | A | 0.136685 | 0.000281163 | 32.36531792 |
| | 3 | 12206489 | −0.0851654 | 0.0149476 | 1.20E-08 | rs11922042 | C | T | 0.017207 | 0.000282003 | 32.46204671 |
| | 4 | 69482383 | 0.0275884 | 0.00500046 | 3.40E-08 | rs368166328 | CT | C | 0.240065 | 0.00026443 | 30.43866251 |
| | 4 | 970112 | 0.0213176 | 0.00366314 | 5.90E-09 | rs12510382 | G | A | 0.459916 | 0.000294194 | 33.86585433 |
| | 4 | 89704367 | −0.0206633 | 0.00371979 | 2.80E-08 | rs13118477 | A | G | 0.390372 | 0.000268064 | 30.85709015 |
| | 4 | 100045141 | −0.0242172 | 0.00394198 | 8.10E-10 | rs1139490 | T | C | 0.696979 | 0.000327845 | 37.74083254 |
| | 4 | 103188709 | −0.0689495 | 0.00690187 | 1.70E-23 | rs13107325 | T | C | 0.074282 | 0.000866453 | 99.79784075 |

| | | | | | | | | | | | |
|---|---|---|---|---|---|---|---|---|---|---|---|
| | 5 | 55857025 | -0.0449221 | 0.0046154 | 2.20E-22 | rs11429307 | GT | G | 0.191517 | 0.000822501 | 94.73137947 |
| | 5 | 75003678 | 0.0244178 | 0.00370526 | 4.40E-11 | rs2307111 | C | T | 0.396595 | 0.000377228 | 43.42783116 |
| | 6 | 43757896 | -0.0345011 | 0.00362671 | 1.90E-21 | rs998584 | A | C | 0.482313 | 0.000785763 | 90.49668622 |
| | 6 | 34335088 | -0.0608089 | 0.00887135 | 7.20E-12 | rs147627829 | A | G | 0.04407 | 0.000408103 | 46.98371133 |
| | 6 | 127519234 | -0.0288313 | 0.00364156 | 2.40E-15 | rs10457487 | A | C | 0.484321 | 0.000544389 | 62.68244996 |
| | 6 | 98583488 | 0.0241374 | 0.0037129 | 8.00E-11 | rs72934503 | G | A | 0.451389 | 0.000367103 | 42.26169608 |
| | 6 | 31287454 | -0.0285779 | 0.00394556 | 4.40E-13 | rs9265113 | T | C | 0.407634 | 0.000455657 | 52.46090432 |
| | 6 | 161092438 | -0.0526888 | 0.00601046 | 1.80E-18 | rs11751347 | T | C | 0.102257 | 0.000667304 | 76.84465143 |
| | 7 | 1066297 | -0.0333688 | 0.00368059 | 1.20E-19 | rs3735687 | G | A | 0.422805 | 0.000713722 | 82.19374082 |
| | 7 | 28224053 | 0.0211864 | 0.00380784 | 2.60E-08 | rs849336 | G | A | 0.655418 | 0.000268926 | 30.95632547 |
| | 7 | 73037956 | 0.060429 | 0.00530554 | 4.70E-30 | rs34060476 | G | A | 0.13482 | 0.001125991 | 129.72512 |

| | | | | | | | | | | | |
|---|---|---|---|---|---|---|---|---|---|---|---|
| | 7 | 73123473 | －0.0532962 | 0.00840344 | 2.30E-10 | rs72555385 | G | A | 0.049304 | 0.00349397 | 40.22265513 |
| | 7 | 94953895 | －0.0250874 | 0.00365852 | 7.00E-12 | rs705379 | A | G | 0.477476 | 0.00408428 | 47.02111184 |
| | 7 | 130437476 | －0.0310732 | 0.00402436 | 1.20E-14 | rs7810507 | A | G | 0.281085 | 0.00517781 | 59.61708891 |
| | 8 | 144306597 | 0.0282139 | 0.00469691 | 1.90E-09 | rs11434143 | GT | G | 0.189177 | 0.00313443 | 36.08230064 |
| | 8 | 9173358 | 0.0340209 | 0.00364956 | 1.10E-20 | rs7012814 | A | G | 0.474265 | 0.00754528 | 86.89664983 |
| | 8 | 126506694 | 0.0304119 | 0.00375684 | 5.70E-16 | rs112875651 | A | G | 0.392379 | 0.00569098 | 65.52909368 |
| | 8 | 9239958 | －0.0346922 | 0.00611202 | 4.40E-08 | rs11786900 | C | G | 0.096383 | 0.00260496 | 29.98565794 |
| | 8 | 19733244 | 0.0511052 | 0.0044144 | 5.40E-31 | rs1561748 | C | G | 0.267445 | 0.01163252 | 134.0229801 |
| | 8 | 19980272 | 0.0401357 | 0.00712259 | 1.80E-08 | rs71205961 | CTTTTTT | C | 0.911027 | 0.00275841 | 31.7525234 |
| | 8 | 19820933 | 0.13489 | 0.00601132 | 1.60E-111 | rs145391587 | C | A | 0.100287 | 0.00435628 | 503.5148308 |
| | 8 | 116610180 | 0.0284256 | 0.00366438 | 8.70E-15 | rs2737205 | C | T | 0.561358 | 0.00522617 | 60.1742554 |

| | | | | | | | | | | | |
|---|---|---|---|---|---|---|---|---|---|---|---|
| | 9 | 107661742 | －0.0378462 | 0.00411292 | 3.50E-20 | rs2740488 | C | A | 0.265313 | 0.00073522 | 84.67135078 |
| | 9 | 15303583 | 0.0495441 | 0.00527304 | 5.70E-21 | rs1215112 | A | G | 0.86303 | 0.00766516 | 88.27835015 |
| | 9 | 107647019 | 0.0423026 | 0.00582969 | 4.00E-13 | rs11789603 | T | C | 0.108822 | 0.00457338 | 52.65451918 |
| | 9 | 107586753 | 0.031415 | 0.00538392 | 5.40E-09 | rs2066714 | C | T | 0.12893 | 0.00295761 | 34.04625788 |
| | 10 | 113934384 | －0.0386631 | 0.00405021 | 1.30E-21 | rs2803619 | C | G | 0.724355 | 0.00791202 | 91.12356761 |
| | 10 | 94839724 | 0.0251094 | 0.00363926 | 5.20E-12 | rs4418728 | T | G | 0.450703 | 0.00413485 | 47.60350764 |
| | 10 | 45998984 | 0.0324886 | 0.00434916 | 8.00E-14 | rs148063610 | CAAATAAAT | C | 0.236628 | 0.00484656 | 55.80121772 |
| | 10 | 113983758 | －0.0339356 | 0.00518677 | 6.00E-11 | rs60847460 | T | C | 0.14333 | 0.00371833 | 42.80648189 |
| | 10 | 65191645 | 0.0200906 | 0.00362029 | 2.90E-08 | rs7924036 | T | G | 0.504241 | 0.00267532 | 30.79580387 |
| | 11 | 64004643 | －0.0493802 | 0.00865309 | 1.20E-08 | rs12294913 | G | C | 0.049058 | 0.002829 | 32.56534623 |
| | 11 | 117156893 | 0.216789 | 0.0184075 | 5.10E-32 | rs1047964 | T | G | 0.010073 | 0.0012038 | 138.7002737 |

| | | | | | | | | | | | |
|---|---|---|---|---|---|---|---|---|---|---|---|
| | 11 | 61603510 | -0.0774636 | 0.00378754 | 5.70E-93 | rs174576 | A | C | 0.350043 | 0.003621578 | 418.286019 |
| | 11 | 118957388 | -0.0246513 | 0.00381572 | 1.00E-10 | rs141949189 | T | A | 0.431973 | 0.00362545 | 41.73679628 |
| | 11 | 75531695 | -0.0447629 | 0.0059052 | 3.40E-14 | rs10899133 | T | C | 0.105503 | 0.00499049 | 57.45921123 |
| | 11 | 47322007 | -0.0416472 | 0.00443255 | 5.70E-21 | rs200644264 | CG | C | 0.788465 | 0.00766521 | 88.27887356 |
| | 11 | 47362465 | 0.0379848 | 0.00464735 | 3.00E-16 | rs10769254 | C | G | 0.18914 | 0.00580162 | 66.80383246 |
| | 11 | 116648917 | 0.0896966 | 0.00533424 | 1.90E-63 | rs964184 | C | G | 0.86726 | 0.02450946 | 282.74785 |
| | 11 | 116916060 | -0.134387 | 0.01832 | 2.20E-13 | rs144018203 | C | G | 0.010653 | 0.00467362 | 53.80913761 |
| | 12 | 124637888 | 0.0231095 | 0.00371045 | 4.70E-10 | rs11057469 | A | G | 0.423885 | 0.00336957 | 38.79005392 |
| | 12 | 125265201 | -0.0433452 | 0.00393192 | 2.90E-28 | rs921919 | A | G | 0.66951 | 0.01054889 | 121.52548598 |
| | 12 | 20470199 | 0.0280322 | 0.00457122 | 8.70E-10 | rs11045171 | G | A | 0.197429 | 0.00326664 | 37.60473842 |
| | 12 | 57843711 | 0.0271406 | 0.00422005 | 1.30E-10 | rs2229357 | A | G | 0.240937 | 0.00359286 | 41.3614874 |

| | | | | | | | | | | | |
|---|---|---|---|---|---|---|---|---|---|---|---|
| | 12 | 109871179 | 0.0263177 | 0.00361671 | 3.40E-13 | rs7308864 | G | A | 0.523292 | 0.00459898 | 52.94938772 |
| | 12 | 123348440 | 0.0513008 | 0.00782498 | 5.50E-11 | rs142265900 | AGAT | A | 0.057566 | 0.00373347 | 42.98077631 |
| | 12 | 124496316 | 0.0305995 | 0.00391884 | 5.80E-15 | rs1054852 | G | A | 0.377477 | 0.00529512 | 60.96857549 |
| | 12 | 125434716 | 0.025638 | 0.00423107 | 1.40E-09 | rs4078216 | A | G | 0.24095 | 0.00318949 | 36.71641904 |
| | 12 | 125326153 | -0.0447101 | 0.00447933 | 1.80E-23 | rs7136506 | C | T | 0.215774 | 0.00864972 | 99.62712943 |
| | 15 | 43820717 | -0.0675826 | 0.0113714 | 2.80E-09 | rs55707100 | T | C | 0.025877 | 0.00306832 | 35.32111223 |
| | 15 | 40986355 | 0.0208239 | 0.00365876 | 1.30E-08 | rs5030789 | G | A | 0.567326 | 0.00281402 | 32.39281917 |
| | 15 | 58580781 | -0.0672214 | 0.00439016 | 6.40E-53 | rs1560390 | C | T | 0.219673 | 0.02033121 | 234.4481779 |
| | 15 | 58737661 | -0.155853 | 0.00515909 | 1.80E-200 | rs488490 | A | C | 0.852654 | 0.07867673 | 912.5918308 |
| | 15 | 59058720 | -0.0501538 | 0.00816293 | 8.00E-10 | rs77516617 | A | G | 0.052809 | 0.00327918 | 37.74922113 |
| | 15 | 58678512 | 0.162205 | 0.00396318 | 1.00E-200 | rs10468017 | T | C | 0.295937 | 0.01434684 | 1675.071431 |

| | | | | | | | | | | | |
|---|---|---|---|---|---|---|---|---|---|---|---|
| | 16 | 56993324 | 0.210122 | 0.00384938 | 1.00E-200 | rs3764261 | A | C | 0.324412 | 0.025237872 | 2979.572334 |
| | 16 | 57049137 | -0.143131 | 0.0106278 | 2.40E-41 | rs75911530 | A | G | 0.032885 | 0.001573582 | 181.3732008 |
| | 16 | 81534790 | 0.0360196 | 0.00392592 | 4.50E-20 | rs2925979 | C | T | 0.698949 | 0.0073092 | 84.17581549 |
| | 16 | 57151796 | -0.0246446 | 0.00404521 | 1.10E-09 | rs8058512 | T | C | 0.724045 | 0.00322414 | 37.11537521 |
| | 16 | 56752882 | -0.0815539 | 0.0124665 | 6.10E-11 | rs75714888 | A | C | 0.021624 | 0.00371733 | 42.79498104 |
| | 16 | 56915357 | 0.0730158 | 0.0131597 | 2.90E-08 | rs55779455 | C | G | 0.019854 | 0.00267435 | 30.78464647 |
| | 16 | 15130594 | -0.0297339 | 0.00392434 | 3.50E-14 | rs6498540 | G | A | 0.302064 | 0.00498593 | 57.40674876 |
| | 16 | 67976320 | -0.0848873 | 0.00999024 | 1.90E-17 | rs4986970 | T | A | 0.033919 | 0.0062698 | 72.19814672 |
| | 17 | 37808759 | -0.0389168 | 0.00670072 | 6.30E-09 | rs34695955 | G | T | 0.079539 | 0.0029302 | 33.73057589 |
| | 17 | 41926126 | -0.138256 | 0.0103721 | 1.60E-40 | rs72836561 | T | C | 0.031482 | 0.01541548 | 177.6752918 |
| | 17 | 66823805 | -0.0456531 | 0.00778948 | 4.60E-09 | rs112001035 | A | G | 0.05973 | 0.00298392 | 34.34915558 |

| | | | | | | | | | | | |
|---|---|---|---|---|---|---|---|---|---|---|---|
| | 18 | 57867526 | -0.0236616 | 0.00411123 | 8.60E-09 | rs688671 | G | A | 0.267133 | 0.00287748 | 33.12356873 |
| | 18 | 47126172 | 0.0712698 | 0.0101336 | 2.00E-12 | rs9955201 | A | G | 0.033777 | 0.00429625 | 49.46249478 |
| | 18 | 47160953 | 0.0619043 | 0.00475777 | 1.10E-38 | rs7241918 | T | G | 0.8229 | 0.01468889 | 169.2883996 |
| | 18 | 47109955 | 0.213874 | 0.0158256 | 1.30E-41 | rs77960347 | G | A | 0.013234 | 0.01584527 | 182.6367066 |
| | 19 | 11347493 | -0.0526592 | 0.00679938 | 9.60E-15 | rs737337 | C | T | 0.076789 | 0.00520926 | 59.97943072 |
| | 19 | 45413576 | -0.0322984 | 0.00448099 | 5.70E-13 | rs75627662 | T | C | 0.20606 | 0.00451243 | 51.9525083 |
| | 19 | 45448465 | 0.0416902 | 0.00378801 | 3.60E-28 | rs5167 | G | T | 0.352266 | 0.01051433 | 121.1262232 |
| | 19 | 8429323 | 0.187101 | 0.0129953 | 5.40E-47 | rs116843064 | A | G | 0.019851 | 0.01798004 | 207.2869924 |
| | 20 | 44113286 | -0.0528405 | 0.00863654 | 9.50E-10 | rs76070947 | G | C | 0.046949 | 0.00325166 | 37.43231714 |
| | 20 | 43008981 | -0.0707424 | 0.0110252 | 1.40E-10 | rs150224153 | T | C | 0.029237 | 0.00357622 | 41.16983047 |
| | 20 | 44055710 | 0.0211632 | 0.00364938 | 6.70E-09 | rs3092498 | C | T | 0.517382 | 0.00292139 | 33.62923202 |

| | | | | | | | | | | | |
|---|---|---|---|---|---|---|---|---|---|---|---|
| | 20 | 44563007 | -0.160003 | 0.00465483 | 1.00E-200 | rs1057208 | T | C | 0.186277 | 0.010162605 | 1181.519937 |
| | 22 | 21916166 | -0.0278053 | 0.00465017 | 2.20E-09 | rs5754100 | C | T | 0.188291 | 0.00310581 | 35.75279203 |

| exposure | chr.exposure | pos.exposure | beta.exposure | se.exposure | pval.exposure | SNP | effect_allele.exposure | other_allele.exposure | eaf.exposure | R2 | F |
|---|---|---|---|---|---|---|---|---|---|---|---|
| Total lipid levels in medium HDL \|\| id:ebi-a-GCST90092898 | 1 | 26987646 | -0.101858 | 0.0129347 | 3.40E-15 | rs193084249 | G | A | 0.023362 | 0.00538563 | 62.01118357 |
| | 1 | 109821307 | -0.0516319 | 0.0045661 | 1.20E-29 | rs583104 | T | G | 0.773376 | 0.01109823 | 127.8603612 |
| | 1 | 150940625 | 0.0390781 | 0.00461926 | 2.70E-17 | rs267738 | G | T | 0.219652 | 0.00621505 | 71.56728086 |
| | 1 | 230305312 | 0.0507523 | 0.00392741 | 3.40E-38 | rs1321257 | A | G | 0.613223 | 0.01448978 | 166.9903697 |
| | 1 | 220970028 | 0.034549 | 0.00419243 | 1.70E-16 | rs2642438 | G | A | 0.70385 | 0.00589761 | 67.90969428 |
| | 1 | 110232983 | 0.0402203 | 0.00428604 | 6.30E-21 | rs140584594 | G | A | 0.730101 | 0.00764608 | 88.0583663 |
| | 1 | 40035928 | -0.0365795 | 0.00482482 | 3.40E-14 | rs3768321 | T | G | 0.196524 | 0.00499217 | 57.47854298 |
| | 1 | 62906489 | -0.0511882 | 0.00403711 | 7.70E-37 | rs638714 | T | G | 0.345938 | 0.01395034 | 160.7648142 |

| | | | | | | | | | | | |
|---|---|---|---|---|---|---|---|---|---|---|---|
| | 1 | 161194641 | 0.0238813 | 0.00395091 | 1.50E-09 | rs4656292 | G | A | 0.619567 | 0.00317378 | 36.53542027 |
| | 1 | 182154990 | -0.0249795 | 0.00405336 | 7.20E-10 | rs61805076 | C | T | 0.334055 | 0.00329903 | 37.97778047 |
| | 1 | 2338126 | 0.0219196 | 0.00384581 | 1.20E-08 | rs3795269 | A | C | 0.438618 | 0.00282202 | 32.4849388 |
| | 2 | 227092802 | -0.0256463 | 0.00399535 | 1.40E-10 | rs2176040 | G | A | 0.646742 | 0.00357913 | 41.2033216 |
| | 2 | 21231524 | 0.06099 | 0.00472067 | 3.50E-38 | rs676210 | A | G | 0.205766 | 0.01448349 | 166.9177166 |
| | 2 | 27730940 | -0.0461811 | 0.00391289 | 3.80E-32 | rs1260326 | C | T | 0.603982 | 0.01208928 | 139.2918546 |
| | 2 | 211540507 | -0.024611 | 0.00412315 | 2.40E-09 | rs1047891 | A | C | 0.314984 | 0.00309498 | 35.62810338 |
| | 2 | 20363666 | -0.0273817 | 0.00384725 | 1.10E-12 | rs35135293 | T | C | 0.516755 | 0.00439968 | 50.65384902 |
| | 3 | 15592561 | -0.0220507 | 0.00403713 | 4.70E-08 | rs28818616 | C | T | 0.349768 | 0.00259167 | 29.83264396 |
| | 4 | 103188709 | -0.0652315 | 0.00729892 | 4.00E-19 | rs13107325 | T | C | 0.074282 | 0.00693568 | 79.87115702 |
| | 6 | 31319923 | -0.0357792 | 0.00479632 | 8.70E-14 | rs36057735 | G | C | 0.19903 | 0.00483312 | 55.64644223 |

| | | | | | | | | | | | |
|---|---|---|---|---|---|---|---|---|---|---|---|
| | 6 | 42915021 | 0.0281151 | 0.00383783 | 2.40E-13 | rs9471972 | A | G | 0.536187 | 0.00466612 | 53.66612659 |
| | 6 | 34223098 | –0.0335924 | 0.00565446 | 2.80E-09 | rs112495680 | G | A | 0.133414 | 0.00306591 | 35.29329823 |
| | 6 | 32458328 | 0.024794 | 0.00412329 | 1.80E-09 | rs113301125 | C | G | 0.526458 | 0.00314095 | 36.15745752 |
| | 8 | 19750044 | –0.0653556 | 0.00941676 | 3.90E-12 | rs59347135 | G | C | 0.045803 | 0.00418382 | 48.16760587 |
| | 8 | 19766233 | –0.124936 | 0.0144417 | 5.10E-18 | rs139915535 | G | A | 0.018003 | 0.00649903 | 74.83952265 |
| | 8 | 9184231 | 0.10043 | 0.00666808 | 2.90E-51 | rs4240624 | A | G | 0.909234 | 0.01967264 | 226.8390264 |
| | 8 | 144300760 | 0.0292003 | 0.00427901 | 8.80E-12 | rs56960668 | A | G | 0.277733 | 0.00404487 | 46.56723762 |
| | 8 | 116624987 | 0.0341623 | 0.00388595 | 1.50E-18 | rs2737214 | G | C | 0.558518 | 0.00671121 | 77.28447127 |
| | 8 | 19824667 | 0.0817955 | 0.00423555 | 4.30E-83 | rs15285 | T | C | 0.286526 | 0.0323018 | 372.9337169 |
| | 9 | 107647019 | 0.0508402 | 0.00616309 | 1.60E-16 | rs11789603 | T | C | 0.108822 | 0.00590954 | 68.0471489 |
| | 9 | 15304782 | 0.0353489 | 0.00551372 | 1.40E-10 | rs686030 | A | C | 0.859548 | 0.00357026 | 41.10123242 |

| | | | | | | | | | | | |
|---|---|---|---|---|---|---|---|---|---|---|---|
| | 9 | 107661742 | -0.0375225 | 0.00434814 | 6.20E-18 | rs2740488 | C | A | 0.265313 | 0.00646678 | 74.4678726 |
| | 10 | 45998984 | 0.0322525 | 0.00459851 | 5.00E-13 | rs148063610 | CAAATAAAT | C | 0.236628 | 0.00453424 | 52.20369364 |
| | 10 | 113983758 | -0.0372585 | 0.00548414 | 1.10E-11 | rs60847460 | T | C | 0.14333 | 0.00400915 | 46.15578264 |
| | 10 | 113931690 | -0.0382212 | 0.00425847 | 2.80E-19 | rs2792703 | T | C | 0.719165 | 0.00699504 | 80.55526014 |
| | 11 | 47460901 | -0.0355279 | 0.00409142 | 3.80E-18 | rs11039238 | C | T | 0.32218 | 0.00654786 | 75.40209286 |
| | 11 | 61604967 | -0.0439408 | 0.00597591 | 1.90E-13 | rs2072113 | T | C | 0.115677 | 0.00469587 | 54.0655064 |
| | 11 | 126225876 | -0.0410276 | 0.00742809 | 3.30E-08 | rs112771035 | G | C | 0.07137 | 0.00265018 | 30.50635808 |
| | 11 | 116727936 | 0.0710654 | 0.00489427 | 9.00E-48 | rs625145 | T | A | 0.188128 | 0.01828682 | 210.8302937 |
| | 11 | 75451281 | -0.0697759 | 0.00526454 | 4.30E-40 | rs559355 | T | A | 0.157979 | 0.01524124 | 175.6638863 |
| | 12 | 103556109 | -0.0237871 | 0.00385271 | 6.70E-10 | rs7956099 | C | T | 0.474982 | 0.0033113 | 38.11911496 |
| | 12 | 109873227 | 0.0306138 | 0.00382501 | 1.20E-15 | rs6606717 | C | A | 0.523165 | 0.00556314 | 64.05629032 |

| | | | | | | | | | | | |
|---|---|---|---|---|---|---|---|---|---|---|---|
| | 12 | 125324798 | -0.0550259 | 0.00588806 | 9.20E-21 | rs61941676 | A | C | 0.127218 | 0.00758321 | 87.33379005 |
| | 12 | 125259888 | -0.0372113 | 0.00416622 | 4.20E-19 | rs838876 | G | A | 0.681442 | 0.00692719 | 79.77333321 |
| | 14 | 24759319 | -0.0220218 | 0.0038506 | 1.10E-08 | rs941507 | C | G | 0.46081 | 0.00284131 | 32.70704192 |
| | 15 | 58680954 | -0.0973615 | 0.0039269 | 1.10E-135 | rs2043085 | C | T | 0.612685 | 0.05313171 | 614.7057576 |
| | 15 | 58723426 | 0.101625 | 0.00462899 | 7.91E-107 | rs1077835 | G | A | 0.219713 | 0.04170673 | 481.9711961 |
| | 16 | 56993324 | 0.1594 | 0.00408269 | 1.00E-200 | rs3764261 | A | C | 0.324412 | 0.01307259 | 1524.320451 |
| | 16 | 67976320 | -0.0620813 | 0.0105957 | 4.70E-09 | rs4986970 | T | A | 0.033919 | 0.00298212 | 34.32849395 |
| | 16 | 81534790 | 0.0272143 | 0.00416387 | 6.30E-11 | rs2925979 | C | T | 0.698949 | 0.00371049 | 42.71618556 |
| | 16 | 57049137 | -0.134929 | 0.011272 | 5.10E-33 | rs75911530 | A | G | 0.032885 | 0.01243543 | 143.2851213 |
| | 17 | 41926126 | -0.125536 | 0.0109642 | 2.40E-30 | rs72836561 | T | C | 0.031482 | 0.01137839 | 131.0916794 |
| | 17 | 66823805 | -0.0506066 | 0.0082342 | 8.00E-10 | rs112001035 | A | G | 0.05973 | 0.00328112 | 37.77147548 |

| | | | | | | | | | | | |
|---|---|---|---|---|---|---|---|---|---|---|---|
| | 17 | 76391653 | 0.0212498 | 0.0038411 | 3.20E-08 | rs4969141 | T | C | 0.489528 | 0.000265874 | 30.60489674 |
| | 17 | 26713970 | −0.0309947 | 0.00382791 | 5.60E-16 | rs967645 | T | C | 0.516505 | 0.000569373 | 65.56074651 |
| | 18 | 47109955 | 0.325493 | 0.0167183 | 2.00E-84 | rs77960347 | G | A | 0.013234 | 0.003282945 | 379.0456617 |
| | 18 | 47160953 | 0.0974937 | 0.00502614 | 8.10E-84 | rs7241918 | T | G | 0.8229 | 0.003258809 | 376.2499015 |
| | 18 | 47120600 | 0.0945219 | 0.0095307 | 3.50E-23 | rs9953437 | A | G | 0.042992 | 0.000853959 | 98.35755839 |
| | 19 | 11228745 | −0.0273005 | 0.00390969 | 2.90E-12 | rs2569550 | C | T | 0.593326 | 0.000423511 | 48.75835079 |
| | 19 | 45442528 | 0.0534587 | 0.00407933 | 3.10E-39 | rs12977604 | G | C | 0.499959 | 0.001490061 | 171.7321317 |
| | 19 | 11202306 | 0.0499243 | 0.00588677 | 2.20E-17 | rs6511720 | T | G | 0.119788 | 0.000624584 | 71.92208764 |
| | 19 | 11347657 | −0.124287 | 0.0010389 | 5.50E-33 | rs737338 | T | C | 0.035187 | 0.001242101 | 143.1186959 |
| | 19 | 54814524 | 0.0225863 | 0.00408593 | 3.20E-08 | rs1761457 | G | A | 0.326718 | 0.000265452 | 30.55630195 |
| | 19 | 8461663 | 0.0245049 | 0.00393092 | 4.60E-10 | rs12976739 | A | G | 0.396147 | 0.00033757 | 38.8606366 |

| | | | | | | | | | | | |
|---|---|---|---|---|---|---|---|---|---|---|---|
| | 19 | 45393516 | −0.0979648 | 0.0177145 | 3.20E-08 | rs117310449 | T | C | 0.011985 | 0.0026568 | 30.58262556 |
| | 19 | 45413233 | 0.162088 | 0.0070611 | 1.30E-116 | rs1065853 | T | G | 0.080581 | 0.04557909 | 526.92579 |
| | 20 | 46476143 | −0.0263887 | 0.00468 | 1.70E-08 | rs3859588 | A | T | 0.217254 | 0.00276196 | 31.79338272 |
| | 20 | 46340596 | 0.0367149 | 0.00473919 | 9.40E-15 | rs4239651 | C | T | 0.794242 | 0.00521246 | 60.01625759 |
| | 20 | 17597531 | −0.0326692 | 0.00512616 | 1.90E-10 | rs2236252 | T | C | 0.167033 | 0.00352803 | 40.61487648 |
| | 20 | 43042364 | −0.112737 | 0.0111827 | 6.70E-24 | rs1800961 | T | C | 0.0302 | 0.00882366 | 101.6323696 |
| | 21 | 46271452 | −0.0210517 | 0.00383663 | 4.10E-08 | rs235314 | T | C | 0.532702 | 0.00261549 | 30.1069638 |
| | 22 | 21982892 | −0.0428341 | 0.0049585 | 5.70E-18 | rs2298428 | T | C | 0.182552 | 0.00648022 | 74.62272596 |
| exposure | chr.exposure | pos.exposure | beta.exposure | se.exposure | pval.exposure | SNP | effect_allele.exposure | other_allele.exposure | eaf.exposure | R2 | F |
| Total lipid levels in sma | 1 | 62963737 | 0.0728806 | 0.0042967 | 1.60E-64 | rs1002687 | A | G | 0.644763 | 0.02493802 | 287.7042434 |
| | 1 | 110232983 | 0.0394141 | 0.00460597 | 1.20E-17 | rs140584594 | G | A | 0.730101 | 0.00635883 | 73.2240036 |

| | | | | | | | | | | | |
|---|---|---|---|---|---|---|---|---|---|---|---|
| 11 HDL \|\| id: ebi-a-GCST90092950 | 1 | 221032881 | 0.037596 | 0.00678185 | 3.00E-08 | rs4846335 | A | C | 0.103754 | 0.00266971 | 30.73118977 |
| | 1 | 66073835 | 0.0259107 | 0.00445644 | 6.10E-09 | rs11417022 | GT | G | 0.390629 | 0.00293662 | 33.8045078 |
| | 1 | 150940625 | 0.0417145 | 0.00496406 | 4.30E-17 | rs267738 | G | T | 0.219652 | 0.00613234 | 70.61427311 |
| | 1 | 23792009 | 0.0374954 | 0.00675801 | 2.90E-08 | rs612177 | T | G | 0.893313 | 0.00267421 | 30.78298808 |
| | 1 | 161194641 | 0.0494498 | 0.00424583 | 2.40E-31 | rs4656292 | G | A | 0.619567 | 0.01177294 | 135.6426696 |
| | 2 | 27730940 | -0.106016 | 0.00420309 | 2.20E-140 | rs1260326 | C | T | 0.603982 | 0.05497987 | 636.2062066 |
| | 2 | 20367135 | -0.0249045 | 0.00412758 | 1.60E-09 | rs11895352 | T | C | 0.475363 | 0.00316242 | 36.40467074 |
| | 3 | 98416900 | 0.02352 | 0.0041385 | 1.30E-08 | rs10935473 | T | G | 0.442432 | 0.00280582 | 32.29841185 |
| | 3 | 119546197 | -0.0251693 | 0.00423658 | 2.80E-09 | rs4261913 | T | C | 0.6194 | 0.00306599 | 35.29424561 |
| | 3 | 141169982 | -0.0245761 | 0.00417334 | 3.90E-09 | rs9878347 | C | A | 0.429825 | 0.00301245 | 34.6777449 |
| | 4 | 3443931 | -0.0381968 | 0.00425764 | 2.90E-19 | rs13108218 | G | A | 0.615355 | 0.00698884 | 80.4838122 |

| | | | | | | | | | | | |
|---|---|---|---|---|---|---|---|---|---|---|---|
| | 4 | 39414993 | 0.0260347 | 0.00423664 | 8.00E-10 | rs11940694 | G | A | 0.605104 | 0.00328029 | 37.76195611 |
| | 6 | 31303922 | 0.0263449 | 0.00426842 | 6.70E-10 | rs28752924 | C | T | 0.445537 | 0.00330908 | 38.09353436 |
| | 6 | 32587213 | 0.0409595 | 0.0042446 | 4.90E-22 | rs28383314 | C | T | 0.622964 | 0.00808495 | 93.11683496 |
| | 6 | 161111700 | -0.0979054 | 0.0173592 | 1.70E-08 | rs186696265 | T | C | 0.014352 | 0.00276329 | 31.80873009 |
| | 7 | 73037956 | -0.0813952 | 0.00602756 | 1.50E-41 | rs34060476 | G | A | 0.13482 | 0.00158204 7 | 182.3504995 |
| | 8 | 126506694 | -0.0584135 | 0.00427068 | 1.40E-42 | rs112875651 | A | G | 0.392379 | 0.01623004 | 187.0789173 |
| | 8 | 9184231 | 0.099535 | 0.00716603 | 7.30E-44 | rs4240624 | A | G | 0.909234 | 0.0167363 | 192.9242631 |
| | 8 | 11689763 | -0.0317462 | 0.00436566 | 3.50E-13 | rs34120986 | A | G | 0.371642 | 0.00459279 | 52.87813498 |
| | 9 | 136155000 | 0.0443892 | 0.00533209 | 8.40E-17 | rs635634 | T | C | 0.183543 | 0.00601854 | 69.30301775 |
| | 9 | 107647019 | 0.0364696 | 0.00661998 | 3.60E-08 | rs11789603 | T | C | 0.108822 | 0.00263649 | 30.34877023 |
| | 10 | 5257647 | -0.0349675 | 0.00570095 | 8.60E-10 | rs75406471 | A | G | 0.154659 | 0.00326802 | 37.62072454 |

| | | | | | | | | | | | |
|---|---|---|---|---|---|---|---|---|---|---|---|
| | 10 | 81105684 | -0.0228736 | 0.00415255 | 3.60E-08 | rs4246944 | T | C | 0.433587 | 0.000263583 | 30.34113032 |
| | 11 | 75474195 | -0.0761129 | 0.00707964 | 5.90E-27 | rs72997616 | A | C | 0.094104 | 0.001003346 | 115.5810519 |
| | 11 | 10324061 | 0.0250753 | 0.00414643 | 1.50E-09 | rs7924631 | G | A | 0.552641 | 0.000317686 | 36.57093395 |
| | 11 | 5701074 | -0.0452858 | 0.00801331 | 1.60E-08 | rs11601507 | A | C | 0.069168 | 0.000277442 | 31.93689223 |
| | 11 | 116705516 | -0.0904484 | 0.00454705 | 4.80E-88 | rs525028 | A | G | 0.708551 | 0.003426449 | 395.6715008 |
| | 12 | 4384844 | -0.102109 | 0.0160066 | 1.80E-10 | rs76895963 | G | T | 0.020556 | 0.000353483 | 40.69324413 |
| | 14 | 24871530 | 0.0275605 | 0.00417497 | 4.10E-11 | rs11620783 | T | C | 0.431558 | 0.000378526 | 43.57725989 |
| | 14 | 58701638 | -0.0239938 | 0.00434305 | 3.30E-08 | rs11627138 | C | T | 0.653479 | 0.000265146 | 30.52114786 |
| | 15 | 60902512 | -0.0328964 | 0.00572487 | 9.10E-09 | rs12591786 | T | C | 0.158555 | 0.000286836 | 33.01854575 |
| | 15 | 58723675 | -0.0284442 | 0.00499565 | 1.20E-08 | rs1800588 | T | C | 0.21552 | 0.000281627 | 32.41872215 |
| | 16 | 57007446 | -0.0421447 | 0.00561376 | 6.00E-14 | rs11076176 | G | T | 0.167744 | 0.000489506 | 56.3599634 |

| | | | | | | | | | | | |
|---|---|---|---|---|---|---|---|---|---|---|---|
| | 17 | 7080316 | -0.0452428 | 0.00523108 | 5.20E-18 | rs55714927 | T | C | 0.190994 | 0.00649571 | 74.80121951 |
| | 17 | 26694861 | -0.0249356 | 0.00410997 | 1.30E-09 | rs704 | A | G | 0.476026 | 0.00319754 | 36.80906569 |
| | 17 | 66463383 | -0.0303017 | 0.00496442 | 1.00E-09 | rs2952290 | A | G | 0.778744 | 0.0032363 | 37.25541518 |
| | 18 | 47181668 | 0.075969 | 0.0053621 | 1.50E-45 | rs2156552 | T | A | 0.821797 | 0.0174116 | 200.7222209 |
| | 18 | 47109955 | 0.250915 | 0.0179682 | 2.60E-44 | rs77960347 | G | A | 0.013234 | 0.01691615 | 195.0008731 |
| | 19 | 11330942 | -0.107705 | 0.0113826 | 3.00E-21 | rs17699030 | G | A | 0.033711 | 0.00777398 | 89.53260767 |
| | 19 | 19379549 | -0.0884748 | 0.00784472 | 1.70E-29 | rs58542926 | T | C | 0.074399 | 0.01104071 | 127.1969617 |
| | 19 | 45447161 | 0.0321787 | 0.00416595 | 1.10E-14 | rs5157 | C | T | 0.502351 | 0.00518175 | 59.66248466 |
| | 19 | 19458388 | -0.129836 | 0.0198123 | 5.60E-11 | rs182611493 | G | A | 0.012523 | 0.00373036 | 42.94503053 |
| | 19 | 45413233 | 0.0767396 | 0.00758997 | 5.00E-24 | rs1065853 | T | G | 0.080581 | 0.00887495 | 102.2236529 |
| | 20 | 44559748 | 0.173253 | 0.00528619 | 1.00E-200 | rs139953093 | T | G | 0.186317 | 0.09247699 | 1074.158679 |

<table>
<tr><td></td><td>20</td><td>44055710</td><td>-0.0227194</td><td>0.00414481</td><td>4.20E-08</td><td>rs3092498</td><td>C</td><td>T</td><td>0.517382</td><td>0.00261014</td><td>30.04532554</td></tr>
<tr><td></td><td>20</td><td>44113286</td><td>0.0576984</td><td>0.00980901</td><td>4.00E-09</td><td>rs76070947</td><td>G</td><td>C</td><td>0.046949</td><td>0.00300566</td><td>34.59948529</td></tr>
<tr><td></td><td>22</td><td>21982892</td><td>-0.0375139</td><td>0.00532598</td><td>1.90E-12</td><td>rs2298428</td><td>T</td><td>C</td><td>0.182552</td><td>0.00430914</td><td>49.6109819</td></tr>
<tr><td>exposure</td><td>chr.exposure</td><td>pos.exposure</td><td>beta.exposure</td><td>se.exposure</td><td>pval.exposure</td><td>SNP</td><td>effect_allele.exposure</td><td>other_allele.exposure</td><td>eaf.exposure</td><td>R2</td><td>F</td></tr>
<tr><td rowspan="7">Free cholesterol to total lipids ratio in very large HDL || id:ebi-a-GCST90093009</td><td>1</td><td>62906489</td><td>0.0225517</td><td>0.0038541</td><td>7.80E-09</td><td>rs638714</td><td>T</td><td>G</td><td>0.345925</td><td>0.00289631</td><td>33.33288356</td></tr>
<tr><td>1</td><td>182119332</td><td>0.0376622</td><td>0.00391201</td><td>6.10E-22</td><td>rs6694102</td><td>A</td><td>G</td><td>0.320916</td><td>0.0080492</td><td>92.6838165</td></tr>
<tr><td>1</td><td>161193683</td><td>0.0262579</td><td>0.00377482</td><td>3.50E-12</td><td>rs5082</td><td>A</td><td>G</td><td>0.620715</td><td>0.00420374</td><td>48.38602394</td></tr>
<tr><td>1</td><td>230303848</td><td>-0.0344953</td><td>0.00374816</td><td>3.50E-20</td><td>rs1546954</td><td>G</td><td>T</td><td>0.612656</td><td>0.00735622</td><td>84.69856649</td></tr>
<tr><td>1</td><td>26987646</td><td>0.0718665</td><td>0.0123348</td><td>5.90E-09</td><td>rs193084249</td><td>G</td><td>A</td><td>0.023363</td><td>0.00294322</td><td>33.87288583</td></tr>
<tr><td>1</td><td>40087764</td><td>0.025624</td><td>0.00444712</td><td>8.30E-09</td><td>rs61781393</td><td>A</td><td>G</td><td>0.215777</td><td>0.00288471</td><td>33.1992754</td></tr>
<tr><td>1</td><td>93866110</td><td>-0.0512836</td><td>0.00929907</td><td>3.50E-08</td><td>rs150429317</td><td>C</td><td>G</td><td>0.040657</td><td>0.00264274</td><td>30.41378175</td></tr>
</table>

| | | | | | | | | | | | |
|---|---|---|---|---|---|---|---|---|---|---|---|
| | 2 | 21416614 | -0.0285276 | 0.00461912 | 6.60E-10 | rs539845 | T | A | 0.804892 | 0.00331405 | 38.14208535 |
| | 2 | 32851488 | -0.0290371 | 0.005295 | 4.20E-08 | rs72787518 | G | T | 0.139092 | 0.00261307 | 30.07232472 |
| | 2 | 227095876 | 0.0335819 | 0.0038133 | 1.30E-18 | rs1024137 | T | G | 0.64649 | 0.00673607 | 77.55343384 |
| | 2 | 165528876 | -0.0317702 | 0.00374386 | 2.10E-17 | rs13389219 | T | C | 0.392538 | 0.00625489 | 72.0100559 |
| | 2 | 242237902 | -0.0243185 | 0.00381096 | 1.80E-10 | rs59104589 | T | C | 0.358839 | 0.00353787 | 40.71901247 |
| | 2 | 20371380 | 0.0258928 | 0.00369212 | 2.30E-12 | rs907866 | A | G | 0.443189 | 0.00427279 | 49.18117139 |
| | 3 | 50022054 | 0.0227241 | 0.0037439 | 1.30E-09 | rs62262140 | A | G | 0.536321 | 0.00320093 | 36.83980764 |
| | 3 | 98417481 | 0.0202918 | 0.00368857 | 3.80E-08 | rs62276455 | A | T | 0.444535 | 0.00262967 | 30.26338742 |
| | 3 | 124905731 | -0.0686652 | 0.0123444 | 2.70E-08 | rs77036479 | A | G | 0.02292 | 0.00268848 | 30.94039527 |
| | 3 | 12206489 | 0.0968038 | 0.0150952 | 1.40E-10 | rs11922042 | C | T | 0.017202 | 0.00357308 | 41.12439474 |
| | 3 | 185804583 | 0.0319729 | 0.00534311 | 2.20E-09 | rs57912727 | C | A | 0.136677 | 0.00311122 | 35.80702825 |

| | | | | | | | | | | | |
|---|---|---|---|---|---|---|---|---|---|---|---|
| | 4 | 103188709 | 0.0581901 | 0.00696882 | 6.80E-17 | rs13107325 | T | C | 0.074286 | 0.0060563 | 69.72237357 |
| | 4 | 100039499 | 0.0234171 | 0.00413086 | 1.40E-08 | rs2851257 | C | G | 0.72915 | 0.00279225 | 32.13495669 |
| | 5 | 75010002 | -0.021775 | 0.00380595 | 1.10E-08 | rs6864091 | C | T | 0.362504 | 0.00284418 | 32.73275961 |
| | 5 | 55857025 | 0.0362186 | 0.00465962 | 7.70E-15 | rs11429307 | GT | G | 0.191524 | 0.00524838 | 60.41638939 |
| | 6 | 31299685 | 0.0263927 | 0.00389033 | 1.20E-11 | rs9265620 | C | T | 0.370214 | 0.00399864 | 46.02430178 |
| | 6 | 34335088 | 0.0606206 | 0.00895838 | 1.30E-11 | rs147627829 | A | G | 0.044066 | 0.00397832 | 45.79034868 |
| | 6 | 43757896 | 0.0336668 | 0.00366221 | 3.80E-20 | rs998584 | A | C | 0.482326 | 0.00733988 | 84.51029941 |
| | 6 | 34893726 | 0.0458361 | 0.00815413 | 1.90E-08 | rs36037051 | G | A | 0.053446 | 0.00274556 | 31.59748418 |
| | 6 | 127511523 | 0.0248125 | 0.00370083 | 2.00E-11 | rs6569475 | A | G | 0.54604 | 0.00390539 | 44.95057246 |
| | 6 | 161092438 | 0.0404993 | 0.00606934 | 2.50E-11 | rs11751347 | T | C | 0.10225 | 0.00386843 | 44.52506378 |
| | 6 | 98583488 | -0.0247647 | 0.00374922 | 4.00E-11 | rs72934503 | G | A | 0.451402 | 0.00379062 | 43.62914911 |

| | | | | | | | | | | | |
|---|---|---|---|---|---|---|---|---|---|---|---|
| | 7 | 1029585 | 0.0329537 | 0.00376659 | 2.20E-18 | rs9769088 | C | T | 0.390937 | 0.000664835 | 76.54276033 |
| | 7 | 94953895 | 0.0230927 | 0.00369471 | 4.10E-10 | rs705379 | A | G | 0.477458 | 0.000339416 | 39.06440068 |
| | 7 | 73039406 | −0.0292202 | 0.00455309 | 1.40E-10 | rs35493868 | G | C | 0.203686 | 0.00035784 | 41.18569026 |
| | 7 | 28224053 | −0.0227618 | 0.00384559 | 3.20E-09 | rs849336 | G | A | 0.655431 | 0.000304401 | 35.03319102 |
| | 7 | 130449842 | 0.0274275 | 0.00382108 | 7.10E-13 | rs149522 | T | C | 0.404105 | 0.000447607 | 51.52199099 |
| | 8 | 9181395 | −0.0407725 | 0.00613105 | 2.90E-11 | rs2169387 | G | A | 0.899991 | 0.000384228 | 44.22393852 |
| | 8 | 116610180 | −0.0260333 | 0.00370314 | 2.10E-12 | rs2737205 | C | T | 0.561349 | 0.000429361 | 49.42089823 |
| | 8 | 126506694 | −0.0251701 | 0.00379643 | 3.40E-11 | rs112875651 | A | G | 0.392341 | 0.000381895 | 43.95532944 |
| | 8 | 19820933 | −0.0848976 | 0.00607474 | 2.20E-44 | rs145391587 | C | A | 0.100283 | 0.001694687 | 195.3115324 |
| | 8 | 144305667 | −0.0329074 | 0.00554325 | 2.90E-09 | rs79744701 | A | G | 0.12407 | 0.000306208 | 35.24119769 |
| | 8 | 19733244 | −0.0316489 | 0.00446084 | 1.30E-12 | rs1561748 | C | G | 0.267446 | 0.000437305 | 50.3357331 |

| | | | | | | | | | | | |
|---|---|---|---|---|---|---|---|---|---|---|---|
| | 9 | 107647019 | -0.0344047 | 0.00588565 | 5.10E-09 | rs11789603 | T | C | 0.108809 | 0.00296899 | 34.16954031 |
| | 9 | 107661742 | 0.0354111 | 0.00415215 | 1.50E-17 | rs2740488 | C | A | 0.265334 | 0.00631756 | 72.73194764 |
| | 9 | 15303583 | -0.0413442 | 0.00532381 | 8.10E-15 | rs1215112 | A | G | 0.863047 | 0.00523899 | 60.30822425 |
| | 10 | 45998984 | -0.0327515 | 0.00439166 | 8.80E-14 | rs148063610 | CAAATAAAT | C | 0.236625 | 0.00483154 | 55.61570173 |
| | 10 | 94842213 | -0.0225495 | 0.0036696 | 8.00E-10 | rs10882140 | T | G | 0.53343 | 0.00328084 | 37.75972559 |
| | 10 | 113921825 | 0.0260185 | 0.00407242 | 1.70E-10 | rs2792735 | A | G | 0.720359 | 0.00354647 | 40.81801097 |
| | 11 | 47316568 | -0.037997 | 0.00429075 | 8.30E-19 | rs113012334 | GT | G | 0.289463 | 0.00681125 | 78.41960421 |
| | 11 | 61586328 | 0.0618717 | 0.00384772 | 3.50E-58 | rs35473591 | CT | C | 0.341909 | 0.02242296 | 258.5649343 |
| | 11 | 116791391 | -0.0623855 | 0.010487 | 2.70E-09 | rs79489590 | T | C | 0.031333 | 0.00307484 | 35.38809912 |
| | 11 | 117156893 | -0.153425 | 0.0186007 | 1.60E-16 | rs1047964 | T | G | 0.010067 | 0.00590973 | 68.03402072 |
| | 11 | 126225876 | 0.0677691 | 0.00709351 | 1.30E-21 | rs112771035 | G | C | 0.071378 | 0.00792661 | 91.27114865 |

| | | | | | | | | | | | |
|---|---|---|---|---|---|---|---|---|---|---|---|
| | 12 | 121368518 | -0.0231065 | 0.00374104 | 6.60E-10 | rs3999421 | T | A | 0.392026 | 0.00331459 | 38.1483783 |
| | 12 | 124478586 | 0.0354193 | 0.00571572 | 5.80E-10 | rs825510 | C | A | 0.115135 | 0.00333644 | 38.3999366 |
| | 12 | 125259888 | 0.0413967 | 0.00397748 | 2.30E-25 | rs838876 | G | A | 0.681461 | 0.00940584 | 108.319809 |
| | 12 | 57848708 | -0.0245489 | 0.00403923 | 1.20E-09 | rs3809113 | G | C | 0.710436 | 0.00320936 | 36.93680764 |
| | 12 | 125324798 | 0.0548539 | 0.00562132 | 1.70E-22 | rs61941676 | A | C | 0.127212 | 0.00826932 | 95.22058695 |
| | 12 | 109871179 | -0.0232205 | 0.00365235 | 2.00E-10 | rs7308864 | G | A | 0.52328 | 0.00351185 | 40.41948794 |
| | 12 | 125077567 | -0.0246248 | 0.00382029 | 1.20E-10 | rs837500 | T | C | 0.638493 | 0.00360983 | 41.5475301 |
| | 15 | 58580781 | 0.0760053 | 0.0044305 | 5.80E-66 | rs1560390 | C | T | 0.219666 | 0.02551312 | 294.2894708 |
| | 15 | 58737661 | 0.168354 | 0.00520638 | 1.00E-200 | rs488490 | A | C | 0.852643 | 0.09006102 | 1045.604885 |
| | 15 | 58678512 | -0.178154 | 0.0039995 | 1.00E-200 | rs10468017 | T | C | 0.295943 | 0.16952931 | 1984.139504 |
| | 15 | 59058720 | 0.0538562 | 0.00823854 | 6.30E-11 | rs77516617 | A | G | 0.052808 | 0.00371279 | 42.73299732 |

| | | | | | | | | | | | |
|---|---|---|---|---|---|---|---|---|---|---|---|
| | 16 | 15130594 | 0.0279695 | 0.00396363 | 1.70E-12 | rs6498540 | G | A | 0.302067 | 0.00432599 | 49.79384263 |
| | 16 | 56752882 | 0.0747832 | 0.0125918 | 2.90E-09 | rs75714888 | A | C | 0.021625 | 0.00306471 | 35.27157934 |
| | 16 | 56915357 | -0.0760374 | 0.0132935 | 1.10E-08 | rs55779455 | C | G | 0.01985 | 0.00284278 | 32.71660928 |
| | 16 | 57049137 | 0.141158 | 0.0107351 | 1.70E-39 | rs75911530 | A | G | 0.032884 | 0.01500505 | 172.8985117 |
| | 16 | 56993324 | -0.210212 | 0.00388812 | 1.00E-200 | rs3764261 | A | C | 0.324407 | 0.02475978 | 2922.995446 |
| | 16 | 81534790 | -0.0290024 | 0.00396551 | 2.60E-13 | rs2925979 | C | T | 0.698934 | 0.00464685 | 53.48872279 |
| | 16 | 67976320 | 0.0882436 | 0.0100914 | 2.20E-18 | rs4986970 | T | A | 0.033914 | 0.00664149 | 76.46382926 |
| | 16 | 56946502 | 0.136413 | 0.0230139 | 3.10E-09 | rs117738782 | C | T | 0.007459 | 0.00305274 | 35.13367078 |
| | 17 | 7080316 | -0.0266087 | 0.00465036 | 1.10E-08 | rs55714927 | T | C | 0.190994 | 0.00284473 | 32.73909092 |
| | 17 | 37808759 | 0.0405999 | 0.0067647 | 2.00E-09 | rs34695955 | G | T | 0.079548 | 0.00312974 | 36.02013165 |
| | 17 | 37746847 | -0.0269032 | 0.00411283 | 6.10E-11 | rs4795386 | G | A | 0.723034 | 0.00371754 | 42.78768734 |

| | | | | | | | | | | | |
|---|---|---|---|---|---|---|---|---|---|---|---|
| | 17 | 41926126 | 0.106071 | 0.0104738 | 4.20E-24 | rs72836561 | T | C | 0.031467 | 0.00890612 | 102.5598138 |
| | 18 | 47118219 | -0.0647077 | 0.00918203 | 1.80E-12 | rs3786248 | C | T | 0.042089 | 0.00431457 | 49.66231527 |
| | 18 | 47160953 | -0.0621185 | 0.00480252 | 2.90E-38 | rs7241918 | T | G | 0.822891 | 0.01451989 | 167.30000253 |
| | 18 | 47109955 | -0.218294 | 0.0159732 | 1.60E-42 | rs77960347 | G | A | 0.013237 | 0.01620638 | 186.7635797 |
| | 19 | 11347493 | 0.0479931 | 0.00686387 | 2.70E-12 | rs737337 | C | T | 0.076784 | 0.00424743 | 48.88909048 |
| | 19 | 45425178 | -0.0918558 | 0.00837713 | 5.60E-28 | rs190712692 | A | G | 0.05362 | 0.01043902 | 120.230626 |
| | 19 | 54815577 | -0.0219994 | 0.00390951 | 1.80E-08 | rs759819 | C | T | 0.326736 | 0.00275137 | 31.66426901 |
| | 19 | 45448465 | -0.0651813 | 0.00382417 | 3.80E-65 | rs5167 | G | T | 0.352224 | 0.02518646 | 290.51202 |
| | 19 | 8429323 | -0.12811 | 0.0131183 | 1.60E-22 | rs116843064 | A | G | 0.019851 | 0.00828213 | 95.36815835 |
| | 20 | 44076549 | 0.0254578 | 0.00446132 | 1.20E-08 | rs16990165 | T | C | 0.251033 | 0.00282932 | 32.56172774 |
| | 20 | 44563007 | 0.166601 | 0.00469931 | 1.00E-200 | rs1057208 | T | C | 0.186258 | 0.01080585 9 | 1256.838478 |

| | | | | | | | | | | | |
|---|---|---|---|---|---|---|---|---|---|---|---|
| | 20 | 44113286 | 0.0581052 | 0.00871871 | 2.70E-11 | rs76070947 | G | C | 0.046951 | 0.00385877 | 44.41380551 |
| | 22 | 21922456 | 0.0278227 | 0.00466783 | 2.50E-09 | rs11089620 | G | C | 0.189494 | 0.00308692 | 35.52719624 |
| | 22 | 44340904 | -0.0237315 | 0.00434361 | 4.70E-08 | rs2294915 | T | C | 0.231796 | 0.00259374 | 29.8497832 |
| exposure | chr.exposure | pos.exposure | beta.exposure | se.exposure | pval.exposure | SNP | effect_allele.exposure | other_allele.exposure | eaf.exposure | R2 | F |
| Free cholesterol to total lipids ratio in large HDL \|\| id:ebi-a-GCST90092849 | 1 | 55505647 | -0.214652 | 0.0146235 | 8.80E-49 | rs11591147 | T | G | 0.017457 | 0.01868734 | 215.4565355 |
| | 1 | 55521313 | 0.0269116 | 0.00401707 | 2.10E-11 | rs472495 | T | G | 0.649121 | 0.00389837 | 44.87998376 |
| | 1 | 110232983 | -0.0287791 | 0.00429742 | 2.10E-11 | rs140584594 | G | A | 0.730101 | 0.00389549 | 44.84678014 |
| | 1 | 220970028 | 0.0281087 | 0.00420356 | 2.30E-11 | rs2642438 | G | A | 0.70385 | 0.00388393 | 44.71358293 |
| | 1 | 11913757 | 0.02149 | 0.0039142 | 4.00E-08 | rs198375 | C | T | 0.397603 | 0.00261858 | 30.14249921 |
| | 1 | 55492083 | -0.0236179 | 0.00431958 | 4.60E-08 | rs59784135 | G | A | 0.696422 | 0.00259704 | 29.89454638 |
| | 1 | 109817590 | -0.081325 | 0.00461742 | 2.00E-69 | rs12740374 | T | G | 0.221064 | 0.0268827 | 310.2000031 |

| | | | | | | | | | | | |
|---|---|---|---|---|---|---|---|---|---|---|---|
| | 1 | 161193683 | −0.0412514 | 0.00396466 | 2.40E-25 | rs5082 | A | G | 0.620747 | 0.00939831 | 108.2574863 |
| | 1 | 178532188 | −0.0219359 | 0.00384933 | 1.20E-08 | rs4650999 | T | G | 0.533408 | 0.00282105 | 32.47379754 |
| | 2 | 21288226 | 0.0932491 | 0.00497901 | 2.90E-78 | rs563290 | A | G | 0.819463 | 0.03038605 | 350.748444 |
| | 2 | 44055510 | −0.0762858 | 0.00804376 | 2.50E-21 | rs114938914 | T | C | 0.061412 | 0.00780949 | 89.9418164 |
| | 2 | 165514409 | 0.0257074 | 0.0040384 | 1.90E-10 | rs11394662 | AT | A | 0.384759 | 0.00351996 | 40.52192759 |
| | 2 | 27741237 | 0.048176 | 0.00394118 | 2.30E-34 | rs780094 | C | T | 0.615153 | 0.01296696 | 149.4174734 |
| | 2 | 227112754 | −0.0300878 | 0.00400046 | 5.40E-14 | rs2943654 | T | C | 0.64513 | 0.00491293 | 56.56573764 |
| | 2 | 111894720 | −0.0437966 | 0.00784738 | 2.40E-08 | rs17041868 | C | T | 0.063607 | 0.00270587 | 31.14754912 |
| | 3 | 12450838 | −0.145489 | 0.0229815 | 2.40E-10 | rs147667955 | T | C | 0.007221 | 0.00348133 | 40.07707587 |
| | 3 | 98312581 | −0.0220745 | 0.00384794 | 9.70E-09 | rs4857406 | C | G | 0.531506 | 0.00285886 | 32.90922235 |
| | 4 | 103146890 | −0.0391767 | 0.00716254 | 4.50E-08 | rs35518360 | T | A | 0.08273 | 0.00259897 | 29.91672391 |

| | | | | | | | | | | | |
|---|---|---|---|---|---|---|---|---|---|---|---|
| | 4 | 3443931 | 0.0280917 | 0.00397428 | 1.60E-12 | rs13108218 | G | A | 0.615355 | 0.00433954 | 49.96105189 |
| | 5 | 74472939 | 0.033151 | 0.0040006 | 1.20E-16 | rs7707394 | A | G | 0.357075 | 0.00596316 | 68.66500531 |
| | 5 | 74666492 | 0.0448717 | 0.00395057 | 6.70E-30 | rs7733436 | T | C | 0.381464 | 0.01119777 | 129.0083962 |
| | 5 | 55860866 | -0.0289774 | 0.00478168 | 1.40E-09 | rs3936510 | T | G | 0.201952 | 0.00319016 | 36.72402431 |
| | 5 | 32716589 | 0.0239592 | 0.00433295 | 3.20E-08 | rs10057069 | C | T | 0.271258 | 0.00265616 | 30.57522932 |
| | 6 | 31243347 | -0.032856 | 0.00507264 | 9.40E-11 | rs9461680 | T | C | 0.174798 | 0.00364414 | 41.95210414 |
| | 6 | 98583488 | 0.0229136 | 0.003937 | 5.90E-09 | rs72934503 | G | A | 0.451389 | 0.00294253 | 33.87258003 |
| | 6 | 161092438 | -0.0492121 | 0.00637325 | 1.10E-14 | rs11751347 | T | C | 0.102257 | 0.00517833 | 59.62307063 |
| | 6 | 43758873 | -0.0241831 | 0.00386855 | 4.10E-10 | rs6905288 | A | G | 0.568525 | 0.00339448 | 39.07688773 |
| | 7 | 94953895 | -0.029329 | 0.00387986 | 4.10E-14 | rs705379 | A | G | 0.477476 | 0.00496294 | 57.14192327 |
| | 7 | 1029585 | -0.0360487 | 0.00395541 | 8.00E-20 | rs9769088 | C | T | 0.390936 | 0.00721233 | 83.05937395 |

| | | | | | | | | | | | |
|---|---|---|---|---|---|---|---|---|---|---|---|
| | 7 | 28189411 | 0.0221716 | 0.00384997 | 8.50E-09 | rs1635852 | C | T | 0.505957 | 0.00288102 | 33.16437608 |
| | 7 | 73037956 | 0.0881121 | 0.00562652 | 2.80E-55 | rs34060476 | G | A | 0.13482 | 0.002126471 | 245.2357734 |
| | 7 | 75044830 | -0.0232468 | 0.00406828 | 1.10E-08 | rs236651 | A | G | 0.573853 | 0.00283644 | 32.65104906 |
| | 8 | 11435927 | -0.0382996 | 0.00386094 | 3.40E-23 | rs2409799 | C | G | 0.529588 | 0.00854326 | 98.39993772 |
| | 8 | 19830170 | 0.0923099 | 0.0068608 | 2.90E-41 | rs1569209 | G | T | 0.085308 | 0.01570569 | 181.0253912 |
| | 9 | 15298666 | 0.0303567 | 0.00555253 | 4.60E-08 | rs675849 | C | T | 0.860645 | 0.00259661 | 29.88957518 |
| | 10 | 113949664 | -0.0329289 | 0.00431352 | 2.30E-14 | rs10787429 | C | T | 0.725575 | 0.00506132 | 58.27513211 |
| | 10 | 65301725 | 0.0249038 | 0.00384342 | 9.20E-11 | rs7897379 | C | T | 0.479379 | 0.00364695 | 41.9844048 |
| | 11 | 5701074 | 0.0544428 | 0.00747489 | 3.30E-13 | rs11601507 | A | C | 0.069168 | 0.00460748 | 53.04735645 |
| | 11 | 47322007 | -0.0284459 | 0.00469967 | 1.40E-09 | rs200644264 | CG | C | 0.788465 | 0.00318243 | 36.6350748 |
| | 11 | 117156893 | 0.167262 | 0.0195168 | 1.00E-17 | rs1047964 | T | G | 0.010073 | 0.00637812 | 73.44627932 |

| | | | | | | | | | | | |
|---|---|---|---|---|---|---|---|---|---|---|---|
| | 11 | 61557826 | －0.113451 | 0.00403304 | 4.20E-174 | rs102274 | C | T | 0.343868 | 0.006829174 | 791.3052555 |
| | 11 | 10324061 | －0.02413 | 0.00386784 | 4.40E-10 | rs7924631 | G | A | 0.552641 | 0.00338083 | 38.91975479 |
| | 11 | 116648917 | 0.0893976 | 0.0056557 | 2.80E-56 | rs964184 | C | G | 0.86726 | 0.002166356 | 249.8454486 |
| | 12 | 125265201 | －0.0278395 | 0.00417151 | 2.50E-11 | rs921919 | A | G | 0.66951 | 0.00386866 | 44.53779751 |
| | 12 | 112007756 | 0.0228574 | 0.00384645 | 2.80E-09 | rs653178 | T | C | 0.516036 | 0.00306756 | 35.31229357 |
| | 12 | 125347764 | －0.0291983 | 0.00405369 | 5.90E-13 | rs10692614 | TCA | T | 0.638692 | 0.0045062 | 51.88078254 |
| | 12 | 123209159 | 0.0261148 | 0.00473301 | 3.40E-08 | rs2454702 | T | C | 0.209964 | 0.0026447 | 30.44325017 |
| | 13 | 32976651 | －0.0270853 | 0.00405969 | 2.50E-11 | rs61946988 | T | G | 0.451462 | 0.00386639 | 44.51168235 |
| | 15 | 58646053 | －0.0437235 | 0.00489139 | 3.90E-19 | rs11071373 | G | A | 0.194289 | 0.00693835 | 79.90201069 |
| | 15 | 58499265 | 0.100616 | 0.0163602 | 7.70E-10 | rs4775022 | G | A | 0.985144 | 0.00328554 | 37.82242158 |
| | 15 | 58725839 | 0.112364 | 0.00423045 | 1.90E-155 | rs633695 | G | A | 0.292354 | 0.006092839 | 705.462124 |

| | | | | | | | | | | | |
|---|---|---|---|---|---|---|---|---|---|---|---|
| | 15 | 58678512 | 0.149189 | 0.00420569 | 1.00E-200 | rs10468017 | T | C | 0.295937 | 0.010816052 | 1258.321284 |
| | 16 | 56852822 | 0.0261542 | 0.00388326 | 1.60E-11 | rs1561139 | T | G | 0.42589 | 0.00394014 | 45.36097645 |
| | 16 | 15148646 | 0.0368992 | 0.00420882 | 1.80E-18 | rs11075253 | A | C | 0.295789 | 0.00667446 | 76.86093234 |
| | 16 | 81533789 | 0.0249022 | 0.00419907 | 3.00E-09 | rs56823429 | A | C | 0.701601 | 0.00305513 | 35.16913258 |
| | 16 | 56993886 | 0.1184 | 0.00410006 | 2.30E-183 | rs821840 | G | A | 0.324526 | 0.007194157 | 833.9028013 |
| | 17 | 64195431 | 0.073466 | 0.0108578 | 1.30E-11 | rs75003668 | G | A | 0.033354 | 0.00397657 | 45.78060903 |
| | 17 | 66447073 | 0.0233565 | 0.00425077 | 3.90E-08 | rs2302783 | C | T | 0.714873 | 0.00262276 | 30.19067105 |
| | 17 | 41926126 | -0.103937 | 0.0109975 | 3.40E-21 | rs72836561 | T | C | 0.031482 | 0.00775547 | 89.31920847 |
| | 17 | 67081278 | 0.122238 | 0.0129161 | 3.00E-21 | rs77542162 | G | A | 0.022666 | 0.00777686 | 89.56578279 |
| | 19 | 45358353 | 0.0437284 | 0.0038834 | 2.10E-29 | rs10410835 | C | T | 0.523103 | 0.01100569 | 126.7930281 |
| | 19 | 8570490 | 0.0356928 | 0.00596711 | 2.20E-09 | rs201801505 | CA | C | 0.123685 | 0.00310807 | 35.77878608 |

| | | | | | | | | | | | |
|---|---|---|---|---|---|---|---|---|---|---|---|
| | 19 | 45421650 | 0.136312 | 0.0116783 | 1.80E-31 | rs140480140 | A | G | 0.028202 | 0.001182463 | 136.238978 |
| | 19 | 11192915 | -0.106922 | 0.00587463 | 5.10E-74 | rs73015021 | G | A | 0.120658 | 0.002870235 | 331.2573798 |
| | 19 | 11147526 | -0.112027 | 0.018444 | 1.20E-09 | rs73013176 | C | T | 0.011281 | 0.0032061 | 36.9076192 |
| | 19 | 45173951 | -0.102815 | 0.00889559 | 6.70E-31 | rs28807203 | C | A | 0.049207 | 0.00115945 | 133.5844468 |
| | 19 | 45413233 | -0.43769 | 0.00706517 | 1.00E-200 | rs1065853 | T | G | 0.080581 | 0.032272578 | 3837.783476 |
| | 20 | 44563007 | -0.221644 | 0.00493376 | 1.00E-200 | rs1057208 | T | C | 0.186277 | 0.01723449 | 2018.126437 |
| | 20 | 44776543 | -0.0759369 | 0.0106515 | 1.00E-12 | rs2024570 | C | T | 0.962637 | 0.00441454 | 50.82491214 |
| | 20 | 44105636 | -0.0463506 | 0.00600743 | 1.20E-14 | rs6065819 | A | C | 0.116575 | 0.00517013 | 59.52860908 |
| | 20 | 17844684 | -0.0327879 | 0.0040523 | 5.90E-16 | rs2618566 | T | G | 0.660294 | 0.00568551 | 65.46610111 |
| | 20 | 44055710 | 0.0287702 | 0.00386807 | 1.00E-13 | rs3092498 | C | T | 0.517382 | 0.00480486 | 55.32094174 |
| exposure | chr.exposure | pos.exposure | beta.exposure | se.exposure | pval.exposure | SNP | effect_allele.exposure | other_allele.exposure | eaf.exposure | R2 | F |

| | | | | | | | | | | | |
|---|---|---|---|---|---|---|---|---|---|---|---|
| Free cholesterol to total lipids ratio in medium HDL \|\| id: ebi-a-GCST90092897 | 1 | 220970028 | 0.0359371 | 0.00400625 | 3.00E-19 | rs2642438 | G | A | 0.703885 | 0.00698713 | 80.46414759 |
| | 1 | 40035928 | -0.0348095 | 0.00461056 | 4.40E-14 | rs3768321 | T | G | 0.196524 | 0.00495069 | 57.00076265 |
| | 1 | 182119332 | -0.0265187 | 0.00391565 | 1.30E-11 | rs6694102 | A | G | 0.320921 | 0.00398397 | 45.86582207 |
| | 1 | 26987646 | -0.0678549 | 0.0123603 | 4.00E-08 | rs193084249 | G | A | 0.023362 | 0.00261808 | 30.13678074 |
| | 1 | 230304352 | 0.0391352 | 0.00375247 | 1.80E-25 | rs4846921 | A | G | 0.613232 | 0.00944241 | 108.7659882 |
| | 1 | 55505647 | -0.0814009 | 0.0139371 | 5.20E-09 | rs11591147 | T | G | 0.017457 | 0.00296331 | 34.1119097 |
| | 1 | 109817590 | -0.0310165 | 0.00440068 | 1.80E-12 | rs12740374 | T | G | 0.221064 | 0.00431471 | 49.67506585 |
| | 2 | 165528876 | 0.0326975 | 0.0037465 | 2.60E-18 | rs13389219 | T | C | 0.392532 | 0.00661428 | 76.16756562 |
| | 2 | 227105921 | -0.0383378 | 0.00381556 | 9.40E-24 | rs2943650 | T | C | 0.644754 | 0.00876495 | 100.9555625 |
| | 2 | 20363074 | -0.0231696 | 0.00367423 | 2.90E-10 | rs35633876 | T | G | 0.481858 | 0.0034542 | 39.76462096 |
| | 2 | 21237544 | 0.0627809 | 0.00450767 | 4.30E-44 | rs673548 | A | G | 0.206075 | 0.01682721 | 193.9739001 |

| | | | | | | | | | | | |
|---|---|---|---|---|---|---|---|---|---|---|---|
| | 2 | 21471603 | 0.02987 | 0.00455831 | 5.60E-11 | rs1848922 | C | T | 0.795813 | 0.000372987 | 42.93932204 |
| | 2 | 219699999 | -0.0597124 | 0.0094693 | 2.90E-10 | rs78058190 | A | G | 0.050108 | 0.000345411 | 39.76360534 |
| | 3 | 185804583 | -0.0297721 | 0.00534683 | 2.60E-08 | rs57912727 | C | A | 0.136685 | 0.000269341 | 31.00406496 |
| | 4 | 970112 | 0.02062 | 0.00370208 | 2.50E-08 | rs12510382 | G | A | 0.459916 | 0.000269502 | 31.02259874 |
| | 4 | 103188709 | -0.0716693 | 0.00697525 | 9.20E-25 | rs13107325 | T | C | 0.074282 | 0.000916518 | 105.56968942 |
| | 4 | 69373407 | 0.0263044 | 0.00437283 | 1.80E-09 | rs9884390 | C | T | 0.234037 | 0.000314331 | 36.18462405 |
| | 5 | 74472939 | 0.0222206 | 0.0038139 | 5.70E-09 | rs7707394 | A | G | 0.357075 | 0.000294875 | 33.94418706 |
| | 5 | 55857025 | -0.0370418 | 0.00466502 | 2.00E-15 | rs11429307 | GT | G | 0.191517 | 0.00054756 | 63.04775072 |
| | 5 | 75003678 | 0.0331994 | 0.0037451 | 7.70E-19 | rs2307111 | C | T | 0.396595 | 0.000682385 | 78.58254415 |
| | 6 | 34580221 | -0.0310045 | 0.00397558 | 6.30E-15 | rs2744973 | T | C | 0.306849 | 0.000528216 | 60.81923065 |
| | 6 | 31299685 | -0.0279211 | 0.00389321 | 7.40E-13 | rs9265620 | C | T | 0.370213 | 0.000446733 | 51.43299803 |

| | | | | | | | | | | | |
|---|---|---|---|---|---|---|---|---|---|---|---|
| | 6 | 127521127 | −0.0225301 | 0.00370437 | 1.20E-09 | rs4318902 | G | A | 0.556738 | 0.00321329 | 36.99048063 |
| | 6 | 161092438 | −0.0611132 | 0.00607368 | 7.90E-24 | rs11751347 | T | C | 0.102257 | 0.00879516 | 101.3038259 |
| | 6 | 34247047 | −0.0507015 | 0.00884474 | 9.90E-09 | rs77136196 | T | C | 0.045494 | 0.00285457 | 32.85972517 |
| | 6 | 43758873 | −0.0253976 | 0.00368671 | 5.60E-12 | rs6905288 | A | G | 0.568525 | 0.00412212 | 47.45695363 |
| | 6 | 98583488 | 0.0206614 | 0.00375195 | 3.70E-08 | rs72934503 | G | A | 0.451389 | 0.00263441 | 30.324794 |
| | 7 | 94953895 | −0.0204191 | 0.00369751 | 3.30E-08 | rs705379 | A | G | 0.477476 | 0.00264931 | 30.49628446 |
| | 7 | 1029585 | −0.031285 | 0.00376951 | 1.00E-16 | rs9769088 | C | T | 0.390936 | 0.00598185 | 68.88029072 |
| | 7 | 73033559 | 0.0498623 | 0.00543733 | 4.70E-20 | rs34062580 | A | G | 0.130304 | 0.00730211 | 84.0941303 |
| | 7 | 130437476 | −0.0232778 | 0.00406726 | 1.00E-08 | rs7810507 | A | G | 0.281085 | 0.00284544 | 32.754611 |
| | 8 | 19733244 | 0.0551805 | 0.00446135 | 3.90E-35 | rs1561748 | C | G | 0.267445 | 0.01327561 | 152.9787556 |
| | 8 | 9181395 | 0.0755834 | 0.00613144 | 6.50E-35 | rs2169387 | G | A | 0.899996 | 0.01318704 | 151.9568684 |

| | | | | | | | | | | | |
|---|---|---|---|---|---|---|---|---|---|---|---|
| | 8 | 19819724 | 0.139434 | 0.00607316 | 1.20E-116 | rs328 | G | C | 0.100307 | 0.00459485 | 527.1089364 |
| | 8 | 9256631 | -0.0226938 | 0.00391591 | 6.80E-09 | rs17729883 | C | T | 0.334631 | 0.00291753 | 33.58470168 |
| | 8 | 73435156 | 0.0232238 | 0.00413512 | 2.00E-08 | rs6993128 | T | C | 0.728947 | 0.00274008 | 31.541532 |
| | 8 | 116624879 | 0.0268686 | 0.00371096 | 4.50E-13 | rs2245221 | A | G | 0.558968 | 0.00455315 | 52.42156495 |
| | 9 | 107586753 | 0.0374661 | 0.00544083 | 5.70E-12 | rs2066714 | C | T | 0.12893 | 0.00411887 | 47.41755049 |
| | 9 | 107661742 | -0.0491764 | 0.00415639 | 2.70E-32 | rs2740488 | C | A | 0.265313 | 0.01214914 | 139.9823678 |
| | 9 | 15303583 | 0.062774 | 0.00532877 | 4.90E-32 | rs1215112 | A | G | 0.86303 | 0.01204411 | 138.7708049 |
| | 9 | 107647019 | 0.0584636 | 0.0058913 | 3.30E-23 | rs11789603 | T | C | 0.108822 | 0.00855008 | 98.47846796 |
| | 10 | 45998984 | 0.0317767 | 0.00439629 | 4.90E-13 | rs148063610 | CAAATAAAT | C | 0.236628 | 0.00453775 | 52.24411473 |
| | 10 | 113983758 | -0.0317898 | 0.00524297 | 1.30E-09 | rs60847460 | T | C | 0.14333 | 0.00319356 | 36.76319927 |
| | 10 | 65191645 | 0.0221147 | 0.00365952 | 1.50E-09 | rs7924036 | T | G | 0.504241 | 0.00317226 | 36.51794304 |

| | | | | | | | | | | | |
|---|---|---|---|---|---|---|---|---|---|---|---|
| | 10 | 113921825 | -0.0398204 | 0.00407654 | 1.50E-22 | rs2792735 | A | G | 0.72034 | 0.00828439 | 95.4157949 |
| | 11 | 116648917 | 0.0824385 | 0.00538878 | 7.90E-53 | rs964184 | C | G | 0.86726 | 0.02029502 | 234.030094 |
| | 11 | 116916060 | -0.147749 | 0.0185073 | 1.40E-15 | rs144018203 | C | G | 0.010653 | 0.00553497 | 63.73168823 |
| | 11 | 117156893 | 0.210099 | 0.0185957 | 1.30E-29 | rs1047964 | T | G | 0.010073 | 0.01107985 | 127.6483956 |
| | 11 | 61592362 | -0.10349 | 0.00382922 | 7.29E-161 | rs174566 | G | A | 0.348122 | 0.006306973 | 730.4131596 |
| | 11 | 27505954 | 0.0277138 | 0.0043152 | 1.30E-10 | rs12786130 | T | C | 0.761797 | 0.00358284 | 41.24608678 |
| | 11 | 47362465 | 0.0413948 | 0.00469487 | 1.20E-18 | rs10769254 | C | G | 0.18914 | 0.00675062 | 77.73863079 |
| | 11 | 116990507 | 0.0605811 | 0.0102971 | 4.00E-09 | rs116978226 | A | C | 0.033251 | 0.00300681 | 34.6128134 |
| | 11 | 47322007 | -0.0451258 | 0.00447786 | 6.90E-24 | rs200644264 | CG | C | 0.788465 | 0.00881695 | 101.5549878 |
| | 11 | 64004643 | -0.048942 | 0.00874156 | 2.20E-08 | rs12294913 | G | C | 0.049058 | 0.00272308 | 31.3457014 |
| | 11 | 75451281 | -0.0369578 | 0.00502763 | 2.00E-13 | rs559355 | T | A | 0.157979 | 0.00469326 | 54.03536087 |

| | | | | | | | | | | | |
|---|---|---|---|---|---|---|---|---|---|---|---|
| | 11 | 118957388 | -0.0240172 | 0.00385473 | 4.60E-10 | rs141949189 | T | A | 0.431973 | 0.000337212 | 38.81944071 |
| | 12 | 124496316 | 0.0222168 | 0.00396171 | 2.00E-08 | rs1054852 | G | A | 0.377477 | 0.000273194 | 31.44778776 |
| | 12 | 20473758 | 0.0205362 | 0.00368891 | 2.60E-08 | rs7134375 | A | C | 0.430211 | 0.000269228 | 30.99106518 |
| | 12 | 57860826 | 0.0222239 | 0.00393071 | 1.60E-08 | rs4760259 | C | T | 0.681952 | 0.000277697 | 31.96619862 |
| | 12 | 111910219 | 0.0260485 | 0.00366336 | 1.20E-12 | rs10774625 | G | A | 0.502824 | 0.000439145 | 50.55902536 |
| | 12 | 123348440 | 0.0509564 | 0.00791058 | 1.20E-10 | rs142265900 | AGAT | A | 0.057566 | 0.000360427 | 41.49285083 |
| | 12 | 125265201 | -0.0388652 | 0.00397494 | 1.40E-22 | rs921919 | A | G | 0.66951 | 0.000830028 | 95.59894754 |
| | 12 | 125326153 | -0.0376059 | 0.00452833 | 1.00E-16 | rs7136506 | C | T | 0.215774 | 0.000598919 | 68.96492836 |
| | 15 | 58725839 | 0.108856 | 0.0040321 | 1.60E-160 | rs633695 | G | A | 0.292354 | 0.0006293509 | 728.8440403 |
| | 15 | 43757184 | -0.0787064 | 0.0120047 | 5.50E-11 | rs190543502 | C | T | 0.024111 | 0.000373377 | 42.9843067 |
| | 15 | 58569330 | 0.0425205 | 0.00541774 | 4.20E-15 | rs34663616 | A | C | 0.137646 | 0.000534959 | 61.59604953 |

| | | | | | | | | | | | |
|---|---|---|---|---|---|---|---|---|---|---|---|
| | 15 | 58833993 | 0.0626207 | 0.0109298 | 1.00E-08 | rs6078 | A | G | 0.028853 | 0.00285154 | 32.82493376 |
| | 15 | 58678720 | -0.135384 | 0.00385121 | 1.00E-200 | rs261290 | C | T | 0.654645 | 0.01062415 | 1235.756018 |
| | 16 | 67976320 | -0.0980976 | 0.0101005 | 2.70E-22 | rs4986970 | T | A | 0.033919 | 0.0081897 | 94.32427397 |
| | 16 | 81534790 | 0.0330254 | 0.00396924 | 8.80E-17 | rs2925979 | C | T | 0.698949 | 0.00601192 | 69.22674417 |
| | 16 | 15172118 | 0.0354986 | 0.00400153 | 7.20E-19 | rs11644601 | C | T | 0.296695 | 0.00683386 | 78.69782852 |
| | 16 | 57049137 | -0.146939 | 0.0107451 | 1.40E-42 | rs75911530 | A | G | 0.032885 | 0.01622336 | 187.0017578 |
| | 16 | 56752882 | -0.0858122 | 0.0126041 | 9.90E-12 | rs75714888 | A | C | 0.021624 | 0.00402617 | 46.35182568 |
| | 16 | 56993324 | 0.208301 | 0.00389186 | 1.00E-200 | rs3764261 | A | C | 0.324412 | 0.02428749 7 | 2864.578604 |
| | 16 | 57151796 | -0.0270351 | 0.00408984 | 3.80E-11 | rs8058512 | T | C | 0.724045 | 0.00379552 | 43.69540577 |
| | 17 | 66823805 | -0.0472863 | 0.00787202 | 1.90E-09 | rs112001035 | A | G | 0.05973 | 0.0031344 | 36.08201155 |
| | 17 | 38505824 | -0.0249324 | 0.00417566 | 2.40E-09 | rs2586116 | G | C | 0.259676 | 0.00309696 | 35.65088911 |

| | | | | | | | | | | | |
|---|---|---|---|---|---|---|---|---|---|---|---|
| | 17 | 41926126 | -0.140843 | 0.010482 | 3.70E-41 | rs72836561 | T | C | 0.031482 | 0.001566369 | 180.5405145 |
| | 18 | 47179516 | 0.0622277 | 0.00512696 | 6.70E-34 | rs1943973 | A | G | 0.849315 | 0.001278453 | 147.3126672 |
| | 19 | 8429323 | 0.194836 | 0.0131209 | 7.00E-50 | rs116843064 | A | G | 0.019851 | 0.001912372 | 220.4974702 |
| | 19 | 11347493 | -0.0447598 | 0.00686512 | 7.00E-11 | rs737337 | C | T | 0.076789 | 0.00369243 | 42.50817512 |
| | 19 | 45173951 | -0.0489735 | 0.00849647 | 8.20E-09 | rs28807203 | C | A | 0.049207 | 0.00288611 | 33.22291091 |
| | 19 | 45444524 | 0.0267934 | 0.00424947 | 2.90E-10 | rs72235114 | CT | C | 0.294931 | 0.00345326 | 39.75379306 |
| | 19 | 45413233 | -0.232335 | 0.00674817 | 1.00E-200 | rs1065853 | T | G | 0.080581 | 0.010195285 | 1185.358451 |
| | 19 | 46397531 | -0.0327829 | 0.00571005 | 9.40E-09 | rs8105375 | T | C | 0.117639 | 0.00286341 | 32.96153042 |
| | 20 | 44563007 | -0.120668 | 0.0047057 | 5.11E-145 | rs1057208 | T | C | 0.186277 | 0.005681381 | 657.5491321 |
| | 20 | 43008981 | -0.0836449 | 0.0111457 | 6.20E-14 | rs150224153 | T | C | 0.029237 | 0.00489153 | 56.31922713 |
| | 20 | 44733110 | 0.0501911 | 0.00914609 | 4.10E-08 | rs111953743 | G | A | 0.042903 | 0.00261614 | 30.11445021 |

| | | | | | | | | | | | |
|---|---|---|---|---|---|---|---|---|---|---|---|
| | 22 | 21916272 | -0.0307789 | 0.0047895 | 1.30E-10 | rs5754102 | A | C | 0.183262 | 0.00358726 | 41.29697934 |
| exposure | chr.exposure | pos.exposure | beta.exposure | se.exposure | pval.exposure | SNP | effect_allele.exposure | other_allele.exposure | eaf.exposure | R2 | F |
| Free cholesterol to total lipids ratio in small HDL \|\| id:ebi-a-GCST90092949 | 1 | 55521109 | 0.0291592 | 0.0040907 | 1.00E-12 | rs693668 | A | G | 0.648549 | 0.00441323 | 50.80990702 |
| | 1 | 110232983 | -0.0239049 | 0.00437748 | 4.70E-08 | rs140584594 | G | A | 0.730101 | 0.00259063 | 29.82070962 |
| | 1 | 62999675 | -0.0223284 | 0.00408589 | 4.60E-08 | rs7534572 | G | C | 0.64747 | 0.00259431 | 29.86306083 |
| | 1 | 177949888 | -0.032972 | 0.00584539 | 1.70E-08 | rs7517442 | T | C | 0.129594 | 0.00276399 | 31.81676719 |
| | 1 | 55505647 | -0.238926 | 0.0148959 | 6.70E-58 | rs11591147 | T | G | 0.017457 | 0.02230571 | 257.2680063 |
| | 1 | 55695535 | 0.0585948 | 0.0101057 | 6.70E-09 | rs2902875 | T | C | 0.039159 | 0.00292046 | 33.61845895 |
| | 1 | 109817590 | -0.110127 | 0.00470344 | 3.10E-121 | rs12740374 | T | G | 0.221064 | 0.04741165 | 548.2123981 |
| | 1 | 161193683 | -0.0487631 | 0.00403852 | 1.40E-33 | rs5082 | A | G | 0.620747 | 0.01265263 | 145.7909583 |
| | 1 | 234852760 | 0.0310851 | 0.00390997 | 1.90E-15 | rs553427 | T | C | 0.517037 | 0.00548924 | 63.20481138 |

| | | | | | | | | | | | |
|---|---|---|---|---|---|---|---|---|---|---|---|
| | 2 | 136608646 | -0.0248042 | 0.00440966 | 1.90E-08 | rs4988235 | A | G | 0.735825 | 0.00274861 | 31.63972434 |
| | 2 | 21288226 | 0.112522 | 0.00507268 | 5.11E-109 | rs563290 | A | G | 0.819463 | 0.00425735 3 | 492.0309308 |
| | 2 | 44055510 | -0.0686434 | 0.00819508 | 5.50E-17 | rs114938914 | T | C | 0.061412 | 0.00609283 | 70.15903881 |
| | 3 | 12450838 | -0.157559 | 0.0234197 | 1.70E-11 | rs147667955 | T | C | 0.007221 | 0.00393139 | 45.26017499 |
| | 3 | 50052867 | 0.0268123 | 0.0046778 | 9.90E-09 | rs6800913 | A | T | 0.749098 | 0.002854 | 32.85317093 |
| | 4 | 69339933 | 0.0258009 | 0.00464118 | 2.70E-08 | rs35662434 | G | A | 0.233712 | 0.00268466 | 30.90333425 |
| | 5 | 74472939 | 0.0301444 | 0.00407728 | 1.40E-13 | rs7707394 | A | G | 0.357075 | 0.00474743 | 54.65937565 |
| | 5 | 74639259 | 0.0398661 | 0.00404436 | 6.40E-23 | rs3843482 | G | T | 0.373802 | 0.00843595 | 97.16287192 |
| | 5 | 156443066 | 0.0248063 | 0.00408214 | 1.20E-09 | rs4704834 | G | A | 0.644028 | 0.00320776 | 36.92671253 |
| | 6 | 42919222 | -0.0265661 | 0.00396164 | 2.00E-11 | rs9471975 | C | T | 0.583391 | 0.00390597 | 44.96742743 |
| | 6 | 25715657 | -0.0629252 | 0.00774671 | 4.60E-16 | rs116009877 | A | G | 0.07117 | 0.00573005 | 65.97920295 |

| | | | | | | | | | | | |
|---|---|---|---|---|---|---|---|---|---|---|---|
| | 6 | 35006409 | -0.0245241 | 0.00447765 | 4.30E-08 | rs2091074 | T | C | 0.742779 | 0.00260595 | 29.99703446 |
| | 6 | 30055813 | -0.0906224 | 0.0159359 | 1.30E-08 | rs142292417 | A | T | 0.015997 | 0.00280924 | 32.33779324 |
| | 6 | 160684867 | -0.0538881 | 0.0097903 | 3.70E-08 | rs74427859 | T | A | 0.042261 | 0.00263192 | 30.29606301 |
| | 6 | 161010118 | 0.0641434 | 0.00725274 | 9.20E-19 | rs10455872 | G | A | 0.078987 | 0.00679199 | 78.21538503 |
| | 7 | 75044830 | -0.0236606 | 0.00414584 | 1.10E-08 | rs236651 | A | G | 0.573853 | 0.00282941 | 32.57008445 |
| | 7 | 1029585 | -0.0383489 | 0.00403082 | 1.80E-21 | rs9769088 | C | T | 0.390936 | 0.00785905 | 90.51310537 |
| | 7 | 94953895 | -0.0228511 | 0.00395383 | 7.50E-09 | rs705379 | A | G | 0.477476 | 0.00290165 | 33.40186292 |
| | 7 | 72878044 | 0.0283885 | 0.00478497 | 3.00E-09 | rs35797675 | G | T | 0.217235 | 0.00305764 | 35.19807443 |
| | 7 | 21686001 | -0.0236275 | 0.00392635 | 1.80E-09 | rs12700289 | T | C | 0.509417 | 0.00314567 | 36.21178848 |
| | 7 | 28194397 | 0.0224357 | 0.00392471 | 1.10E-08 | rs860262 | A | C | 0.500451 | 0.00283879 | 32.6780819 |
| | 8 | 10809490 | -0.0300042 | 0.00396242 | 3.70E-14 | rs34031674 | C | G | 0.434685 | 0.00497989 | 57.33707601 |

| | | | | | | | | | | | |
|---|---|---|---|---|---|---|---|---|---|---|---|
| | 9 | 15304782 | 0.076076 | 0.00563359 | 3.60E-43 | rs686030 | A | C | 0.859548 | 0.01646323 | 189.7712792 |
| | 9 | 107586753 | 0.0424671 | 0.00581558 | 2.80E-13 | rs2066714 | C | T | 0.12893 | 0.00463138 | 53.32263389 |
| | 9 | 107647019 | 0.0559208 | 0.00629708 | 6.70E-19 | rs11789603 | T | C | 0.108822 | 0.00684799 | 78.86072809 |
| | 9 | 107661742 | -0.0586443 | 0.00444267 | 8.70E-40 | rs2740488 | C | A | 0.265313 | 0.01511816 | 174.243247 |
| | 10 | 114091670 | 0.0429789 | 0.0074449 | 7.80E-09 | rs79172158 | C | T | 0.075202 | 0.00289507 | 33.32616245 |
| | 11 | 5701074 | 0.0506912 | 0.00761814 | 2.90E-11 | rs11601507 | A | C | 0.069168 | 0.00384585 | 44.275115 |
| | 11 | 68562328 | -0.0512962 | 0.00784811 | 6.30E-11 | rs2229738 | T | C | 0.066036 | 0.00371083 | 42.72014098 |
| | 11 | 47334524 | 0.0343051 | 0.00513185 | 2.30E-11 | rs10838689 | A | G | 0.175488 | 0.00388145 | 44.68501665 |
| | 11 | 61586328 | -0.103414 | 0.00411558 | 2.50E-139 | rs35473591 | CT | C | 0.34192 | 0.05456485 | 631.3773961 |
| | 11 | 10358139 | -0.0221589 | 0.00394075 | 1.90E-08 | rs2403309 | T | A | 0.469926 | 0.0027467 | 31.61775858 |
| | 11 | 116617895 | 0.0476983 | 0.00865359 | 3.50E-08 | rs45539536 | T | C | 0.053849 | 0.00263931 | 30.38124901 |

| | | | | | | | | | | | |
|---|---|---|---|---|---|---|---|---|---|---|---|
| | 11 | 117111368 | 0.130158 | 0.0174426 | 8.50E-14 | rs187929675 | T | C | 0.013097 | 0.00483617 | 55.68160468 |
| | 11 | 117156893 | 0.158227 | 0.0198908 | 1.80E-15 | rs1047964 | T | G | 0.010073 | 0.00549554 | 63.27747471 |
| | 12 | 111904371 | 0.0221025 | 0.00392013 | 1.70E-08 | rs4766578 | A | T | 0.503234 | 0.00276156 | 31.78881303 |
| | 13 | 95236825 | -0.0279822 | 0.00408471 | 7.40E-12 | rs9556403 | G | A | 0.352423 | 0.00407621 | 46.92817962 |
| | 13 | 32976654 | -0.0325514 | 0.00413752 | 3.60E-15 | rs61946991 | C | A | 0.451649 | 0.00537549 | 61.89443229 |
| | 14 | 69297986 | -0.0287185 | 0.00475219 | 1.50E-09 | rs1296535 | T | C | 0.778482 | 0.00317242 | 36.51979787 |
| | 14 | 70708353 | -0.0236632 | 0.00419291 | 1.70E-08 | rs4902797 | T | C | 0.322448 | 0.00276687 | 31.84991812 |
| | 15 | 58214313 | -0.0658468 | 0.0112125 | 4.30E-09 | rs78756050 | G | A | 0.032203 | 0.0029959 | 34.48711685 |
| | 15 | 58555798 | -0.0545602 | 0.00441184 | 4.00E-35 | rs16939991 | G | C | 0.26718 | 0.01327175 | 152.9342832 |
| | 15 | 58723939 | 0.263855 | 0.00471209 | 1.00E-200 | rs2070895 | A | G | 0.218679 | 0.02652295 | 3135.42167 |
| | 15 | 59080390 | -0.0725604 | 0.0086401 | 4.50E-17 | rs75888243 | C | A | 0.054074 | 0.00612475 | 70.52681162 |

| | | | | | | | | | | | |
|---|---|---|---|---|---|---|---|---|---|---|---|
| | 15 | 58833993 | 0.12304 | 0.0116501 | 4.50E-26 | rs6078 | A | G | 0.028853 | 0.000968289 | 111.5387546 |
| | 15 | 58683366 | -0.214335 | 0.00400122 | 1.00E-200 | rs1532085 | G | A | 0.613962 | 0.024327528 | 2869.417754 |
| | 16 | 15119607 | -0.0369483 | 0.00471567 | 4.70E-15 | rs8059755 | C | T | 0.55864 | 0.00533167 | 61.38956601 |
| | 16 | 57065556 | -0.0273035 | 0.00427559 | 1.70E-10 | rs289754 | T | C | 0.30631 | 0.00354228 | 40.77903943 |
| | 16 | 67976320 | -0.101024 | 0.0108316 | 1.10E-20 | rs4986970 | T | A | 0.033919 | 0.00755315 | 86.98739908 |
| | 16 | 56991363 | 0.147227 | 0.004176 | 1.00E-200 | rs183130 | T | C | 0.32403 | 0.010685161 | 1242.929219 |
| | 16 | 56852822 | 0.0292361 | 0.00395511 | 1.40E-13 | rs1561139 | T | G | 0.42589 | 0.00474579 | 54.64044107 |
| | 17 | 45766771 | 0.026453 | 0.00394285 | 2.00E-11 | rs56325564 | A | G | 0.482691 | 0.00390978 | 45.01132585 |
| | 17 | 64195431 | 0.0895931 | 0.0110636 | 5.60E-16 | rs75003668 | G | A | 0.033354 | 0.0056951 | 65.5765627 |
| | 17 | 66428002 | 0.0309745 | 0.00541045 | 1.00E-08 | rs11077558 | G | C | 0.845623 | 0.00284715 | 32.77436498 |
| | 17 | 67081278 | 0.0914656 | 0.0131609 | 3.70E-12 | rs77542162 | G | A | 0.022666 | 0.00419522 | 48.29885654 |

| | | | | | | | | | | | |
|---|---|---|---|---|---|---|---|---|---|---|---|
| | 18 | 2876010 | 0.0354631 | 0.00636378 | 2.50E-08 | rs113573417 | T | C | 0.128478 | 0.00269773 | 31.05385573 |
| | 19 | 45413233 | -0.426076 | 0.00719923 | 1.00E-200 | rs1065853 | T | G | 0.080581 | 0.029537478 | 3502.631904 |
| | 19 | 45618169 | 0.0347514 | 0.0062032 | 2.10E-08 | rs62118465 | A | C | 0.117516 | 0.00272639 | 31.3837977 |
| | 19 | 11192915 | -0.104463 | 0.0059861 | 3.40E-68 | rs73015021 | G | A | 0.120658 | 0.0026392558 | 304.529595 |
| | 19 | 45039971 | -0.0909308 | 0.0128023 | 1.20E-12 | rs74747585 | C | T | 0.025173 | 0.00438176 | 50.4473638 |
| | 19 | 11147526 | -0.131936 | 0.0187899 | 2.20E-12 | rs73013176 | C | T | 0.011281 | 0.00428237 | 49.30263615 |
| | 19 | 45173951 | -0.108236 | 0.00906438 | 7.30E-33 | rs28807203 | C | A | 0.049207 | 0.01237434 | 142.5803619 |
| | 19 | 45337918 | 0.144563 | 0.0107062 | 1.50E-41 | rs147711004 | A | G | 0.036038 | 0.0158179 | 182.3207307 |
| | 19 | 45421204 | 0.135901 | 0.0119032 | 3.40E-30 | rs150966173 | T | C | 0.027969 | 0.01131407 | 130.3497779 |
| | 19 | 45215081 | -0.0973072 | 0.0138924 | 2.50E-12 | rs111278137 | A | G | 0.021614 | 0.00426131 | 49.06003409 |
| | 20 | 17844684 | -0.0344087 | 0.00413138 | 8.20E-17 | rs2618566 | T | G | 0.660294 | 0.00602389 | 69.36472941 |

| | | | | | | | | | | | |
|---|---|---|---|---|---|---|---|---|---|---|---|
| | 20 | 44563007 | -0.145839 | 0.00503004 | 7.91E-185 | rs1057208 | T | C | 0.186277 | 0.00725164 | 840.6146036 |
| | 20 | 44733110 | 0.0622581 | 0.00977648 | 1.90E-10 | rs111953743 | G | A | 0.042903 | 0.00352262 | 40.55264123 |
| | 22 | 44324730 | 0.0283435 | 0.00475259 | 2.50E-09 | rs738408 | T | C | 0.216547 | 0.00308962 | 35.56630176 |
