## Supplemental Table S3 for "Mendelian Randomization Study Reveals That Combined Spontaneous Free Cholesterol Diffusion and Reverse Cholesterol Transport Pathways Shift from Pro-Atherogenic to Anti-Atherogenic Effects"

### Supplemental Material

Table S3: The results of the Q-test. Outcome: Emergency coronary revascularization (for ACS) (no controls excluded) || id: finn-b-I9_REVACS_EXNONE

| exposure | method | Q | Q_df | Q_pval |
|---|---|---|---|---|
| Free cholesterol levels in chylomicrons and extremely large VLDL \|\| id:ebi-a-GCST90093044 | MR Egger | 89.76694221 | 52 | 0.000891302 |
| Free cholesterol levels in chylomicrons and extremely large VLDL \|\| id:ebi-a-GCST90093044 | Inverse variance weighted | 93.37786186 | 53 | 0.000518593 |
| Free cholesterol levels in very large VLDL \|\| id:ebi-a-GCST90093020 | MR Egger | 370.536728 | 54 | 9.15E-49 |
| Free cholesterol levels in very large VLDL \|\| id:ebi-a-GCST90093020 | Inverse variance weighted | 391.139358 | 55 | 3.37E-52 |
| Free cholesterol levels in large VLDL \|\| id:ebi-a-GCST90092872 | MR Egger | 102.6539659 | 49 | 1.14E-05 |
| Free cholesterol levels in large VLDL \|\| id:ebi-a-GCST90092872 | Inverse variance weighted | 107.0157861 | 50 | 5.00E-06 |
| Free cholesterol levels in medium VLDL \|\| id:ebi-a-GCST90092920 | MR Egger | 257.0045104 | 59 | 1.55E-26 |
| Free cholesterol levels in medium VLDL \|\| id:ebi-a-GCST90092920 | Inverse variance weighted | 266.5611266 | 60 | 7.84E-28 |
| Free cholesterol levels in small VLDL \|\| id:ebi-a-GCST90092972 | MR Egger | 238.9828385 | 54 | 4.13E-25 |
| Free cholesterol levels in small VLDL \|\| id:ebi-a-GCST90092972 | Inverse variance weighted | 244.7135796 | 55 | 9.31E-26 |
| Free cholesterol levels in very small VLDL \|\| id:ebi-a-GCST90093032 | MR Egger | 225.5059844 | 57 | 6.71E-22 |
| Free cholesterol levels in very small VLDL \|\| id:ebi-a-GCST90093032 | Inverse variance weighted | 226.9165914 | 58 | 7.91E-22 |

| | | | | |
|---|---|---|---|---|
| Free cholesterol levels in VLDL \|\| id:ebi-a-GCST90092998 | MR Egger | 249.3799555 | 58 | 1.43E-25 |
| Free cholesterol levels in VLDL \|\| id:ebi-a-GCST90092998 | Inverse variance weighted | 255.2289835 | 59 | 3.10E-26 |
| Free cholesterol levels in IDL \|\| id:ebi-a-GCST90092835 | MR Egger | 328.9589395 | 55 | 1.12E-40 |
| Free cholesterol levels in IDL \|\| id:ebi-a-GCST90092835 | Inverse variance weighted | 341.7781497 | 56 | 1.27E-42 |
| Free cholesterol levels in large LDL \|\| id:ebi-a-GCST90092860 | MR Egger | 0.118081417 | 1 | 0.731124735 |
| Free cholesterol levels in large LDL \|\| id:ebi-a-GCST90092860 | Inverse variance weighted | 0.321212567 | 2 | 0.851627305 |
| Free cholesterol levels in medium LDL \|\| id:ebi-a-GCST90092908 | MR Egger | 132.9041618 | 46 | 2.26E-10 |
| Free cholesterol levels in medium LDL \|\| id:ebi-a-GCST90092908 | Inverse variance weighted | 138.5195564 | 47 | 5.87E-11 |
| Free cholesterol levels in small LDL \|\| id:ebi-a-GCST90092960 | MR Egger | 150.6999033 | 46 | 4.65E-13 |
| Free cholesterol levels in small LDL \|\| id:ebi-a-GCST90092960 | Inverse variance weighted | 156.4043475 | 47 | 1.12E-13 |
| Free cholesterol levels in LDL \|\| id:ebi-a-GCST90092885 | MR Egger | 143.1503845 | 45 | 3.69E-12 |
| Free cholesterol levels in LDL \|\| id:ebi-a-GCST90092885 | Inverse variance weighted | 146.208302 | 46 | 2.29E-12 |
| Total lipid levels in chylomicrons and extremely large VLDL \|\| id:ebi-a-GCST90093046 | MR Egger | 348.8687805 | 54 | 9.78E-45 |
| Total lipid levels in chylomicrons and extremely large VLDL \|\| id:ebi-a-GCST90093046 | Inverse variance weighted | 372.879493 | 55 | 8.84E-49 |

| | | | | |
|---|---|---|---|---|
| Total lipid levels in very large VLDL \|\| id:ebi-a-GCST90093022 | MR Egger | 339.4043076 | 57 | 8.58E-42 |
| Total lipid levels in very large VLDL \|\| id:ebi-a-GCST90093022 | Inverse variance weighted | 350.0866724 | 58 | 2.39E-43 |
| Total lipid levels in large VLDL \|\| id:ebi-a-GCST90092874 | MR Egger | 368.5564169 | 58 | 9.75E-47 |
| Total lipid levels in large VLDL \|\| id:ebi-a-GCST90092874 | Inverse variance weighted | 383.4850096 | 59 | 4.37E-49 |
| Total lipid levels in medium VLDL \|\| id:ebi-a-GCST90092922 | MR Egger | 229.5631544 | 55 | 3.40E-23 |
| Total lipid levels in medium VLDL \|\| id:ebi-a-GCST90092922 | Inverse variance weighted | 232.2609252 | 56 | 2.48E-23 |
| Total lipid levels in small VLDL \|\| id:ebi-a-GCST90092974 | MR Egger | 225.2680678 | 62 | 2.20E-20 |
| Total lipid levels in small VLDL \|\| id:ebi-a-GCST90092974 | Inverse variance weighted | 228.8153741 | 63 | 1.15E-20 |
| Total lipid levels in very small VLDL \|\| id:ebi-a-GCST90093034 | MR Egger | 297.4083657 | 63 | 4.03E-32 |
| Total lipid levels in very small VLDL \|\| id:ebi-a-GCST90093034 | Inverse variance weighted | 297.4104314 | 64 | 8.83E-32 |
| Total lipid levels in IDL \|\| id:ebi-a-GCST90092837 | MR Egger | 197.6966069 | 52 | 7.57E-19 |
| Total lipid levels in IDL \|\| id:ebi-a-GCST90092837 | Inverse variance weighted | 210.8120199 | 53 | 1.07E-20 |
| Total lipid levels in large LDL \|\| id:ebi-a-GCST90092862 | MR Egger | 87.16683244 | 42 | 5.31E-05 |
| Total lipid levels in large LDL \|\| id:ebi-a-GCST90092862 | Inverse variance weighted | 89.49937937 | 43 | 4.12E-05 |

| | | | | |
|---|---|---|---|---|
| Total lipid levels in medium LDL \|\| id:ebi-a-GCST90092910 | MR Egger | 180.1543542 | 44 | 2.15E-18 |
| Total lipid levels in medium LDL \|\| id:ebi-a-GCST90092910 | Inverse variance weighted | 183.3044873 | 45 | 1.32E-18 |
| Total lipid levels in small LDL \|\| id:ebi-a-GCST90092962 | MR Egger | 195.0037347 | 46 | 2.97E-20 |
| Total lipid levels in small LDL \|\| id:ebi-a-GCST90092962 | Inverse variance weighted | 195.4830486 | 47 | 5.14E-20 |
| Free cholesterol to total lipids ratio in chylomicrons and extremely large VLDL \|\| id:ebi-a-GCST90093045 | MR Egger | 94.55421589 | 13 | 1.87E-14 |
| Free cholesterol to total lipids ratio in chylomicrons and extremely large VLDL \|\| id:ebi-a-GCST90093045 | Inverse variance weighted | 98.51980568 | 14 | 9.11E-15 |
| Free cholesterol to total lipids ratio in very large VLDL \|\| id:ebi-a-GCST90093021 | MR Egger | 127.8213242 | 43 | 2.38E-10 |
| Free cholesterol to total lipids ratio in very large VLDL \|\| id:ebi-a-GCST90093021 | Inverse variance weighted | 130.186352 | 44 | 1.87E-10 |
| Free cholesterol to total lipids ratio in large VLDL \|\| id:ebi-a-GCST90092873 | MR Egger | 343.924155 | 44 | 4.07E-48 |
| Free cholesterol to total lipids ratio in large VLDL \|\| id:ebi-a-GCST90092873 | Inverse variance weighted | 346.383501 | 45 | 3.91E-48 |
| Free cholesterol to total lipids ratio in medium VLDL \|\| id:ebi-a-GCST90092921 | MR Egger | 154.2415722 | 51 | 2.60E-12 |

| | | | | |
|---|---|---|---|---|
| Free cholesterol to total lipids ratio in medium VLDL \|\| id:ebi-a-GCST90092921 | Inverse variance weighted | 154.5874717 | 52 | 4.08E-12 |
| Free cholesterol to total lipids ratio in small VLDL \|\| id:ebi-a-GCST90092973 | MR Egger | 561.6820202 | 65 | 3.50E-80 |
| Free cholesterol to total lipids ratio in small VLDL \|\| id:ebi-a-GCST90092973 | Inverse variance weighted | 575.6410637 | 66 | 2.11E-82 |
| Free cholesterol to total lipids ratio in very small VLDL \|\| id:ebi-a-GCST90093033 | MR Egger | 409.0892422 | 46 | 1.01E-59 |
| Free cholesterol to total lipids ratio in very small VLDL \|\| id:ebi-a-GCST90093033 | Inverse variance weighted | 412.5610053 | 47 | 6.45E-60 |
| Free cholesterol to total lipids ratio in IDL \|\| id:ebi-a-GCST90092836 | MR Egger | 344.5779561 | 43 | 1.07E-48 |
| Free cholesterol to total lipids ratio in IDL \|\| id:ebi-a-GCST90092836 | Inverse variance weighted | 345.8413674 | 44 | 1.75E-48 |
| Free cholesterol to total lipids ratio in large LDL \|\| id:ebi-a-GCST90092861 | MR Egger | 496.6563564 | 79 | 8.34E-62 |
| Free cholesterol to total lipids ratio in large LDL \|\| id:ebi-a-GCST90092861 | Inverse variance weighted | 500.6145343 | 80 | 3.95E-62 |
| Free cholesterol to total lipids ratio in medium LDL \|\| id:ebi-a-GCST90092909 | MR Egger | 404.5940839 | 69 | 5.88E-49 |
| Free cholesterol to total lipids ratio in medium LDL \|\| id:ebi-a-GCST90092909 | Inverse variance weighted | 408.6778007 | 70 | 2.61E-49 |

| | | | | |
|---|---|---|---|---|
| Free cholesterol to total lipids ratio in small LDL \|\| id:ebi-a-GCST90092961 | MR Egger | 425.1567383 | 67 | 1.65E-53 |
| Free cholesterol to total lipids ratio in small LDL \|\| id:ebi-a-GCST90092961 | Inverse variance weighted | 439.8590363 | 68 | 8.18E-56 |
| Free cholesterol levels in very large HDL \|\| id:ebi-a-GCST90093008 | MR Egger | 357.6796741 | 69 | 1.50E-40 |
| Free cholesterol levels in very large HDL \|\| id:ebi-a-GCST90093008 | Inverse variance weighted | 369.4719035 | 70 | 2.82E-42 |
| Free cholesterol levels in large HDL \|\| id:ebi-a-GCST90092848 | MR Egger | 284.5849696 | 85 | 2.36E-23 |
| Free cholesterol levels in large HDL \|\| id:ebi-a-GCST90092848 | Inverse variance weighted | 296.7935947 | 86 | 5.59E-25 |
| Free cholesterol levels in medium HDL \|\| id:ebi-a-GCST90092896 | MR Egger | 276.4798888 | 73 | 1.96E-25 |
| Free cholesterol levels in medium HDL \|\| id:ebi-a-GCST90092896 | Inverse variance weighted | 277.3541374 | 74 | 2.78E-25 |
| Free cholesterol levels in small HDL \|\| id:ebi-a-GCST90092948 | MR Egger | 122.9341548 | 45 | 3.68E-09 |
| Free cholesterol levels in small HDL \|\| id:ebi-a-GCST90092948 | Inverse variance weighted | 122.9355773 | 46 | 6.18E-09 |
| Free cholesterol levels in HDL \|\| id:ebi-a-GCST90092824 | MR Egger | 265.1891961 | 74 | 2.46E-23 |
| Free cholesterol levels in HDL \|\| id:ebi-a-GCST90092824 | Inverse variance weighted | 265.8705479 | 75 | 3.67E-23 |
| Total lipid levels in very large HDL \|\| id:ebi-a-GCST90093010 | MR Egger | 390.7163345 | 75 | 3.22E-44 |
| Total lipid levels in very large HDL \|\| id:ebi-a-GCST90093010 | Inverse variance weighted | 409.6242761 | 76 | 3.30E-47 |

| | | | | |
|---|---|---|---|---|
| Total lipid levels in large HDL \|\| id:ebi-a-GCST90092850 | MR Egger | 289.7906012 | 89 | 4.27E-23 |
| Total lipid levels in large HDL \|\| id:ebi-a-GCST90092850 | Inverse variance weighted | 297.0538212 | 90 | 6.04E-24 |
| Total lipid levels in medium HDL \|\| id:ebi-a-GCST90092898 | MR Egger | 237.8203413 | 62 | 2.07E-22 |
| Total lipid levels in medium HDL \|\| id:ebi-a-GCST90092898 | Inverse variance weighted | 238.34374 | 63 | 3.36E-22 |
| Total lipid levels in small HDL \|\| id:ebi-a-GCST90092950 | MR Egger | 156.8412504 | 44 | 1.41E-14 |
| Total lipid levels in small HDL \|\| id:ebi-a-GCST90092950 | Inverse variance weighted | 158.0060131 | 45 | 1.77E-14 |
| Free cholesterol to total lipids ratio in very large HDL \|\| id:ebi-a-GCST90093009 | MR Egger | 179.3084684 | 74 | 1.01E-10 |
| Free cholesterol to total lipids ratio in very large HDL \|\| id:ebi-a-GCST90093009 | Inverse variance weighted | 185.6413341 | 75 | 2.33E-11 |
| Free cholesterol to total lipids ratio in large HDL \|\| id:ebi-a-GCST90092849 | MR Egger | 327.5476603 | 60 | 1.68E-38 |
| Free cholesterol to total lipids ratio in large HDL \|\| id:ebi-a-GCST90092849 | Inverse variance weighted | 343.829723 | 61 | 4.76E-41 |
| Free cholesterol to total lipids ratio in medium HDL \|\| id:ebi-a-GCST90092897 | MR Egger | 380.5385163 | 81 | 2.47E-40 |
| Free cholesterol to total lipids ratio in medium HDL \|\| id:ebi-a-GCST90092897 | Inverse variance weighted | 381.5347393 | 82 | 3.64E-40 |
| Free cholesterol to total lipids ratio in | MR Egger | 316.8041041 | 66 | 1.89E-34 |

| | | | | |
|---|---|---|---|---|
| small HDL \|\| id:ebi-a-GCST90092949 | | | | |
| Free cholesterol to total lipids ratio in small HDL \|\| id:ebi-a-GCST90092949 | Inverse variance weighted | 319.0587093 | 67 | 1.70E-34 |
